# Prevalence, clusters, and burden of complex tuberculosis multimorbidity in low- and middle-income countries: a systematic review and meta-analysis

**DOI:** 10.1101/2022.09.22.22280228

**Authors:** Alexander Jarde, Ruimin Ma, Olamide O. Todowede, Abdul Latif, Aashifa Yaqoob, Saima Afaq, Tarana Ferdous, Ewan M Tomeny, Laura Rosu, Lucy Elauteri Mrema, Shagoofa Rakhshanda, Sheraz Fazid, Farah Naz Rahman, Uzochukwu Egere, Asma Elsony, Helen Elsey, Kamran Siddiqi, Brendon Stubbs, Najma Siddiqi

## Abstract

**Objectives:** The co-occurrence of tuberculosis (TB) and multiple chronic communicable and non-communicable diseases is a growing concern in low- and middle-income countries (LMICs). Understanding this ‘complex TB multimorbidity’ (cTBMM) can contribute to improved patient care. We aimed to synthesise the prevalence of clusters of cTBMM and their impact on patient outcomes.

**Design:** Systematic review and meta-analysis.

**Setting:** LMICs.

**Papers:** We searched major databases from inception to 01/10/2020 to identify observational studies reporting primary (prevalence or risk) or secondary (related to disease burden) outcome data for people in LMICs with cTBMM. Titles, abstracts and full text screening, data extraction, and quality assessment were done in duplicate by independent reviewers, and random-effects meta-analyses were performed.

**Primary and secondary outcome measures:** Prevalence or risk of cTBMM (primary); any measure of burden of disease (secondary).

**Results:** From 21,884 search results, 82 studies representing 773,828 TB patients were included, reporting data on 78 different clusters of cTBMM. In TB patients, the most prevalent co-occurring conditions where depression and anxiety (15.3%, 95% confidence interval [CI] 10.7%-20.5%, 3 studies, 1,473 participants, I^2^=64%), HIV and anxiety (15.2%, 95%CI 11.3%-19.7%, 4 studies, 1,413 participants, I^2^=77%), and HIV and post-traumatic stress disorder (14.8%, 95%CI 13.9%-15.8%, 2 studies, 5,400 participants). Sparse evidence indicated lower treatment success and higher risks of death in people with cTBMM.

**Conclusions:** Although limited by high heterogeneity, this first systematic review and meta-analysis on cTBMM highlights that multiple conditions co-occurring in TB patients are common and that mental disorders often cluster together or with HIV. Further research assessing the burden of cTBMM and identifying effective health systems responses is required.

**Registration:** Prospero CRD42020214021.

**Strengths and limitations of this study:**

- To our knowledge, this is the first systematic review summarising the prevalence of the different clusters of complex TB multimorbidity (cTBMM) in LMICs.
- While our focus was on cTBMM in LMICs overall, subgroup analyses by type of TB, country, and socio-economic group allow for a more nuanced assessment on cTBMM.
- We used a broad definition for the term ‘chronic condition’ to be as inclusive as possible, but which could lead to heterogeneity in the definition of some clusters of cTBMM.

## BACKGROUND

Tuberculosis (TB) is one of the leading cause of mortality from a single infectious disease globally.[1] With 1.4 million people dying from TB in 2019, only COVID-19 (with ∼2 million deaths in 2020) is responsible for more deaths.[2] TB frequently co-occurs with non-communicable diseases (NCDs), including diabetes, depression, and cancer.[3] Moreover, the relationship between TB and NCDs may be bidirectional[3–5] and several NCDs, such as diabetes and depression, are considered to be risk factors for TB.[6,7] Similarly, chronic communicable diseases (CCDs) such as HIV and TB adversely interact with each other at the population, individual, cellular, and molecular levels.[8]

Multimorbidity, defined as the co-occurrence of two or more chronic conditions in a single individual at one point in time, is also a growing global health concern.[9] In high-income countries (HICs) 30% of adults experience multimorbidity, and its prevalence is also increasing in low- and middle-income countries (LMICs),[10] where NCDs are on the rise[11] and CCDs such as HIV remain a major public health issue.[12] Multimorbidity represents a substantial burden for individuals, increasing their risk of adverse events,[13,14] and puts health systems under strain,[9] particularly in LMICs.

Regarding multimorbidity in people with TB, results from meta-analyses in LMICs indicate that about half of all TB patients had depression,[15] one third in sub-Saharan Africa had HIV,[16] and approximately 15% had diabetes mellitus (DM).[17] The co-occurrence of TB with individual chronic conditions is associated with a range of adverse outcomes, such as higher odds of unsuccessful treatment, non-adherence, or mortality.[18–20] Furthermore, people with TB and an additional chronic condition tend to have significantly worse treatment outcomes than people with TB alone;[13,20–22] and people with TB and multiple chronic conditions, i.e. complex TB multimorbidity (cTBMM),[23] may be at a higher risk of death than people with TB alone or TB with only one chronic condition.[13] While reviews have identified the prevalence and risks associated with TB and individual chronic conditions, there has been no systematic review summarising the evidence concerning the prevalence and associated burden of cTBMM.

Therefore, we conducted a systematic review and meta-analysis of observational studies reporting the prevalence (or incidence), or relative risk (or odds ratio, compared to people without TB) of different disease clusters (combinations of two or more chronic conditions), in people with TB in LMICs. In addition, we attempted to identify the disease burden in people with cTBMM, the effect of chronic conditions on TB outcomes (compared to people with only TB), and the effect of TB on the outcomes of comorbid chronic conditions (compared with people with those conditions but without TB).

## METHODS

The protocol for this systematic review and meta-analysis was registered in PROSPERO (CRD42020214021). We have followed the MOOSE reporting guidelines.[24]

### Search strategy

We searched for articles published since inception to 01/10/2020 in: Medline (Ovid), Embase (Ovid), PsycINFO (Ovid), Social Sciences Citation Index (Web of Science), Science Citation Index (Web of Science), Emerging Sources Citation Index, Conference Proceedings Citation Index (Web of Science), and the WHO Global Index Medicus. To identify further unpublished studies, we searched in the Open Grey database and contacted authors of published conference abstracts to inquire for full text publications (with a follow-up email after one to two weeks). Reference lists of included studies were reviewed. No restrictions were set on study language or date of publication.

We used controlled vocabulary (e.g., exploded MeSH terms for Medline) and free text terms for CCDs, NCDs, and mental disorders, and combined them with terms for ‘tuberculosis’ and ‘comorbidities’ using Boolean operators (Supplemental file).

### Selection criteria

We included observational studies (cross-sectional, case-control, and cohort studies) reporting data for people in LMICs with any type of TB (pulmonary/extra-pulmonary, drug-resistant/susceptible) and at least two additional chronic conditions (i.e., cTBMM). Given the variety of definitions for the term ‘chronic’, we used the inclusive definition suggested by Bernell and Howard, as “something that is continuing or occurring again and again for a long time”.[25] This included, but was not limited to, heart disease, diabetes mellitus (DM), chronic lung disease (e.g. chronic obstructive pulmonary disease), HIV, AIDS, hepatitis B and C, depression, and anxiety disorders. Conditions arising because of TB treatment were not included. In cases of doubt, the consideration of a condition as chronic was decided by consensus of four authors with clinical/research expertise, and reference to the protocol (KS, NS, HE, BS) (Supplementary Table 1 for details). Studies were only included if they reported data in enough detail for calculation of at least one outcome.

A number of protocol changes were made post-registration. We decided to limit our population of interest to the general TB population, excluding studies focusing on children. We also decided to exclude studies focusing on specific subgroups such as healthcare workers and incarcerated people to increase generalisability of findings. Studies focused on patients with a specific type of TB (e.g., extra-pulmonary) remained eligible provided they were not from a ‘specific population’.

### Primary outcomes

Primary outcomes included prevalence (or incidence) of any cluster of two or more chronic conditions in people with TB, and the relative risk (RR, or odds ratio) of each cluster of two or more chronic conditions in people with TB compared to people without TB.

### Secondary outcomes

Secondary outcomes included all measures of disease burden (e.g., years lived with a disability, disability-adjusted life years, mortality), TB treatment outcomes (e.g., death, loss to follow-up, treatment failure, treatment success), and outcomes related to the additional chronic conditions.

### Study selection

We used Covidence (https://www.covidence.org/) for the study selection process, data extraction, and quality assessment. Multiple pairs of authors screened and extracted data as per protocol. After removing ineligible, duplicate, and overlapping papers, two reviewers independently reviewed the full texts of potentially eligible papers against our inclusion and exclusion criteria. Disagreements were resolved by discussion, with a third reviewer available as an arbitrator if consensus could not be reached. Studies reported in languages understood by only one of the reviewers (e.g., Portuguese, Spanish, or French) were assessed by that reviewer, with a second reviewer using Google Translate (https://translate.google.com/) to make the assessment.

### Data extraction

Data were extracted by two reviewers using a piloted form, including study characteristics (reference information, country, study design, data collection period, setting, inclusion and exclusion criteria), sample characteristics (sample size, gender, age, socio-economic status, type of TB, potential confounding factors), and outcome data (overall and stratified by age, gender, socio-economic status, and country). If studies reported data for cTBMM without specifying the chronic conditions, corresponding authors were contacted (twice in one month) for clarification or for additional data.

### Quality assessment

The quality of included observational studies was assessed by two reviewers (arbitrated by a third) using a modified version of the Newcastle Ottawa Scales (NOS, Appendix 1 in supplementary material).[26] Rather than assessing the quality of the studies overall, we assessed the quality of specific outcome data of interest, in line with Cochrane’s recommendation of doing separate risk-of-bias assessments for each trial’s result.[27] For instance, in studies where we extracted outcome data only from a cohort’s baseline characteristics table, quality was assessed as if it were a cross-sectional study. Consequently, one study could be assigned more than one quality score if data for more than one outcome were extracted (e.g., for different cTBMM clusters).

### Data synthesis

Data for each cluster of two or more chronic conditions alongside TB were analysed separately, with the main analyses including the most adjusted effect sizes. First, we described each study in a summary table. Second, random-effects meta-analyses were undertaken to pool the prevalence and risk estimates. Statistical heterogeneity was assessed using the I² statistic and, when pooling prevalence values, we applied the Freeman-Tukey double arcsine transformation to stabilise variances.[28,29] Third, we performed subgroup analyses, where possible, by age, gender, type of TB, socio-economic group (according to the World Bank’s classification),[30] and country, as well as subgroups defined by the studies’ quality assessment (two or fewer stars, i.e. “low quality”, vs those of three or more, i.e. “high quality”). In studies reporting both raw and adjusted data, subgroup analyses pooling separately raw and adjusted data were also planned. Fourth, when there were 10 or more studies in a meta-analysis we assessed the potential for publication bias using funnel plots (visual inspection) and Duval and Tweedie’s trim-and-fill method.[31]

The following changes were made post-registration: given variety in definitions and operationalisations regarding depression, anxiety, alcoholism, drug use, chronic lung disease, and cardiovascular disease, we conducted post-hoc subgroup analyses with more homogeneous definitions (e.g., by depression scales).

All meta-analyses were conducted in STATA 17 using the *metaprop* command.[29]

### Patient and Public Involvement

Patients or the public were not involved in the design, or conduct, or reporting, or dissemination plans of our research.

Ethical approval was not required for this study, as it is a systematic review. The funder of the study had no role in study design, data collection, data analysis, data interpretation, or writing of the report.

## RESULTS

### Study and participant characteristics

From 21,884 search results, the full texts of 659 articles were assessed for inclusion, 577 of which were excluded (Appendix 2), leaving 82 studies included in our analyses (Figure 1, Appendix 3).

**Figure 1:**
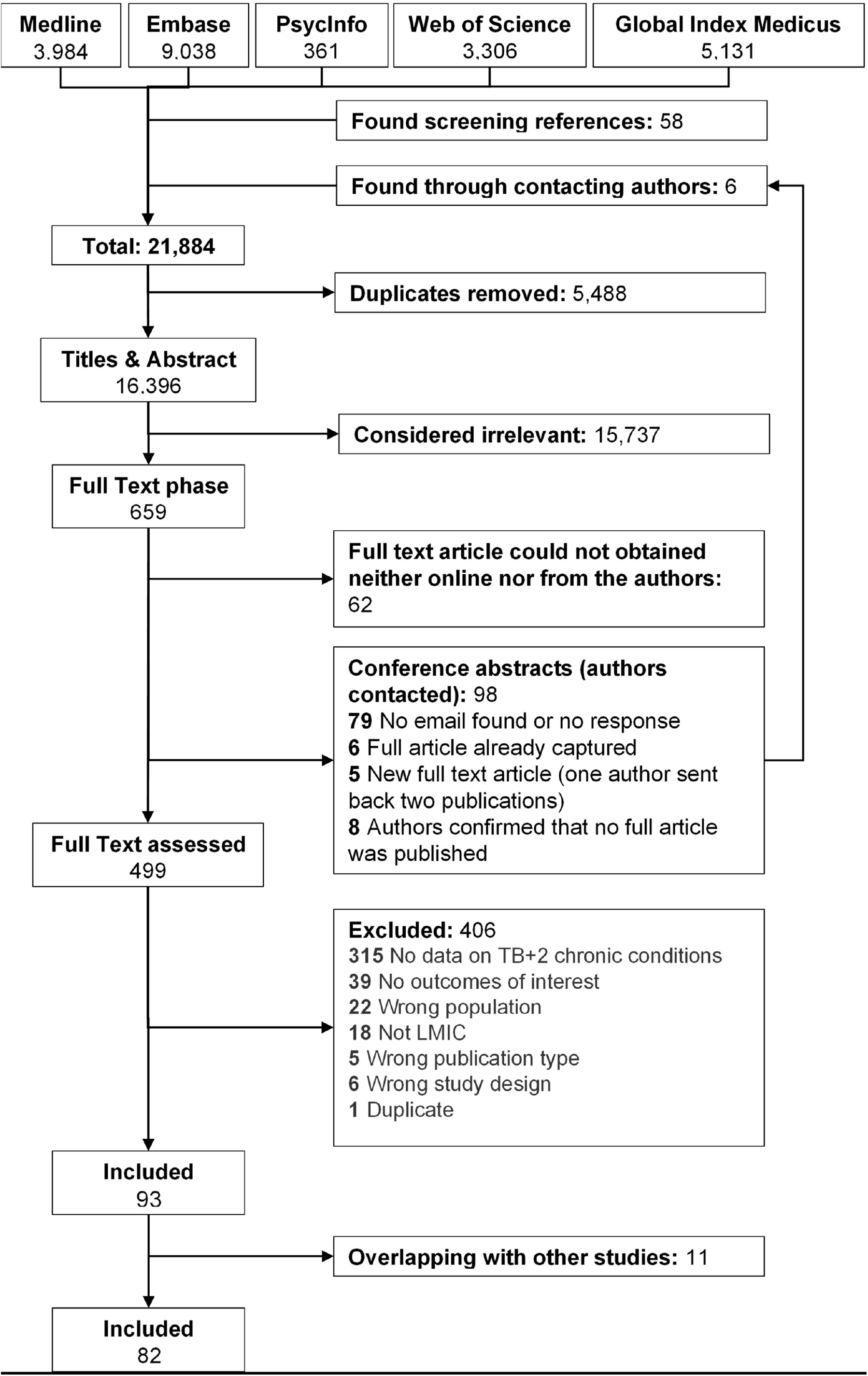
Flow diagram of included and excluded studies

Of these 82 included studies, 36 were conducted in upper-middle-income countries.[S1–S36] The countries with the most studies were India (11 studies),[S37–S47] South Africa (11 studies),[S4,S6,S10,S12– S14,S16,S22,S26,S29,S35] Ethiopia (10 studies), [S48–S57] and Brazil (9 studies).[S1,S3,S5,S9,S15,S19,S21,S31,S33] The data collection period ranged from 1997 to 2019 (Supplementary Table 2) and a total of 773,828 people with TB (46% female) were included. In study design, 58 were cross-sectional,[S1–S4,S6,S8–S10,S14–S17,S19–S24,S27–S35,S37–S41,S43,S45,S46,S48–S70] two were case-control,[S18,S71] and 22 were cohort studies. Of these 22, all reported prevalence data,[S5,S7,S11– S13,S25,S26,S36,S42,S44,S47,S72–S82] with only four[S13,S25,S42,S81] reporting longitudinal data of interest (treatment outcomes). Three of the included studies were published in a non-English language (Spanish).[S2,S24,S27] We identified 178 different cTBMM clusters, i.e. combinations of TB and two or more chronic conditions. For most cTBMM clusters the type of TB was not specified (129 clusters). Pulmonary TB (PTB) was specified in 26 clusters, and multidrug-resistant TB (MDR-TB) in 12. The most common chronic conditions reported as part of a cTBMM cluster were HIV (105 clusters), DM (78 clusters) and depression (20 clusters). The cTBMM clusters most commonly reported were TB and HIV+DM (42 studies),[S2,S3,S5– S7,S11,S13,S14,S17,S18,S22,S23,S25–S27,S29,S33,S35–S39,S41–S45,S47,S54,S55,S57,S61– S63,S65,S67,S68,S71,S73,S77,S79,S80] followed by TB and HIV+depression (10 studies).[S1,S48– S53,S56,S69,S70] Most remaining cTBMM clusters were reported in only one or two studies (Supplementary Table 2).

### Primary outcomes

Table 1 shows the pooled prevalence of each cTBMM cluster reported in at least two studies. The most prevalent clusters involved a mental health condition, either with another mental health condition or with HIV. Three clusters had pooled prevalence values over 10%: TB+depression+anxiety (15.3%, 95%CI 10.7%-20.5%, 3 studies, 1,473 participants), TB+HIV+anxiety (15.2%, 95%CI 11.3%-19.7%, 4 studies, 1,413 participants), and TB+HIV+post-traumatic stress disorder (PTSD, 14.8%, 95%CI 13.9%-15.8%, 2 studies, 5,400 participants); albeit with estimates ranging from 2.4% to 22.1% and with only two to four studies contributing to each meta-analysis (Table 1).

**Table 1:** Meta-analysis of the pooled prevalence of each cluster of tuberculosis + two or more chronic conditions reported in at least two studies.

| Cluster | Prevalence (95%CI)* | Number of studies (participants) | Range | I<sup>2</sup>† |
| --- | --- | --- | --- | --- |
| TB+Depression + Anxiety | 15.27% (10.70% - 20.47%) | 3 (1,473) | 12.78% - 22.09% | 63.81% |
| TB+HIV + Anxiety | 15.23% (11.25% - 19.67%) | 4 (1,413) | 10.07% - 16.40% | 77.37% |
| TB+HIV + PTSD | 14.84% (13.9%-15.8%) | 2 (5,400) | 2.40% - 16.63% | .. |
| TB+HIV + HBV | 9.70% (2.37%-21.04%) | 3 (792) | 2.21% - 20.57% | 94.81% |
| TB+HIV + Depression | 9.66% (6.96% - 12.74%) | 9 (3,418) | 5.06% - 16.28% | 87.47% |
| TB+HIV + Alcoholism | 9.29% (4.98%-14.72%) | 4 (6,963) | 5.00%-16.55% | 96.34% |
| TB+DM + HCV | 7.41% (4.99%-10.24%) | 2 (395) | 2.19%-11.24% | .. |
| TB+HIV + Hypothyroidism | 5.37% (0.90% - 12.94%) | 3 (1,108) | 2.30% - 10.84% | 94.00% |
| TB+HIV + Drug use | 5.21% (2.73% - 8.39%) | 4 (2,584) | 2.74% - 5.87% | 86.19% |
| TB+Drug use + HBV | 3.44% (0% - 12.05%) | 3 (763) | 0% - 9.78% | 95.20% |
| TB+Depression + Chronic lung disease | 1.73% (0.65% - 3.24%) | 2 (437) | 1.15% - 2.86% | .. |
| TB+HBV + HCV | 1.56% (0.60%-2.90%) | 2 (500) | 1.00%-2.00% | .. |
| TB+HIV + HCV | 1.47% (0.36% - 3.21%) | 6 (1,550) | 0.33% - 4.98% | 79.36% |
| TB+DM + Drug use | 1.03% (0.22% - 2.34%) | 5 (7,476) | 0.17% - 6.76% | 90.85% |
| TB+HIV + DM | 0.98% (0.70% - 1.29%) | 40 (526,827) | 0% - 8.33% | 97.79% |
| TB+DM + Alcoholism | 0.69% (0.47% - 0.94%) | 2 (5,218) | 0.63% - 0.76% | .. |
| TB+DM + Chronic kidney disease | 0.50% (0.20% - 0.89%) | 2 (2,275) | 0.55% - 12.22% | .. |
| TB+HIV + Chronic liver disease | 0.48% (0.03% - 1.27%) | 2 (682) | 0.50% - 2.33% | .. |
| TB+DM + Chronic lung disease | 0.23% (0%-0.98%) | 3 (45,143) | 0.03%-1.57% | 96.08% |
| TB+HIV + Chronic lung disease | 0.10% (0% - 0.64%) | 2 (682) | 0.17% - 1.16% | .. |
| TB+DM + Heart disease | 0.05% (0.02% - 0.07%) | 2 (40,495) | 0.07% - 2.28% | .. |
| TB+DM + Chronic liver disease | 0% (0% - 0%) | 2 (181,485) | 0% - 0.03% | .. |

Notes: DM: Diabetes Mellitus; HBV: Hepatitis B Virus; HCV: Hepatitis C Virus; HIV: Human immunodeficiency virus; PTSD: Post-traumatic stress disorder; TB: Tuberculosis.
\* Pooled prevalence and 95% confidence intervals from random-effects meta-analysis when the number of studies is $>1$ .
† I-squared values reported only for meta-analyses with $>2$ studies

Among the cTBMM clusters reported in only one study, only TB+drug use+alcoholism had a prevalence ≥10% (10.0%, 160 participants). Overall, most other clusters (65 of 78) had a prevalence under 1% (Supplementary Table 3).

Four studies reported sufficient data to calculate the pooled RR of having HIV+DM in TB patients compared to non-TB patients, which showed a five-fold increased risk among TB patients (RR 5.5, 95%CI 2.3-13.2, 4 studies, 230,319 participants, I^2^ = 82%). Moreover, from the data reported in one study (148 participants)[S18] we calculated a RR of 66.8 (95%CI 14.8-301.9) of DM+drug use in TB patients compared to non-TB patients.

### Secondary outcomes

Table 2 shows the pooled estimates for prevalence of secondary outcomes in each cTBMM cluster reported in at least two studies. The treatment success rate (patients who were cured or completed treatment) was lowest among people with TB+HIV+drug use (25.4%, 95%CI 22.6%-28.4%, 2 studies, 1,690 participants). Among the cTBMM clusters reported in only one study (Supplementary Table 4), treatment success was reported to be as low as 3.9% (95%CI 0.1%-19.6%) in TB+HIV+silicosis, 3.5% (95%CI 0.1%-17.8%) in TB+HIV+chronic kidney disease, and 0% (95%CI 0%-70.8%) in TB+HIV+mental disorder.

**Table 2:** Meta-analysis of the pooled secondary outcomes of each cluster of tuberculosis + two or more chronic conditions reported in at least two studies.

| Cluster and outcomes | Prevalence (95%CI)* | Number of studies (participants) | Range | I<sup>2</sup>† |
| --- | --- | --- | --- | --- |
| **TB+HIV + DM** |  |  |  |  |
| Treatment success | 36.30% (15.63% - 59.33%) | 5 (4,903) | 21.05% - 100% | 59.16% |
| Treatment failure | 62.17% (36.55% - 85.13%) | 4 (4,723) | 0% - 78.95% | 69.26% |
| **TB+HIV + Drug use** |  |  |  |  |
| Treatment success | 25.44% (22.62% - 28.36%) | 2 (1,690) | 21.43% - 25.89% | .. |
| **TB+HIV + Hepatitis** |  |  |  |  |
| Treatment success | 54.29% (44.38% - 64.04%) | 2 (2,132) | 4.39% - 70.89% | .. |
| Treatment failure | 45.71% (35.96% - 55.62%) | 2 (2,132) | 29.11% - 95.65% | .. |

*Note:* Treatment success represents patients who were cured or completed treatment; treatment failure represents patients where sputum smear or culture remained positive at month five or later during treatment. DM: Diabetes Mellitus; HIV: Human immunodeficiency virus; TB: Tuberculosis
\* Pooled prevalence and 95% confidence intervals from random-effects meta-analysis when the number of studies is >1.
† I-squared values reported only for meta-analyses with >2 studies

Based on individual participant data provided by the authors of one study,[S21] we calculated RRs of each treatment outcome (death, cure and loss to follow-up) in people with cTBMM compared to people with TB alone (Supplementary Table 5). While the number of patients in each cTBMM cluster was very small, in the clusters with more than 100 patients, the RRs of death were significantly higher in the patients with cTBMM, compared to patients with TB alone. In patients with TB+DM+mental disorder, the RR of death was 1.9 (95%CI 1.3-2.8, 118 participants), in TB+AIDS+DM, the RR of death was 4.7 (95%CI 3.8-5.8, 110 participants), and in TB+AIDS+mental disorder, the RR of death was 4.0 (95%CI 3.2-5.0, 143 participants).

Finally, one study[S1] assessed the quality of life (QoL) in people with TB+HIV+depression and TB+HIV+anxiety, reporting the mean values and standard deviation for each domain of the World Health Organization QoL scale for people with HIV (WHOQOL-HIV, Supplementary Table 2). Another study[S49] used this scale to report that 63.2% and 74.7% of people with TB+HIV+depression had poor physical health and poor psychological health, respectively.

### Subgroup analyses

We were unable to conduct subgroup analyses by gender or age, given the lack of data reported separately for each group (only one study[S54] reported the prevalence of PTB+HIV+DM separately for females, 3.7%, and males, 6.0%). For the cTBMM clusters in which subgroup analyses by type of TB were feasible, we found higher prevalence values among studies focused on people with PTB than in studies where the type of TB was not specified, although differences were not always statistically significant, and heterogeneity remained high (Table 3). Similarly, the few studies focused on drug resistant TB and MDR-TB also tended to report a higher prevalence of cTBMM.

**Table 3:** Subgroup analyses pooling prevalence of each cluster of tuberculosis + two or more chronic conditions by type of tuberculosis.

| Cluster and subgroups (p-values)<sup>‡</sup> | Prevalence (95%CI)<sup>*</sup> | Number of studies | range | I<sup>2</sup><sup>†</sup> |
| --- | --- | --- | --- | --- |
| **TB+ Depression + Anxiety** | **15.27% (10.70% - 20.47%)** | **3 (1,473)** | **12.78% - 22.09%** | **63.81%** |
| *By type of TB (p=0.59)* |  |  |  |  |
| Pulmonary TB | 13.14% (11.36% - 15.02%) | 2 (1,338) | 12.78% - 22.09% | .. |
| Multi-drug resistant TB | 14.81% (9.8% - 21.78%) | 1 (135) | .. | .. |
| **TB+HIV + Anxiety** | **15.23% (11.25% - 19.67%)** | **4 (1,413)** | **10.07% - 16.40%** | **77.37%** |
| *By type of TB (p=0.08)* |  |  |  |  |
| TB (unspecified) | 14.08% (10.21% - 18.45%) | 3 (1,327) | 10.07% - 16.40% | .. |
| Pulmonary TB | 22.09% (14.62% - 31.95%) | 1 (86) | .. | .. |
| **TB+HIV + PTSD** | **14.84% (13.9%-15.8%)** | **2 (5,400)** | **2.40% - 16.63%** | **..** |
| *By type of TB: all were TB (unspecified)* |  |  |  |  |
| **TB+HIV + HBV** | **9.70% (2.37%-21.04%)** | **3 (792)** | **2.21% - 20.57%** | **94.81%** |
| *By type of TB: all were TB (unspecified)* |  |  |  |  |
| **TB+HIV + Depression** | **9.66% (6.96% - 12.74%)** | **9 (3,418)** | **5.06% - 16.28%** | **87.47%** |
| *By type of TB (p=0.06)* |  |  |  |  |
| TB (unspecified) | 9.17% (6.48% - 12.26%) | 8 (3,332) | 5.06% - 17.07% | 88.12% |
| Pulmonary TB | 16.28% (9.95% - 25.49%) | 1 (86) | .. | .. |
| **TB+HIV + Alcoholism** | **9.29% (4.98%-14.72%)** | **4 (6,963)** | **5.00%-16.55%** | **96.34%** |
| *By type of TB (p=0.31)* |  |  |  |  |
| TB (unspecified) | 10.24% (5.22% - 16.68%) | 3 (6,877) | 5.00% - 16.55% | .. |
| Pulmonary TB | 5.81% (2.51% - 12.9%) | 1 (86) | .. | .. |
| **TB+DM + HCV** | **7.41% (4.99%-10.24%)** | **2 (395)** | **2.19%-11.24%** | **..** |
| *By type of TB (p<0.005)* |  |  |  |  |
| TB (unspecified) | 2.19% (0.75% - 6.24%) | 1 (137) | .. | .. |
| Pulmonary TB | 11.24% (7.94% - 15.68%) | 1 (258) | .. | .. |
| **TB+HIV + Hypothyroidism** | **5.37% (0.90% - 12.94%)** | **3 (1,108)** | **2.30% - 10.84%** | **94.00%** |
| *By type of TB (p<0.005)* |  |  |  |  |
| Drug resistant TB | 2.3% (1.35% - 3.9%) | 1 (565) | .. | .. |
| Multi-drug resistant TB | 9.49% (7.13% - 12.14%) | 2 (543) | 4.40% - 10.84% | .. |
| **TB+HIV + Drug use** | **5.21% (2.73% - 8.39%)** | **4 (2,584)** | **2.74% - 5.87%** | **86.19%** |
| *By type of TB: all were TB (unspecified)* |  |  |  |  |
| **TB+HBV + Drug use** | **3.44% (0% - 12.05%)** | **3 (763)** | **0% - 9.78%** | **95.20%** |
| *By type of TB: all were TB (unspecified)* |  |  |  |  |
| **TB+Depression + Chronic lung disease** | **1.73% (0.65% - 3.24%)** | **2 (437)** | **1.15% - 2.86%** | **..** |
| *By type of TB: all were TB (unspecified)* |  |  |  |  |
| **TB+HBV + HCV** | **1.56% (0.60%-2.90%)** | **2 (500)** | **1.00%-2.00%** | **..** |
| *By type of TB: all were TB (unspecified)* |  |  |  |  |
| **TB+HIV + HCV** | **1.47% (0.36% - 3.21%)** | **6 (1,550)** | **0.33% - 4.98%** | **79.36%** |
| *By type of TB (p=0.45)* |  |  |  |  |
| TB (unspecified) | 1.31% (0.13% - 3.42%) | 5 (1,292) | 0.33% - 4.98% | 83.07% |
| Pulmonary TB | 2.33% (1.07% - 4.98%) | 1 (258) | .. | .. |
| **TB+DM + Drug use** | **1.03% (0.22% - 2.34%)** | **5 (7,476)** | **0.17% - 6.76%** | **90.85%** |
| *By type of TB (p=0.08)* |  |  |  |  |
| TB (unspecified) | 0.17% (0.06% - 0.31%) | 2 (5,452) | 0.18% - 2.21% | .. |
| Pulmonary TB | 1.4% (0.05% - 4.14%) | 3 (2,024) | 0.33% - 6.76% | .. |
| **TB+HIV + DM** | **0.98% (0.70% - 1.29%)** | **40 (526,827)** | **0% - 8.33%** | **97.79%** |
| *By type of TB (p=0.28)* |  |  |  |  |
| TB (unspecified) | 0.79% (0.53% - 1.1%) | 27 (520,674) | 0% - 8.33% | 98.26% |
| pulmonary TB | 2.48% (0.18% - 6.8%) | 7 (3,325) | 0% - 8.05% | 96.56% |
| extra-pulmonary TB | 0.16% (0% - 0.61%) | 2 (1,259) | 0.57% - 2.70% | .. |
| drug resistant TB | 1.35% (0.32% - 2.94%) | 3 (1,517) | 0.71% - 2.44% | .. |
| Multi-drug resistant TB | 1.92% (0.34% - 10.12%) | 1 (52) | .. | .. |
| **TB+DM + Alcoholism** | **0.69% (0.47% - 0.94%)** | **2 (5,218)** | **0.63% - 0.76%** | **..** |
| *By type of TB (p=0.98)* |  |  |  |  |
| TB (unspecified) | 0.76% (0.55% - 1.04%) | 1 (4,900) | .. | .. |
| Pulmonary TB | 0.63% (0.17% - 2.26%) | 1 (318) | .. | .. |
| **TB+DM + Chronic kidney disease** | **0.50% (0.20% - 0.89%)** | **2 (2,275)** | **0.55% - 12.22%** | **..** |
| *By type of TB (p<0.005)* |  |  |  |  |
| Drug resistant TB | 12.22% (6.96% - 20.57%) | 1 (90) | .. | .. |
| Pulmonary TB | 0.55% (0.31% - 0.96%) | 1 (2,185) | .. | .. |
| **TB+HIV + Chronic liver disease** | **0.48% (0.03% - 1.27%)** | **2 (682)** | **0.50% - 2.33%** | **..** |
| *By type of TB (p=0.11)* |  |  |  |  |
| TB (unspecified) | 0.5% (0.17% - 1.47%) | 1 (596) | .. | .. |
| Pulmonary TB | 2.33% (0.64% - 8.09%) | 1 (86) | .. | .. |
| **TB+DM + Chronic lung disease** | **0.23% (0%-0.98%)** | **3 (45,143)** | **0.03%-1.57%** | **96.08%** |
| *By type of TB (p=0.01)* |  |  |  |  |
| TB (unspecified) | 0.05% (0.03% - 0.08%) | 2 (45,016) | 0.03% - 0.45% | .. |
| Multi-drug resistant TB | 1.57% (0.43% - 5.56%) | 1 (127) | .. | .. |
| **TB+HIV + Chronic lung disease** | **0.10% (0% - 0.64%)** | **2 (682)** | **0.17% - 1.16%** | **..** |
| *By type of TB (p=0.16)* |  |  |  |  |
| TB (unspecified) | 0.17% (0.03% - 0.94%) | 1 (596) | .. | .. |
| Pulmonary TB | 1.16% (0.21% - 6.3%) | 1 (86) | .. | .. |
| **TB+DM + Heart disease** | **0.05% (0.02% - 0.07%)** | **2 (40,495)** | **0.07% - 2.28%** | **..** |
| *By type of TB (p<0.005)* |  |  |  |  |
| TB (unspecified) | 0.07% (0.05% - 0.1%) | 1 (39,881) | .. | .. |
| Pulmonary TB | 2.28% (1.36% - 3.79%) | 1 (614) | .. | .. |
| **TB+DM + Chronic liver disease** | **0% (0% - 0%)** | **2 (181,485)** | **0% - 0.03%** | **..** |
| *By type of TB: all were TB (unspecified)* |  |  |  |  |

*Note.* Subgroups by quality classification were only reported when significant differences were found. We were unable to do subgroup analyses by gender or age, given the lack of data reported separately for each gender or different age groups. See supplementary Figures 1-22 for subgroups by country. DM: Diabetes Mellitus; HBV: Hepatitis B Virus; HCV: Hepatitis C Virus; HIV: Human immunodeficiency virus; PTSD: Post-traumatic stress disorder; TB: Tuberculosis.
\* Pooled prevalence and 95% confidence intervals from random-effects meta-analysis when the number of studies is >1.
† I-squared values reported only for meta-analyses with >2 studies
‡ P-values from the test for heterogeneity between sub-groups

Subgroup analyses by country income classification found that higher income classifications tended to have higher prevalence of cTBMM. However, for most of the cTBMM clusters there were subgroups with only one study (Supplementary Table 6). Similarly, conducting subgroup analyses by countries resulted in multiple subgroups with only one (or very few) studies in each group (Supplementary Figures 1-22).

Quality scores ranged from one to seven stars (more stars indicating higher quality), with most studies receiving three or four (78%, Supplementary Table 7). In most of the meta-analyses, we found no significant differences, except for the clusters TB+HIV+DM, where changes were minimal, and TB+hepatitis B virus+drug use, where pooling studies with quality scores of three or more stars resulted in a significant increase in the estimated pooled prevalence (7.6%, 95%CI 5.5% -10.0%, 2 studies, Supplementary Table 6). Comparison of the pooled estimates of raw and adjusted data was not possible, as only raw (unadjusted) data were available.

Our post-hoc subgroup analyses to explore the impact of different definitions of depression, anxiety, alcoholism, drug use, chronic lung disease, and cardiovascular disease found that the prevalence of TB+HIV+depression varied significantly based on how depression was measured, ranging from 6% for studies using the Patient Health Questionnaire (PHQ-9)[32] to 13% in studies using the Hospital Anxiety and Depression Scale.[33] The definition of alcoholism also significantly affected the prevalence of HIV+alcoholism in people with TB, ranging from 5% when alcoholism was defined as ‘alcohol use that interfered with family, health, or work’, to 17% when defined as ‘alcohol dependence or addiction’. Finally, different definitions of drug use significantly affected the results for the prevalence of the clusters TB+HBV+drug use and TB+DM+drug use (Supplementary Table 8).

### Bias across studies (publication bias)

Only one meta-analysis included more than 10 studies (TB+HIV+DM prevalence). The visual inspection of the funnel plot showed an association between the effect sizes and their standard errors (no funnel shape), although this could be explained by the low pooled prevalence (<0.2%) and the impossibility of a study’s prevalence to go <0%. This was supported by the results of the trim-and-fill method, which imputed no additional studies (Supplementary Figure 23).

## DISCUSSION

To the best of our knowledge this is the first systematic review and meta-analysis of cTBMM and its associated burden. We identified multiple clusters of conditions that were present in ∼10-15% of TB patients, with HIV and mental health disorders being at least one of the conditions in these clusters. This emphasises the need for more global health research focused on mental health.[34,35] Furthermore, considering the synergistic relationship between these conditions and TB,[5,8] the high prevalence of cTBMM clusters including these conditions highlights the need for an integrated approach to care for cTBMM.

Since 2004, the World Health Organization (WHO) has had guidelines for delivering integrated TB and HIV comorbidity services.[36] A similar integration between TB programmes and mental health services has been called for by the WHO, but guidelines are yet to be published.[37] Despite the recent interest in TB and specific comorbidities, there has been limited policy focus on better integrated treatment for coexisting conditions with TB, leave alone for cTBMM, particularly in LMICs where there is a pressing need. Increased emphasis on people-centred care offers a valuable framework for addressing multimorbidities,[38,39] however, robust evaluations of such health system-wide interventions in LMICs are urgently needed. In fact, there is still little evidence on effective interventions to treat depression and anxiety disorders in TB.[40]

Our review found scarce data on treatment outcomes in people with cTBMM, which were mostly in the form of prevalence values, with little evidence regarding the risk of poorer outcomes for people with cTBMM compared to people with TB alone. However, we identified preliminary evidence that people with cTBMM have low treatment success rates and, compared to people with only TB, higher risks of death. This is in line with a study that found increased risks of adverse outcomes with increasing number of comorbidities.[13]

Given the wider international focus on identifying, preventing, and managing multimorbidity in other contexts, it is essential that people with cTBMM are not left behind. There is an important role for well conducted prospective cohort studies to identify common patterns of cTBMM and their impact on TB treatment outcomes; and for high quality clinical trials of interventions to prevent and treat multimorbidity in TB. Such research would be valuable in shaping services in LMICs, allowing earlier identification of TB patients at risk of co- and multimorbidities, and the development of better-integrated scalable models of care. Given the substantial burden and prevalence of TB and multimorbidity in LMICs — and the current gaps in our understanding — there is an urgent need for policy makers and funders to progress research in this area.

### Strengths and Limitations

To our knowledge, this is the first systematic review summarising the prevalence of the different clusters of cTBMM in LMICs. While the exclusion of studies with data from HICs might have contributed to the small number of studies with data on the different clusters, given the differences between HICs and LMICs in terms of the prevalence of chronic conditions, risk factors, and health services, we feel our focus on LMICs makes our results more representative of the populations with the highest burden of cTBMM.

Another strength of our study was our comprehensive search strategy without language restrictions and contacting authors of relevant abstracts.

Our review has several limitations that should be considered when interpreting the results. First, the full text of 62 articles could not be found despite our efforts. Including these studies may have increased the precision of some results.

Second, the I^2^ values were, for the most part, extremely high and remained unexplained by our subgroup analyses. However, this is not uncommon in the pooling of prevalence estimates in observational research. Nonetheless, pooled effect estimates should be interpreted with care. Despite the large heterogeneity present in the meta-analyses, the range of values remained relatively narrow, providing a useful benchmark for the prevalence of each cTBMM cluster.

Third, except for the case of TB+HIV+DM, very few studies contributed to results for each cTBMM cluster, with most being reported by only one study, which may limit the generalisability of those findings. Considering the high heterogeneity observed in those cTBMM clusters where a meta-analysis was possible, caution should be exercised when interpreting results where there was only one data point.

Fourth, definitions and measurement tools for some chronic conditions varied between studies, potentially contributing to the heterogeneity in results. This was particularly the case for depression and anxiety, where the same term could span a range of severities.

Finally, few of the included studies focused primarily on cTBMM, with most of the data in our review extracted from baseline characteristics tables or subgroup analyses. This limited our results to almost only prevalence data and findings regarding the burden of disease and treatment outcomes were sparse. However, this increases our confidence in the absence of publication bias in our meta-analyses, as the information regarding cTBMM is unlikely to have affected the publication status or reporting of a study.

## Conclusion

Further research is urgently needed for a more comprehensive picture of cTBMM for LMIC overall. Our systematic review highlights the importance of considering mental health conditions among people with TB, as the majority of cTBMM clusters present in ∼10-15% of TB patients had a mental health condition, often comorbid with HIV.

## Data Availability

All the collected data is in the public domain.

## Acknowledgements

We would like to thank Ol’ga Aryamkina, Monica Pontino, David Moore, Amer Hayat Khan, Larisa Kiryuhina, Huy Ngoc Le, Baty Florent, Jyotsna Joshi, Helder Bastos, and Saurabh Mehta, who kindly provided us with the pdf files for their studies. We are especially thankful to Barbara Reis-Santos and Olusola Adejumo, who provided raw and additional unpublished data from their studies.

This review is part of the TB Multimorbidity Network project (https://www.impactsouthasia.com/tbmm/), which is funded by the Medical Research Council (MRC Grant reference MC_PC_MR/T037806/1).

## Contributions

NS, BS, KS, and HE conceptualised the study; AJ, RM, AE, HE, KS, BS, and NS contributed to the design of the study; AJ, RM, OT, AL, AY, SA, TF, EMT, LR, LM, SR, SF, FNR, UE, and NS contributed to the titles and abstracts screening; AJ, RM, OT, AL, AY, SA, TF, EMT, LR, LM, SR, SF, and FNR contributed to the full text screening; AJ, RM, OT, AL, AY, SA, TF, EMT, LR, and LM contributed to data extraction and quality assessment; AJ summarised the results, did the analyses, and wrote the first draft of the report with input from BS and NS; all authors contributed to the interpretation of the results and revised the manuscript. All authors had full access to all the data in the study and had final responsibility for the decision to submit for publication.

## Data sharing

All the collected data is in the public domain.

