## Supplementary for "Prevalence, clusters, and burden of complex tuberculosis multimorbidity in low- and middle-income countries: a systematic review and meta-analysis"

### Supplementary Table 1: List of conditions considered or not chronic conditions for this review

| condition/risk factor | Should be considered as a comorbidity in this review? | Description or details |
|---|---|---|
| HIV | Yes | |
| Diabetes mellitus (DM) | Yes | |
| smoking | No | Would not fall under ‘substance abuse’, even if it is reported as ‘high dependence smoking’ |
| Drinking alcohol | No | No, unless it is something like 'harmful use of alcohol' or 'alcohol dependence' |
| Substance use / drug abuse | Yes | usually reserved for illicit substances |
| Cavitary disease | No | |
| Acute Respiratory Distress Syndrome | No | Acute |
| Leukopenia | no | |
| Chronic diarrhea | no | Unless it’s inflammatory bowel disorders, no. |
| Malnourishment | No | |
| Obesity | No | |
| BMI | No | |
| Anaemia | No | |
| drug-induced hepatotoxicity/ liver injury | No | |
| Intestinal parasites | No | |
| hypothyroidism | Yes | |
| hyperthyroidism | Yes | |
| hypokalaemia | No | |
| hearing defect | Yes | |
| Hypertension | No | |
| Dyslipidemia | No | |
| Skin conditions | no | |
| deep venous thrombosis | NO | |
| sacroiliitis | NO | |
| severe epistaxis | No | |
| seizures (cause not determined) | Yes if called epilepsy | Yes only if called epilepsy |
| haemorrhoids/fistula-in-ano | No | |
| dermatitis | No | |
| cerebrovascular accident | Yes | |
| scabies | No | |
| pneumothorax | No | |
| pancreatitis | No | Unless Long term pancreatitis and it is clearly specified |
| cryptococcal IRIS (immune reconstitution inflammatory syndrome) adenitis | No | |
| cryptoccocus | No | complications of HIV |
| visual impairment (reported as a symptom of DM) | No (reported as a complication) | |
| pulmonary fungal infection | No | complications of HIV |
| Candida coinfection | No | complications of HIV |

| | | |
|---|---|---|
| Aspergilius coinfection | No | complications of HIV |
| chronic airflow obstruction | Yes | As a proxy for COPD |
| HTLV (Human T-Cell LymphotropicVirus) | No | complications of HIV |
| pneumoconiosis | Yes | |
| hyponatremia | No | complications of HIV |
| Hepatitis B virus (HBV) | Yes | |
| Hepatitis C virus (HCV) | Yes | |
| Acquired immunodeficiency syndrome (AIDS) | Yes | |
| Autoimmune diseases | Yes | |
| Arthritis | Yes | |
| T. pallidum | Yes | |
| Cancer | Yes | Any type of cancer |
| Heart disease / Cardiopathies | Yes | |
| cardiomyopathy | No | Can be acute and/or reversible |
| Heart failure | Yes | |
| Cardiovascular disease | Yes | |
| cerebrovascular accident | Yes | Cerebrovascular disease is a form of cardiovascular disease |
| Chronic kidney disease / Chronic renal failure | Yes | No if acute renal failure- Yes if chronic renal failure |
| Chronic liver disease / Cirrhosis / Chronic hepatic dysfunction | Yes | |
| Depression | Yes | Clinical diagnostic or assessed with a validated scale (e.g. PHQ-9) |
| Anxiety | Yes | Clinical diagnostic of an anxiety disorder or assessed with a validated scale |
| Panic disorder | Yes | |
| Obsessive compulsive disorder (OCD) | Yes | |
| Post-traumatic stress disorder (PTSD) | Yes | |
| Mental disorder | Yes | Umbrella term for conditions such as PTSD, OCD, depression, anxiety disorders, etc. |
| Pneumonia | No | Acute condition |
| Chronic obstructive pulmonary disease (COPD) | Yes | COPD is form of chronic lung disease |
| Chronic lung disease | Yes | |
| Pulmonary edema | No | Symptom |
| Asthma | Yes | |
| Cor pulmonale | Yes | |
| Unstable angina | Yes | |
| Chronic corticosteroid therapy | No | Treatment |

### Supplementary Table 2: Characteristics of included studies reporting prevalence or risk of tuberculosis multimorbidity clusters and their burden.

| Lead Author, year | Country (period), World Bank income classification | Study design and quality * | Setting | Inclusion and exclusion criteria | Sample characteristics | Outcomes |
|---|---|---|---|---|---|---|
| Carvalho 2002 [S5] | Brazil (1995-1997), Upper Middle | Coh *** | Hospital setting | Included: All pulmonary TB patients with a positive acid-fast bacillus | Sample size: 86<br>Gender: Female: 31.4%; Male: 68.6%<br>Age: median (range): 37 years (18-76) | PTB +HIV +Alcoholism (CAGE alcohol screening test): 5/86 (5.81%)<br>PTB +HIV +DM: 4/86 (4.65%)<br>PTB +HIV +Heart disease: 4/86 (4.65%)<br>PTB +HIV +Cancer: 3/86 (3.49%)<br>PTB +HIV +Autoimmune diseases: 3/86 (3.49%)<br>PTB +HIV +Chronic liver disease: 2/86 (2.33%)<br>PTB +HIV +Chronic lung disease (COPD): 1/86 (1.16%) |
| Blal 2005 [S9] | Brazil (1999-2002), Upper Middle | CS *** | Tuberculosis Reference Center | Included: TB patients who had been serologically tested for HBV infection | Sample size: 209<br>Age: mean (SD): 39.7 (11.6) | TB +HIV +HBV: 43/209 (20.57%) |
| Krupitsky 2006 [S78] | Russia (2000-2001), Lower Middle | Coh **** | TB hospital | Included: TB patients treated in the city TB hospital | Sample size: 160<br>Gender: Female: 31.9%; Male: 68.1%<br>Age: mean (SD): 39.8 (1.0) | TB +Drug use (drug abuse) +Alcoholism (DSM-IV criteria): 16/160 (10%) |
| Richards 2006 [S58] | Georgia (2001), Low | CS ** | In-patient TB hospitals | Included: Hospital in-patients aged ≥ 18 years. | Sample Size: 272<br>Gender: Female: 27.6%; Male: 72.4%<br>Age: median (range): 35 (18-74)<br>Education: completed primary education: 93%; completed high school: 16% | TB +HIV +HCV: 1/272 (0.37%) |
| Kuniholm 2008 [S59] | Georgia (1997-1998), Lower Middle | CS ** | In-patient tuberculosis hospitals | Included: Hospital patients aged 18 to 65 years and who consented to answer a confidential questionnaire and be tested for HIV, HCV, and HBV. | Sample size: 300<br>Gender: Female: 29.7%; Male: 80.7%<br>Age: 18-27: 31.7%; 28-37: 42.0%; ≥38: 26.3%<br>Education: Primary: 12.7%; Secondary: 57.7%; University: 29.7% | TB +HIV +HCV: 1/300 (0.33%)<br>TB +HBV +HCV: 6/300 (2%) |
| Pando 2008 [S30] | Argentina (2001), Upper Middle | CS *** | 3 public hospitals in Buenos Aires | Included: Patients with TB aged ≥18 years who attended one of three public hospitals in Buenos Aires | Sample size: 181 (of 205) with full serological data<br>Gender: Female: 34.1%: Male: 65.9%<br>Age: mean (SD): 34.8 (14.1); 18-24: 27%; 25-29: 20%; 30-44: 29%; ≥45: 23%<br>Education: High educational level (high school equivalent): 59%<br>Occupation: Employees: 80.5% | TB +HIV +HBV: 4/181 (2.21%)<br>TB +HIV +HCV: 3/181 (1.66%)<br>TB +HIV +HBV+HCV: 8/181 (4.42%)<br>TB +HIV +Drug use (use of illicit drugs): 17/205 (8.29%)<br>TB +HBV +Drug use (use of illicit drugs): 9/205 (4.39%)<br>TB +HCV +Drug use (use of illicit drugs): 9/205 (4.39%)<br>TB +Drug use (use of illicit drugs) +T. pallidum: 1/205 (0.49%) |
| Sirinak 2008 [S81] | Thailand (2005-2006), Lower Middle | Coh ***(*)† | Public TB treatment facilities and national infectious diseases disease referral hospital. | Included: Adults aged ≥18 years with documented HIV infection and active TB disease who had received anti-TB therapy for <4 weeks<br>Excluded: Prisoners and pregnant women. | Sample size: 769<br>Gender: Female: 30%; Male: 70%<br>Age: median (IQR): 34 (30-41)<br>Education: >6th grade: 39%<br>Occupation: Employed: 59%<br>Types of TB: PTB: 60%; EPTB: 30%; PTB+EPTB: 10% | TB +HIV +HCV:<br>Cured: 63 (30.00%) of 210 with TB+HIV+HCV<br>Treatment completed: 65/210 (30.95%)<br>Treatment failure: 2/210 (0.95%)<br>Death: 37/210 (17.62%)<br>Defaulted: (20/210 (9.52%)<br>TB +HIV +HBV:<br>Cured: 7 (16.28%) of 43 with TB+HIV+HBV<br>Treatment completed: 16/43 (37.21%) |

| | | | | | | |
|---|---|---|---|---|---|---|
| | | | | | | Treatment failure: 1/43 (2.33%)<br>Death: 5/43 (11.63%)<br>Defaulted: 8/43 (18.60%)<br>TB +HIV +HBV+HCV:<br>Cured: 6 (22.22%) of 27 with TB+HIV+HBV+HCV<br>Treatment completed: 9/27 (33.33%)<br>Treatment failure: 0/27 (0%)<br>Death: 7/27 (25.93%)<br>Defaulted: 1/27 (3.70%) |
| Deribew 2009 [S49] | Ethiopia (February to April 2009), Low | CS **** | 3 specialized hospitals | Included: HIV patients with new TB and who were in the intensive phase of anti-TB treatment.<br>Exclusion: Patients aged <15 years, the presence of an opportunistic infection or a known chronic illness like DM and hypertension. | Sample size: HIV+TB: 124<br>Gender: Female: 50%; Male 50%<br>Age: 15-24: 9.7%; 25-34: 42.7%; ≥ 35: 47.6%<br>Education: Illiterate: 24.2%; Literate: 75.8%<br>Occupation: Government employee: 13.7%; Private employee: 12.1%; Merchant: 8.1%; Farmer: 10.5%; Housewives: 12.1%; Daily laborer: 17.7%; No Job: 25.8%<br>Types of TB: PTB: 83%; EPTB: 17% | TB +HIV +Depression (Kessler 10 scale):<br>Poor physical health (below the median in the physical domain of the WHOQOL-HIV): 12 (63.2%) of 19 with TB +HIV +Depression and poor physical health data<br>Poor psychological health (below the mean in the psychological domain of the WHOQOL-HIV): 59 (74.7%) of 79 with TB +HIV +Depression and poor psychological health |
| Pepper 2010 [S12] | South Africa (2008-2009), Upper Middle | Coh * | Tuberculosis clinic (a primary-level outpatient health care facility) | Included: Patients ≥ 18 years diagnosed with TB whose HIV-1 status and clinic records were available. | Sample size: 292<br>Gender: Female: 46.9%; Male: 53.1%<br>Age: Median (IQR) in HIV-1 infected: 35 (30-41); in HIV-1 uninfected: 38 (29-49)<br>Types of TB: PTB: 67.1%; EPTB: 31.8% | TB +HIV +Cardiovascular disease (Cerebrovascular accident): 3/292 (1.03%) |
| Gupta 2011 [S37] | India (2005-2006), Low | CS *** | Tertiary care hospital | Included: PTB and EPTB patients attending hospital | Sample size: 229<br>Gender: Female: 51; Male: 178<br>Age: <20: 8.7%; 21-30: 25.8%; 31-40: 18.3%; 41-50: 20.1%, 51-60: 17%; >60: 10.0%<br>Occupation: Unemployed: 9.2%; Housewives or students: 24.5%; Labourers and workers: 66.4%<br>Types of TB: PTB: 192; EPTB: 37 | EPTB +HIV +DM: 1/37 (2.7%) |
| Heredia 2011 [S17] | Mexico (2008-2009), Upper Middle | CS **** | A specific region of Guerrero State health jurisdiction # 2 (North Zone) | Included: Patients with PTB | Sample size: 107<br>Gender: Female: 44.9%; Male 55.1%<br>Age: mean (SD): 48.3 (17.2)<br>Types of TB: all PTB | PTB +HIV +DM: 0/107 (0%)<br>TB +DM +Heart failure: 0/107 (0%)<br>TB +DM +Chronic liver disease: 0/107 (0%)<br>TB +DM +Unstable angina: 0/107 (0%) |
| Reis 2011 [S31] | Brazil (2008-2010), Upper Middle | CS ***** | Out-patient and in-patient units at reference hospital for infectious diseases. | Included: Patients with clinical diagnosis of TB. | Sample size: 402<br>Gender: Female: 28.1%; Male: 71.9%<br>Age: mean (range): 44.1 (3-86); <30: 18.7%; 30-39: 24.4%; 40-49: 23.1%; ≥50: 33.8%<br>Education: Nearly 80.0% had received ≤8 years of formal education<br>Monthly income: ≤US$750: 95.1%; ≤1 Brazilian minimum wage/month (~US$300): 63.8% | TB +HIV +HCV: 20/402 (4.98%)<br>TB +HIV +Drug use (intravenous drug use): 11/402 (2.74%) |
| Manosuthi 2012 [S80] | Thailand (2008), Lower Middle | Coh *** | 6 hospitals in Bangkok and Nonthaburi, Thailand | Included: TB-infected patients age > 15 years. | Sample size: 813<br>Gender: Female: 37.9%; Male: 62%<br>Age: mean (SD): 41 (14)<br>Types of TB: PTB: 65.6%; EPTB: 22.6%; PTB & EPTB: 11.8% | TB +HIV +DM: 7/596 (1.17%)<br>TB +HIV + Chronic lung disease: 1/596 (0.17%)<br>TB +HIV +Chronic kidney disease: 1/596 (0.17%)<br>TB +HIV +Chronic liver disease: 3/596 (0.5%) |

| | | | | | | |
|---|---|---|---|---|---|---|
| Peltzer 2012 [S10] | South Africa (2011), Upper Middle | CS *** | Public primary care clinics in 3 provinces in South Africa. | Included: New TB treatment or retreatment patient (within one month on treatment) and ≥ 18 years | Sample size: 4900<br>Gender: Female: 45.5%; Male: 54.5%<br>Age: mean (SD): 36.2 (11.5), range: 18-93 years; 18-34: 36.6%; 35-44: 41.9%; ≥45: 21.5%<br>Education: grade 7 or less: 26.3%; grade 8-11: 45.9%; grade 12 or more: 27.7%<br>Poverty index: low: 35%; medium: 48.2%; high: 16.9% | TB +HIV +Alcoholism (alcohol use disorders identification test (AUDIT)): 552/4900 (11.27%)<br>TB +DM +Alcoholism (alcohol use disorders identification test (AUDIT)): 37/4900 (0.76%) |
| Jimenez-Corona 2013 [S11] | Mexico (1995-2010), Upper Middle | Coh *** | 12 municipalities in the Orizaba Health Jurisdiction in Veracruz State. | Included: Consecutive patients aged ≥ 15 years with acid-fast bacilli or Mycobacterium tuberculosis in sputum samples. | Sample size: 1262<br>Gender: Female: 42.16%; Male: 57.84%<br>Age: 45.32 (18.18)<br>Education: >6 years: 67.14% | PTB +HIV +DM: 2/1262 (0.16%)<br>PTB +DM +Drug use (use of illicit drugs): 6/1262 (0.48%) |
| Abdelsalam 2013 [S66] | Sudan (2010-2011), Lower Middle | CS ** | Tuberculosis center at Tropical Diseases Teaching Hospital | Included: Confirmed TB patients on antituberculous treatment and managed in TB center. | Sample size: 200<br>Gender: Female: 36.5%; Male: 63.5%<br>Age: <40: 70%; ≥40: 30%<br>Education: Educated: 67.5%, Illilterate: 32.5%<br>Occupation: Employed: 57.5%; Unemployed: 42.5% | TB +HBV +HCV: 2/200 (1%)<br>TB +HBV +Drug use (intravenous drug use): 0/200 (0%) |
| Peltzer 2013 [S16] | South Africa (2011), Upper Middle | CS *** | 42 public primary care clinics | Included: Patients aged ≥18 years with a new TB treatment or retreatment. | Sample size: 4900<br>Gender: Female: 45.5%; Male: 54.5%<br>Age: mean (SD) 36.2 (11.5); 18-24: 13.4%; 25-34: 38.1%; 35-44: 27.1%; ≥45: 21.5%<br>Poverty index: Low: 35.0 %; Medium: 48.2 %; High: 16.9 %<br>Education: Grade 7 or less: 26.3%; Grade 8-11: 45.9%; Grade 12 or more: 27.7% | TB +HIV +PTSD: 815/4900 (16.63%)<br>TB +PTSD +Alcoholism (alcohol use disorders identification test (AUDIT)): 346/4900 (7.06%) |
| Alavi 2012 [S18] | Iran (2008-2010), Upper Middle | CC ***** | Teaching hospital | Included: Patients aged ≥15 years diagnosed with TB. | Sample size: Patients with TB: 148; All patients: 2124<br>Gender:<br>Patients with TB: Female: 46%; Male: 54%<br>All patients: Female: 45%; Male: 55% | PTB +DM +Drug use (use of illicit drugs): 10/148 (6.76%); RR 66.76 (14.76 - 301.88)<br>PTB +HIV +DM: 9/148 (6.08%); RR 120.16 (15.33 - 942.05) |
| Aires 2012 [S19] | Brazil (2008-2010), Upper Middle | CS ***** | Out-patient and in-patient units at the a reference hospital for infectious diseases | Included: Patients diagnosed with TB.<br>Excluded: Children. | Sample size: 402<br>Gender: Female: 28.1%; Male: 71.9%<br>Age: <30 years: 18.7%; 30-39: 24.4%; 40-49: 23.1%; ≥50: 33.8%<br>Monthly income: ≤1 minimum salary (US$300): 63.8%; >1 minimum salary: 31.3% | TB +HIV +HBV: 41/402 (10.2%)<br>TB +HBV +Drug use (use of illicit drugs): 35/358 (9.78%) |
| Faurholt-Jepsen 2012 [S79] | Tanzania (2006-2008), Low | Coh *** | Urban setting | Included: Patients newly diagnosed with active PTB.<br>Excluded: Patients aged <15 years, pregnant or lactating women, or who were terminally ill. | Sample size: 1205<br>Gender: Female: 40.9%; Male: 59.1%<br>Age: mean (SD): 36.6 (13.0)<br>Occupation: (TB with Diabetes) Farmer/Fisherman: 38.3%; Businessman/Employed: 37.8%; Housewife: 11.7%; Unemployed: 4.6%<br>Types of TB: PTB: 100% | PTB +HIV +DM: 97/1205 (8.05%) |
| Modongo 2012 [S28] | Botswana (2007-2012), Upper Middle | CS **** | Botswana (no further details reported) | Included: all MDR-TB patients in Botswana since January 2007 | Sample size: 452 patients included (213 had TSH level checked)<br>Gender: Female: 44.9%; Male: 55.1%<br>Age: mean (range): 37 (28-48) | MDR-TB +HIV +Hypothyroidism: 49/452 (10.84%) |

| | | | | | | |
|---|---|---|---|---|---|---|
| Naik 2013 [S38] | India (2012), Lower Middle | CS **** | Peripheral health institutions of Bangarpet (population 0.5 million) | Included: All TB patients registered from January to September 2012 | Sample size: 362<br>Gender: Female: 32%; Male: 68%<br>Age: median (IQR): 40 (27-56); <40: 47.8%; ≥ 40: 52.2%<br>Types of TB: new smear-positive PTB: 44.8%; New smear-negative PTB: 12.4%; New EPTB: 26.2%; Retreatment: 16.6% | TB +HIV +DM: 1/358 (0.28%) |
| Khanna 2013 [S39] | India (2012), Lower Middle | CS **** | Chest clinic at a tertiary care teaching hospital | Included: All adult TB patients | Sample size: 458<br>Gender: Female (51%); Male: 49%<br>Age: <40: 71.4%; ≥40: 28.6%<br>Types of TB: PTB: 62.4%; EPTB: 37.6% | TB +HIV +DM: 0/458 (0%) |
| van den Heuvel 2013 [S70] | Zambia (2009-2010), Low | CS **** | Primary health care centres (urban and rural) | Included: Patients with confirmed HIV and TB disease status who were started on ART or TB treatment within one month<br>Excluded: Patient's age < 16 years | Sample size: 500 TB patients<br>Gender: Female: 37.4%, Male: 62.6%<br>Age: mean (IQR) for TB+HIV coinfected: 33.9 (30-40); for TB only: 31.0 (26-39)<br>Education: No education: 4.6%; Primary: 30.4%; Secondary: 63.2%; Tertiary: 9.2%<br>Occupation: Employed: 22% | TB +HIV +Depression (MINI): 34/500 (6.8%)<br>TB +HIV +Anxiety (any anxiety disorder, MINI): 82/500 (16.4%)<br>TB +HIV +Anxiety (Generalised Anxiety Disorder, MINI): 21/500 (4.2%)<br>TB +HIV +Obsessive compulsive disorder: 11/500 (2.2%)<br>TB +HIV +Panic disorder: 11/500 (2.2%)<br>TB +HIV +PTSD: 12/500 (2.4%) |
| de Oliveira 2013 [S15] | Brazil (2010), Upper Middle | CS *** | Several referral hospitals | Included: Admitted patients with TB as a primary or associated diagnosis. | Sample size: 278<br>Gender: Female: 42.1%; Male: 57.9%<br>Age: Mean (SD): 38.0 (18.0); 0-19: 10.1%; 20-29: 24.1%; 30-39: 22.3%; 40-49: 19.4%; 50+: 24.1%<br>Education (years of schooling): ≤4: 41.4%; ≥5: 58.6%<br>Income (minimum wage): <1: 25.9%; 1-3: 60.4%; >3: 13.7% | TB +HIV +Alcoholism (alcohol dependence/addiction): 46/278 (16.55%)<br>TB +HIV +Drug use (use of illicit drugs): 24/278 (8.63%) |
| Achanta 2013 [S41] | India (2012), Lower Middle | CS ***** | 19 peripheral health institutions | Included: all TB patients registered between January to September 2012 | Sample size: 374<br>Gender: Female: 36.9%; Male: 63.1%<br>Age:< 40: 50.5%; ≥40: 49.5% | TB +HIV +DM: 0/381 (0%) |
| Kibirige 2013 [S68] | Uganda (2011-2012), Low | CS **** | Pulmonology wards of Mulago hospital | Included: Patients with confirmed TB aged ≥18 years and admitted on the pulmonology wards.<br>Excluded: Patients on anti TB drugs. | Sample size: 260<br>Gender: Female: 43.8%; Male: 56.2%<br>Age: 18-35: 59.2%; 36-50: 35%; >50: 5.8%<br>Education: No formal education: 6.9%; Primary level: 64.6%; Secondary level: 20.8%; Tertiary level: 7.7%<br>Occupation: Unemployed: 22.3%; Self employed: 13.1%; Unskilled: 56.5%; Skilled: 8.1%<br>Types of TB: PTB: 75.8%; EPTB: 23.2% | TB +HIV +DM: 13/260 (5%) |
| Reis- Santos 2013 [S21] | Brazil (2011), Upper Middle | CS **** | National Notification System (SINAN) | Included: TB cases registered in the National Notification System (SINAN) in 2011<br>Excluded: Those subjects with missing information on TB treatment outcome. | Sample size: 39,881<br>Gender: Female: 31.7%; Male: 67.2%<br>Age: < 20: 8.5%; 20-39: 45.7%; 40-59: 33.2%; >60: 14.0%<br>Education: illiterate: 9%; 1-4 years: 33%; 5-8 years: 30%; >8 years: 28%<br>Types of TB: PTB: 83.3%; EPTB: 12.5%; PTB+EPTB: 3.1% | In addition to the outcomes below, risk ratios of treatment outcomes in each cluster compared to people with TB but without additional chronic conditions could be calculated and are reported in the Supplementary Table 4.<br>TB +AIDS +Chronic lung disease (COPD): 2/39881 (0.01%)<br>Died (any cause): 2 / 2 (100%); Died (other): 2 / 2 (100%);<br>TB +AIDS +Cancer: 17/39881 (0.04%)<br>Died (any cause): 10 / 17 (58.8%); Died (TB): 2 / 17 (11.8%); Died (other): 8 / 17 (47.1%); LFU: 2 / 17 (11.8%); Cured: 5 / 17 (29.4%);<br>TB +AIDS +DM: 110/39881 (0.28%) |

| | | | | | | |
|---|---|---|---|---|---|---|
| | | | | | | Died (any cause): 48 / 110 (43.6%); Died (TB): 10 / 110 (9.1%); Died (other): 38 / 110 (34.5%); LFU: 16 / 110 (14.5%); Cured: 44 / 110 (40%);<br>TB +AIDS +Heart disease: 8/39881 (0.02%)<br>Died (any cause): 4 / 8 (50%); Died (TB): 1 / 8 (12.5%); Died (other): 3 / 8 (37.5%); LFU: 2 / 8 (25%); Cured: 2 / 8 (25%);<br>TB +AIDS +Mental disorder: 143/39881 (0.36%)<br>Died (any cause): 53 / 143 (37.1%); Died (TB): 13 / 143 (9.1%); Died (other): 40 / 143 (28%); LFU: 45 / 143 (31.5%); Cured: 44 / 143 (30.8%);<br>TB +Arthritis +Cancer: 2/39881 (0.01%)<br>Cured: 2 / 2 (100%);<br>TB +Cancer +Chronic lung disease (COPD): 6/39881 (0.02%)<br>Died (any cause): 4 / 6 (66.7%); Died (TB): 1 / 6 (16.7%); Died (other): 3 / 6 (50%); Cured: 2 / 6 (33.3%);<br>TB +Cancer +Heart disease: 3/39881 (0.01%)<br>Died (any cause): 2 / 3 (66.7%); Died (other): 2 / 3 (66.7%); Cured: 1 / 3 (33.3%);<br>TB +DM +Arthritis: 4/39881 (0.01%)<br>Died (any cause): 1 / 4 (25%); Died (other): 1 / 4 (25%); LFU: 1 / 4 (25%); Cured: 2 / 4 (50%);<br>TB +DM +Chronic lung disease (COPD): 13/39881 (0.03%)<br>Died (any cause): 9 / 13 (69.2%); Died (TB): 4 / 13 (30.8%); Died (other): 5 / 13 (38.5%); LFU: 1 / 13 (7.7%); Cured: 3 / 13 (23.1%);<br>TB +DM +Cancer: 15/39881 (0.04%)<br>Died (any cause): 11 / 15 (73.3%); Died (TB): 1 / 15 (6.7%); Died (other): 10 / 15 (66.7%); Cured: 4 / 15 (26.7%);<br>TB +DM +Heart disease: 28/39881 (0.07%)<br>Died (any cause): 8 / 28 (28.6%); Died (TB): 3 / 28 (10.7%); Died (other): 5 / 28 (17.9%); Cured: 20 / 28 (71.4%);<br>TB +DM +Mental disorder: 118/39881 (0.3%)<br>Died (any cause): 21 / 118 (17.8%); Died (TB): 9 / 118 (7.6%); Died (other): 12 / 118 (10.2%); LFU: 18 / 118 (15.3%); Cured: 78 / 118 (66.1%);<br>TB +Heart disease +Chronic lung disease (COPD): 8/39881 (0.02%)<br>Died (any cause): 4 / 8 (50%); Died (TB): 1 / 8 (12.5%); Died (other): 3 / 8 (37.5%); Cured: 4 / 8 (50%);<br>TB +Mental disorder +Arthritis: 2/39881 (0.01%)<br>LFU: 1 / 2 (50%); Cured: 1 / 2 (50%);<br>TB +Mental disorder +Chronic lung disease (COPD): 5/39881 (0.01%)<br>Died (any cause): 3 / 5 (60%); Died (TB): 1 / 5 (20%); Died (other): 2 / 5 (40%); Cured: 2 / 5 (40%);<br>TB +Mental disorder +Cancer: 6/39881 (0.02%)<br>Died (any cause): 4 / 6 (66.7%); Died (other): 4 / 6 (66.7%); Cured: 2 / 6 (33.3%);<br>TB +Mental disorder +Heart disease: 2/39881 (0.01%)<br>Cured: 2 / 2 (100%);<br>TB +AIDS +DM +Chronic lung disease (COPD): 1/39881 (0%)<br>Died (any cause): 1 / 1 (100%); Died (other): 1 / 1 (100%);<br>TB +AIDS +DM +Mental disorder: 23/39881 (0.06%) |

| | | | | | | |
|---|---|---|---|---|---|---|
| | | | | | | Died (any cause): 6 / 23 (26.1%); Died (TB): 3 / 23 (13%); Died (other): 3 / 23 (13%); LFU: 5 / 23 (21.7%); Cured: 11 / 23 (47.8%);<br>TB +AIDS +Mental disorder +Cancer: 2/39881 (0.01%)<br>Died (any cause): 2 / 2 (100%); Died (other): 2 / 2 (100%);<br>TB +DM +Cancer +Heart disease: 1/39881 (0%)<br>Died (any cause): 1 / 1 (100%); Died (other): 1 / 1 (100%);<br>TB +DM +Heart disease +Chronic lung disease (COPD): 2/39881 (0.01%)<br>Died (any cause): 1 / 2 (50%); Died (other): 1 / 2 (50%); Cured: 1 / 2 (50%);<br>TB +DM +Mental disorder +Arthritis: 1/39881 (0%)<br>LFU: 1 / 1 (100%);<br>TB +DM +Mental disorder +Chronic lung disease (COPD): 3/39881 (0.01%)<br>Died (any cause): 3 / 3 (100%); Died (TB): 1 / 3 (33.3%); Died (other): 2 / 3 (66.7%);<br>TB +DM +Mental disorder +Cancer: 1/39881 (0%)<br>Died (any cause): 1 / 1 (100%); Died (other): 1 / 1 (100%);<br>TB +DM +Mental disorder +Heart disease: 1/39881 (0%)<br>Cured: 1 / 1 (100%); |
| Dave 2013 [S43] | India (2012), Lower Middle | CS ***** | Tuberculosis unit in the Anand district. | Included: All TB patients (adults and children) registered from January to September 2012. | Sample size: 556<br>Gender: Female: 33.3%; Male: 66.7%<br>Age: median (IQR): 35 (25-50); <35: 42.4%; ≥35: 57.6%<br>Types of TB: PTB: 86.7%; EPTB: 13.3% | TB +HIV +DM: 1/539 (0.19%) |
| KV 2013 [S47] | India (2010-2011), Lower Middle | Coh ***** | Revised National Tuberculosis Control Programme | Included: All TB cases aged ≥14 years.<br>Excluded: Transfer- in cases. | Sample size: 3116<br>Gender: Female: 32%; Male: 68%<br>Age: mean (SD): 46 (17); 15-44: 42.8%; ≥45: 57.2%;<br>Types of TB: PTB: 72%; EPTB: 28% | TB +HIV +DM: 5/3116 (0.16%) |
| Adem 2014 [S53] | Ethiopia (2010), Low | CS *** | University specialized Hospital and TB follow up clinic in health center. | Included: Patients aged ≥ 12 years | Sample size: 222<br>Gender: Female: 43.24%; Male: 56.76%<br>Age: 12-18: 4.50%; 19-40: 66.67%; 41-65: 24.33%; >65: 4.50%<br>Education: Illiterate: 11.7%; Literate: 8.11%; Primary: 36.04%; Secondary: 26.58%; Tertiary: 17.57%<br>Occupation: Government employee: 29.28%; Merchant: 10.81%; Daily laborer: 13.96%; Student: 9.46%; Farmer: 8.11%; Other: 28.38%<br>Types of TB: PTB: 82.4%; EPTB: 17.6% | TB +HIV +Depression (Kessler 10 scale): 28/222 (12.61%) |
| do Prado 2014 [S33] | Brazil (2007-2011), Upper Middle | CS **** | Database of the national TB reporting system (SINAN/TB). | Included: TB cases aged ≥ 15 years.<br>Excluded: missing data for HIV status. | Sample size: 243,672<br>Gender: Female: 67.7%; Male: 32.3%<br>Age: < 20: 5.2%; 20-39: 49.7%; 40-59: 34.9%; ≥60: 10.2%<br>Education: <4 years: 41.6%; 4-7 years: 26.0%; >8 years: 32.4%<br>Types of TB: PTB: 80.3%; EPTB: 15.2%; PTB & EPTB: 4.4% | TB +HIV +DM: 1095/243676 (0.45%) |

| | | | | | | |
|---|---|---|---|---|---|---|
| Damtew 2014 [S57] | Ethiopia (2014), Low | CS **** | Referral TB hospital | Included: patients with active PTB aged ≥ 15 years | Sample size: 120<br>Gender: Female: 44.2%; Male: 55.8%<br>Age: mean (SD): 37.3 (13.3); range: 15 - 86; ≤24: 11.7%; 25-44: 61.7%; 45-64: 20.8%; ≥65: 5.8%<br>Education: no formal education: 30.8%; grade 1-6: 23.3%; grade 7-12: 35.8%; diploma and above: 10%<br>Occupation: self-employed: 71.7%; employed: 20%; student: 5.8%; unemployed: 2.5% | TB +HIV +DM: 10/120 (8.33%) |
| Getachew 2014 [S54] | Ethiopia (2011-2012), Low | CS **** | Referral university hospital with DOTS follow-up clinic. | Included: All consecutive PTB patients aged ≥ 15 years. | Sample size: 199<br>Gender: Female: 41.2%; Male: 58.8%<br>Age: mean (SD): 33.9 (13.9); range: 14 - 80; ≤24: 30.7%; 25-44: 47.7%; 45-64: 16.6%; ≥65: 5.03%<br>Education: no formal schooling: 51.8%; grade 1-6: 5.7%; grade 7-12: 21.2%; diploma and above: 21.2%<br>Occupation: government employed: 7.5%; self-employed: 43.7%; student: 22.1%; homemaker: 21.1%; retired: 0.5%; unemployed: 5.1% | PTB +HIV +DM: 10/199 (5.03%)<br>Female: 3.66%; Male: 5.99% |
| Viswanathan 2014 [S45] | India (2011), Lower Middle | CS ** | 5 TB units | Included: New smear-positive TB patients who underwent DM screening.<br>Excluded: TB patients with pre-diabetes were excluded. | Sample size: 245<br>Gender: Female: 25.7%; Male: 74.3%<br>Age: ≥40: 58% | TB +HIV +DM: 9/245 (3.67%) |
| Tabarsi 2014 [S36] | Iran (2012-2013), Upper Middle | Coh **** | The National Research Institution of Tuberculosis and Lung Diseases, Masih Daneshvari Hospital | Included: All newly diagnosed TB patients aged ≥ 15 years who were hospitalized and who had HbA1c measurements at the time of registration and 3 months later. | Sample size: 317<br>Gender: Female: 48.6%; Male: 51.4%<br>Age: mean (SD): 55.33 (21.04); median (IQR): 58 (35.5 - 73.5)<br>Types of TB: PTB: 93.4% | TB +HIV +DM: 0/317 (0%)<br>TB +DM +Drug use (opium use): 7/317 (2.21%) |
| Kelly 2016 [S4] | South Africa (2014), Upper Middle | CS **** | Community-based setting | Included: Age ≥ 18 years receiving community-based treatment for MDR-TB, currently in the intensive phase of treatment and who had completed at least one month of treatment.<br>Excluded: XDR-TB | Sample size: 121<br>Gender: Female: 51.2%; Male: 48.8%<br>Age: Mean (SD): 33.1 (8.8); <24: 19.8%; 25-34: 37.2%; 35-44: 33.1%; 45+: 9.9%<br>Education: Less than secondary: 6.6%; Completed some secondary: 93.4%<br>Occupation: Unemployed: 30.6%; Employed: 69.4%<br>Types of TB: 100% MDR-TB | MDR-TB +HIV +Hypothyroidism: 4/91 (4.4%) |
| Oni 2015 [S22] | South Africa (Sep 2012-May 2013), Upper Middle | CS **** | A primary care health facility | Included: Persons identified from the electronic pharmacy and CDU databases as having been prescribed medicines for at least one of the four most prevalent chronic diseases (HIV, TB, DM, Hypertension). | Sample size: 14364 (388 patients had TB)<br>Gender and age not reported for TB patients | TB +HIV +DM: 6/14364 (0.04%) |
| Duko 2015 [S50] | Ethiopia (2014), Low | CS *** | Sodo University Hospital and Sodo Health Center | Included: Patients aged ≥18 years with TB<br>Excluded: Critically ill patients | Sample size: 417<br>Gender: Female: 42.2%; Male: 57.8%<br>Age: Mean (SD): 34.52 11.01); 8-24: 21.1%; 25-49: 69.8%; > = 50: 9.1% | TB +HIV +Depression (HADS depression subscale): 34/417 (8.15%)<br>TB +HIV +Anxiety (HADS anxiety subscale): 42/417 (10.07%) |

| | | | | | | |
|---|---|---|---|---|---|---|
| | | | | | Monthly income: <735 ETB (Ethiopian birrs): 50.8%; 735-1176 ETB: 20.6%; ≥ 1176 ETB: 28.6%<br>Education: No formal education: 18.7%; Primary education: 30.9%; Secondary education: 28.1%; Higher education: 22.3%<br>Types of TB: PTB: 68.8%; EPTB: 31.2% | |
| Delgado-Sánchez 2015 [S34] | Mexico (2000-2012), Upper Middle | CS ***** | National Tuberculosis Registry | Included: PTB patients aged ≥ 20 years<br>Excluded: Patients without information of previous DM diagnosis | Sample size: 181,378<br>Gender: Female: 36.49%; Male: 63.5%<br>Age: median (IQR): 43 (32-60)<br>Types of TB: DR-TB: 8.44%; MDR-TB: 43.39% | TB +DM +Chronic liver disease: 49/181378 (0.03%) |
| Magee 2015 [S73] | Georgia (2011-2014), Lower Middle | Coh **** | The National Center for TB and Lung Disease (largest anti-tuberculosis treatment and referral facility) | Included: Newly diagnosed patients aged ≥ 35 years with confirmed PTB and with no history of previous anti-tuberculosis treatment. | Sample size: 318<br>Gender: Female: 24.8%; Male: 75.2%<br>Age: median (IQR): 49 (42-58)<br>Monthly income (USD): median (IQR): 132 (47-412)<br>Types of TB: MDR-TB: 15.4%, XDR-TB: 0.9% | PTB +HIV +DM: 1/318 (0.31%)<br>PTB +DM +Alcoholism (≥ 5 drinks per day, ≥3 days/week): 2/318 (0.63%) |
| Workneh 2016 [S55] | Ethiopia (2013-2014), Low | CS ***** | DOTS clinics at selected health facilities | Included: New TB patients and DM patients newly diagnosed for TB<br>Excluded: Age < 15 years, re-treatment cases, known or suspected MDR-TB cases | Sample size: 1314<br>Gender: Female: 47.3%; Male: 52.7%<br>Age: mean 35.74 (SD 15.26); 15-40 years: 70.6%; 41-64 years: 21.7%; 65-89 years: 7.7%<br>62.9% from urban area; 37.1% from rural area<br>Education: no formal schooling: 47.0%; 1-6 grade: 19.2%; 7-12+1: 27.5%; Diploma and above: 6.2%<br>Occupation: Unemployed: 28.6%; Student 8.5%; Self-employed: 55.2%; Government employed: 7.7%<br>Monthly income (USD): No income: 12.7%; ≤18.9: 38.5%; 19-37.9 23.1%; ≥ 19-37.9: 23.1%<br>Types of TB: PTB: 57.6%; EPTB: 41.4% | TB +HIV +DM: 22/1314 (1.67%) |
| Weimann 2016 [S6] | South Africa (2008 (wave 1) and 2012 (wave 3)), Upper Middle | CS *** | Households in South Africa | Included: Age ≥ 15 years | (Data for Wave 1 only)<br>S. size: 18526<br>Gender: Female: 56.31%; Male: 43.69%<br>Age: 15-24: 30.65%; 25-34: 19.75%; 35-44: 16.63%; 45-44: 13.8%; 55-64: 9.57%; 65: 9.59%<br>SES: Not Deprived: 66.77%; Vulnerable: 18.71%; Deprived: 11.90%; Severe Poverty 2.63% | (only available for wave 3) TB +HIV +DM: 2509/229079 (1.1%); RR 5.16 (4.96 - 5.37) |
| Appana 2016 [S29] | South Africa (NR), Upper Middle | CS *** | District hospital | Included: MDR-TB in-patients aged ≥ 18 admitted at the hospital. | Sample size: 52<br>Gender: Female: 48%; Male: 52%<br>Age: mean (range): 34 (18-56) | MDR-TB +HIV +DM: 1/52 (1.92%) |
| Velásquez 2015 [S77] | Peru (2005-2008), Lower Middle | Coh **** | Local health care establishments. | Included: Patients with confirmed TB or presumptive TB with respiratory symptoms for at least 2 weeks and who met Peruvian National TB Programme criteria for drug-susceptibility testing referral. | Sample size: 1701<br>Gender: Female: 35.3%; Male: 64.7%<br>Age: mean (SD): 33.6 (15.4)<br>Education: Did not begin secondary level education: 20.5%<br>Occupation: Unemployed: 37.5%<br>Types of TB: PTB: 94.4%; EPTB (with or without | TB +HIV +DM: 1/1701 (0.06%)<br>TB +HIV +Alcoholism (alcohol use that interfered with family, health, or work): 85/1699 (5%)<br>TB +HIV +Drug use (use of illicit drugs): 57/1699 (3.35%) |

| | | | | | | |
|---|---|---|---|---|---|---|
| | | | | | PTB): 5.6%; Mono-resistant: 9.0%; MDR-TB: 24.0% | |
| Boillat-Blanco 2016 [S71] | Tanzania (2012-2013), Low | CC **** | Regional hospital and connected health facilities. | Cases: Consecutive adult patients (aged ≥ 18 years) presenting in the participating hospitals with new active TB.<br>Controls: Adults accompanying patients.<br>Exclusion: Having a biological relationship to a case, tuberculosis history, symptoms or signs of tuberculosis, other acute infection, or major trauma within the last 3 months. | Sample size: 530 cases, 491 control<br>Gender: Cases: Female: 36%; Male: 64%; Controls: Female:48%; Male: 52%<br>Age: mean (SD): Cases: 35.9 (12); Controls: 36.7 (13)<br>SES status: Cases: low: 32%; medium: 48%; high: 20%; Controls: low: 18%; medium: 50%; high: 33%<br>Types of TB: EPTB: 6% | TB +HIV +DM: 8/530 (1.51%); RR 3.71 (0.79 - 17.37) |
| Wu 2016 [S72] | China (2007-2008), Lower Middle | Coh ** | District tuberculosis hospital | Included: all new PTB patients who lived in Changning district during 2007-2008<br>Excluded: Patients who were infected with non-tuberculosis mycobacteria. | Sample size: 201<br>Gender: Female: 35.8%; Male: 64.2%<br>Age: median (range): 48 (15-86); <50: 54.2%; ≥50: 45.8%; | PTB +DM +HBV: 2/201 (1%) |
| Dos Santos 2017 [S1] | Brazil (2013-2015), Upper Middle | CS *** | Tertiary care hospital | Included: Hospitalized PTB with age ≥18 years and who began treatment after hospitalization.<br>Excluded: Patients already receiving treatment at admission | Sample size: 86<br>Gender: Female: 30.2%; Male: 69.8%<br>Age: mean (SD): 44.6 (15.4)<br>Education: <8 years of schooling: 66.3%<br>Types of TB: PTB: 100% | PTB +HIV +Depression (HADS depression subscale): 14/86 (16.28%)<br>WHOQOL-HIV domain - Physical : 10.9 ±2.8 (n=14)<br>WHOQOL-HIV domain - Psychological 10.7 ±2.3 (n=14)<br>WHOQOL-HIV domain - Social 10.4 ±3.4 (n=14)<br>WHOQOL-HIV domain - Environmental 10.7 ±2.1 (n=14)<br>WHOQOL-HIV domain - Level of independence 12.1 ±2.3 (n=14)<br>WHOQOL-HIV domain - Spiritual 9.4 ±3.9 (n=14)<br>PTB +Depression (HADS depression subscale) +Anxiety (HADS anxiety subscale): 19/86 (22.09%)<br>PTB +HIV +Anxiety (HADS anxiety subscale): 19/86 (22.09%)<br>WHOQOL-HIV domain - Physical : 11.1 ±3.1 (n=19)<br>WHOQOL-HIV domain - Psychological 11.3 ±2.7 (n=19)<br>WHOQOL-HIV domain - Social 11.9 ±3.9 (n=19)<br>WHOQOL-HIV domain - Environmental 10.9 ±2.2 (n=19)<br>WHOQOL-HIV domain - Level of independence 12.0 ±2.3 (n=19)<br>WHOQOL-HIV domain - Spiritual 10.6 ±4.0 (n=19) |
| Munoz-Torrico 2017 [S7] | Mexico (2010-2015), Upper Middle | Coh **** | The INER's (National Institute of Respiratory Diseases) TB clinic, as the national reference centre for TB in Mexico city | Included: Patients with MDR-TB | Sample size: 90<br>Gender: Female: 37.8%; Male: 62.2%<br>Age: mean (SD) 49.5 (11.4) in TB patients with DM (n=49), 36.4 (14.1) in TB patients without DM (n=41)<br>Types of TB: Rifampicin resistant TB or MDR-TB: 81.1%; pre-XDR-TB: 12.2%; XDR-TB: 6.7% | DR-TB +DM +chronic kidney disease: 11/90 (12.22%)<br>DR-TB +HIV +DM: 1/90 (1.11%) |
| Ambaw 2017 [S48] | Ethiopia (2014-2016), Low | CS *** | Primary care centers from five districts in Ethiopia | Included: People attending the selected health centres for TB treatment who were within one month of starting anti-TB treatment.<br>Excluded: People planning to move out of the study area. Those too ill to be interviewed at baseline. Those who | Sample size: 657<br>Gender: Female: 45.8%; Male: 54.2%<br>Age: median (IQR): 30 (16); 18-29: 46.4%; 30-39: 24.5%; 40-49: 11.4%; 50-59: 7.9%; 60-69: 5.3%; 70-79: 2.6%; 80-89: 1.8%<br>Education: Unable to read and write: 34.6%; Able to read & write only: 10.8%; Grade 1-4: 11.3%; | TB +HIV +Depression (PHQ-9): 40/657 (6.09%) |

| | | | | | | |
|---|---|---|---|---|---|---|
| | | | | were admitted to in-patient unit for more than 5 days in the last month. People with MDR-TB. People on re-treatment for TB, who had experienced previous treatment failures. | Grade 5-8: 18.0%; Grade 9-12: 16.1%; College graduate: 9.3%<br>Occupation: Government employee: 9.4%; Unemployed: 5.6%; Self-employed: 20.4%; Farmer: 26.8%; Student: 6.1%; Housewife: 16.9%; Daily labourer: 6.7%; Others: 8.1%<br>Income: < 1USD/person/day: 84.5%; ≥ 1USD/person/day: 15.2% | |
| Piparva 2017 [S42] | India (2013-2014), Lower Middle | Coh *** | District TB centre | Included: TB cases registered for treatment.<br>Excluded: Patients with MDR-TB, XDR-TB, and receiving non-DOTS treatment. | Sample size: 1340<br>Gender: Female: 28.95%; Male: 71.04%<br>Age: <15: 5.59%; 15-44: 66.19%; 45-64: 20.44%; ≥65: 7.76%<br>Types of TB: PTB: 78.95%, EPTB: 21.04% | TB +HIV +DM: 3/1081 (0.28%)<br>Unsuccessful outcome: 0 (0%) of 3 with TB+HIV+DM |
| Jalal 2017 [S23] | Malaysia (2003 - 2012), Upper Middle | CS ***** | Hospitals and health clinics in all ten districts | Included: All cases diagnosed with TB/HIV co-infection and registered to the Kelantan Health Department from 2003 up to 2012. | Sample size: 1510<br>Gender: Female: 8.9%; Male: 91.1%<br>Age: mean (SD): 35.7 (7.98)<br>Occupation: Working: 43.2%; not working: 56.8% | TB +HIV +DM:<br>Treatment success: 12 (38.7%) of 31 with TB+HIV+DM)<br>Treatment failure: 19/31 (61.3%)<br>TB +HIV +Drug use (drug abuse):<br>Treatment success: 225 (25.9%) of 869 with TB+DM+drug abuse<br>Treatment failure: 644/869 (74.1%) |
| Vashakidze 2017 [S74] | Georgia (2014 - 2015), Lower Middle | Coh **** | National Center for Tuberculosis and Lung Diseases | Included: Patients undergoing therapeutical surgery for their TB | Sample size: 137<br>Females: 30.8%; Males: 69.8%<br>Age: Median: 43.5 years (men), 28 years (women)<br>Types of TB: Drug-sensitive TB: 56.9%; Mono resistant TB: 1.5%; Rifampicin resistant TB: 1.5%; MDR-TB: 30.7%; XDR-TB: 9.5%; MDR/XDR-TB: 40.1% | TB +DM +HCV: 3/137 (2.19%)<br>TB +HIV +HCV: 1/137 (0.73%) |
| Owiti 2017 [S61] | Kenya (2016), Lower Middle | CS *** | 5 major health facilities (referral hospitals and health centres) | Included: Drug-susceptible TB patients aged ≥18 years registered for treatment. | Sample size: 454<br>Gender: Female: 40%; Male: 60%<br>Age: median (IQR): 34 (27-42); 18-39: 65%; ≥40: 35%<br>Education: median years (IQR): 12 (8-14); No formal schooling: 0.8%; Primary: 30%; Secondary: 36%; Tertiary: 34%<br>Occupation: Formal: 12%; Informal: 62%; None: 26%<br>Monthly income (USD/month): median (IQR): 70 (30-150)<br>Types of TB: PTB: 86% EPTB: 65 (14%) | TB +HIV +DM: 9/454 (1.98%) |
| Ekeke 2017 [S63] | Nigeria (2015), Lower Middle | CS ***** | Health facilities notifying and treating the highest proportion of TB patients. | Included: Patients aged ≥ 15 years with newly diagnosed TB starting treatment.<br>Excluded: Previously-treated TB patients. | Sample size: 2094<br>Gender: Female: 43.6%; Male: 56.4%<br>Age: mean (SD): 40.7 (15.7); ≤25: 17.1%; 26-35: 27.9%; 36-45: 21.9%; 46-55: 14.6%; 56-65: 11.0%; ≥ 65: 7.5%<br>Types of TB: PTB: 95.3%; EPTB: 4.7% | TB +HIV +DM: 33/2094 (1.58%) |
| Oni 2017 [S35] | South Africa (2013-2015), Upper Middle | CS ******* | TB clinic | Included: Patients aged ≥ years and had not received >48h of TB chemotherapy.<br>Excluded: Those who were critically | Sample size: 414 with TB (852 in total)<br>Gender: Female: 42.6%; Male: 57.5%<br>Age: median (IQR): 36 (30-43); 18-24: 10.4%; 25-34: 36.5%; 35-44: 33.3%; 45-54: 12.8%; ≥55: 7% | TB +HIV +DM: 31/414 (7.49%); RR 2.19 (1.20 - 3.99) |

| | | | | | | |
|---|---|---|---|---|---|---|
| | | | | ill and in need of emergency clinical care. | Education: up to primary: 32.3%; up to secondary: 64.3%; higher education: 3.5%<br>Occupation: unemployed: 74.8%<br>Monthly income (South African Rands): 0: (2.6%); 1-1600: 46.1%; 1601-3200: 29.7%; 3201-6400: 17.9%; 6401-12800: 3.5%; ≥ 12801: 0.3% | |
| Shyamala 2018 [S40] | India (2016), Lower Middle | CS ** | Tertiary care hospital | Included: PTB and EPTB patients diagnosed and registered under DOTS | Sample size: 262<br>Gender: Female: 32.71%; Male: 67.28%<br>Age: mean (range): 43.50 (18 to 69); <19: 9.3%; 20-39: 49.5%; 40-59: 31.8%; ≥60: 9.3%<br>Occupation: Employed: 25.23%; Homemakers: 23.37%; Students: 17.75%; Labourers: 15.89%; Business: 8.4%; Painter: 5.6%; farmer: 3.7%<br>Types of TB: PTB: 80.38%; EPTB: 19.62% | TB +Depression (PHQ-9) +Asthma: 4/262 (1.53%)<br>TB +Depression (PHQ-9) +Chronic lung disease (COPD): 3/262 (1.15%)<br>TB +Depression (PHQ-9) +Hyperthyroidism: 3/262 (1.15%)<br>TB +DM +Depression (PHQ-9): 11/262 (4.2%) |
| Javed 2018 [S64] | Pakistan (NR), Lower Middle | CS ** | Different TB hospitals of Faisalabad, Lahore, and Gojra | Included: Patients having clinical symptoms of PTB with positive Ziehl-Neelsen staining for acid-fast bacilli. | Sample size: 366 | PTB +DM +HCV: 29/258 (11.24%)<br>PTB +HIV +HCV: 6/258 (2.33%) |
| Azeez 2018 [S13] | South Africa (2010-2016), Upper Middle | Coh ***(*) | Two different hospitals | Included: New TB/HIV coinfected patients aged ≥15 years who had started TB treatment.<br>Excluded: Outpatients, inpatients with incomplete information, and patients who died before or during TB treatment. | Sample size: 910<br>Gender: Female: 41.8%; Male: 58.2%<br>Age: median (IQR): 37 (27-47); 15-25: 29.3%; 26-45: 48.0%; >45: 22.8%<br>Education: Illiterate: 71.0%; Literate: 29%<br>Types of TB: MDR-TB: 62.6%; XDR-TB: 37.4% | MDR-TB +HIV +DM:<br>Unsuccessful outcomes: 30 (78.9%) of 38 with TB+HIV+DM<br>MDR-TB +HIV +Hepatitis:<br>Unsuccessful treatments: 22 (95.7%) of 23 with TB+HIV+Hepatitis<br>MDR-TB +HIV +Liver diseases:<br>Unsuccessful outcome: 19 (86.4%) of 22 with TB+HIV+liver diseases<br>MDR-TB +HIV +Silicosis:<br>Unsuccessful outcome: 25 (96.2%) of 26 with TB+HIV+silicosis<br>MDR-TB +HIV +Chronic kidney disease:<br>Unsuccessful outcome: 28 (96.6%) of 29 with TB+HIV+renal failure |
| Shokri 2018 [S20] | Iran (2008-2015), Upper Middle | CS ** | Hospital | Included: PTB patients aged ≥ 18 years of age<br>Excluded: Patients with incomplete sputum test results and/or radiological changes. | Sample size: 207<br>Gender: Female: 34.3%; Male: 65.7%<br>Age: 16-30: 20.3%; 31-50: 24.2%; 51-70: 26.1%; >71: 29.5%<br>Occupation: Housewife: 30.4%; Farmer: 11.1%; Unemployed: 4.3%; Other: 26%; Not specified: 28%<br>Types of TB: PTB: 100% | TB +DM +Cardiovascular disease: 9/207 (4.35%) |
| Echazarreta 2018 [S24] | Argentina (2015-2016), Upper Middle | CS *** | Referral hospitals for TB patients from urban areas | Included: TB patients aged ≥15 years who were hospitalized or treated by outpatient clinics with confirmed PTB or EPTB, as well as patients who, although not bacteriologically confirmed, had clinical and radiological characteristics consistent with TB and were prescribed TB treatment. | Sample size: 378<br>Gender: Female: 47.8%; Male: 58.2%<br>Age: median (range): 37 (15-89 years); 15-45: 65%; >45: 35%<br>Types of TB: PTB: 69.6%; EPTB (with or without PTB): 28.8% | TB +AIDS +Drug use (drug addiction): 29/376 (7.71%)<br>TB +AIDS +Alcoholism (alcohol ingestion of >50g/day (women) or >70g/day (men)): 14/376 (3.72%) |
| Pawar 2018 [S46] | India (2012), Lower Middle | CS **** | Drug-Resistant Tuberculosis Centre in hospital. | Included: All confirmed MDR PTB patients admitted for pre-treatment evaluation. | Sample size: 127<br>Gender: Female: 20.4%; Male: 79.5%<br>Age: mean: 38.6 years | MDR-TB +Chronic lung disease (COPD) +Silicosis: 3/127 (2.36%)<br>MDR-TB +DM +Chronic lung disease (COPD): 2/127 (1.57%)<br>MDR-TB +Chronic lung disease (COPD) +Cor pulmonale: 1/127 (0.79%) |

| | | | | | | |
|---|---|---|---|---|---|---|
| | | | | Excluded: Incomplete records being transferred to some other unit. | | |
| Alinaitwe 2018 [S69] | Uganda (NR), Low | CS *** | TB clinic. | Included: Patients aged ≥ 18 years attending TB clinic. | Sample size: 308<br>Gender: Female: 38.76%; Male: 61.24%<br>Age: mean (SD): 36 (10.8); <30: 31.27%; 30-50: 59.61%; >50: 9.12%;<br>Education: None: 4.56%; Primary: 43.97%; Secondary: 39.74%; University/Technical: 11.73%<br>Occupation: Employed: 60.26%; No formal employment: 15.64%; Unemployed: 24.10%<br>Types of TB: PTB: 79.15%; EPTB: 17.92%; PTB & EPTB: 2.93% | TB +HIV +Depression (MINI): 44/308 (14.29%)<br>TB +Depression (MINI) +Drug use (drug abuse): 10/308 (3.25%) |
| Wang 2018 [S32] | China (2014-2015), Upper Middle | CS ** | 3 hospitals | Included: PTB patients who were inpatients and newly diagnosed or were undergoing treatment via DOT<br>Excluded: Patients with drug resistance, EPTB without PTB, patients who were pregnant, critically ill, or had communication problems; and patients with severe illnesses, such as cancer. | Sample size: 1252<br>Gender: Female: 40.97%; Male: 59.03%<br>Age: mean (SD): 44.35 (17.36); 18-30: 30.35%; 31-50: 29.63%; 51-80: 37.22%; >80: 2.80%<br>Monthly income (Yuan): <1,500: 25.48%; 1,500-3,000: 44.33%; 3,000-5,000: 22.28%; 5,000-10,000: 3.75%; >10,000: 1.04%<br>Education: Elementary: 9.66%; Middle school: 34.27%; High school: 27.24%; Vocational/college: 26.52%; Undergraduate: 1.28%; None: 0.96%<br>Occupation: Unemployed: 57.41%; Employed: 42.11% | PTB +Depression (HADS depression subscale) +Anxiety (HADS anxiety subscale): 160/1252 (12.78%) |
| Mukhtar 2018 [S76] | Pakistan (2013-2014), Lower Middle | Coh *** | Tertiary care hospital | Included: New PTB patients aged ≥ 15 years registered for antituberculosis treatment. | Sample size: 614<br>Gender: Female: 49%: Male: 51%<br>Age: 15-19: 22%; 20-24: 23%; 25-29: 11%; 30-39: 15%; 40-49: 11%; >50: 18%<br>Education: illiterate: 52%; primary: 14%; matriculation: 24%; intermediate: 5%; bachelors: 3%; masters and above: 2%<br>Income category (Rupees): Nil (income in the form of loans/ help from relatives/extended family/friends): 63%; <5000: 7%; 5100-8000: 11%; 8100-11000: 9%; 11100-14000: 4%; 14100-17000: 3% | PTB +DM +Drug use (drug abuse): 2/614 (0.33%)<br>PTB +DM +Heart disease: 14/614 (2.28%) |
| Pande 2018 [S44] | India (2015-2016), Lower Middle | Coh ***** | Tertiary teaching hospital | Included: All hospitalized patients ≥ 18 years with PTB or EPTB<br>Excluded: Pregnant TB patients. | Sample size: 728<br>Gender: Female: 29%; Male: 71%<br>Age: 18-40: 42%; 41-60: 39%; ≥60: 18%; unknown: 2%<br>Types of TB: PTB: 56%; EPTB: 44% | TB +HIV +DM: 2/728 (0.27%) |
| Yaneth-Giovanetti 2019 [S2] | Colombia (2015), Upper Middle | CS *** | Public Hospital | Included: TB patients who attended treatment at the Anti-tuberculosis League of the Public Hospital | Sample size: 70<br>Gender: Female: 45.7%; Male: 54.3%<br>Age: Mean (range): 35.8 (7-88); > 40: 51.4%; <40: 48.6%<br>Types of TB: PTB 100% | TB +HIV +DM: 0/70 (0%) |

| | | | | | | |
|---|---|---|---|---|---|---|
| Castro 2019 [S3] | Brazil (2016), Upper Middle | CS *(*) | Registered in the Brazilian Information System for Notifiable Diseases (SINAN) | Included: New TB cases with closure status (cured, abandoned, death from TB and death from other causes).<br>Exclusion: Cases without a defined closure status | Sample size: 180<br>Gender: Female: 24.4%; Male: 75.6%<br>Age: 0-9: 1.7%; 10-19: 0.6%; 20-29: 15.6%; 30-39: 36.7%; 40-49: 25.6%; 50-59: 15.0%; ≥60: 5.0%<br>Education: 1st to 4th grades: 14.4%; 5th to 8thgrades: 18.9%; High school: 7.2%; Higher education: 3.3%; Illiterate: 0.6%; Not applicable: 1.7%; Ignored: 53.9%<br>Types of TB: PTB 60.6%; EPTB: 28.9%; PTB & EPTB: 10.6% | TB +HIV +DM:<br>Cure: 1 (33%) of 3 with TB+HIV+Diabetes<br>Abandonment: 1/3 (33%)<br>Death TB: 0/3 (0%)<br>Death other causes: 1/3 (33%)<br>TB +HIV +Drug use (use of illicit drugs):<br>Cure: 9 (21%) of 42 with TB+HIV+Illicit drug use<br>Abandonment: 18/42 (43%)<br>Death TB: 1/42 (2%)<br>Death other causes: 14/42 (33%)<br>TB +HIV +Mental disorder:<br>Cure: 0 (0%) of 3 with TB+HIV+Mental illness<br>Abandonment: 2/3 (67%)<br>Death TB: 0/3 (0%)<br>Death other causes: 1/3 (33%) |
| Chin 2019 [S82] | Zimbabwe (2011-2014), Low | Coh * | Harare (not further specified) | Included: Adult participants requiring retreatment of TB<br>Excluded: Active TB | Sample size: 486 initially recruited, 175 with follow-up included in analysis<br>Gender: Female: 41.7%; Male: 58.3%<br>Age: mean (IQR): 41 (33-48)<br>Income: US $0 ≤99/month: 21.1%; US $100-199/month: 25.1%;US $200-299/month: 21.7%; US $300-2000/month: 31.4%<br>Types of TB: Unconfirmed: 21.1%; Drug-susceptible TB: 29.1%; Rifampin-monoresistant: 22.9%; Multidrug-resistant: 26.9% | TB +Depression (CES-D) +Chronic lung disease: 5/175 (2.86%)<br>TB +Depression (CES-D) +PTSD: 6/175 (3.43%)<br>TB +PTSD +Chronic lung disease: 2/175 (1.14%) |
| Munseri 2019 [S67] | Tanzania (2016-2017), Low | CS ***** | Four TB clinics in Dar es Salaam, Tanzania | Included: Patient's age ≥18 years on TB treatment | Sample size: 660<br>Gender: Female: 37%; Male: 63%<br>Age: mean (SD): 39.2 (14.3); ≤ 24: 13.2%; 25-34: 27.4%; 35-44: 26.1%; ≥45: 33.3%<br>Education: No formal education: 9.8%; Primary: 65.2%; Secondary: 18.3%; College: 6.7%<br>Occupation: Unemployed: 73.3%; Retired: 4.7%; Employed: 22%<br>Types of TB: PTB: 87.4%; EPTB: 12.6% | TB +HIV +DM: 11/660 (1.67%) |
| Azovtzeva 2019 [S8] | Russia (2016-2018), Upper Middle | CS ** | City Tuberculosis Hospital in St. Petersburg (n = 94) and the Novgorod Clinical Specialized Center in Veliky Novgorod (n = 43) | Included: Cases of co-morbidity (HIV infection, TB and chronic hepatitis), in which TB was the dominant diagnosis | Sample size: 137<br>Gender: Female: 30.7%; Male: 69.3%<br>Age: mean (SD): 37.4 (0.71)<br>Occupation: non-working: 88.4% | TB +HIV +Hepatitis:<br>Mortality: 8 (7.55%) of106 with HIV+TB+hepatitis |
| Walker 2019 [S75] | Nepal (2015-2016), Low | Coh *** | Two urban TB clinics | Included: Patients aged ≥18 years with MDR-TB.<br>Excluded: Drug-susceptible TB or XDR-TB. | Sample size: 135<br>Gender: Female: 24.4%; Male: 75.6%<br>Age: median (IQR): 30 (23-43)<br>Occupation (before MDR-TB diagnosis): Not working: 2.2%; Housewife: 10.4%; Student: 16.3%; Paid work: 71.1%<br>Occupation (after MDR-TB diagnosis): Not working: 69.6%; Housewife: 4.4%; Student: 5.2%; | MDR-TB +Depression (HSCL-25) +Anxiety (HSCL-25): 20/135 (14.81%) |

| | | | | | | |
|---|---|---|---|---|---|---|
| | | | | | Paid work: 20.7%;<br>Types of TB: all MDR-TB | |
| Whitehouse 2019 [S14] | South Africa (2014-2016), Upper Middle | CS **** | 10 DR-TB hospitals (5 urban, 5 rural) | Included: Patients with DR-TB (rifampicin-resistant) aged ≥ 18 enrolled in a parent cluster-randomized trial assessing case management by nurses of patients with DR-TB. | Sample size: 900<br>Gender: Female: 46.3%; Male: 53.7%<br>Age: mean (SD): 36.2 (10.8)<br>Types of TB: all DR TB | DR-TB +HIV +DM: 21/862 (2.44%) |
| Khan 2019 [S25] | Malaysia (2006 - 2008), Upper Middle | Coh **** | Selected hospitals and prisons | Included: EPTB patients<br>Excluded: Patients who had both PTB and EPTB. | Sample size: 1222<br>Gender: Female: 36.3%; Male: 63.7%<br>Age: <15: 4.5%; 16-25: 14.5%; 26-35: 24.2%; 36-45: 18.3%; 46-55: 19.1%; 56-65: 13.9%; ≥66: 5.5%<br>Education: Unknown: 71.7%; Primary: 9.1%; Secondary: 6.9%; College: 3.4%; University: 0.7%; Diploma: 1.6%; No formal education: 6.6%<br>Occupation: Unknown: 15.4%; Employed: 28%; Unemployed: 56.6%<br>Types of TB: PTB: 86.9% | EPTB +HIV +DM: 7/1222 (0.57%)<br>Treatment success: 2 (28.6%) of 7 with EPTB +HIV +DM<br>EPTB +DM +Hepatitis: 11/1222 (0.9%)<br>Treatment success: 3 (27.3%) of 11 with EPTB +DM +Hepatitis<br>EPTB +HIV +Hepatitis: 79/1222 (6.46%)<br>Treatment success: 56 (70.9%) of 79 with EPTB +HIV +Hepatitis |
| Dasa 2019 [S51] | Ethiopia (2017), Low | CS **** | 8 hospitals and 3 health centers in eastern Ethiopia | Included: TB patients who were under anti TB treatments for more than one month. | Sample size: 403<br>Gender: Female: 40.7%; Male: 59.3%<br>Age: ≤24: 31.0%; 25-34: 30.5%; >35: 38.5%<br>Education: No formal education: 25.8%; Primary education: 35.5%; Secondary and above: 38.7%<br>Occupation: Employed: 13.9%; Private: 32.5%; Student: 17.1%; Farmer/Daily laborers: 36.5%<br>Income (monthly): 25th percentile: 30.8%; 50th percentile: 27.0%; 75th percentile: 42.2%<br>Types of TB: MDR-TB: 24.8% | TB +HIV +Depression (PHQ-9): 28/403 (6.95%) |
| Sharman 2019 [S26] | South Africa (2009-2015), Upper Middle | Coh **** | Umkhanyakude district of rural KwaZulu-Natal, South Africa | Included: Adults aged ≥15 years who had been surveyed for a population-based HIV and health surveillance study and who had been tested for HIV | Sample size: 47334<br>Gender: Female: 53%; Male: 47%<br>Age: median (IQR): 32 (20-52) | TB +HIV +DM: 327/12118 (2.7%) |
| Cespedes 2019 [S27] | Paraguay (2016-2017), Upper Middle | CS **** | All the health services of the Ministry of Public Health and Welfare Social and Social Security notify the cases to the PNCT (National TB Control Program) | Included: Patients with any type of TB, with or without DM, and registered in the official database of the National TB Control Program.<br>Excluded: DR-TB cases | Sample size: 5135<br>Gender: Female: 29.85%; Male: 70,15%<br>Age: <15: 8,3%; 15-30: 33,9%; 31-45: 25,1%; 46-60: 16,8%; >61: 15,8%<br>Types of TB: PTB: 85,7%; EPTB: 9,5%; PTB & EPTB: 4,8% | TB +HIV +DM: 9/5135 (0.18%)<br>TB +DM +Chronic lung disease (COPD): 23/5135 (0.45%)<br>TB +DM +Drug use (psychoactive drug addiction): 9/5135 (0.18%) |
| Molla 2019 [S52] | Ethiopia (2018), Low | CS **** | Specialized hospital. | Included: Patients aged ≥ 15 visiting an outpatient TB clinic. | Sample size: 415<br>Gender: Female: 46.5%; Male: 53.5%<br>Age: mean (SD): 34.56 (12.6); range: 15-78; 15-24: 23.1%; 25-34: 33.74%; 35-44: 20%; 45-54: 14.7%; >54: 8.4%<br>Education: No formal education: 19%; Primary: 28.4%; Secondary: 22.7%; Preparatory: 15.2%; College and above: 14.7%<br>Occupation: Trading: 23.4%; Farming: 24.34%; | TB +HIV +Depression (PHQ-9): 21/415 (5.06%) |

| | | | | | | |
|---|---|---|---|---|---|---|
| | | | | | Government employee: 32.52%; Private employee: 14.94%; Others (housewife and students): 4.82%<br>Monthly income (Ethiopian Birrs): <1,539 ETB: 59.3%; ≥ 1,539 ETB: 40.7%<br>Types of TB: PTB: 83%; EPTB: 17%; non-MDR-TB: 74%; MDR-TB: 26% | |
| Ugarte-Gil 2020 [S60] | Indonesia, Peru, Romania, South Africa (2014-2016), Lower Middle | CS **** | Indonesia: 44 community health centres and a referral hospital. Peru: 3 primary health facilities and 1 secondary level hospital. Romania: 2 secondary level hospitals. South Africa: 6 community health care clinics. | Included: PTB patients presenting for treatment at study sites.<br>Excluded: Patients with DM. | Sample size: 2185 (Indonesia n=748; Peru n=600; Romania n=506 and South Africa n=331)<br>Gender: Female: 38.8%; Male: 61.2%<br>Age: median (IQR): 36.6 (26.0-49.1)<br>SES index: Quintile 1: 28.7%; Quintile 2: 22.9%; Quintile 3: 20.6%; Quintile 4: 16.4%; Quintile 5: 9.5%<br>Types of TB: all PTB | PTB +DM +Chronic kidney disease: 12/2185 (0.55%) |
| Mohammed-hussein 2020 [S56] | Ethiopia (2019), Low | CS **** | 39 health facilities | Included: PTB patients who visited the selected health facilities during the study period.<br>Excluded: Critically ill patients and those on treatment for less than a month | Sample size: 410<br>Gender: Female: 49.5%; Male: 51.5%<br>Age: mean (SD): 31.85 (12.42); 18-24: 32.2%; 25-34: 35.4%; 35-44: 13.9%; 45-54: 11.2%; ≥55: 7.3%<br>Education: No formal education:22%; Primary (1-8): 35.9%; Secondary: 26.6%; College and above (15.6%)<br>Occupation: Daily laborer: 13.7%; Farmer: 21.2%; Government employee: 12.9%; Merchant: 9.3%; Student: 14.4%; Housewife: 16.8%; Non-government organization/private employee: 11.7% | TB +HIV +Depression (HADS depression subscale): 70/410 (17.07%)<br>TB +HIV +Anxiety (HADS anxiety subscale): 66/410 (16.1%) |
| White 2020 [S65] | Philippines (2017), Lower Middle | CS **** | Three TB-DOTS clinics in an urban setting and two sites from a rural setting. | Included: Non-pregnant patients aged ≥18 years with ongoing TB treatment, including patients who had stopped attending the TB DOTS clinic or visited infrequently. | Sample size: 637<br>Gender: Female: 30.0%; Male: 70.0%<br>Age: 18-40: 42.7%; 41-65: 44.1%; >65: 13.2%<br>Education: Primary: 34.2%; Secondary: 39.4%; Tertiary: 23.1%; Vocational: 3.3%<br>Types of TB: MDR-TB: 5.2%; non-MDR-TB: 94.8% | TB +HIV +DM: 1/637 (0.16%) |
| Adejumo 2020 [S62] | Nigeria (2014-2017), Lower Middle | CS **** | All (314) public and private health facilities providing TB services in Lagos State | Included: DR-TB patients with laboratory tests data.<br>Excluded: Patients who could not be reached (wrong contact details or moved to another state) | Sample size: 565<br>Gender: Female: 38.2%; Male: 61.8%<br>Age: mean (SD): 34.2 (12.5); <15: 2.5%; 15-24: 19.8%; 25-34: 33.8%; 35-44: 24.2%; 45-54: 12.2%; ≥55: 7.4% | DR-TB +HIV +Hypothyroidism: 13/565 (2.3%)<br>DR-TB +HIV +DM: 4/565 (0.71%)<br>DR-TB +HIV +Hypothyroidism + DM: 2/565 (0.35%)<br>DR-TB +Hypothyroidism +DM: 4/565 (0.71%) |

AIDS: Acquired immune deficiency syndrome; ART: Antiretroviral therapy; AUDIT: Alcohol use disorders identification test; CC: Case-Control study; CDU: Chronic Dispensing Unit; Coh: Cohort study; COPD: Chronic obstructive pulmonary disease ; CS: Cross-sectional study; DM: Diabetes Mellitus; DOTS: Directly Observed Treatment Short-course; DR-TB: Drug-resistant TB; DSM-IV: Diagnostic and Statistical Manual of Mental Disorders, version 4; EPTB: Extra-pulmonary TB; HADS: Hospital Anxiety and Depression Scale; HbA1c: glycated haemoglobin; HBV: Hepatitis B Virus; HCV: Hepatitis C Virus; HIV: Human immunodeficiency virus; HSCL-25: Hopkins Symptom Checklist-25; IQR: Interquartile range; Kessler 10 scale: Kessler Psychological Distress Scale; LFU: Loss to follow-up; MDR-TB: Multidrug resistant TB; MINI: Mini-International Neuropsychiatric Interview; PHQ-9: Patient Health Questionnaire; PTB: Pulmonary TB; PTSD: Post-traumatic stress disorder; RR: Risk Ratio; Standard deviation; SES: Socio-economic status; SINAN: Brazil's Information System for Notifiable Diseases, Sistema de Informação de Agravos de Notificação; T. pallidum: Treponema pallidum; Tuberculosis; TSH: thyroid stimulating hormone; WHOQOL-HIV: Quality of life assessment developed by the WHO for people with HIV; XDR-TB: Extensively drug-resistant TB.

[*] Quality assessed using a modified version of the Newcastle-Ottawa Scale
[†] Treatment outcomes have one additional * because length of follow-up was =>6 months

### Supplementary Table 3: Prevalence of each cluster of tuberculosis multimorbidity, grouped by the most prevalent comorbidities

| Cluster | Prevalence (95%CI)* | Number of studies (participants) | Range | $I^2$ † |
|---|---|---|---|---|
| **TB+HIV+…** | | | | |
| TB+HIV + Anxiety | 15.23% (11.25% - 19.67%) | 4 (1,413) | 10.07% - 16.40% | 77.37% |
| TB+HIV + PTSD | 14.84% (13.9%-15.8%) | 2 (5,400) | 2.40% - 16.63% | - |
| TB+HIV + HBV | 9.70% (2.37%-21.04%) | 3 (792) | 2.21% - 20.57% | 94.81% |
| TB+HIV + Depression | 9.66% (6.96% - 12.74%) | 9 (3,418) | 5.06% - 16.28% | 87.47% |
| TB+HIV + Alcoholism | 9.29% (4.98%-14.72%) | 4 (6,963) | 5.00%-16.55% | 96.34% |
| TB+ HIV + Hepatitis | 6.46% (5.15% - 7.99%) | 1 (1,222) | | |
| TB+HIV + Hypothyroidism | 5.37% (0.90% - 12.94%) | 3 (1,108) | 2.30% - 10.84% | 94.00% |
| TB+HIV + Drug use (illicit) | 5.21% (2.73% - 8.39%) | 4 (2,584) | 2.74% - 5.87% | 86.19% |
| TB+HIV + Heart disease | 4.65% (1.28% - 11.48%) | 1 (86) | | |
| TB+HIV + HBV + HCV | 4.42% (1.93% - 8.52%) | 1 (181) | | |
| TB+HIV + Cancer | 3.49% (0.73% - 9.86%) | 1 (86) | | |
| TB+HIV + Autoimmune diseases | 3.49% (0.73% - 9.86%) | 1 (86) | | |
| TB+HIV + OCD | 2.2% (1.1% - 3.9%) | 1 (500) | | |
| TB+HIV + Panic disorder | 2.2% (1.1% - 3.9%) | 1 (500) | | |
| TB+HIV + HCV | 1.47% (0.36% - 3.21%) | 6 (1,550) | 0.33% - 4.98% | 79.36% |
| TB+HIV + Cardiovascular disease | 1.03% (0.21% - 2.97%) | 1 (292) | | |
| TB+HIV + DM | 0.98% (0.70% - 1.29%) | 40 (526,827) | 0% - 8.33% | 97.79% |
| TB+HIV + Chronic liver disease | 0.48% (0.03% - 1.27%) | 2 (682) | 0.50% - 2.33% | - |
| TB+HIV + DM +Hypothyroidism | 0.35% (0.04% - 1.27%) | 1 (565) | | |
| TB+ HIV + Chronic kidney disease | 0.17% (0% - 0.93%) | 1 (596) | | |
| TB+HIV + Chronic lung disease | 0.10% (0% - 0.64%) | 2 (682) | 0.17% - 1.16% | - |
| **TB+AIDS+…** | | | | |
| TB+AIDS + Drug use | 7.71% (5.23% - 10.89%) | 1 (376) | | |
| TB+AIDS + Alcoholism | 3.72% (2.05% - 6.17%) | 1 (376) | | |
| TB+AIDS + Mental disorder | 0.36% (0.3% - 0.42%) | 1 (39,881) | | |
| TB+AIDS + DM | 0.28% (0.23% - 0.33%) | 1 (39,881) | | |
| TB+AIDS + DM+Mental disorder | 0.06% (0.04% - 0.09%) | 1 (39,881) | | |
| TB+AIDS + Cancer | 0.04% (0.02% - 0.07%) | 1 (39,881) | | |
| TB+AIDS + Heart disease | 0.02% (0.01% - 0.04%) | 1 (39,881) | | |
| TB+AIDS + Chronic lung disease | 0.01% (0% - 0.02%) | 1 (39,881) | | |
| TB+AIDS + Mental disorder+Cancer | 0.01% (0% - 0.02%) | 1 (39,881) | | |
| TB+AIDS + DM+Chronic lung disease | <0.01% (0% - 0.01%) | 1 (39,881) | | |
| **TB+DM+…** | | | | |
| TB+DM + HCV | 7.41% (4.99%-10.24%) | 2 (395) | 2.19%-11.24% | - |
| TB+DM + Cardiovascular disease | 4.35% (2.01% - 8.09%) | 1 (207) | | |
| TB+ DM + Depression | 4.2% (2.11% - 7.39%) | 1 (262) | | |
| TB+DM + Drug use | 1.03% (0.22% - 2.34%) | 5 (7,476) | 0.17% - 6.76% | 90.85% |
| TB+DM + HBV | 1.00% (0.12% - 3.55%) | 1 (201) | | |
| TB+ DM + Hepatitis | 0.9% (0.45% - 1.6%) | 1 (1,222) | | |

| | | | | |
|---|---|---|---|---|
| TB+DM + Hypothyroidism | 0.71% (0.19% - 1.8%) | 1 (565) | | |
| TB+DM + Alcoholism | 0.69% (0.47% - 0.94%) | 2 (5,218) | 0.63% - 0.76% | - |
| TB+DM + Chronic kidney disease | 0.50% (0.20% – 0.89%) | 2 (2,275) | 0.55% - 12.22% | - |
| TB+DM + Mental disorder | 0.3% (0.24% - 0.35%) | 1 (39,881) | | |
| TB+DM + Chronic lung disease | 0.23% (0%-0.98%) | 3 (45,143) | 0.03%-1.57% | 96.08% |
| TB+DM + Heart disease | 0.05% (0.02% - 0.07%) | 2 (40,495) | 0.07% - 2.28% | - |
| TB+DM + Cancer | 0.04% (0.02% - 0.06%) | 1 (39,881) | | |
| TB+DM + Arthritis | 0.01% (0% - 0.03%) | 1 (39,881) | | |
| TB+DM +Heart disease+Chronic lung disease | 0.01% (0% - 0.02%) | 1 (39,881) | | |
| TB+DM + Mental disorder+Chronic lung disease | 0.01% (0% - 0.02%) | 1 (39,881) | | |
| TB+DM + Cancer+Heart disease | <0.01% (0% - 0.01%) | 1 (39,881) | | |
| TB+DM + Mental disorder+Arthritis | <0.01% (0% - 0.01%) | 1 (39,881) | | |
| TB+DM + Mental disorder+Cancer | <0.01% (0% - 0.01%) | 1 (39,881) | | |
| TB+DM + Mental disorder + Heart disease | <0.01% (0% - 0.01%) | 1 (39,881) | | |
| TB+DM + Heart failure | 0% (0% - 3.39%) | 1 (107) | | |
| TB+DM + Unstable angina | 0% (0% - 3.39%) | 1 (107) | | |
| TB+DM + Chronic liver disease | 0% (0% - 0%) | 2 (181,485) | 0% - 0.03% | - |
| **TB+Depression+…** | | | | |
| TB+Depression + Anxiety | 15.27% (10.70% - 20.47%) | 3 (1,473) | 12.78% - 22.09% | 63.81% |
| TB+Depression + PTSD | 3.43% (1.27% - 7.31%) | 1 (175) | | |
| TB+Depression + Drug use | 3.25% (1.57% - 5.89%) | 1 (308) | | |
| TB+Depression + Chronic lung disease | 1.73% (0.65% - 3.24%) | 2 (437) | 1.15% - 2.86% | - |
| TB+Depression + Asthma | 1.53% (0.42% - 3.86%) | 1 (262) | | |
| TB+Depression + hyperthyroidism | 1.15% (0.24% - 3.31%) | 1 (262) | | |
| **TB+Chronic lung disease+…** | | | | |
| TB+Chronic lung disease + Silicosis | 2.36% (0.49% - 6.75%) | 1 (127) | | |
| TB+Chronic lung disease + PTSD | 1.14% (0.14% - 4.07%) | 1 (175) | | |
| TB+Chronic lung disease + Cor pulmonale | 0.79% (0.02% - 4.31%) | 1 (127) | | |
| TB+Chronic lung disease + Heart disease | 0.02% (0.01% - 0.04%) | 1 (39,881) | | |
| TB+Chronic lung disease + Cancer | 0.02% (0.01% - 0.03%) | 1 (39,881) | | |
| TB+Chronic lung disease + Mental disorder | 0.01% (0% - 0.03%) | 1 (39,881) | | |
| **TB+Drug use+…** | | | | |
| TB+Drug use + Alcoholism | 10% (5.82% - 15.73%) | 1 (160) | | |
| TB+Drug use + HCV | 4.39% (2.03% - 8.17%) | 1 (205) | | |
| TB+Drug use + HBV | 3.44% (0% - 12.05%) | 3 (763) | 0% - 9.78% | 95.20% |
| TB+Drug use + T. pallidum | 0.49% (0.01% - 2.69%) | 1 (205) | | |
| **TB+Cancer+…** | | | | |
| TB+Cancer + Mental disorder | 0.02% (0.01% - 0.03%) | 1 (39,881) | | |
| TB+Cancer + Arthritis | 0.01% (0% - 0.02%) | 1 (39,881) | | |
| TB+Cancer + Heart disease | 0.01% (0% - 0.02%) | 1 (39,881) | | |
| **TB+other+…** | | | | |
| TB+PTSD + Alcoholism | 7.06% (6.36% - 7.81%) | 1 (4,900) | | - |
| TB+HBV + HCV | 1.56% (0.60%-2.90%) | 2 (500) | 1.00%-2.00% | - |

| | | |
|---|---|---|
| TB+Heart disease + Mental disorder | 0.01% (0% - 0.02%) | 1 (39,881) |
| TB+Arthritis + Mental disorder | 0.01% (0% - 0.02%) | 1 (39,881) |

*Notes:* AIDS: Acquired immune deficiency syndrome; DM: Diabetes Mellitus; HBV: Hepatitis B Virus; HCV: Hepatitis C Virus; HIV: Human immunodeficiency virus; OCD: Obsessive compulsive disorder; PTSD: Post-traumatic stress disorder.

[*] Pooled prevalence and 95% confidence intervals from random-effects meta-analysis when the number of studies is >1.

[†] I-squared values reported only for meta-analyses with >2 studies

### Supplementary Table 4: Prevalence of treatment outcomes in each cluster

| Cluster | Died | Cured | Lost to follow-up | Treatment success | Treatment failure |
|---|---|---|---|---|---|
| TB+HIV + DM | 33.33% (0.84% - 90.57%) | 33.33% (0.84% - 90.57%) | 33.33% (0.84% - 90.57%) | 36.30% (15.63% - 59.33%, 5 studies, 4,903 participants, range: 21.05% to 100%, $I^2$=59.16%) | 62.17% (36.55% - 85.13%, 4 studies, 4,723 participants, range: 0% to 78.95%, $I^2$ = 69.26%) |
| TB+HIV + Drug use | 35.71% (21.55% - 51.97%) | 21.43% (10.3% - 36.81%) | 42.86% (27.72% - 59.04%) | 25.44% (22.62% - 28.36%, 2 studies, 1,690 participants, range: 21.43% to 25.89%, $I^2$=NA) | |
| TB+HIV + HBV | 11.63% (3.89% - 25.08%) | 16.28% (6.81% - 30.7%) | 18.6% (8.39% - 33.4%) | 53.49% (37.65% - 68.82%) | 2.33% (0.06% - 12.29%) |
| TB+HIV + HCV | 17.62% (12.72% - 23.46%) | 30% (23.89% - 36.69%) | 9.52% (5.91% - 14.33%) | 60.95% (54% - 67.59%) | .95% (0.12% - 3.4%) |
| TB+HIV + HBV+HCV | 25.93% (11.11% - 46.28%) | 22.22% (8.62% - 42.26%) | 3.7% (0.09% - 18.97%) | 55.56% (35.33% - 74.52%) | 0% (0% - 12.77%) |
| TB+HIV + Hepatitis | 7.55% (3.31% - 14.33%) | | | 54.29% (44.38% - 64.04%, 2 studies, 2,132 participants, range 4.39% to 70.89%, $I^2$=NA) | 45.71% (35.96% - 55.62%, 2 studies, 2,132 participants, range: 29.11% to 95.65%, $I^2$=NA) |
| TB+HIV + Liver diseases | | | | 13.64% (2.91% - 34.91%) | 86.36% (65.09% - 97.09%) |
| TB+HIV + Mental disorder | 33.33% (0.84% - 90.57%) | 0% (0% - 70.76%) | 66.67% (9.43% - 99.16%) | 0% (0% - 70.76%) | |
| TB+HIV + Chronic kidney disease | | | | 3.45% (0.09% - 17.76%) | 96.55% (82.24% - 99.91%) |
| TB+HIV + Silicosis | | | | 3.85% (0.1% - 19.64%) | 96.15% (80.36% - 99.9%) |
| TB+HIV + Depression | | | | | |
| TB+HIV + Anxiety | | | | | |
| TB+AIDS + Cancer | 58.82% (32.92% - 81.56%) | 29.41% (10.31% - 55.96%) | 11.76% (1.46% - 36.44%) | | |
| TB+AIDS + Chronic lung disease | 100% (15.81% - 1%) | 0% (0% - 84.19%) | 0% (0% - 84.19%) | | |
| TB+AIDS + DM | 43.64% (34.2% - 53.42%) | 40% (30.78% - 49.78%) | 14.55% (8.55% - 22.54%) | | |
| TB+AIDS + DM +Chronic lung disease (COPD) | 100% (2.5% - 1%) | 0% (0% - 97.5%) | 0% (0% - 97.5%) | | |
| TB+AIDS + DM +Mental disorder | 26.09% (10.23% - 48.41%) | 47.83% (26.82% - 69.41%) | 21.74% (7.46% - 43.7%) | | |
| TB+AIDS + Heart disease | 50% (15.7% - 84.3%) | 25% (3.19% - 65.09%) | 25% (3.19% - 65.09%) | | |
| TB+AIDS + Mental disorder | 37.06% (29.14% - 45.53%) | 30.77% (23.33% - 39.03%) | 31.47% (23.97% - 39.76%) | | |
| TB+AIDS + Mental disorder +Cancer | 100% (15.81% - 1%) | 0% (0% - 84.19%) | 0% (0% - 84.19%) | | |
| TB+DM + Arthritis | 25% (0.63% - 80.59%) | 50% (6.76% - 93.24%) | 25% (0.63% - 80.59%) | | |
| TB+DM + Cancer | 73.33% (44.9% - 92.21%) | 26.67% (7.79% - 55.1%) | 0% (0% - 21.8%) | | |
| TB+DM + Cancer +Heart disease | 100% (2.5% - 1%) | 0% (0% - 97.5%) | 0% (0% - 97.5%) | | |

| | | | | | |
|---|---|---|---|---|---|
| TB+DM + Chronic lung disease | 69.23% (38.57% - 90.91%) | 23.08% (5.04% - 53.81%) | 7.69% (0.19% - 36.03%) | | |
| TB+DM + Heart disease | 28.57% (13.22% - 48.67%) | 71.43% (51.33% - 86.78%) | 0% (0% - 12.34%) | | |
| TB+DM+ Heart disease +Chronic lung disease (COPD) | 50% (1.26% - 98.74%) | 50% (1.26% - 98.74%) | 0% (0% - 84.19%) | | |
| TB+DM + Hepatitis | | | | 27.27% (6.02% - 60.97%) | 72.73% (39.03% - 93.98%) |
| TB+DM + Mental disorder | 17.8% (11.37% - 25.91%) | 66.1% (56.81% - 74.56%) | 15.25% (9.3% - 23.03%) | | |
| TB+DM + Mental disorder +Arthritis | 0% (0% - 97.5%) | 0% (0% - 97.5%) | 100% (2.5% - 1%) | | |
| TB+DM + Mental disorder +Cancer | 100% (2.5% - 1%) | 0% (0% - 97.5%) | 0% (0% - 97.5%) | | |
| TB+DM+Mental disorder +Chronic lung disease (COPD) | 100% (29.24% - 1%) | 0% (0% - 70.76%) | 0% (0% - 70.76%) | | |
| TB+DM + Mental disorder +Heart disease | 0% (0% - 97.5%) | 100% (2.5% - 1%) | 0% (0% - 97.5%) | | |
| TB+Arthritis + Cancer | 0% (0% - 84.19%) | 100% (15.81% - 1%) | 0% (0% - 84.19%) | | |
| TB+Cancer + Chronic lung disease | 66.67% (22.28% - 95.67%) | 33.33% (4.33% - 77.72%) | 0% (0% - 45.93%) | | |
| TB+Cancer + Heart disease | 66.67% (9.43% - 99.16%) | 33.33% (0.84% - 90.57%) | 0% (0% - 70.76%) | | |
| TB+Heart disease + Chronic lung disease | 50% (15.7% - 84.3%) | 50% (15.7% - 84.3%) | 0% (0% - 36.94%) | | |
| TB+Mental disorder + Arthritis | 0% (0% - 84.19%) | 50% (1.26% - 98.74%) | 50% (1.26% - 98.74%) | | |
| TB+Mental disorder + Cancer | 66.67% (22.28% - 95.67%) | 33.33% (4.33% - 77.72%) | 0% (0% - 45.93%) | | |
| TB+Mental disorder + Chronic lung disease | 60% (14.66% - 94.73%) | 40% (5.27% - 85.34%) | 0% (0% - 52.18%) | | |
| TB+Mental disorder + Heart disease | 0% (0% - 84.19%) | 100% (15.81% - 1%) | 0% (0% - 84.19%) | | |

Note: Unless otherwise stated, each result was reported by only one study. Given the high heterogeneity in most of the meta-analyses, we also report the range of effect sizes pooled to better assess the heterogeneity in the results. AIDS: Acquired immune deficiency syndrome; COPD: Chronic obstructive pulmonary disease; DM: Diabetes Mellitus; HBV: Hepatitis B Virus; HCV: Hepatitis C Virus; HIV: Human immunodeficiency virus; TB: Tuberculosis.

### Supplementary Table 5: Risk ratios of treatment outcomes in each cluster

| Cluster | n | Died | Cured | LFU |
|---|---|---|---|---|
| TB+AIDS + DM +Chronic lung disease (COPD) | 1 | RR 10.79 (10.42 - 11.16) | | |
| TB+AIDS + DM +Mental disorder | 23 | RR 2.81 (1.41 - 5.6) | RR .62 (0.41 - .96) | RR 1.6 (0.74 - 3.49) |
| TB+AIDS + Mental disorder | 143 | RR 4 (3.22 - 4.96) | RR .4 (0.31 - .51) | RR 2.32 (1.82 - 2.96) |
| TB+AIDS + Mental disorder +Cancer | 2 | RR 10.79 (10.42 - 11.16) | | |
| TB+AIDS + DM | 110 | RR 4.71 (3.8 - 5.84) | RR .52 (0.42 - .66) | RR 1.07 (0.68 - 1.69) |
| TB+DM + Mental disorder +Arthritis | 1 | | | RR 7.38 (7.18 - 7.59) |
| TB+DM + Mental disorder | 118 | RR 1.92 (1.3 - 2.83) | RR .86 (0.76 - .98) | RR 1.13 (0.74 - 1.72) |
| TB+DM + Chronic lung disease | 13 | RR 7.47 (5.19 - 10.75) | RR .3 (0.11 - .81) | RR .57 (0.09 - 3.73) |
| TB+Mental disorder + Cancer | 6 | RR 7.19 (4.08 - 12.67) | RR .44 (0.14 - 1.35) | |
| TB+DM + Cancer +Heart disease | 1 | RR 10.79 (10.42 - 11.16) | | |
| TB+AIDS + Heart disease | 8 | RR 5.39 (2.69 - 10.79) | RR .33 (0.1 - 1.08) | RR 1.85 (0.56 - 6.13) |
| TB+AIDS + Cancer | 17 | RR 6.34 (4.26 - 9.46) | RR .38 (0.18 - .8) | RR .87 (0.24 - 3.19) |
| TB+DM + Mental disorder +Cancer | 1 | RR 10.79 (10.42 - 11.16) | | |
| TB+DM + Heart disease | 28 | RR 3.08 (1.71 - 5.54) | RR .93 (0.74 - 1.18) | |
| TB+DM+ Heart disease +Chronic lung disease (COPD) | 2 | RR 5.39 (1.35 - 21.57) | RR .65 (0.16 - 2.61) | |
| TB+DM + Cancer | 15 | RR 7.91 (5.82 - 10.75) | RR .35 (0.15 - .81) | |
| TB+Cancer + Heart disease | 3 | RR 7.19 (3.23 - 16.02) | RR .44 (0.09 - 2.16) | |
| TB+Mental disorder + Heart disease | 2 | | RR 1.31 (1.3 - 1.31) | |
| TB+DM+Mental disorder +Chronic lung disease (COPD) | 3 | RR 10.79 (10.42 - 11.16) | | |
| TB+Cancer + Chronic lung disease | 6 | RR 7.19 (4.08 - 12.67) | RR .44 (0.14 - 1.35) | |
| TB+AIDS + Chronic lung disease | 2 | RR 10.79 (10.42 - 11.16) | | |
| TB+Mental disorder + Chronic lung disease | 5 | RR 6.47 (3.16 - 13.25) | RR .52 (0.18 - 1.53) | |
| TB+Mental disorder + Arthritis | 2 | | RR .65 (0.16 - 2.61) | RR 3.69 (0.92 - 14.76) |
| TB+Arthritis + Cancer | 2 | | RR 1.31 (1.3 - 1.31) | |
| TB+Heart disease + Chronic lung disease | 8 | RR 5.39 (2.69 - 10.79) | RR .65 (0.33 - 1.31) | |
| TB+DM + Arthritis | 4 | RR 2.7 (0.49 - 14.73) | RR .65 (0.25 - 1.74) | RR 1.85 (0.34 - 10.08) |
| TB+DM + Mental disorder +Heart disease | 1 | | RR 1.31 (1.3 - 1.31) | |

***Note:*** Risk ratios calculated from individual participant data provided by the authors of one study.(Reis-Santos 2013). n is the number of people with each cluster. The total number of people with TB only is 31,959. AIDS: Acquired immune deficiency syndrome; COPD: Chronic obstructive pulmonary disease; DM: Diabetes Mellitus; HIV: Human immunodeficiency virus; TB: Tuberculosis.

### Supplementary Table 6: Subgroup analyses pooling prevalence of each cluster of tuberculosis + two or more chronic conditions by country's income level, and quality score

| Cluster and subgroups (p-values) ‡ | Prevalence (95%CI)* | Number of studies | range | $I^2$ † |
|---|---|---|---|---|
| **TB+ Depression + Anxiety** | **15.27% (10.70% - 20.47%)** | **3 (1,473)** | **12.78% - 22.09%** | **63.81%** |
| *By country's income level (p=0.59)* | | | | |
| Low | 14.81% (9.8% - 21.78%) | 1 (135) | .. | .. |
| Upper Middle | 13.14% (11.36% - 15.02%) | 2 (1,338) | 12.78% - 22.09% | |
| *By quality score (p=0.06)* | | | | |
| <=2* | 12.78% (11.04% - 14.74%) | 1 (1,252) | .. | .. |
| ≥3* | 17.49% (12.71% - 22.84%) | 2 (221) | 14.81% - 22.09% | .. |
| **TB+HIV + Anxiety** | **15.23% (11.25% - 19.67%)** | **4 (1,413)** | **10.07% - 16.40%** | **77.37%** |
| *By country's income level (p=0.08)* | | | | |
| Low | 14.08% (10.21% - 18.45%) | 3 (1,327) | 10.07% - 16.40% | .. |
| Upper Middle | 22.09% (14.62% - 31.95%) | 1 (86) | .. | .. |
| **TB+HIV + PTSD** | **14.84% (13.9%-15.8%)** | **2 (5,400)** | **2.40% - 16.63%** | **..** |
| *By country's income level (p<0.005)* | | | | |
| Low | 2.4% (1.38% - 4.15%) | 1 (500) | .. | .. |
| Upper Middle | 16.63% (15.62% - 17.7%) | 1 (4900) | .. | .. |
| **TB+HIV + HBV** | **9.70% (2.37%-21.04%)** | **3 (792)** | **2.21% - 20.57%** | **94.81%** |
| *By country's income level: all were Upper Middle* | | | | |
| **TB+HIV + Depression** | **9.66% (6.96% - 12.74%)** | **9 (3,418)** | **5.06% - 16.28%** | **87.47%** |
| *By country's income level (p=0.06)* | | | | |
| Low | 9.17% (6.48% - 12.26%) | 8 (3,332) | 5.06% - 17.07% | 88.12% |
| Upper Middle | 16.28% (9.95% - 25.49%) | 1 (86) | **..** | **..** |
| **TB+HIV + Alcoholism** | **9.29% (4.98%-14.72%)** | **4 (6,963)** | **5.00%-16.55%** | **96.34%** |
| *By country's income level (p<0.005)* | | | | |
| Lower Middle | 5% (4.06% - 6.14%) | 1 (1,699) | .. | .. |
| Upper Middle | 11.54% (7.5% - 16.29%) | 3 (5,264) | 5.81% - 16.55% | .. |
| **TB+DM + HCV** | **7.41% (4.99%-10.24%)** | **2 (395)** | **2.19%-11.24%** | **..** |
| *By country's income level: all were Lower Middle* | | | | |
| *By quality score (p<0.005)* | | | | |
| <=2* | 11.24% (7.94% - 15.68%) | 1 (258) | .. | .. |
| ≥3* | 2.19% (0.75% - 6.24%) | 1 (137) | .. | .. |
| **TB+HIV + Hypothyroidism** | **5.37% (0.90% - 12.94%)** | **3 (1,108)** | **2.30% - 10.84%** | **94.00%** |
| *By country's income level (p<0.005)* | | | | |
| Lower Middle | 2.3% (1.35% - 3.9%) | 1 (565) | .. | .. |
| Upper Middle | 9.49% (7.13% - 12.14%) | 2 (543) | 4.40% - 10.84% | .. |
| **TB+HIV + Drug use** | **5.21% (2.73% - 8.39%)** | **4 (2,584)** | **2.74% - 5.87%** | **86.19%** |
| *By country's income level (p=0.15)* | | | | |
| Lower Middle | 3.35% (2.6% - 4.32%) | 1 (1,699) | .. | .. |
| Upper Middle | 6.13% (2.45% - 11.26%) | 3 (885) | 2.74% - 8.63% | .. |
| **TB+HBV + Drug use** | **3.44% (0% - 12.05%)** | **3 (763)** | **0% - 9.78%** | **95.20%** |

| | | | | |
|---|---|---|---|---|
| *By country's income level (p<0.005)* | | | | |
| Lower Middle | 0% (0% - 1.88%) | 1 (200) | .. | .. |
| Upper Middle | 7.59% (5.52% - 9.95%) | 2 (563) | 4.39% - 9.78% | .. |
| *By quality score (p<0.005)* | | | | |
| <=2* | 0% (0% - 1.88%) | 1 (200) | .. | .. |
| ≥3* | 7.59% (5.52% - 9.95%) | 2 (563) | 4.39% - 9.78% | .. |
| **TB+Depression + Chronic lung disease** | **1.73% (0.65% - 3.24%)** | **2 (437)** | **1.15% - 2.86%** | **..** |
| *By country's income level (p=0.20)* | | | | |
| Low | 2.86% (1.23% - 6.51%) | 1 (175) | .. | .. |
| Lower Middle | 1.15% (0.39% - 3.31%) | 1 (262) | .. | .. |
| **TB+HBV + HCV** | **1.56% (0.60%-2.90%)** | **2 (500)** | **1.00%-2.00%** | **..** |
| *By country's income level: all were Lower Middle* | | | | |
| **TB+HIV + HCV** | **1.47% (0.36% - 3.21%)** | **6 (1,550)** | **0.33% - 4.98%** | **79.36%** |
| *By country's income level (p<0.005)* | | | | |
| Low | 0.37% (0.06% - 2.05%) | 1 (272) | .. | .. |
| Lower Middle | 0.99% (0.1% - 2.55%) | 3 (695) | 0.33% - 2.33% | .. |
| Upper Middle | 3.76% (2.33% - 5.5%) | 2 (583) | 1.66% - 4.98% | .. |
| *By quality score (p=0.21)* | | | | |
| <=2* | 0.82% (0.05% - 2.25%) | 3 (830) | 0.33% - 2.33% | .. |
| ≥3* | 2.36% (0.46% - 5.45%) | 3 (720) | 0.73% - 4.98% | .. |
| **TB+DM + Drug use** | **1.03% (0.22% - 2.34%)** | **5 (7,476)** | **0.17% - 6.76%** | **90.85%** |
| *By country's income level (p=0.12)* | | | | |
| Lower Middle | 0.33% (0.09% - 1.18%) | 1 (614) | .. | .. |
| Upper Middle | 1.36% (0.22% - 3.31%) | 4 (6,862) | 0.18% - 6.76% | .. |
| **TB+HIV + DM** | **0.98% (0.70% - 1.29%)** | **40 (526,827)** | **0% - 8.33%** | **97.79%** |
| *By country's income level (p<0.005)* | | | | |
| Low | 3.22% (1.44% - 5.61%) | 9 (4,554) | 0.44% - 8.33% | 92.11% |
| Lower Middle | 0.46% (0.18% - 0.85%) | 15 (13,271) | 0% - 3.67% | 84.35% |
| Upper Middle | 0.87% (0.48% - 1.34%) | 16 (509,002) | 0% - 7.49% | 98.91% |
| *By quality score (p=0.01)* | | | | |
| <=2* | 3.67% (1.94% - 6.83%) | 1 (245) | .. | .. |
| ≥3* | 0.94% (0.67% - 1.25%) | 39 (526,582) | 0% - 8.05% | 97.83% |
| **TB+DM + Alcoholism** | **0.69% (0.47% - 0.94%)** | **2 (5,218)** | **0.63% - 0.76%** | **..** |
| *By country's income level (p=0.98)* | | | | |
| Lower Middle | 0.63% (0.17% - 2.26%) | 1 (318) | .. | **..** |
| Upper Middle | 0.76% (0.55% - 1.04%) | 1 (4,900) | .. | **..** |
| **TB+DM + Chronic kidney disease** | **0.50% (0.20% – 0.89%)** | **2 (2,275)** | **0.55% - 12.22%** | **..** |
| *By country's income level (p<0.005)* | | | | |
| Lower Middle | 0.55% (0.31% - 0.96%) | 1 (2,185) | .. | **..** |
| Upper Middle | 12.22% (6.96% - 20.57%) | 1 (90) | .. | **..** |
| **TB+HIV + Chronic liver disease** | **0.48% (0.03% - 1.27%)** | **2 (682)** | **0.50% - 2.33%** | **..** |
| *By country's income level (p=0.11)* | | | | |

| | | | | |
|---|---|---|---|---|
| Lower Middle | 0.5% (0.17% - 1.47%) | 1 (596) | .. | .. |
| Upper Middle | 2.33% (0.64% - 8.09%) | 1 (86) | .. | .. |
| **TB+DM + Chronic lung disease** | **0.23% (0%-0.98%)** | **3 (45,143)** | **0.03%-1.57%** | **96.08%** |
| *By country's income level (p=0.01)* | | | | |
| Lower Middle | 1.57% (0.43% - 5.56%) | 1 (127) | | |
| Upper Middle | 0.05% (0.03% - 0.08%) | 2 (45,016) | 0.03% – 0.45% | |
| **TB+HIV + Chronic lung disease** | **0.10% (0% - 0.64%)** | **2 (682)** | **0.17% - 1.16%** | **..** |
| *By country's income level (p=0.16)* | | | | |
| Lower Middle | 0.17% (0.03% - 0.94%) | 1 (596) | .. | .. |
| Upper Middle | 1.16% (0.21% - 6.3%) | 1 (86) | .. | .. |
| **TB+DM + Heart disease** | **0.05% (0.02% - 0.07%)** | **2 (40,495)** | **0.07% - 2.28%** | **..** |
| *By country's income level (p<0.005)* | | | | |
| Lower Middle | 2.28% (1.36% - 3.79%) | 1 (614) | .. | .. |
| Upper Middle | 0.07% (0.05% - 0.1%) | 1 (39,881) | .. | .. |
| **TB+DM + Chronic liver disease** | **0% (0% - 0%)** | **2 (181,485)** | **0% - 0.03%** | **..** |
| *By country's income level: all were Upper Middle* | | | | |

*Note.* Subgroups by quality classification were only reported when significant differences were found. We were unable to do subgroup analyses by gender or age, given the lack of data reported separately for each gender or different age groups. See supplementary Figures 1-22 for subgroups by country. DM: Diabetes Mellitus; HBV: Hepatitis B Virus; HCV: Hepatitis C Virus; HIV: Human immunodeficiency virus; PTSD: Post-traumatic stress disorder; TB: Tuberculosis.

[*] Pooled prevalence and 95% confidence intervals from random-effects meta-analysis when the number of studies is >1.

[†] I-squared values reported only for meta-analyses with >2 studies

[‡] P-values from the test for heterogeneity between sub-groups

### Supplementary Table 7: Quality assessment for each study

| Lead Author, year | 1) | 2) | 3) | 4) | 5) | 5b) | 6) | Total score |
|---|---|---|---|---|---|---|---|---|
| Abdelsalam 2013 | - | - | - | NA | ** | NA | NA | 2 ** |
| Achanta 2013 | * | * | * | NA | ** | NA | NA | 5 ***** |
| Adejumo 2020 | * | - | * | NA | ** | NA | NA | 4 **** |
| Adem 2014 | * | - | * | NA | * | NA | NA | 3 *** |
| Aires 2012 | * | - | * | NA | ** | NA | NA | 5 ***** |
| Alavi 2012 | * | - | * | NA | ** | NA | NA | 5 ***** |
| Alinaitwe 2018 | * | * | - | NA | * | NA | NA | 3 *** |
| Ambaw 2017 | * | - | * | NA | * | NA | NA | 3 *** |
| Appana 2016 | * | - | - | NA | ** | NA | NA | 3 *** |
| Azeez 2018 | * | - | - | NA | ** | * | NA | 3 ***(*) |
| Azovtzeva 2019 | - | - | - | NA | ** | - | NA | 2 ** |
| Blal 2005 | * | - | - | NA | ** | NA | NA | 3 *** |
| Boillat-Blanco 2016 | * | * | - | NA | ** | NA | NA | 4 **** |
| Carvalho 2002 | * | - | * | NA | * | NA | NA | 3 *** |
| Castro 2019 | * | - | - | NA | - | * | NA | 1 *(*) |
| Cespedes 2019 | * | - | * | NA | ** | NA | NA | 4 **** |
| Chin 2019 | - | - | - | NA | * | NA | NA | 1 * |
| Damtew 2014 | * | * | - | NA | ** | NA | NA | 4 **** |
| Dasa 2019 | * | * | * | NA | * | NA | NA | 4 **** |
| Dave 2013 | * | * | * | NA | ** | NA | NA | 5 ***** |
| de Oliveira 2013 | * | - | * | NA | ** | NA | NA | 3 *** |
| Delgado-Sánchez 2015 | * | * | * | NA | ** | NA | NA | 5 ***** |
| Deribew 2009 | * | * | * | NA | * | NA | NA | 4 **** |
| do Prado 2014 | * | * | - | NA | ** | NA | NA | 4 **** |
| Dos Santos 2017 | * | * | - | NA | * | NA | NA | 3 *** |
| Duko 2015 | * | * | * | NA | - | NA | NA | 3 *** |
| Echazarreta 2018 | * | - | - | NA | ** | NA | NA | 3 *** |
| Ekeke 2017 | * | * | * | NA | ** | NA | NA | 5 ***** |
| Faurholt-Jepsen 2012 | * | - | - | NA | ** | NA | NA | 3 *** |
| Getachew 2014 | * | * | - | NA | ** | NA | NA | 4 **** |
| Gupta 2011 | * | - | - | NA | ** | NA | NA | 3 *** |
| Heredia 2011 | * | - | * | NA | ** | NA | NA | 4 **** |
| Jalal 2017 | * | * | * | NA | ** | - | NA | 5 ***** |
| Javed 2018 | - | - | - | NA | ** | NA | NA | 2 ** |
| Jimenez-Corona 2013 | * | - | - | NA | ** | NA | NA | 3 *** |
| Kelly 2016 | * | * | - | NA | ** | NA | NA | 4 **** |
| Khan 2019 | * | * | - | NA | ** | - | NA | 4 **** |
| Khanna 2013 | * | * | - | NA | ** | NA | NA | 4 **** |
| Kibirige 2013 | * | * | - | NA | ** | NA | NA | 4 **** |
| Krupitsky 2006 | * | - | * | NA | ** | NA | NA | 4 **** |
| Kuniholm 2008 | - | - | - | NA | ** | NA | NA | 2 ** |
| KV 2013 | * | * | * | NA | ** | NA | NA | 5 ***** |
| Magee 2015 | * | - | * | NA | ** | NA | NA | 4 **** |
| Manosuthi 2012 | * | - | - | NA | ** | NA | NA | 3 *** |
| Modongo 2012 | * | * | - | NA | ** | NA | NA | 4 **** |
| Mohammed-hussein 2020 | * | * | * | NA | * | NA | NA | 4 **** |
| Molla 2019 | * | * | * | NA | * | NA | NA | 4 **** |
| Mukhtar 2018 | * | * | - | NA | * | NA | NA | 3 *** |
| Munoz-Torrico 2017 | * | - | * | NA | ** | NA | NA | 4 **** |
| Munseri 2019 | * | * | * | NA | ** | NA | NA | 5 ***** |
| Naik 2013 | * | * | - | NA | ** | NA | NA | 4 **** |
| Oni 2015 | * | * | - | NA | ** | NA | NA | 4 **** |
| Oni 2017 | * | * | * | ** | ** | NA | NA | 7 ******* |
| Owiti 2017 | * | - | - | NA | ** | NA | NA | 3 *** |
| Pande 2018 | * | * | * | NA | ** | NA | NA | 5 ***** |
| Pando 2008 | * | - | - | NA | ** | NA | NA | 3 *** |
| Pawar 2018 | * | * | - | NA | ** | NA | NA | 4 **** |
| Peltzer 2012 | * | - | * | NA | * | NA | NA | 3 *** |
| Peltzer 2013 | * | - | * | NA | * | NA | NA | 3 *** |
| Pepper 2010 | * | - | - | NA | - | NA | NA | 1 * |
| Piparva 2017 | * | - | - | NA | ** | - | NA | 3 *** |
| Reis 2011 | * | * | * | NA | ** | NA | NA | 5 ***** |

| | | | | | | | | |
|---|---|---|---|---|---|---|---|---|
| Reis-Santos 2013 | * | - | * | NA | ** | - | NA | 4 **** |
| Richards 2006 | - | - | - | NA | ** | NA | NA | 2 ** |
| Sharman 2019 | * | * | - | NA | ** | NA | NA | 4 **** |
| Shokri 2018 | - | - | - | NA | ** | NA | NA | 2 ** |
| Shyamala 2018 | * | - | - | NA | * | NA | NA | 2 ** |
| Sirinak 2008 | - | - | - | NA | ** | * | NA | 3 ***(*) |
| Tabarsi 2014 | * | * | - | NA | ** | NA | NA | 4 **** |
| Ugarte-Gil 2020 | * | * | - | NA | ** | NA | NA | 4 **** |
| van den Heuvel 2013 | * | * | * | NA | * | NA | NA | 4 **** |
| Vashakidze 2017 | * | * | - | NA | ** | NA | NA | 4 **** |
| Velásquez 2015 | * | * | - | NA | ** | NA | NA | 4 **** |
| Viswanathan 2014 | - | - | - | NA | ** | NA | NA | 2 ** |
| Walker 2019 | * | * | - | NA | * | NA | NA | 3 *** |
| Wang 2018 | * | - | - | NA | * | NA | NA | 2 ** |
| Weimann 2016 | * | - | * | NA | * | NA | NA | 3 *** |
| White 2020 | * | * | - | NA | ** | NA | NA | 4 **** |
| Whitehouse 2019 | * | * | - | NA | ** | NA | NA | 4 **** |
| Workneh 2016 | * | * | * | NA | ** | NA | NA | 5 ***** |
| Wu 2016 | - | - | - | NA | ** | NA | NA | 2 ** |
| Yaneth-Giovanetti 2019 | * | - | - | NA | ** | NA | NA | 3 *** |

1) Representativeness of the sample: *: Truly (all subjects or random sampling) or somewhat (non-random sampling) representative of the average in the target population. -: Selected group of users or no description of the sampling strategy.
2) Sample size: *: Justified and satisfactory; -: Not justified.
3) Non-respondents: *: Comparability between respondents and non-respondents characteristics is established, or the response rate is satisfactory (>80%); -: The response rate is unsatisfactory or not reported, or the comparability between respondents and non-respondents is unsatisfactory or not reported.
4) Comparability: **: Study controls, either in the design (e.g. matching) or in the analyses (adjusted results), for two or more potential confounders (e.g. age, gender, deprivation, severity, TB type, severity of disease); NA: Not applicable (no two groups are compared).
5) Assessment of the outcome of interest: **: Record linkage or independent blind assessment; *: Validation scale or self report; -: No description.
5b) Appropriate length of follow-up: *: Yes (≥ 6 months); -: Not reported; NA: Not applicable
6) Statistical test: NA: Not applicable (raw data extracted).

### Supplementary Table 8: Post-hoc (unplanned) subgroup analyses grouping similar exposure definitions

| Cluster and subgroups (p-values)‡ | Prevalence (95%CI)* | Number of studies | range | $I^2$ † |
|---|---|---|---|---|
| **TB+Depression + Anxiety** | **15.27% (10.70% - 20.47%)** | **3 (1,473)** | **12.78% - 22.09%** | **63.81%** |
| *By definition of anxiety (p=0.59)* | | | | |
| HADS anxiety subscale | 13.14% (11.36% - 15.02%) | 2 (1,338) | 12.78% - 22.09% | - |
| HSCL-25 | 14.81% (9.8% - 21.78%) | 1 (135) | - | - |
| *By definition of depression (p=0.59)* | | | | |
| HADS depression subscale | 13.14% (11.36% - 15.02%) | 2 (1,338) | 12.78% - 22.09% | - |
| HSCL-25 | 14.81% (9.8% - 21.78%) | 1 (135) | - | - |
| **TB+HIV + Anxiety** | **15.23% (11.25% - 19.67%)** | **4 (1,413)** | **10.07% - 16.40%** | **77.37%** |
| *By definition of anxiety (p=0.73)* | | | | |
| Any anxiety disorder (MINI) | 16.4% (13.41% - 19.9%) | 1 (500) | - | - |
| HADS anxiety subscale | 15.04% (9.44% - 21.66%) | 3 (913) | 10.07% - 22.09% | - |
| **TB+HIV + Depression** | **9.66% (6.96% - 12.74%)** | **9 (3,418)** | **5.06% - 16.28%** | **87.47%** |
| *By definition of depression (p<0.005)* | | | | |
| PHQ-9 | 6.01% (4.84% - 7.29%) | 3 (1,475) | 5.06% - 6.95% | - |
| Kessler-10 scale | 12.61% (8.87% - 17.62%) | 1 (222) | - | - |
| HADS depression subscale | 13.29% (7.13% - 20.96%) | 3 (913) | 8.15% - 17.07% | - |
| MINI | 9.36% (7.43% - 11.47%) | 2 (808) | 6.80% - 14.29% | - |
| **TB+HIV + Alcoholism** | **9.29% (4.98%-14.72%)** | **4 (6,963)** | **5.00%-16.55%** | **96.34%** |
| *By definition of Alcoholism (p<0.005)* | | | | |
| CAGE alcohol screening test | 5.81% (2.51% - 12.9%) | 1 (86) | 5.81 | |
| alcohol dependence/addiction | 16.55% (12.64% - 21.36%) | 1 (278) | 16.55 | |
| alcohol use disorders identification test (AUDIT) | 11.27% (10.41% - 12.18%) | 1 (4,900) | - | - |
| alcohol use that interfered with family, health, or work | 5% (4.06% - 6.14%) | 1 (1,699) | 5 | |
| **TB+HIV + Drug use** | **5.21% (2.73% - 8.39%)** | **4 (2,584)** | **2.74% - 5.87%** | **86.19%** |
| *By definition of drug use (p=0.09)* | | | | |
| Use of illicit drugs | 6.31% (2.65% - 11.35%) | 3 (2,182) | 3.35% - 8.63% | - |
| Intravenous drug use | 2.74% (1.53% - 4.83%) | 1 (402) | - | - |
| **TB+HBV + Drug use** | **3.44% (0% - 12.05%)** | **3 (763)** | **0% - 9.78%** | **95.20%** |
| *By definition of drug use (p<0.005)* | | | | |
| Intravenous drug use | 0% (0% - 1.88%) | 1 (200) | - | - |
| Use of illicit drugs | 7.59% (5.52% - 9.95%) | 2 (563) | 4.39% - 9.78% | - |
| **TB+Depression + Chronic lung disease** | **1.73% (0.65% - 3.24%)** | **2 (437)** | **1.15% - 2.86%** | **-** |
| *By definition of chronic lung disease (p=0.20)* | | | | |
| Chronic lung disease | 2.86% (1.23% - 6.51%) | 1 (175) | - | - |
| Chronic obstructive pulmonary disease (COPD) | 1.15% (0.39% - 3.31%) | 1 (262) | - | - |
| *By definition of depression (p=0.20)* | | | | |
| CES-D | 2.86% (1.23% - 6.51%) | 1 (175) | - | - |
| PHQ-9 | 1.15% (0.39% - 3.31%) | 1 (262) | - | - |
| **TB+DM + Drug use** | **1.03% (0.22% - 2.34%)** | **5 (7,476)** | **0.17% - 6.76%** | **90.85%** |
| *By definition of drug use (p<0.005)* | | | | |

| | | | | |
|---|---|---|---|---|
| Use of illicit drugs | 0.68% (0.27% - 1.22%) | 2 (1,410) | 0.48% - 6.76% | - |
| Psychoactive drug addiction | 0.18% (0.09% - 0.33%) | 1 (5,135) | - | - |
| Opium use | 2.21% (1.07% - 4.49%) | 1 (317) | - | - |
| Drug abuse | 0.33% (0.09% - 1.18%) | 1 (614) | - | - |
| **TB+DM + Alcoholism** | **0.69% (0.47% - 0.94%)** | **2 (5,218)** | **0.63% - 0.76%** | **-** |
| *By type of TB (p=0.98)* | | | | |
| alcohol use disorders identification test (AUDIT) | 0.76% (0.55% - 1.04%) | 1 (4,900) | - | - |
| ≥ 5 drinks per day, >=3 days/week | 0.63% (0.17% - 2.26%) | 1 (318) | - | - |
| **TB+DM + Chronic lung disease** | **0.23% (0%-0.98%)** | **3 (45,143)** | **0.03%-1.57%** | **96.08%** |
| *By definition of chronic lung disease: all were chronic obstructive pulmonary disease (COPD)* | | | | |
| **TB+HIV + Chronic lung disease** | **0.10% (0% - 0.64%)** | **2 (682)** | **0.17% - 1.16%** | **-** |
| *By definition of chronic lung disease (p=0.16)* | | | | |
| Chronic lung disease | 0.17% (0.03% - 0.94%) | 1 (596) | - | - |
| Chronic obstructive pulmonary disease (COPD) | 1.16% (0.21% - 6.3%) | 1 (86) | - | - |

*Note.* DM: Diabetes Mellitus; HBV: Hepatitis B Virus; HCV: Hepatitis C Virus; HIV: Human immunodeficiency virus; PTSD: Post-traumatic stress disorder; TB: Tuberculosis.

[*] Pooled prevalence and 95% confidence intervals from random-effects meta-analysis when the number of studies is >1.

[†] I-squared values reported only for meta-analyses with >2 studies

[‡] P-values from the test for heterogeneity between sub-groups

**Supplementary Figure 1a: Subgroup analyses pooling prevalence of TB+HIV+DM by country (India, Ethiopia, Georgia, Brazil)**

Due to the high number of countries for which there were data, results were split in three figures for clarity

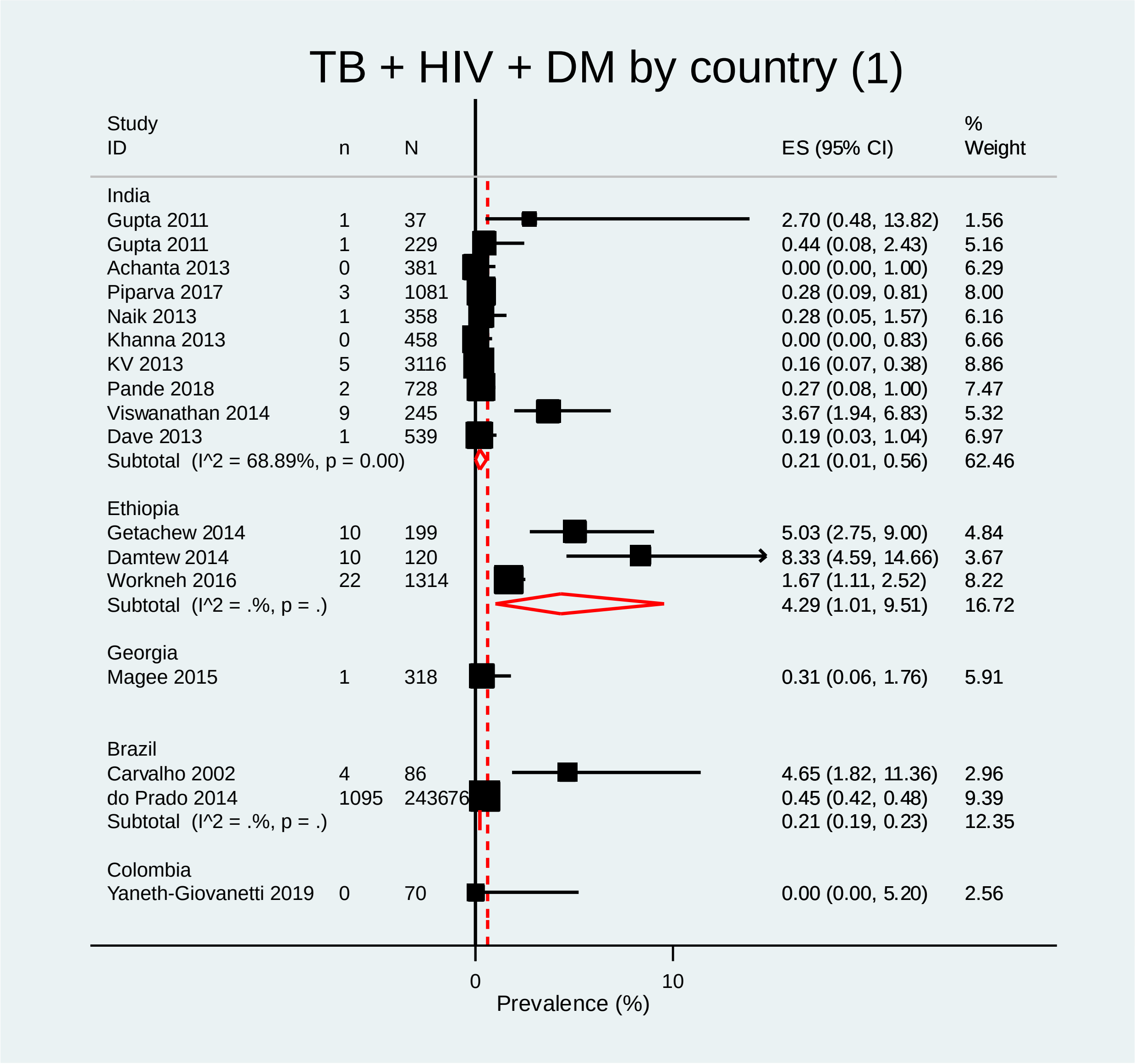

**Supplementary Figure 1b: Subgroup analyses pooling prevalence of TB+HIV+DM by country (Nigeria, Kenya, Peru, Malaysia, Mexico, Paraguay, Iran)**

Due to the high number of countries for which there were data, results were split in three figures for clarity

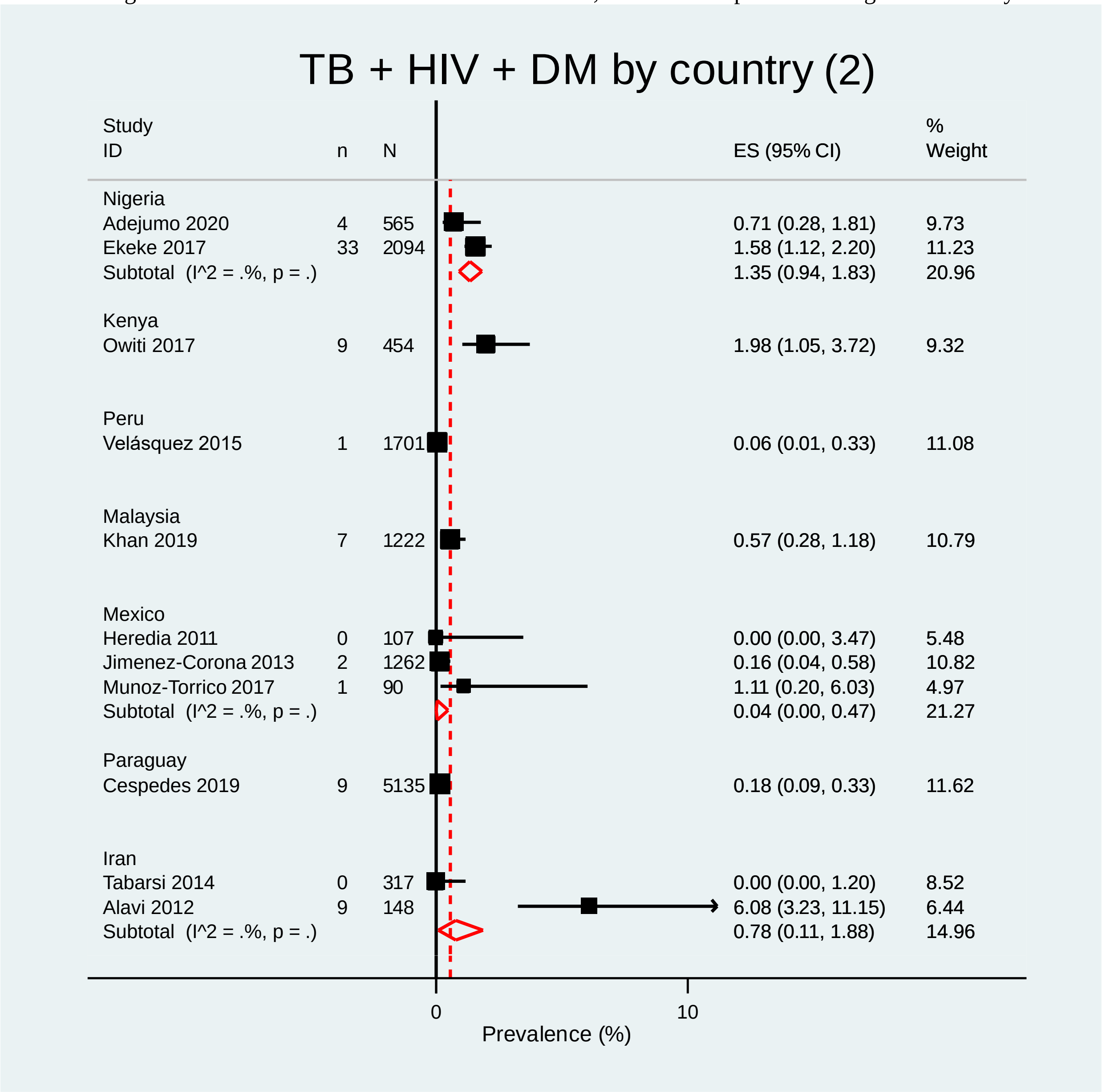

**Supplementary Figure 1c: Subgroup analyses pooling prevalence of TB+HIV+DM by country (Tanzania, Uganda, Thailand, Philippines, South Africa)**
Due to the high number of countries for which there were data, results were split in three figures for clarity

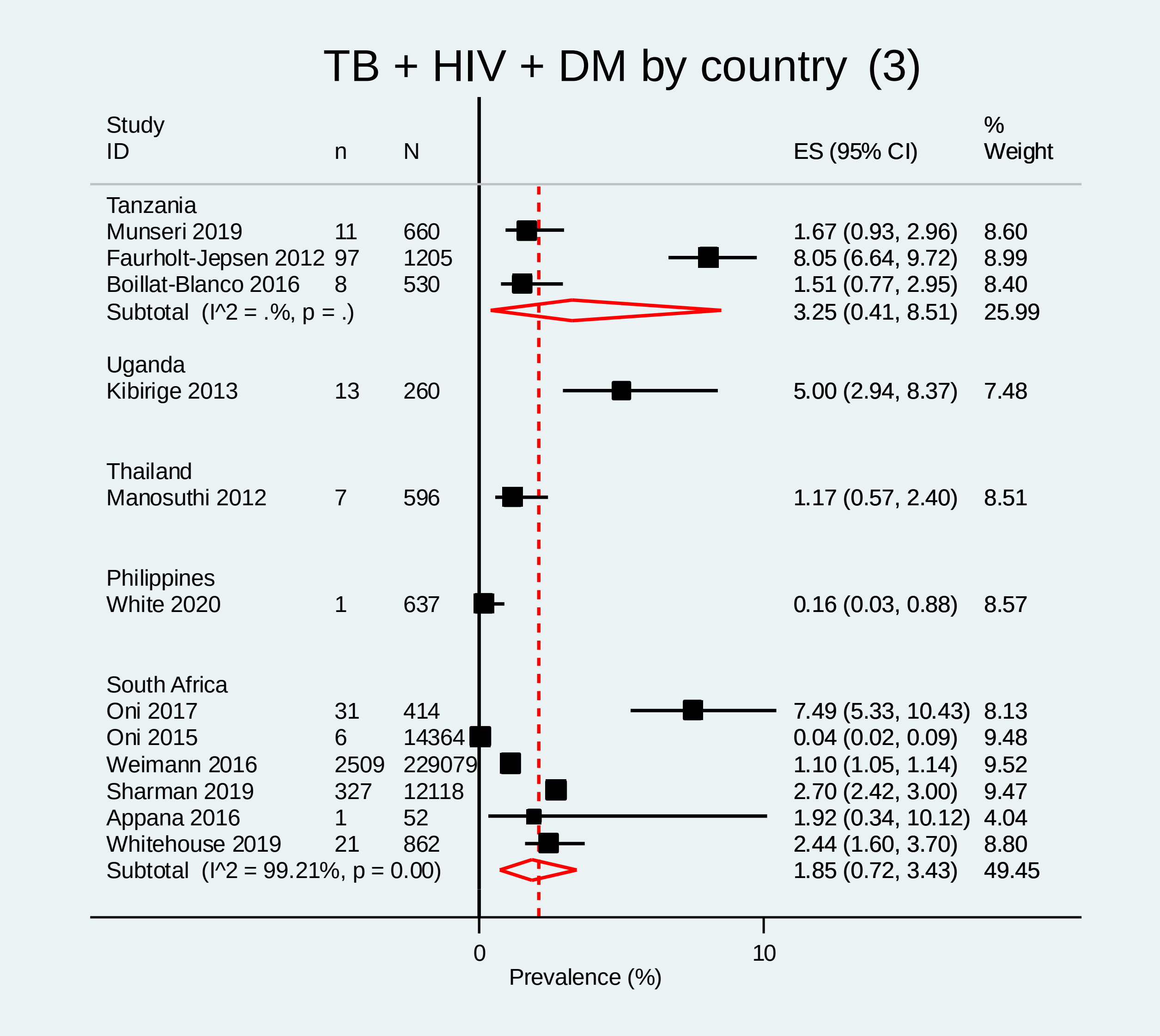

**Supplementary Figure 2: Subgroup analyses pooling prevalence of TB+HIV+Depression by country**

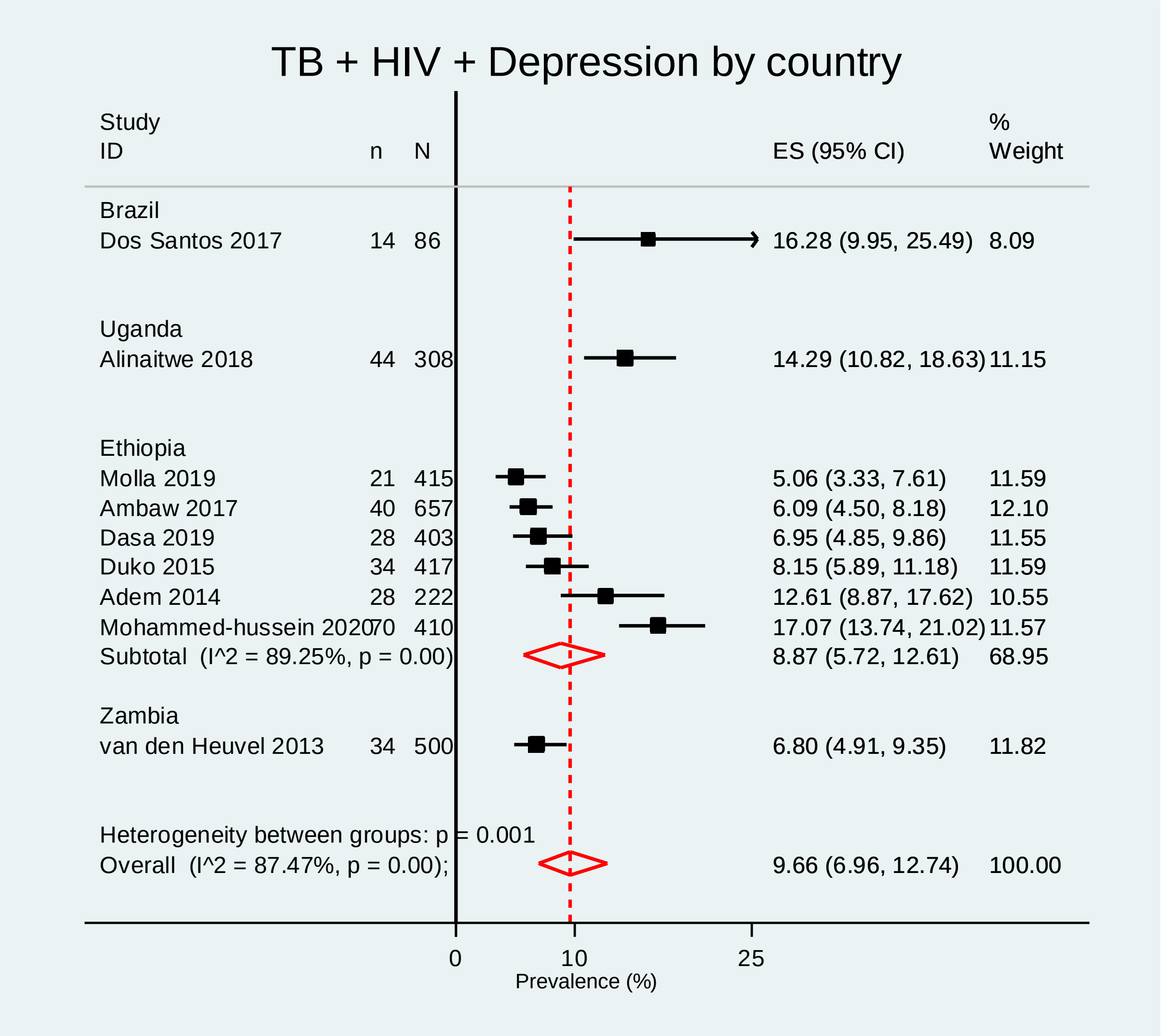

**Supplementary Figure 3: Subgroup analyses pooling prevalence of TB+HIV+HCV by country**

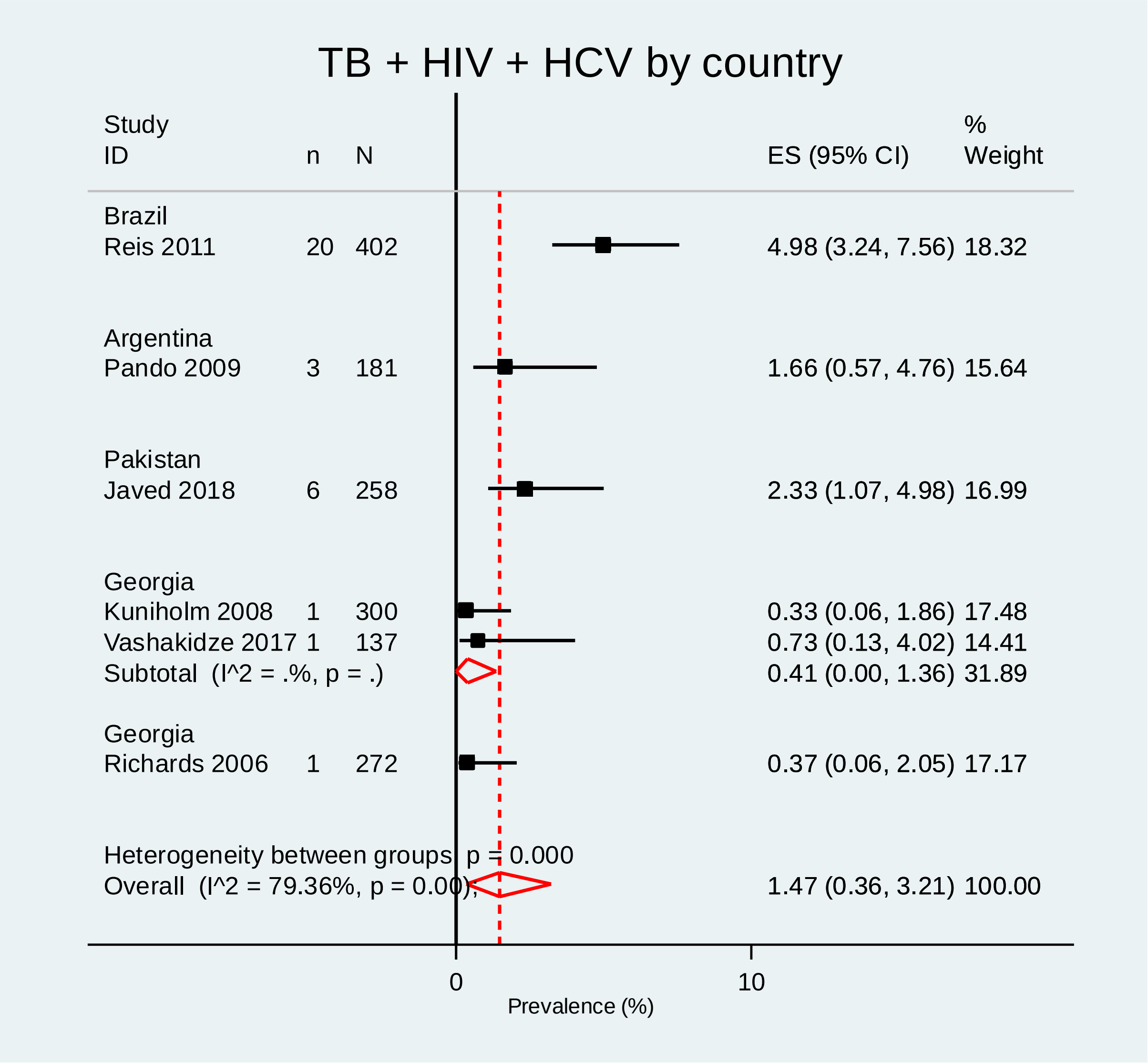

**Supplementary Figure 4: Subgroup analyses pooling prevalence of TB+HIV+Drug use by country**

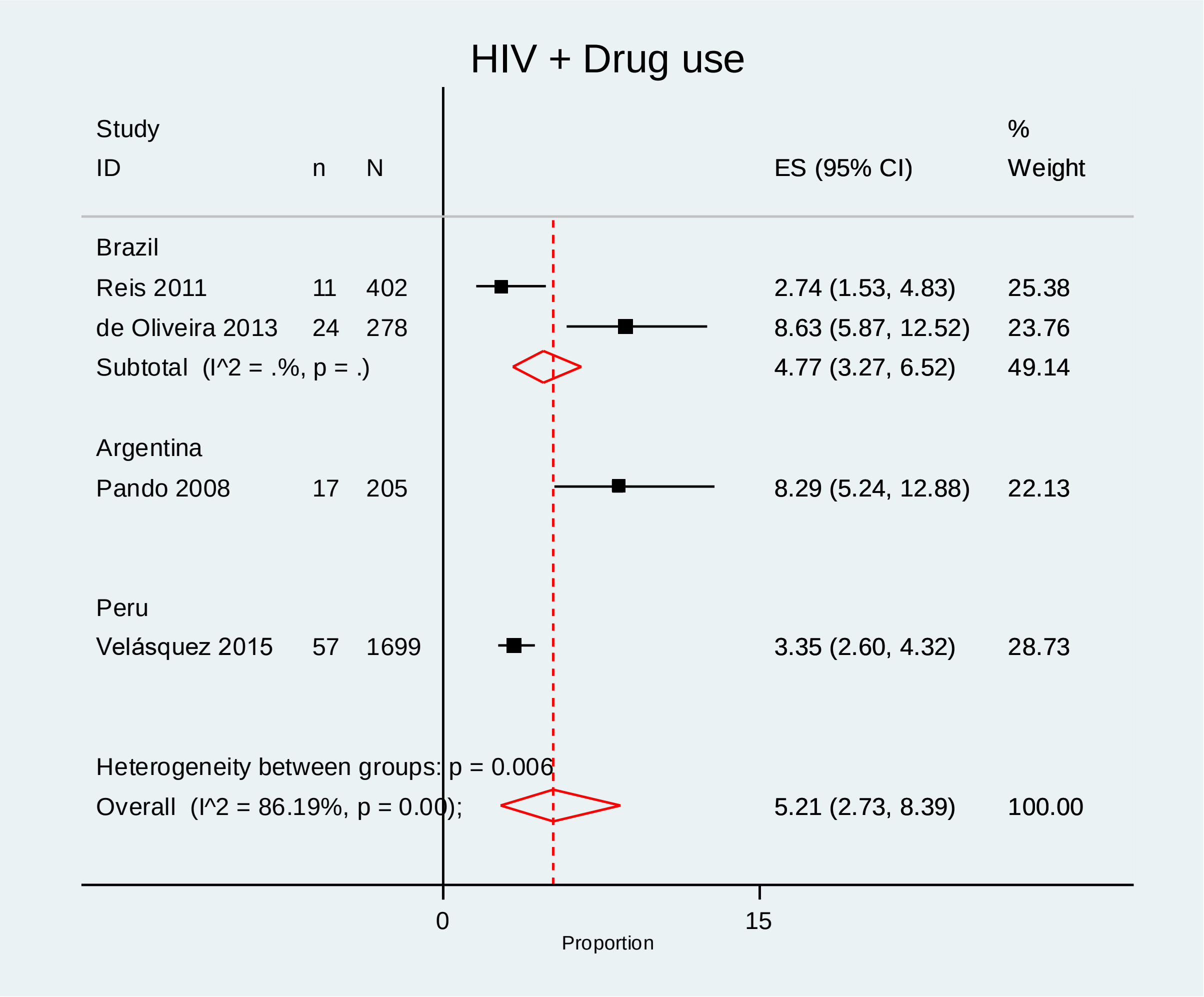

**Supplementary Figure 5: Subgroup analyses pooling prevalence of TB+DM+Drug use by country**

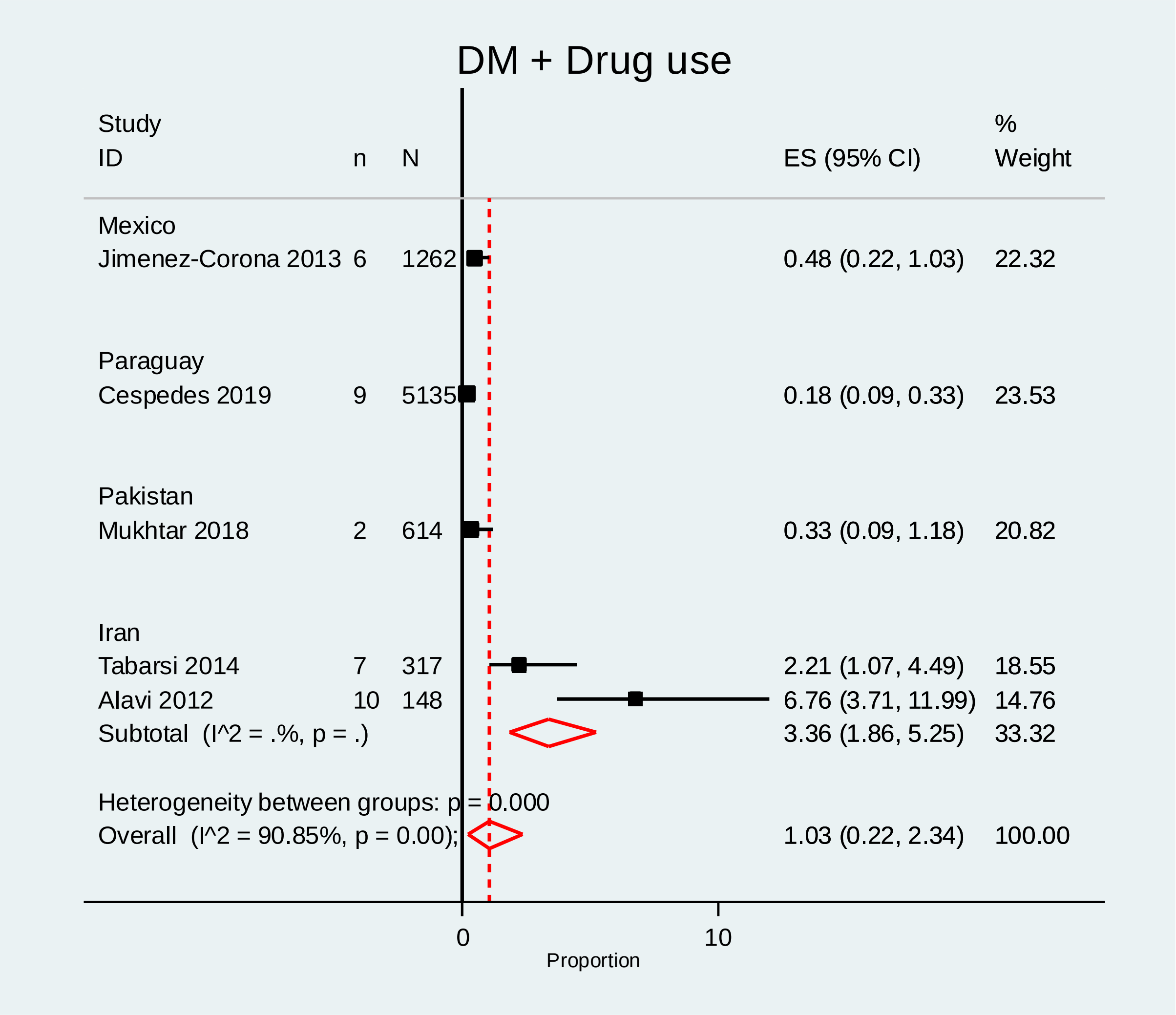

**Supplementary Figure 6: Subgroup analyses pooling prevalence of TB+HIV+Anxiety by country**

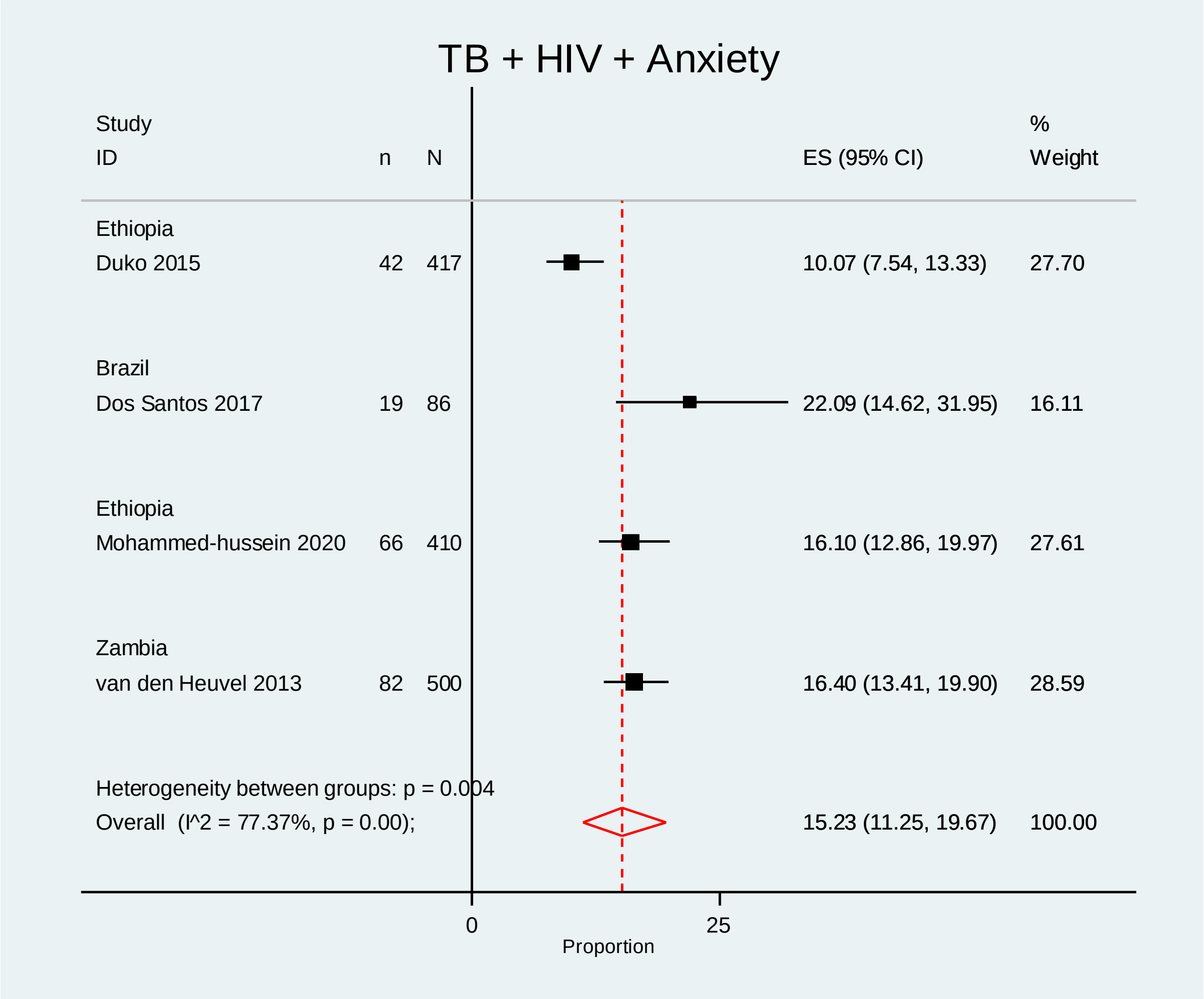

**Supplementary Figure 7: Subgroup analyses pooling prevalence of TB+HIV+Alcoholism by country**

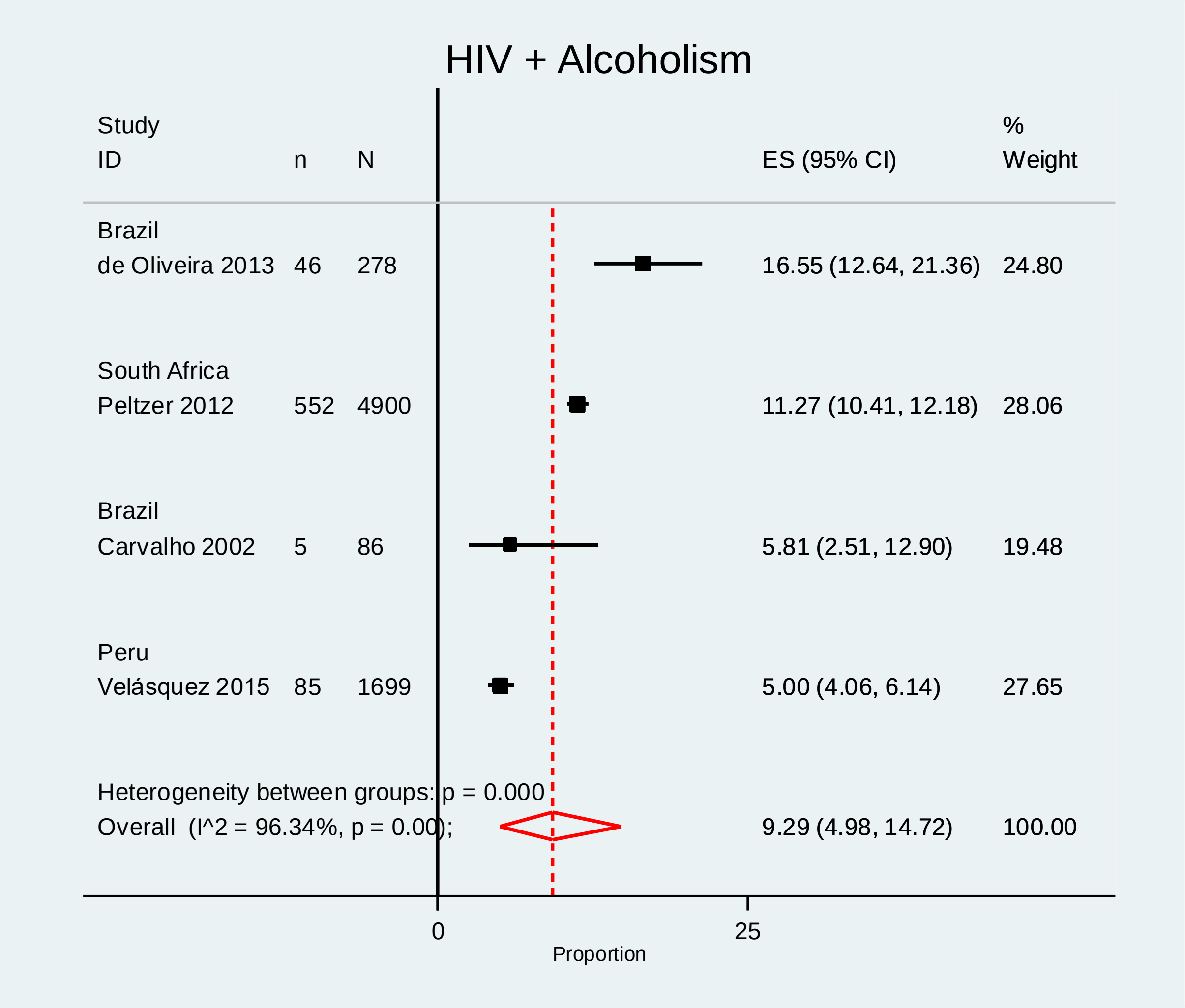

**Supplementary Figure 8: Subgroup analyses pooling prevalence of TB+HIV+HBV by country**

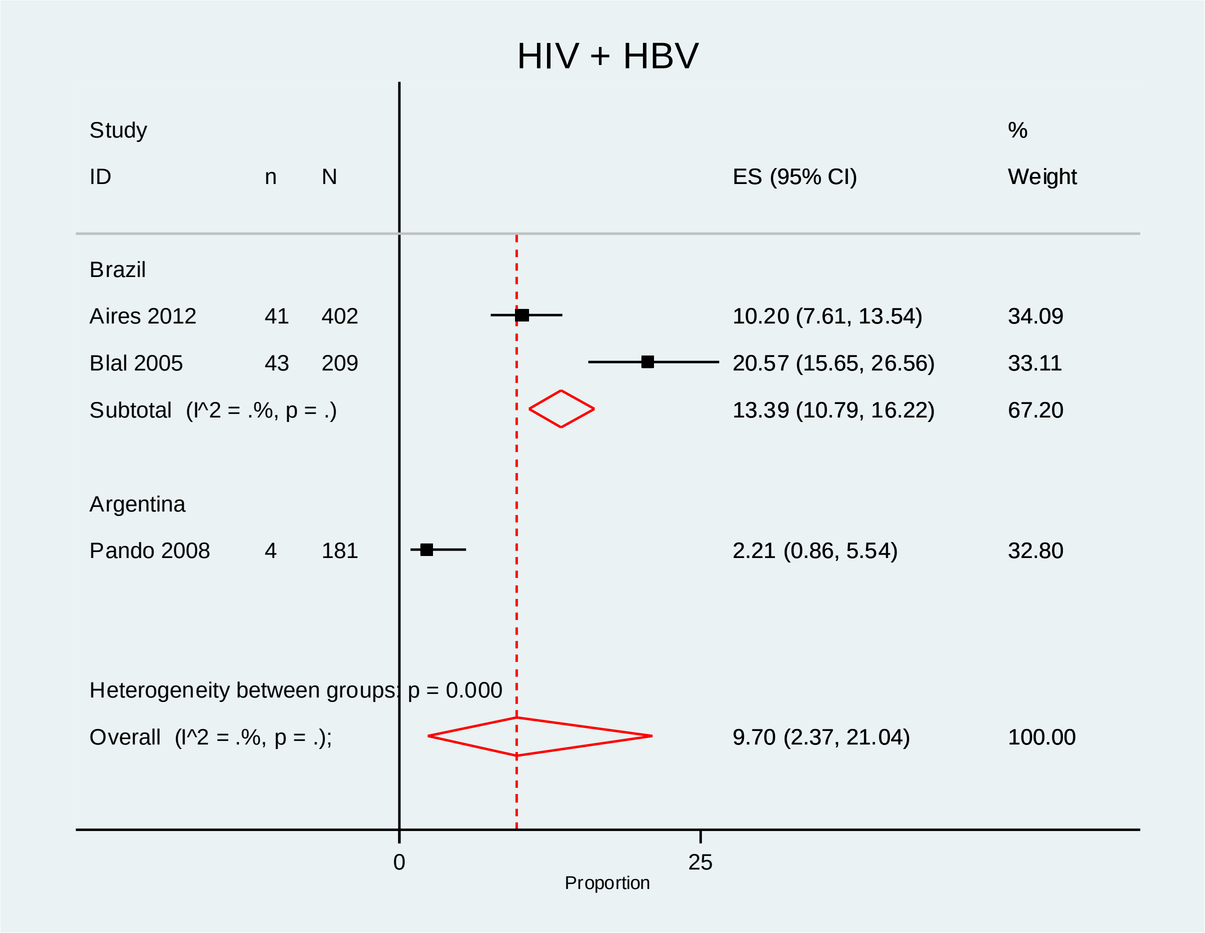

**Supplementary Figure 9: Subgroup analyses pooling prevalence of TB+DM+Chronic lung disease by country**

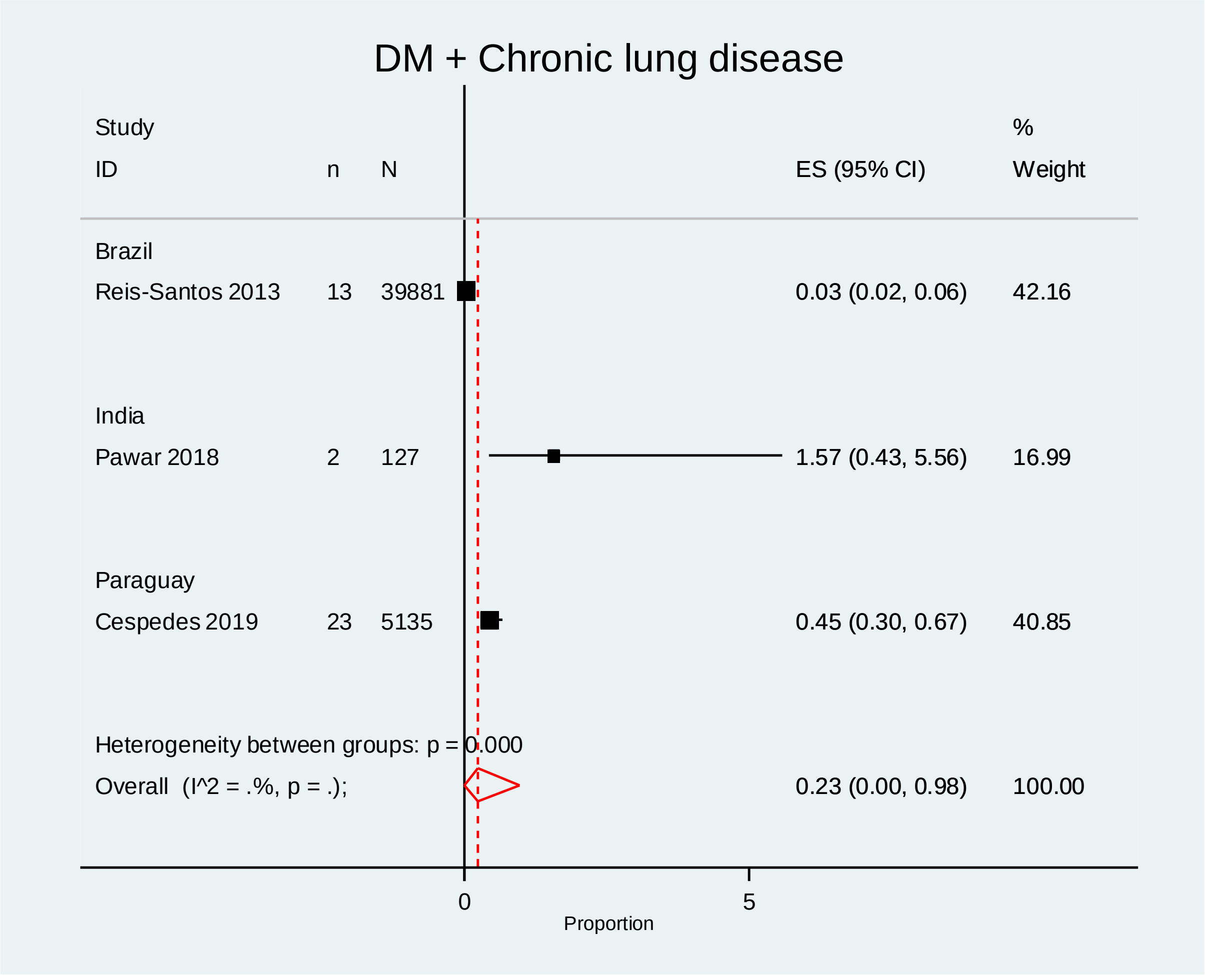

**Supplementary Figure 10: Subgroup analyses pooling prevalence of TB+Depression+Anxiety by country**

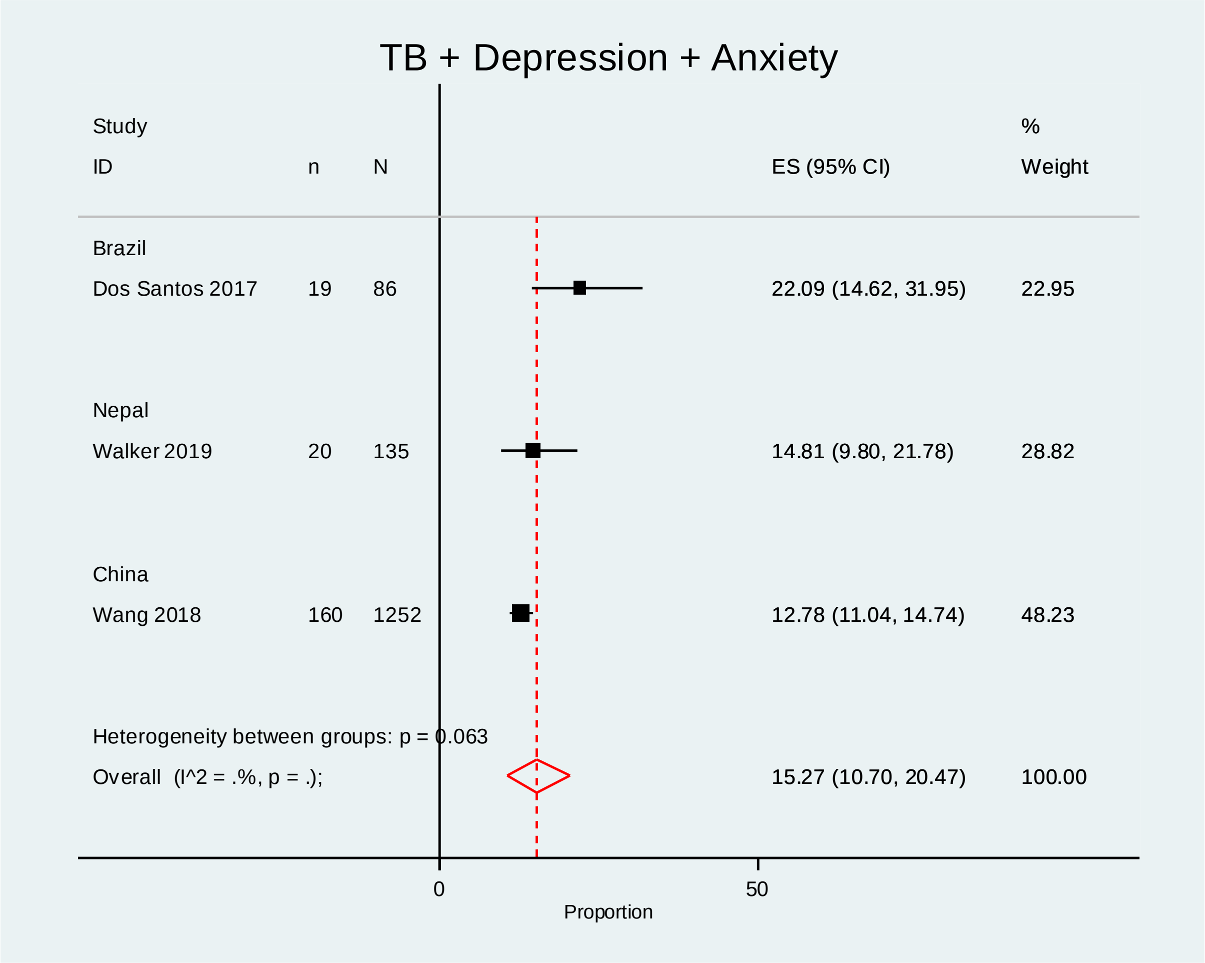

**Supplementary Figure 11: Subgroup analyses pooling prevalence of TB+HBV+Drug use by country**

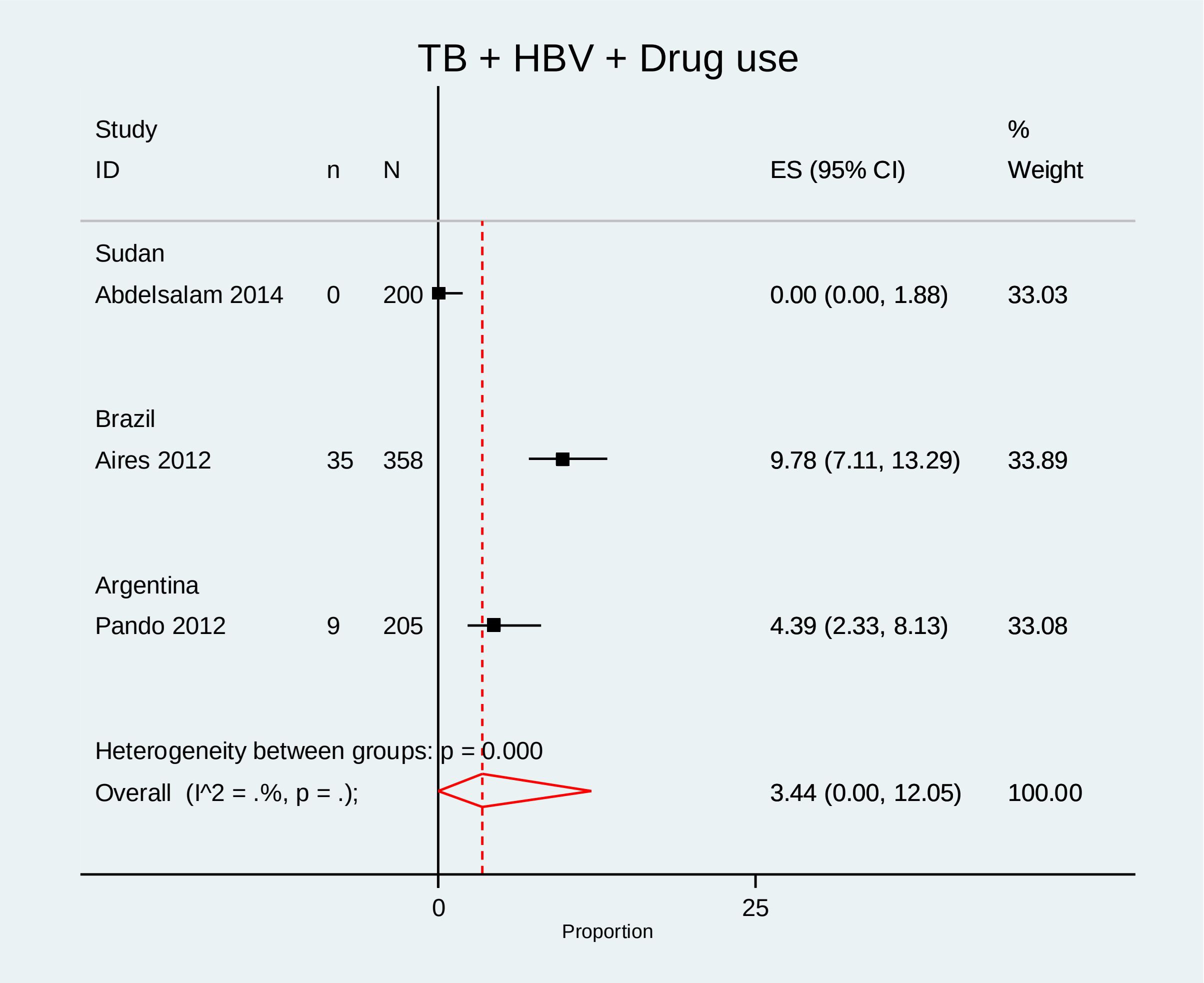

**Supplementary Figure 12: Subgroup analyses pooling prevalence of TB+HIV+Hypothyroidism by country**

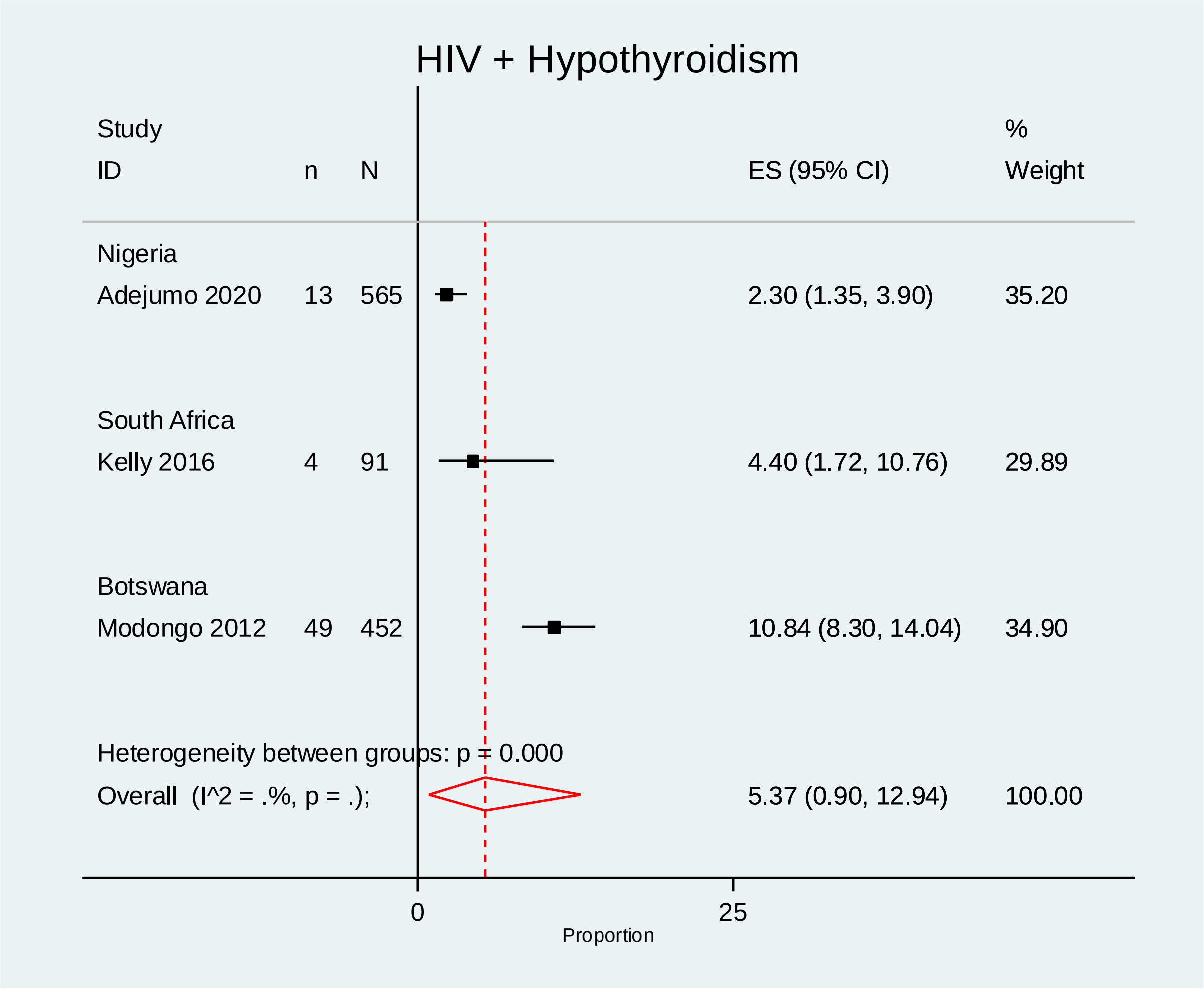

**Supplementary Figure 13: Subgroup analyses pooling prevalence of TB+DM+Alcoholism by country**

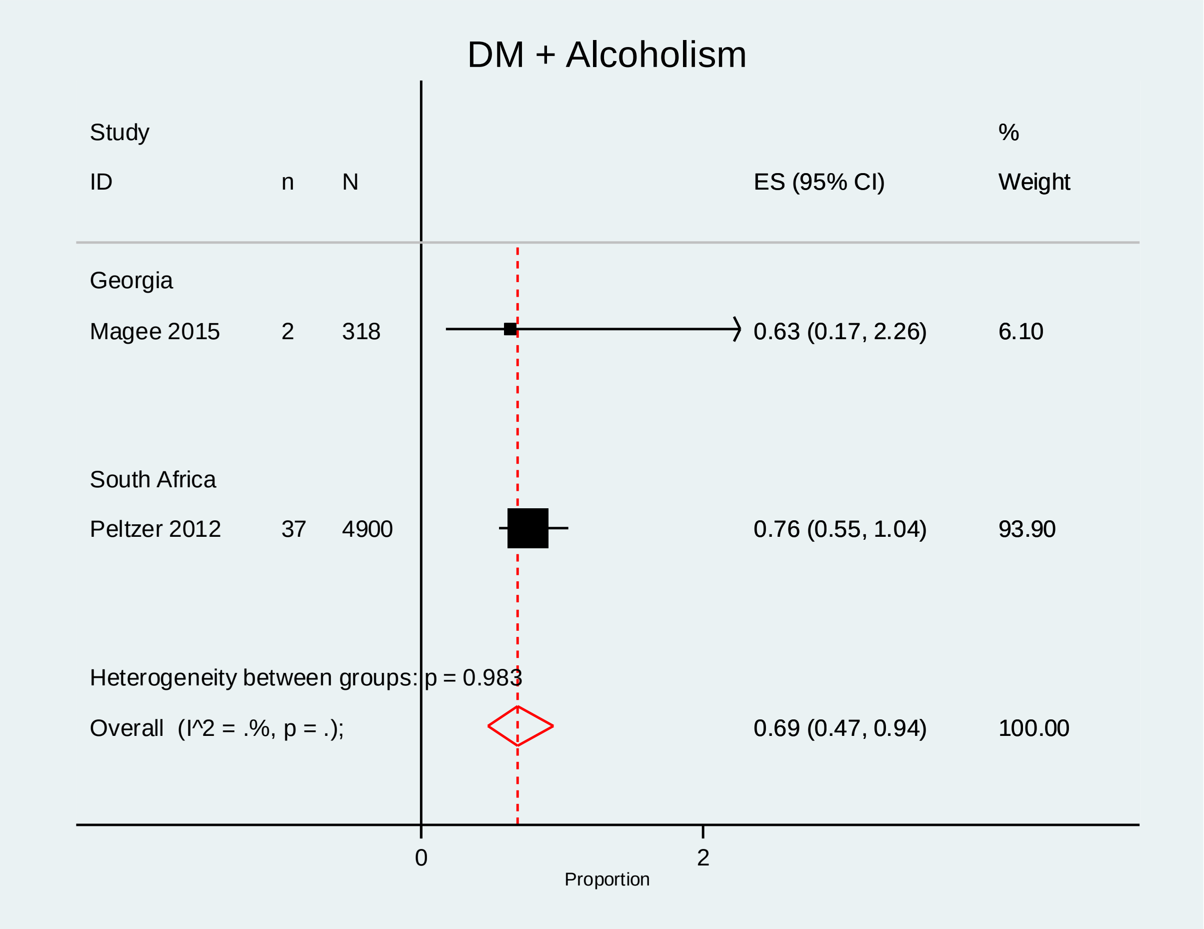

**Supplementary Figure 14: Subgroup analyses pooling prevalence of TB+DM+Chronic kidney disease by country**

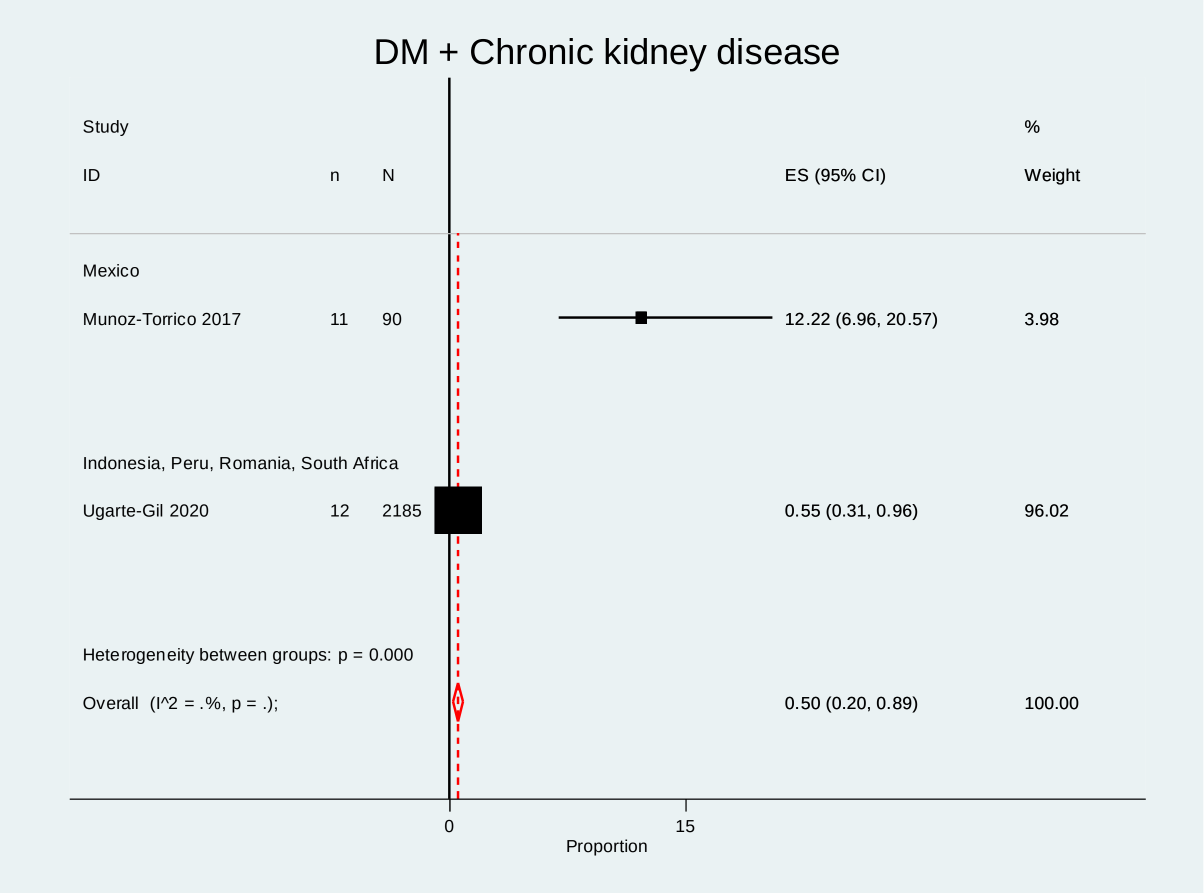

**Supplementary Figure 15: Subgroup analyses pooling prevalence of TB+ DM+ Chronic liver disease by country**

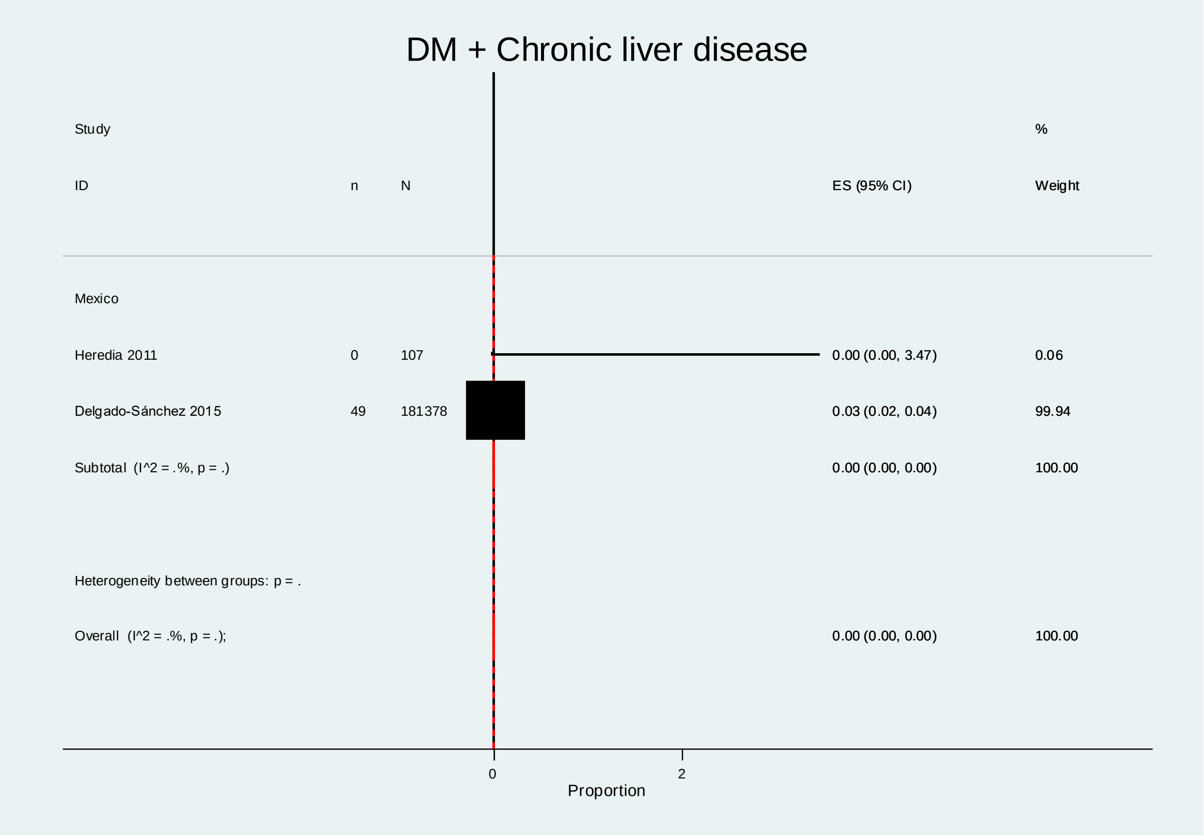

**Supplementary Figure 16: Subgroup analyses pooling prevalence of TB+ DM + HCV by country**

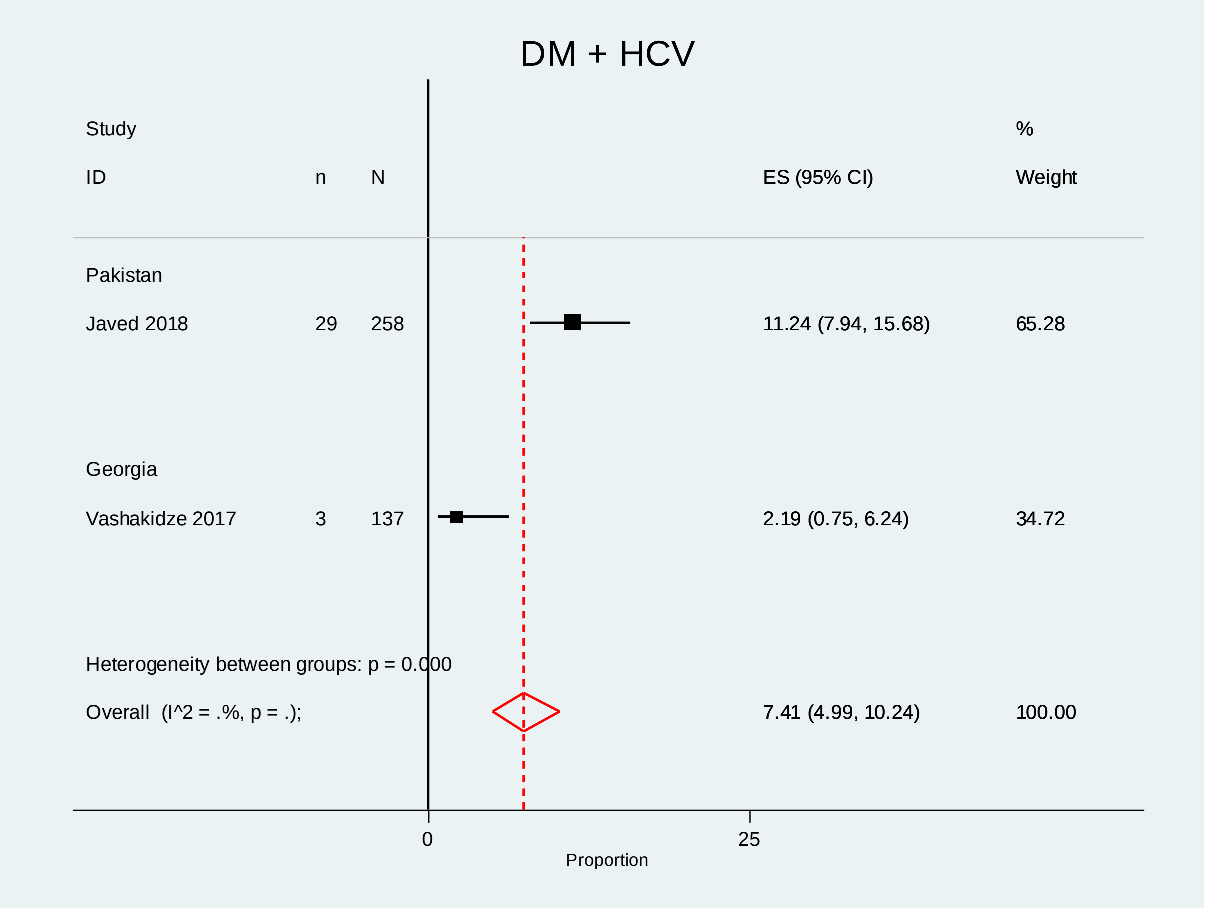

**Supplementary Figure 17: Subgroup analyses pooling prevalence of TB+DM+Heart Disease by country**

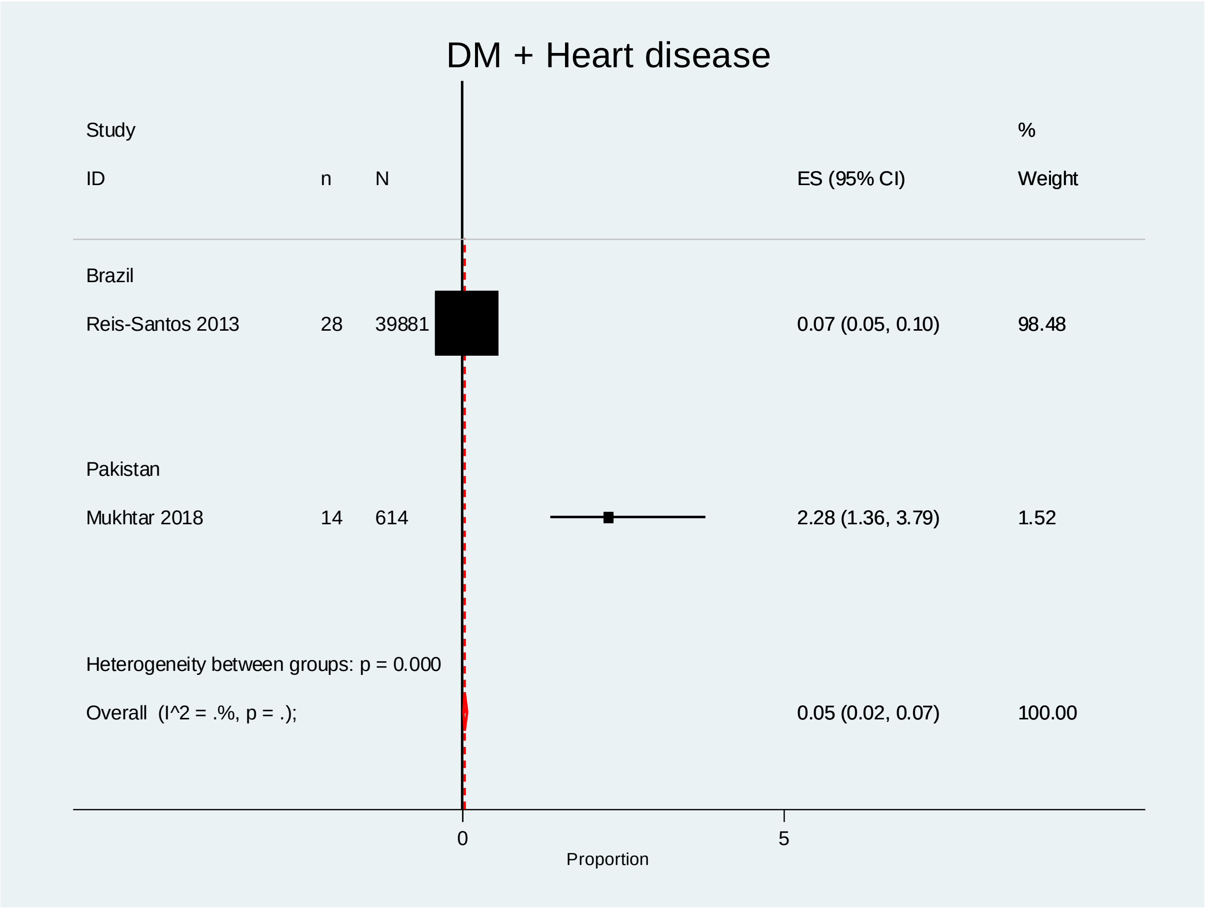

**Supplementary Figure 18: Subgroup analyses pooling prevalence of TB+Depression+Chronic lung disease by country**

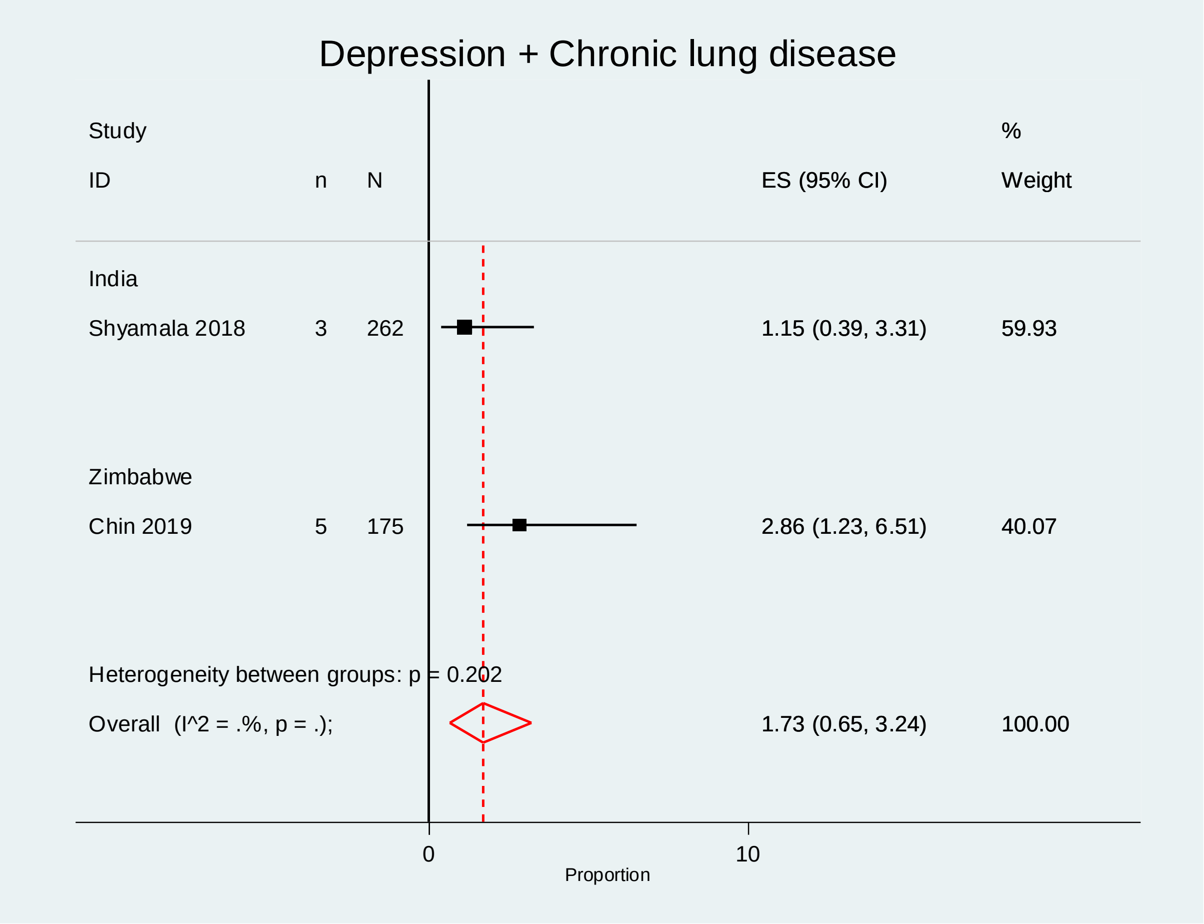

**Supplementary Figure 19: Subgroup analyses pooling prevalence of TB+HBV+HCV by country**

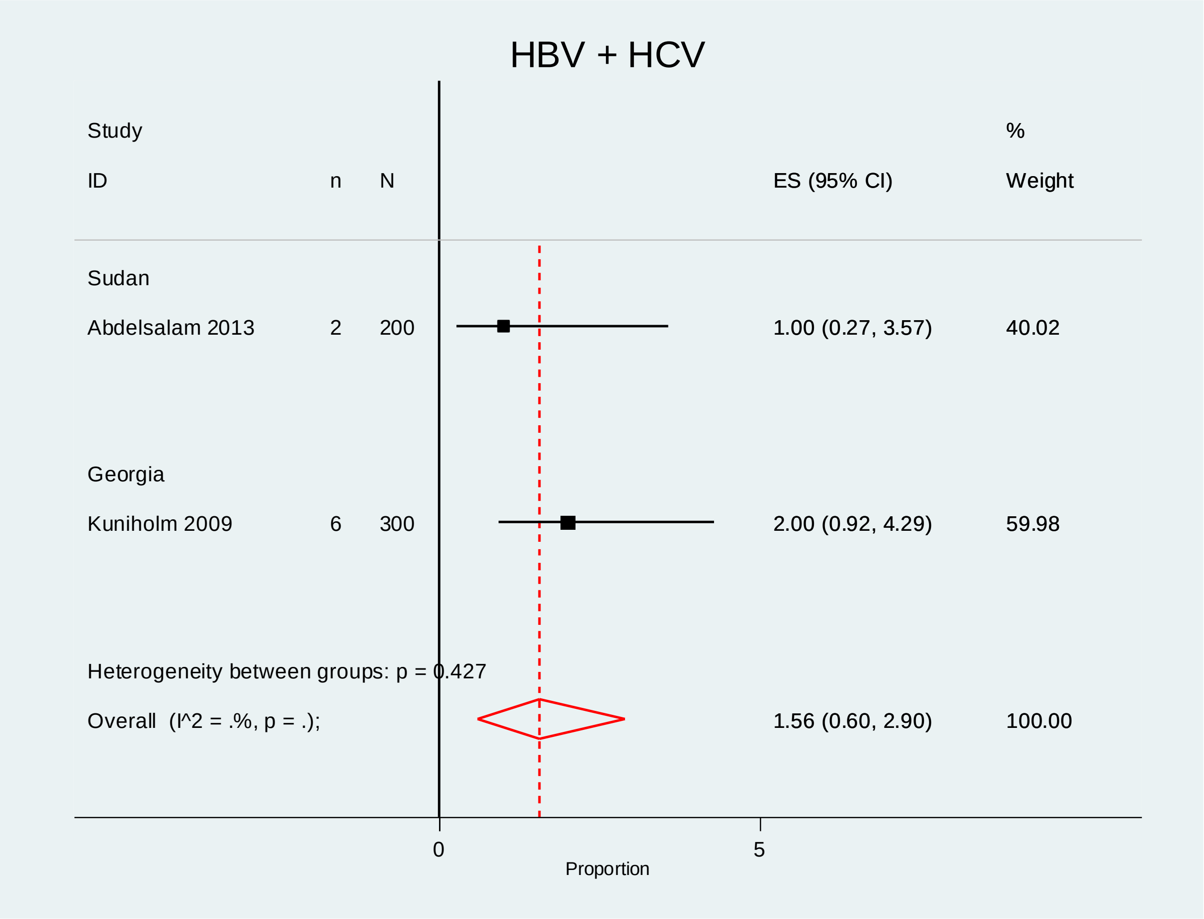

**Supplementary Figure 20: Subgroup analyses pooling prevalence of TB+HIV+Chronic liver disease by country**

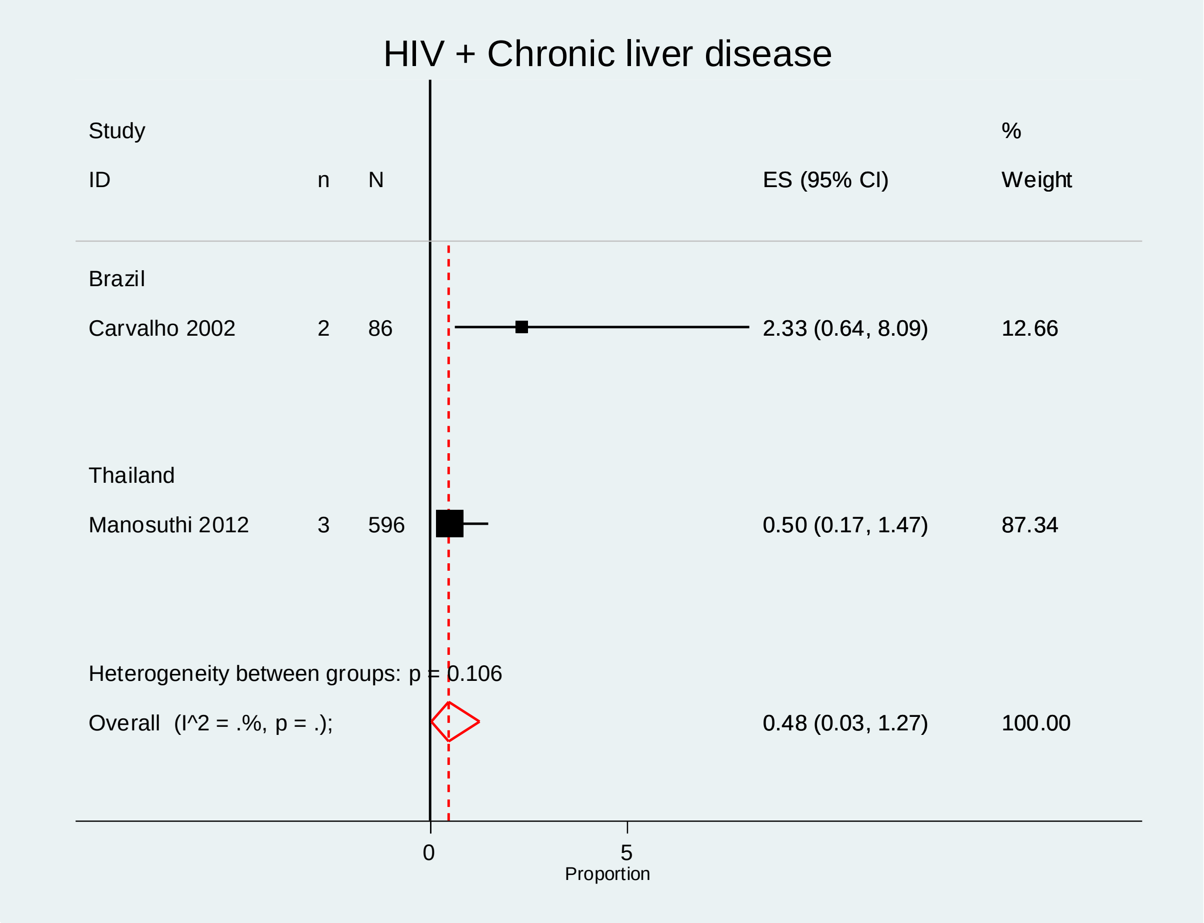

**Supplementary Figure 21: Subgroup analyses pooling prevalence of TB+HIV+Chronic lung disease by country**

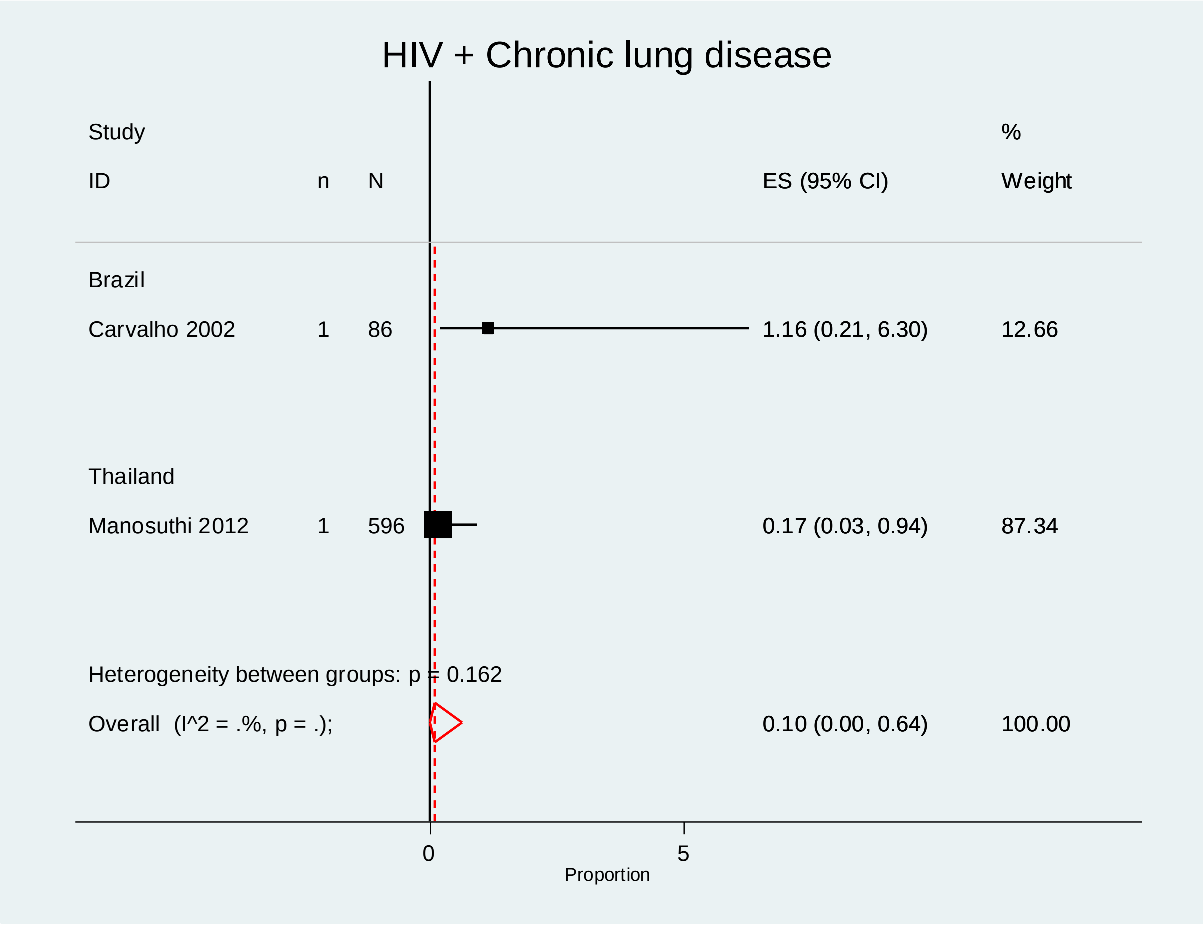

**Supplementary Figure 22: Subgroup analyses pooling prevalence of TB+HIV+PTSD by country**

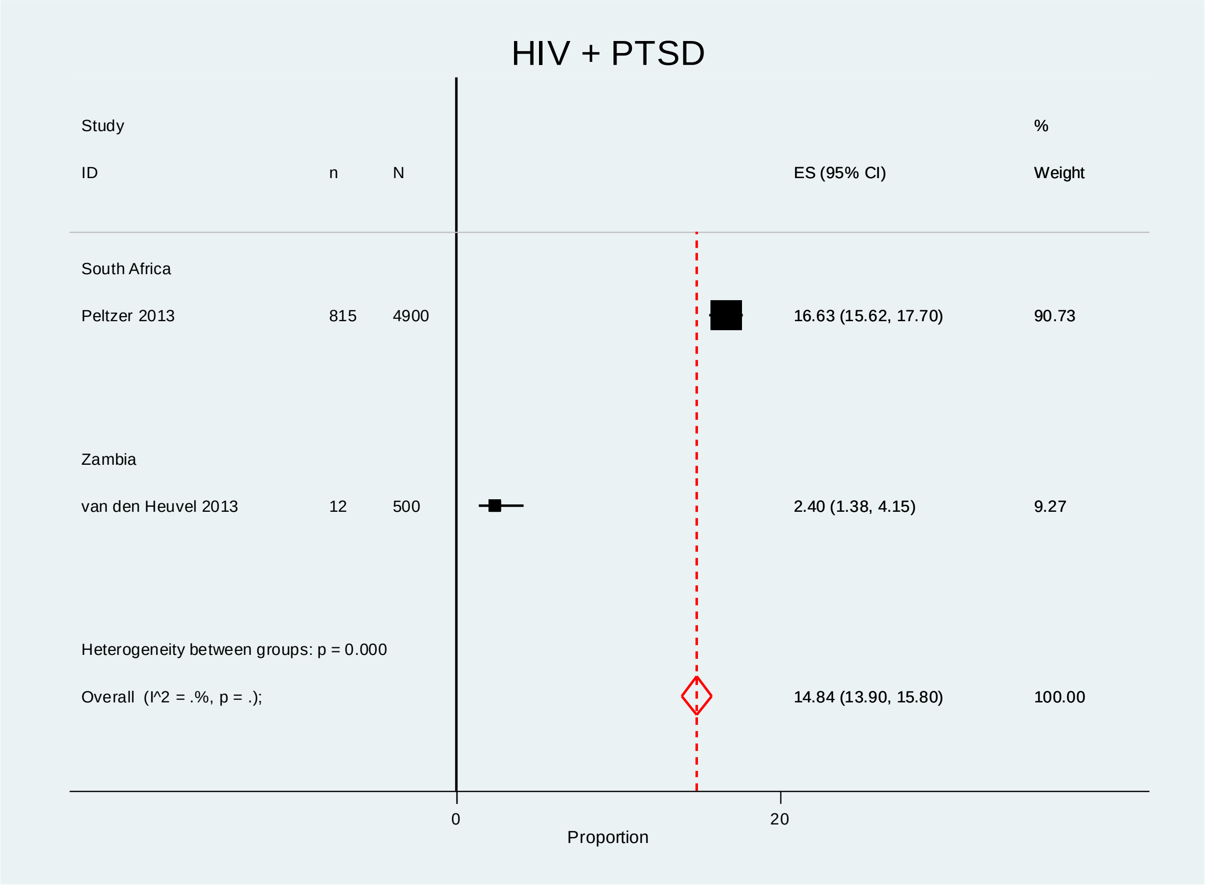

**Supplementary Figure 23: Funnel plot for the meta-analysis of TB + HIV + DM prevalence generated using the trim-and-fill method**

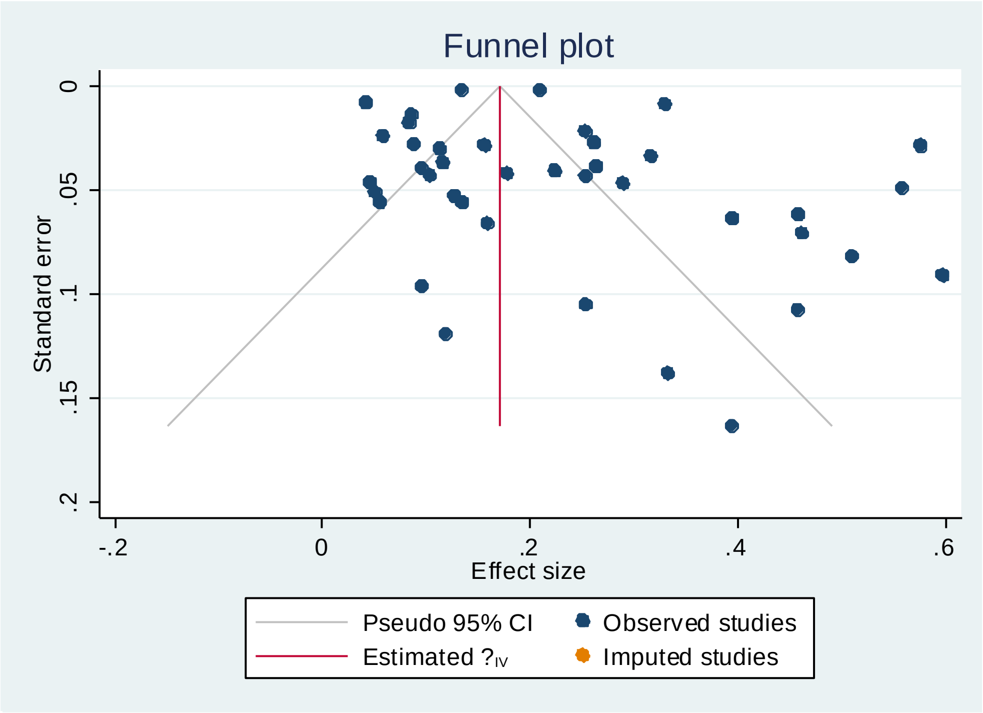
