## Appendix 1 for "Prevalence, clusters, and burden of complex tuberculosis multimorbidity in low- and middle-income countries: a systematic review and meta-analysis"

#### APPENDIX 1: Quality assessment tool (modified Newcastle-Ottawa Scale)

### <u>CODING MANUAL FOR QUALITY ASSESSMENT (modified Newcastle-Ottawa Scale)</u>

*SELECTION*

1) **Representativeness of the sample:**
   *Note that, while we are interested in any type of TB, a study's target population might be MDR-TB only. In this item we want to assess the study's sample representativeness of <u>their</u> target population (e.g. is their sample representative of the average patient with MDR-TB?)*
   a) Truly representative of the average in the target population. (all subjects or random sampling) *
   b) Somewhat representative of the average in the target population. (non-random sampling) *
   c) Selected group of users.
   d) No description of the sampling strategy.
2) **Sample size:**
   a) Justified and satisfactory *
   b) Not justified.
3) **Non-respondents:**
   *If those who did not want to participate are different who do, this could introduce bias.*
   a) Comparability between respondents and non-respondents characteristics is established, and the response rate is satisfactory. *
   b) The response rate is unsatisfactory, or the comparability between respondents and non-respondents is unsatisfactory.
   c) No description of the response rate or the characteristics of the responders and the non-responders.

*COMPARABILITY*

1) **The subjects in different outcome groups are comparable, based on the study design or analysis. Confounding factors are controlled.**
   *Note that this refers to the outcomes we are extracting. The study might report adjusted analyses for other outcomes that we are not interested in.*
   a) Study controls, either in the design (e.g. matching) or in the analyses (adjusted results), for <u>two or more</u> potential confounders (e.g. age, gender, deprivation, severity, TB type, severity of disease…) **
   b) Study controls for only one potential confounder *
   c) No potential confounder was controlled for.
   d) Not applicable (no two groups are compared)

*OUTCOME*

1) **Assessment of the outcome of interest:**
   *If we are extracting prevalence data, how is the comorbidity assessed?*
   a) Independent blind assessment. **
   b) Record linkage. **
   c) Self report. *
   d) Validated scale
   e) No description.
2) **For outcomes that require follow-up (e.g. treatment outcomes), was follow-up long enough for outcomes to occur?**
   a) Yes (≥ 6 months) *
   b) No (≤ 6 months)
   c) Not reported
3) **Statistical test:**
   *If data extracted is the result of a statistical test. If raw data (e.g. number of subjects with comorbidity) is extracted, mark as not applicable.*
   a) The statistical test used to analyze the data is clearly described and appropriate, and the measurement of the association is presented, including confidence intervals and the probability level (p value). *
   b) Not applicable (raw data extracted)
   c) The statistical test is not appropriate, not described or incomplete.
