## Appendix 2 for "Prevalence, clusters, and burden of complex tuberculosis multimorbidity in low- and middle-income countries: a systematic review and meta-analysis"

**APPENDIX 2: Reasons for exclusions for each reference screened in full text**

**- No data on TB+2 chronic conditions**

A, Sagmak Tartar; S, Ozer Balin; A, Akbulut. Is the distribution of geriatric infections different in Eastern Turkey? Retrospective evaluation of geriatric infections. Turk Geriatri Dergisi, 2018; 21(2):184-192

Aamir, Siddiqua; Aisha. Co-morbid anxiety and depression among pulmonary tuberculosis patients.. J Coll Physicians Surg Pak, 2010; 20(10):703-4

Agbor, Ako A; Bigna, Jean Joel R; Billong, Serges Clotaire; Tejiokem, Mathurin Cyrille; Ekali, Gabriel L; Plottel, Claudia S; Noubiap, Jean Jacques N; Abessolo, Hortence; Toby, Roselyne; Koulla-Shiro, Sinata. Factors associated with death during tuberculosis treatment of patients co-infected with HIV at the Yaounde Central Hospital, Cameroon: an 8-year hospital-based retrospective cohort study (2006-2013).. PLoS ONE, 2014; 9(12):e115211

Ahmad, N; Javaid, A; Basit, A; Afridi, A K; Khan, M A; Ahmad, I; Sulaiman, S A S; Khan, A H. Management and treatment outcomes of MDR-TB: results from a setting with high rates of drug resistance.. Int J Tuberc Lung Dis, 2015; 19(9):1109-ii

Ai, Xianqin; Men, Ke; Guo, Liujia; Zhang, Tianhua; Zhao, Yan; Sun, Xiaolu; Zhang, Hongwei; He, Guangxue; van der Werf, Marieke J; van den Hof, Susan. Factors associated with low cure rate of tuberculosis in remote poor areas of Shaanxi Province, China: a case control study.. BMC Public Health, 2010; 10(100968562):112

Alavi S.M.; Salami N.. The causes and risk factors of tuberculosis deaths in khuzestan. Acta Med. Iran., 2009; 47(2):89-92

Alavi-Naini, Roya; Moghtaderi, Ali; Metanat, Maliheh; Mohammadi, Mehdi; Zabetian, Mahnaz. Factors associated with mortality in tuberculosis patients.. J. res. med. sci., 2013; 18(1):52-5

Alba, Sandra; Rood, Ente; Bakker, Mirjam I; Straetemans, Masja; Glaziou, Philippe; Sismanidis, Charalampos. Development and validation of a predictive ecological model for TB prevalence.. Int J Epidemiol, 2018; 47(5):1645-1657

Ali, Nihal; Gupta, Nitin; Saravu, Kavitha. Malnutrition as an important risk factor for drug-induced liver injury in patients on anti-tubercular therapy: an experience from a tertiary care center in South India.. Drug discov. ther., 2020; 14(3):135-138

Allan J., Hsiao; Connor A., Emdin. <The> association between development assistance for health and malaria, HIV and tuberculosis mortality: a cross-national analysis. J. Epidemiol. Glob. Health, 2015; 5(1):41-48

Almeida, Paulo CÃ©sar de. Mortalidade por mÃºltiplas causas como instrumento de vigilÃ¢ncia epidemiolÃ³gica da tuberculose apÃ³s o advento da AIDS. , 1996; ():99-99

Alvarez G.G.; Thembela B.L.; Muller F.J.; Clinch J.; Singhal N.; Cameron D.W.. Tuberculosis at Edendale Hospital in Pietermaritzburg, KwaZulu Natal, South Africa. Int. J. Tuberc. Lung Dis., 2004; 8(12):1472-1478

Ambaw, F; Mayston, R; Hanlon, C; Alem, A. Is depression associated with pathways to care and diagnosis delay in people with tuberculosis in Ethiopia?.. Glob Ment Health (Camb), 2019; 6(101659641):e20

Angaali, Neeliam; Kadasu, Rajashekar; Patil, Madhusudhan Apparao; Teja, Vijay Dharma. Identification of Non-tuberculous Mycobacteria by Partial Gene Sequencing and Matrix Assisted Laser Desorption Ionisation Time of Flight at a Tertiary care Hospital Telangana, India. JOURNAL OF CLINICAL AND DIAGNOSTIC RESEARCH, 2020; 14(8):

Anunnatsiri, Siriluck; Chetchotisakd, Ploenchan; Wanke, Christine. Factors associated with treatment outcomes in pulmonary tuberculosis in northeastern Thailand.. Southeast Asian J Trop Med Public Health, 2005; 36(2):324-30

Arroyo-HernÃ¡ndez, Margarita del RocÃo; Torres-Chang, Julio Hector. Influencia de la sintomatologÃa depresiva en la adherencia al tratamiento antituberculoso en pacientes del Hospital Santa MarÃa del Socorro de Ica, 2018. Rev. mÃ©d. panacea, 2019; 8(2):58-63

Aryamkina, O L; Savonenkova, L N; Ruzov, V I; Midlenko, V I; Gnoevy Kh, V V; Razin, V A; Gamaev, R Kh. [The chronic hepatitis under pulmonary, extra-pulmonary and abdominal tuberculosis.].. Klin Lab Diagn, 2017; 62(8):462-467

Asemahagn, Mulusew Andualem; Alene, Getu Degu; Yimer, Solomon Abebe. Tuberculosis infectious pool and associated factors in East Gojjam Zone, Northwest Ethiopia.. BMC polm. med., 2019; 19(1):229

Atif, Muhammad; Sulaiman, Syed Azhar Syed; Shafie, Asrul Akmal; Ali, Irfhan; Asif, Muhammad; Babar, Zaheer-Ud-Din. Treatment outcome of new smear positive pulmonary tuberculosis patients in Penang, Malaysia.. BMC Infect Dis, 2014; 14(100968551):399

Augusto, Claudio Jose; Carvalho, Wania da Silva; Goncalves, Alan Douglas; Ceccato, Maria das Gracas Braga; Miranda, Silvana Spindola de. Characteristics of tuberculosis in the state of Minas Gerais, Brazil: 2002-2009.. J Bras Pneumol, 2013; 39(3):357-64

Aydin, I O; Ulusahin, A. Depression, anxiety comorbidity, and disability in tuberculosis and chronic obstructive pulmonary disease patients: applicability of GHQ-12.. Gen Hosp Psychiatry, 2001; 23(2):77-83

Ayin C.T.; Ayetey J.. Pulmonary tuberculosis trends and treatment outcomes in the gomoa west district, Ghana. Value Health, 2017; 20(9):A781-A782

Azhar S.; Umer R.Z.; Shahid M.. Bidirectional screening of comorbid tuberculosis and diabetes and its effect on tuberculosis treatment outcomes. Value Health, 2016; 19(3):A11-A12

Babalik A.; Kclaslan; Z.; Kizilta S.; Gencer S.; Ongen G.. A retrospective case-control study, factors affecting treatment outcomes for pulmonary tuberculosis in Istanbul, Turkey. Balkan Med. J., 2013; 30(2):204-210

Babalik, Aylin; Arda, Hulya; Bakirci, Nadi; Agca, Sinem; Oruc, Korkmaz; Kiziltas, Sule; Cetintas, Gulgun; Calisir, Haluk C. Management of and risk factors related to hepatotoxicity during tuberculosis treatment.. Tuberk. Toraks, 2012; 60(2):136-44

Balabanova, Yanina; Ignatyeva, Olga; Fiebig, Lena; Riekstina, Vija; Danilovits, Manfred; Jaama, Kadri; Davidaviciene, Edita; Radiulyte, Birute; Popa, Christina Marcela; Nikolayevskyy, Vladyslav; Drobniewski, Francis. Survival of patients with multidrug-resistant TB in Eastern Europe: what makes a difference?.. Thorax, 2016; 71(9):854-61

Bandyopadhyay, SK; Bandyopadhyay, R; Dutta, A. Profile of tuberculous meningitis with or without HIV infection and the predicators of adverse outcome. West Indian med. j, 2009; 58(6):589-592

Barbosa, Isabelle Ribeiro; Henrique, Glauber Lucena. CaracterizaÃ§Ã£o dos casos de tuberculose em um municÃpio prioritÃ¡rio no estado do Rio Grande do Norte. Rev. APS, 2014; 17(1):

Barbosa, Isabelle Ribeiro; Silva, LÃliam Ã• gna AraÃºjo Santos; Freitas, ClÃ©bio Jarlison Rego de; Silva, Elisa Marianna Abreu; Costa, Ã• ris do CÃ©u Clara. Aspectos epidemiolÃ³gicos da tuberculose no estado do Rio Grande do Norte de 2005 a 2010. ACM arq. catarin. med, 2013; 42(4):67-72

Bates, Matthew; Ahmed, Yusuf; Chilukutu, Lophina; Tembo, John; Cheelo, Busiku; Sinyangwe, Sylvester; Kapata, Nathan; Maeurer, Markus; O'Grady, Justin; Mwaba, Peter; Zumla, Alimuddin. Use of the Xpert( R) MTB/RIF assay for diagnosing pulmonary tuberculosis comorbidity and multidrug-resistant TB in obstetrics and gynaecology inpatient wards at the University Teaching Hospital, Lusaka, Zambia.. Trop Med Int Health, 2013; 18(9):1134-1140

Bates, Matthew; O'Grady, Justin; Mwaba, Peter; Chilukutu, Lophina; Mzyece, Judith; Cheelo, Busiku; Chilufya, Moses; Mukonda, Lukundo; Mumba, Maxwell; Tembo, John; Chomba, Mumba; Kapata, Nathan; Rachow, Andrea; Clowes, Petra; Maeurer, Markus; Hoelscher, Michael; Zumla, Alimuddin.

Evaluation of the burden of unsuspected pulmonary tuberculosis and co-morbidity with non-communicable diseases in sputum producing adult inpatients.. PLoS ONE, 2012; 7(7):e40774

Beare, N A V; Kublin, J G; Lewis, D K; Schijffelen, M J; Peters, R P H; Joaki, G; Kumwenda, J; Zijlstra, E E. Ocular disease in patients with tuberculosis and HIV presenting with fever in Africa.. Br J Ophthalmol, 2002; 86(10):1076-9

Beare, Nav; Kublin, J G; Lewis, D K; Schijffelen, M J; Peters, Rph; Joaki, G; Kumwenda, K; Zijlstra, E E. Ocular disease in patients with TB and HIV presenting with fever in Malawi.. Malawi Med J, 2003; 15(1):44414

Behnaz, Fatemah; Mohammadzadeh, Mahmoud; Mohammadzade, Golnaz. Five-year assessment of time of sputum smears conversion and outcome and risk factors of tuberculosis patients in central iran.. tuberc. res. treat. (print), 2015; 2015(101576351):609083

Bei, Chengli; Fu, Manjiao; Zhang, Yao; Xie, Hebin; Yin, Ke; Liu, Yanke; Zhang, Li; Xie, Bangruan; Li, Fang; Huang, Hua; Liu, Yuhong; Yang, Li; Zhou, Jing. Mortality and associated factors of patients with extensive drug-resistant tuberculosis: an emerging public health crisis in China.. BMC Infect Dis, 2018; 18(1):261

Benea O. E.; Benea S.; Ducu G.; Cozma A.; Dumitriu R.; Podani M.; Avram F.. Factors Associated with Unfavorable Clinical Outcome in HIV-TB Coinfected Patients. Jnl. Gastrointest. Liver Dis., 2012; 21(4):46

Bhering, Marcela; Duarte, Raquel; Kritski, Afranio. Predictive factors for unfavourable treatment in MDR-TB and XDR-TB patients in Rio de Janeiro State, Brazil, 2000-2016.. PLoS ONE, 2019; 14(11):e0218299

Boulle, Andrew; Davies, Mary-Ann; Hussey, Hannah; Ismail, Muzzammil; Morden, Erna; Vundle, Ziyanda; Zweigenthal, Virginia; Mahomed, Hassan; Paleker, Masudah; Pienaar, David; Tembo, Yamanya; Lawrence, Charlene; Isaacs, Washiefa; Mathema, Hlengani; Allen, Derick; Allie, Taryn; Bam, Jamy-Lee; Buddiga, Kasturi; Dane, Pierre; Heekes, Alexa; Matlapeng, Boitumelo; Mutemaringa, Themba; Muzarabani, Luckmore; Phelanyane, Florence; Pienaar, Rory; Rode, Catherine; Smith, Mariette; Tiffin, Nicki; Zinyakatira, Nesbert; Cragg, Carol; Marais, Frederick; Mudaly, Vanessa; Voget, Jacqueline; Davids, Jody; Roodt, Francois; van Zyl Smit, Nellis; Vermeulen, Alda; Adams, Kevin; Audley, Gordon; Bateman, Kathleen; Beckwith, Peter; Bernon, Marc; Blom, Dirk; Boloko, Linda; Botha, Jean; Boutall, Adam; Burmeister, Sean; Cairncross, Lydia; Calligaro, Gregory; Coccia, Cecilia; Corin, Chadwin; Daroowala, Remy; Dave, Joel A; De Bruyn, Elsa; De Villiers, Martin; Deetlefs, Mimi; Dlamini, Sipho; Du Toit, Thomas; Endres, Wilhelm; Europa, Tarin; Fieggan, Graham; Figaji, Anthony; Frankenfeld, Petro; Gatley, Elizabeth; Gina, Phindile; Govender, Evashan; Grobler, Rochelle; Gule, Manqoba Vusumuzi; Hanekom, Christoff; Held, Michael; Heynes, Alana; Hlatswayo, Sabelo; Hodkinson, Bridget; Holtzhausen, Jeanette; Hoosain, Shakeel; Jacobs, Ashely; Kahn, Miriam; Kahn, Thania; Khamajeet, Arvin; Khan, Joubin; Khan, Riaasat; Khwitshana, Alicia; Knight, Lauren; Kooverjee, Sharita; Krogscheepers, Rene; Jacque Kruger, Jean; Kuhn, Suzanne; Laubscher, Kim; Lazarus, John; Le Roux, Jacque; Lee Jones, Scott; Levin, Dion; Maartens, Gary; Majola, Thina; Manganyi, Rodgers; Marais, David; Marais, Suzaan; Maritz, Francois; Maughan, Deborah; Mazondwa, Simthandile; Mbanga, Luyanda; Mbatani, Nomonde; Mbena, Bulewa; Meintjes, Graeme; Mendelson, Marc; Moller, Ernst; Moore, Allison; Ndebele, Babalwa; Nortje, Marc; Ntusi, Ntobeko; Nyengane, Funeka; Ofoegbu, Chima; Papavarnavas, Nectarios; Peter, Jonny; Pickard, Henri; Pluke, Kent; Raubenheimer, Peter J; Robertson, Gordon; Rozmiarek, Julius; Sayed, A; Scriba, Matthias; Sekhukhune, Hennie; Singh, Prasun; Smith, Elsabe; Soldati, Vuyolwethu; Stek, Cari; van den Berg, Robert; van der Merwe, Le Roux; Venter, Pieter; Vermooten, Barbra; Viljoen, Gerrit; Viranna, Santhuri; Vogel, Jonno; Vundla, Nokubonga; Wasserman, Sean; Zitha, Eddy; Lomas-Marais, Vanessa; Lombard, Annie; Stuve, Katrin; Viljoen, Werner; Basson, De Vries; Le Roux, Sue; Linden-Mars, Ethel; Victor, Lizanne; Wates, Mark; Zwanepoel, Elbe; Ebrahim, Nabilah; Lahri, Sa'ad; Mnguni, Ayanda; Crede, Thomas; de Man, Martin; Evans, Katya; Hendrikse, Clint; Naude, Jonathan; Parak, Moosa; Szymanski, Patrick; Van Koningsbruggen, Candice; Abrahams, Riezaah; Allwood, Brian; Botha, Christoffel; Henndrik Botha, Matthys; Broadhurst, Alistair; Claasen, Dirkie; Daniel, Che; Dawood, Riyaadh; du

Preez, Marie; Du Toit, Nicolene; Erasmus, Kobie; Koegelenberg, Coenraad F N; Gabriel, Shiraaz; Hugo, Susan; Jardine, Thabiet; Johannes, Clint; Karamchand, Sumanth; Lalla, Usha; Langenegger, Eduard; Louw, Eize; Mashigo, Boitumelo; Mhlana, Nonte; Mnqwazi, Chizama; Moodley, Ashley; Moodley, Desiree; Moolla, Saadiq; Mowlana, Abdurasiet; Nortje, Andre; Olivier, Elzanne; Parker, Arifa; Paulsen, Chane; Prozesky, Hans; Rood, Jacques; Sabela, Tholakele; Schrueder, Neshaad; Sithole, Nokwanda; Sithole, Sthembiso; Taljaard, Jantjie J; Titus, Gideon; Van Der Merwe, Tian; van Schalkwyk, Marije; Vazi, Luthando; Viljoen, Abraham J; Yazied Chothia, Mogamat; Naidoo, Vanessa; Alan Wallis, Lee; Abbass, Mumtaz; Arendse, Juanita; Armien, Rizqa; Bailey, Rochelle; Bello, Muideen; Carelse, Rachel; Forgus, Sheron; Kalawe, Nosi; Kariem, Saadiq; Kotze, Mariska; Lucas, Jonathan; McClaughlin, Juanita; Murie, Kathleen; Najjaar, Leilah; Petersen, Liesel; Porter, James; Shaw, Melanie; Stapar, Dusica; Williams, Michelle; Aldum, Linda; Berkowitz, Natacha; Girran, Raakhee; Lee, Kevin; Naidoo, Lenny; Neumuller, Caroline; Anderson, Kim; Begg, Kerrin; Boerlage, Lisa; Cornell, Morna; de Waal, Renee; Dudley, Lilian; English, Rene; Euvrard, Jonathan; Groenewald, Pam; Jacob, Nisha; Jaspan, Heather; Kalk, Emma; Levitt, Naomi; Malaba, Thoko; Nyakato, Patience; Patten, Gabriela; Schneider, Helen; Shung King, Maylene; Tsondai, Priscilla; Van Duuren, James; van Schaik, Nienke; Blumberg, Lucille; Cohen, Cheryl; Govender, Nelesh; Jassat, Waasila; Kufa, Tendesayi; McCarthy, Kerrigan; Morris, Lynn; Hsiao, Nei-Yuan; Marais, Ruan; Ambler, Jon; Ngwenya, Olina; Osei-Yeboah, Richard; Johnson, Leigh; Kassanjee, Reshma; Tamuhla, Tsaone. Risk factors for COVID-19 death in a population cohort study from the Western Cape Province, South Africa.. Clin Infect Dis, 2020; (a4j, 9203213):

CaÃ±as Castillo, Karen Gabriela; Imtyaz Ahmad, Moh; Navas, Trina. Tuberculosis: caracterÃsticas epidemiolÃ³gicas en un hospital tipo IV. Med. interna (Caracas), 2015; 31(1):31-43

Camara, A; Sow, M S; Toure, A; Diallo, O H; Kaba, I; Bah, B; Diallo, T H; Diallo, M S; Guilavogui, T; Sow, O Y. [Treatment outcome, survival and their risk factors among new tuberculosis patients co-infected with HIV during the Ebola outbreak in Conakry].. Rev Epidemiol Sante Publique, 2017; 65(6):419-426

Chalya, Phillipo L; Mchembe, Mabula D; Mshana, Stephen E; Rambau, Peter F; Jaka, Hyasinta; Mabula, Joseph B. Clinicopathological profile and surgical treatment of abdominal tuberculosis: a single centre experience in northwestern Tanzania.. BMC Infect Dis, 2013; 13(100968551):270

Chandra N.. Study of socio-demographic and psychosocial factors in patients of mdr tb and xdr tb with psychiatric comorbidities. Indian J. Psychiatry, 2016; 58(5 Supplement 1):S107

Chandra P.; Singh S.; Singh B.K.. Study of psychiatric co - morbidity in cases of tuberculosis patients undergoing treatment. Indian J. Public Health Res. Dev., 2011; 2(2):111-113

Charoensakulchai S.; Limsakul M.; Saengungsumalee I.; Usawachoke S.; Udomdech A.; Pongsaboripat A.; Kaewput W.; Sakboonyarat B.; Rangsin R.; Suwannahitatorn P.; Mungthin M.; Piyaraj P.. Characteristics of poor tuberculosis treatment outcomes among patients with pulmonary tuberculosis in community hospitals of Thailand. Am. J. Trop. Med. Hyg., 2020; 102(3):553-561

Chatterjee S.; Agrawal D.; Parchand S.M.; Sahu A.. Visual outcome and prognostic factors in cataract surgery in ocular tuberculosis. Indian J Ophthalmol, 2020; 68(9):1894-1900

Chittoor G.; Arya R.; Farook V.S.; David R.; Puppala S.; Resendez R.G.; Rivera-Chavira B.E.; Leal-Berumen I.; Zenteno-Cuevas R.; Lopez-Alvarenga J.C.; Bastarrachea R.A.; Curran J.E.; Dhandayuthapani S.; Gonzalez L.; Blangero J.; Crawford M.H.; Vlasich E.M.; Escobedo L.G.; Duggirala R.. Epidemiologic investigation of tuberculosis in a Mexican population from Chihuahua State, Mexico: A pilot study. Tuberculosis, 2013; 93(SUPPL.):S71-S77

Chittoor, Geetha; Arya, Rector; Farook, Vidya S; David, Randy; Puppala, Sobha; Resendez, Roy G; Rivera-Chavira, Blanca E; Leal-Berumen, Irene; Zenteno-Cuevas, Roberto; Lopez-Alvarenga, Juan Carlos; Bastarrachea, Raul A; Curran, Joanne E; Dhandayuthapani, Subramanian; Gonzalez, Lupe; Blangero, John; Crawford, Michael H; Vlasich, Esteban M; Escobedo, Luis G; Duggirala, Ravindranath. Epidemiologic investigation of tuberculosis in a Mexican population from Chihuahua State, Mexico: a pilot study.. Tuberculosis (Edinb), 2013; 93 Suppl(d4x, 100971555):S71-7

Chong, V H; Rajendran, N. Tuberculosis peritonitis in Negara Brunei Darussalam.. Ann Acad Med Singapore, 2005; 34(9):548-52

Chowdary, K. P. R.; Sumalatha, G.; Gouri, K. Sri; Dani, K. J. S.. A PROSPECTIVE OBSERVATIONAL STUDY ON PRESCRIPTION PATTERN, DRUG UTILIZATION AND AUDIT FOR THE TREATMENT OF TUBERCULOSIS IN A TERTIARY CARE HOSPITAL IN ANDHRA PRADESH. INDO AMERICAN JOURNAL OF PHARMACEUTICAL SCIENCES, 2017; 4(6):1636-1640

Chua Jamie R; Mejia-Christina-Irene-D; Berba Regina P. Prevalence, clinical profile, and treatment outcomes of adult patients diagnosed with disseminated tuberculosis seen at University of the Philippines Manila-Philippine General Hospital Tuberculosis Directly Observed Treatment Short Course (TB-DOTS) Clinic. Acta Medica Philippina, 2017; ():300-309

Chung-Tae KIM; Nam-Soo RHU; Dong-Il CHO. Clinical charateristics of elderly patients with plmonary tuberculosis. Tuberculosis and Respiratory Diseases, 2000; ():432-440

Coelho, Andrea Gobetti Vieira. Tuberculose multirresistente e extensivamente resistente em Ã¡rea metropolitana de elevada incidÃªncia - municÃpio de Santos (SP), Brasil. , 2014; ():[101]-[101]

Coelho, Andrea Gobetti Vieira; Zamarioli, Liliana Aparecida; Perandones, Carmen ArgÃ¼ello; Cuntiere, Ivonete; Waldman, Eliseu Alves. CaracterÃsticas da tuberculose pulmonar em Ã¡rea hiperendÃªmica: municÃpio de Santos (SP). J. bras. pneumol, 2009; 35(10):998-1007

Cohen, Danielle B; Davies, Geriant; Malwafu, Wakisa; Mangochi, Helen; Joekes, Elizabeth; Greenwood, Simon; Corbett, Liz; Squire, S Bertel. Poor outcomes in recurrent tuberculosis: More than just drug resistance?.. PLoS ONE, 2019; 14(5):e0215855

Contreras, Carmen C; Millones, Ana K; Santa Cruz, Janeth; Aguilar, Margot; Clendenes, Martin; Toranzo, Miguel; Llaro, Karim; Lecca, Leonid; Becerra, Mercedes C; Yuen, Courtney M. Addressing tuberculosis patients' medical and socio-economic needs: a comprehensive programmatic approach.. Trop Med Int Health, 2017; 22(4):505-511

Corrales-Alvarez, Milena; de la Pava-Salgado, Elmer; Hurtado-TobÃ³n, Luis H.. Incidencia del VIH en la Tuberculosis, en Armenia, Colombia. Rev. salud pÃºblica, 2011; 13(6):1022-1030

Correa de Adjounian, M; HernÃ¡ndez, C; AlveÃ¡rez, O; GonzÃ¡lez Rico, S; Pedroza, R; CÃ©spedes, G; RodrÃguez, B; GÃ³mez, M. Micobacterias en muestras de autopsias. Rev. Soc. Venez. Microbiol, 2004; 24(44228):44508

Coutinho, Regina Claudia Gayoso de Azeredo. Fatores associados ao Ã³bito em pacientes com tuberculose multirresistente tratados nos centros de referÃªncia brasileiros de 2005 a 2012: anÃ¡lise de sobrevivÃªncia. , 2016; ():94-94

D, Vijayaraju; M, Manjula.. A study on adverse drug reaction associated co-morbid symptoms on newly diagnosed TB patients with and without diabetes mellitus. , 2019; 6():

Ð¡Ñ‹Ñ• Ð¾ÐµÐ²Ð°, Ð'. Ð'.; Ð”ÐµÐ½Ð¸Ñ• Ð¾Ð²Ð°, Ð• . Ð'.; Ð• ÐµÑ„ÐµÐ´Ð¾Ð²Ð°, Ð• . Ð".; ÐšÐ¾Ð²Ð°Ð»ÐµÐ²Ð°, Ð¡. Ð• .; ÐšÐ¸Ñ€ÑŽÑ…Ð¸Ð½Ð°, Ð›. Ð”.. ÐžÑ• Ð¾Ð±ÐµÐ½Ð½Ð¾Ñ• Ñ‚Ð¸ Ñ„ÑƒÐ½ÐºÑ†Ð¸Ð¾Ð½Ð°Ð»ÑŒÐ½Ð¾Ð³Ð¾ Ñ• Ð¾Ñ• Ñ‚Ð¾Ñ• Ð½Ð¸Ñ• Ð´Ñ‹Ñ…Ð°Ñ‚ÐµÐ»ÑŒÐ½Ñ‹Ñ… Ð¼Ñ‹ÑˆÑ† Ñƒ Ð¿Ð°Ñ†Ð¸ÐµÐ½Ñ‚Ð¾Ð² Ñ• Ñ‚ÑƒÐ±ÐµÑ€ÐºÑƒÐ»ÐµÐ·Ð¾Ð¼ Ð»ÐµÐ³ÐºÐ¸Ñ…. Ð¢ÑƒÐ±ÐµÑ€ÐºÑƒÐ»ÐµÐ· Ð¸ Ñ• Ð¾Ñ†Ð¸Ð°Ð»ÑŒÐ½Ð¾-Ð·Ð½Ð°Ñ‡Ð¸Ð¼Ñ‹Ðµ Ð·Ð°Ð±Ð¾Ð»ÐµÐ²Ð°Ð½Ð¸Ñ• , 2018; (4):41548

Damasceno, Glauciene Santana; Guaraldo, Lusiele; Engstrom, Elyne Montenegro; Theme Filha, Mariza Miranda; Souza-Santos, Reinaldo; Vasconcelos, Ana Gloria Godoi; Rozenfeld, Suely. Adverse reactions to antituberculosis drugs in Manguinhos, Rio de Janeiro, Brazil.. Clinics, 2013; 68(3):329-37

Danilla DÃŸvila, Mario; MartÃnez-Merizalde Huatuco, Nelson; Gave ZÃŸrate, Jorge. La tuberculosis en el Hospital Arzobispo Loayza. Enfer. tÃ³rax (Lima), 2004; 48(2):160-166

De Melo F.A.F.; Afiune J.B.; Ide Neto J.; De Almeida E.A.; Spada D.T.A.; Antelmo A.N.L.; Cruz M.L.. Epidemiological features of multidrug-resistant tuberculosis in a reference service in Sao Paulo city. Rev. Soc. Bras. Med. Trop., 2003; 36(1):27-34

Devkota, J; Devkota, N; Lohani, SP. Health Related Quality of Life, Anxiety and Depression among Tuberculosis Patients in Kathmandu, Nepal. , 2017; 4():

Dewan, P K; Arguin, P M; Kiryanova, H; Kondroshova, N V; Khorosheva, T M; Laserson, K; Kluge, H; Jakubowiak, W; Wells, C; Kazionny, B. Risk factors for death during tuberculosis treatment in Orel, Russia.. Int J Tuberc Lung Dis, 2004; 8(5):598-602

Dixit, Ramakant; Arya, Manoj Kumar; Panjabi, Mukesh; Gupta, Avinash; Paramez, A R. Clinical profile of patients having splenic involvement in tuberculosis.. INDIAN J. TUBERC., 2010; 57(1):25-30

Donoso G., Felipe; Capella S., Daniela; Segovia M., Roberto; Pinto I., Andrea; Roblero C., Juan Pablo; PeÃ±a M., Carlos; Estela P., Ricardo; Alonso T., Faustino. Frecuencia y factores asociados a daÃ±o hepÃ¡tico inducido por medicamentos en tratamiento antituberculosis. Gastroenterol. latinoam, 2013; 24(2):63-66

Edwards, T; White, L V; Lee, N; Castro, M C; Saludar, N R; Faguer, B N; Fuente, N D; Mayoga, F; Ariyoshi, K; Garfin, A M C G; Solon, J A; Cox, S E. Effects of comorbidities on quality of life in Filipino people with tuberculosis.. Int J Tuberc Lung Dis, 2020; 24(7):712-719

El-Sony, A; Enarson, D; Khamis, A; Baraka, O; Bjune, G. Relation of grading of sputum smears with clinical features of tuberculosis patients in routine practice in Sudan.. Int J Tuberc Lung Dis, 2002; 6(2):91-7

Espindola, Lidia Codorniz Delamare. Estudo da mortalidade por tuberculose em Campo Grande - MS, 2001 a 2008. , 2010; ():ix,44-ix,44

F.C, Ugoeze; P.U, Ele; A.E, Anyabolu; E.H, Enemuo; R.C, Okonkwo; E.O, Umeh; I.U, Ezeani; U.U, Onyeonoro. Pulmonary tuberculosis and diabetes mellitus co-morbidity in a Nigerian tertiary hospital. African Journal of Respiratory Medicine, 2020; 14(2):13-17

Farazi, Aliasghar; Sofian, Masoomeh; Jabbariasl, Mansoureh; Keshavarz, Sara. Adverse reactions to antituberculosis drugs in Iranian tuberculosis patients.. tuberc. res. treat. (print), 2014; 2014(101576351):412893

Fatemeh SARVI; Abbas MOGHIMBEIGI; Hossein MAHJUB; Mahshid NASEHI; Mahmoud KHODADOST. Factors associated with mortality from tuberculosis in Iran: an application of a generalized estimating equation-based zero-inflated negative binomial model to national registry data. Epidemiology and Health, 2019; ():e2019032-e2019032

Fescina, Pablo MartÃn; Membriani, Evangelina; Limongi, Leticia; Putruele, Ana. Incidencia de la resistencia a drogas en tuberculosis y su asociaciÃ³n a comorbilidades en pacientes tratados en un hospital universitario. Rev. am. med. respir, 2013; 13(2):64-70

Fleming M.F.; Krupitsky E.; Tsoy M.; Zvartau E.; Brazhenko N.; Jakubowiak W.; McCaul M.E.. Alcohol and drug use disorders, HIV status and drug resistance in a sample of Russian TB patients. Int. J. Tuberc. Lung Dis., 2006; 10(5):565-570

Foocharoen, Chingching; Nanagara, Ratanavadee; Foocharoen, Thanit; Mootsikapun, Pirun; Suwannaroj, Siraphop; Mahakkanukrauh, Ajanee. Clinical features of tuberculous septic arthritis in Khon Kaen, Thailand: a 10-year retrospective study.. Southeast Asian J Trop Med Public Health, 2010; 41(6):1438-46

Furin, J J; Mitnick, C D; Shin, S S; Bayona, J; Becerra, M C; Singler, J M; Alcantara, F; Castanieda, C; Sanchez, E; Acha, J; Farmer, P E; Kim, J Y. Occurrence of serious adverse effects in patients receiving community-based therapy for multidrug-resistant tuberculosis.. Int J Tuberc Lung Dis, 2001; 5(7):648-55

Gadoev, Jamshid; Asadov, Damin; Harries, Anthony D; Parpieva, Nargiza; Tayler-Smith, Katie; Isaakidis, Petros; Ali, Engy; Hinderaker, Sven Gudmund; Ogtay, Gozalov; Ramsay, Andrew;

Jalolov, Avazbek; Dara, Masoud. Recurrent tuberculosis and associated factors: A five - year countrywide study in Uzbekistan.. PLoS ONE, 2017; 12(5):e0176473

Gallego, Claudio; Salomone, CÃ©sar; Poropat, Alejandra. Results obtained by using a self-administered treatment for tuberculosis. Rev. am. med. respir, 2017; 17(2):152-156

Gao, Xiao-feng; Chen, Jian; Yang, Xiao-dong; Sun, Xin; Li, You-ping; Qin, Wen-xia. [Study on the results of treating tuberculosis inpatient in the general hospitals: a correspondence analysis].. Chung Hua Liu Hsing Ping Hsueh Tsa Chih, 2006; 27(8):716-20

Garcia-Goez J.F.; Rodriguez-Tabares J.F.; Orozco-Erazo C.E.; Parra-Lara L.G.; Velez J.D.; Moncada P.A.; Rosso F.. An appraisal of the isoniazid resistant tuberculosis in Colombia: A underestimated problem in Colombia?. Infectio, 2020; 24(3):173-181

Garrido, Marlucia da Silva; Penna, Maria Lucia; Perez-Porcuna, Tomas M; de Souza, Alexandra Brito; Marreiro, Leni da Silva; Albuquerque, Bernardino Claudio; Martinez-Espinosa, Flor Ernestina; Buhrer-Sekula, Samira. Factors associated with tuberculosis treatment default in an endemic area of the Brazilian Amazon: a case control-study.. PLoS ONE, 2012; 7(6):e39134

Gaude, Gajanan S; Chaudhury, Alisha; Hattiholi, Jyothi. Drug-induced hepatitis and the risk factors for liver injury in pulmonary tuberculosis patients.. J. family med. prim., 2015; 4(2):238-43

Gayoso, Regina; Dalcolmo, Margareth; Braga, Jose Ueleres; Barreira, Draurio. Predictors of mortality in multidrug-resistant tuberculosis patients from Brazilian reference centers, 2005 to 2012.. Braz J Infect Dis, 2018; 22(4):305-310

Gazetta, Claudia Eli; Takayanagui, Angela M. M; Costa JÃºnior, Moacir Lobo da; Villa, Teresa Cristina Scatena; Vendramini, Silvia Helena F. Aspectos epidemiolÃ³gicos da tuberculose em SÃ£o JosÃ© do Rio Preto - SP, a partir das notificaÃ§Ãµes da doenÃ§a em um Hospital - Escola (1993-1998). PulmÃ¤o RJ, 2003; 12(3):155-162

GBD 2017 Disease and Injury Incidence and Prevalence Collaborators. Global, regional, and national incidence, prevalence, and years lived with disability for 354 diseases and injuries for 195 countries and territories, 1990-2017: a systematic analysis for the Global Burden of Disease Study 2017.. Lancet, 2018; 392(10159):1789-1858

Getachew T.; Bayray A.; Weldearegay B.. Survival and predictors of mortality among patients under multi-drug resistant tuberculosis treatment in Ethiopia: St. Peter's specialized tuberculosis hospital, Ethiopia. Int. J. Pharm. Sci. Res., 2013; 4(2):772-783

Golsha R.; Rezaei S.R.; Shafiee A.; Najafi L.; Dashti M.; Roshandel G.. Pulmonary tuberculosis and some underlying conditions in Golestan Province of Iran, during 2001-2005. J. Clin. Diagn. Res., 2009; 3(1):1302-1306

Gomes N.M.F.; Bastos M.C.M.; Marins R.M.; Barbosa A.A.; Soares L.C.P.; De Abreu A.M.O.W.; Souto Filho J.T.D.. Differences between Risk Factors Associated with Tuberculosis Treatment Abandonment and Mortality. Pulm. Med., 2015; 2015((Gomes, Bastos, Marins, Barbosa, Soares, De Abreu, Souto Filho) Faculdade de Medicina de Campos (FMC), Campos dos Goytacazes, RJ 28035-581, Brazil):546106

Gomes, Teresa; Reis-Santos, Barbara; Bertolde, Adelmo; Johnson, John L; Riley, Lee W; Maciel, Ethel Leonor. Epidemiology of extrapulmonary tuberculosis in Brazil: a hierarchical model.. BMC Infect Dis, 2014; 14(100968551):9

GonÃ§alves, Berenice das Dores; Cavalini, Luciana Tricai; Valente, Joaquim GonÃ§alves. Monitoramento epidemiolÃ³gico da tuberculose em um hospital geral universitÃ¡rio. J. bras. pneumol, 2010; 36(3):347-355

Goncalves, Maria Jacirema Ferreira; Ferreira, Alaidistania A. Factors associated with length of hospital stay among HIV positive and HIV negative patients with tuberculosis in Brazil.. PLoS ONE, 2013; 8(4):e60487

GonzÃ¡lez, Alejandra; FernÃ¡ndez Casares, Marcelo; Baldini, MatÃas; Monteverde, Alfredo. Tuberculosis pulmonar de campos inferiores. Medicina (B.Aires), 2010; 70(5):434-436

GonzÃ¡lez, Claudio; SÃ¡enz, CÃ©sar; Herrmann, Eduardo; Jajati, MÃ³nica; Kaplan, Paula; MonzÃ³n, Dora. Tratamiento directamente observado de la tuberculosis en un hospital de la Ciudad de Buenos Aires. Medicina (B.Aires), 2012; 72(5):371-379

Gonzalez, Alejandra; Fernandez Casares, Marcelo; Baldini, Matias; Monteverde, Alfredo. [Lower lung field tuberculosis].. Medicina (B Aires), 2010; 70(5):434-6

Gonzalez, Claudio; Saenz, Cesar; Herrmann, Eduardo; Jajati, Monica; Kaplan, Paula; Monzon, Dora. [Directly observed treatment for tuberculosis in a Buenos Aires City hospital].. Medicina (B Aires), 2012; 72(5):371-9

Gothankar, Jayashree Sachin; Patil, Usha Pravin; Gaikwad, Sunil R.; Kamble, Sulakshana B.. Care seeking behaviour and various delays in Tuberculosis patients registered under RNTCP in Pune city. INDIAN JOURNAL OF COMMUNITY HEALTH, 2016; 28(1):48-53

Grandjean L.; Crossa A.; Gilman R.; Moore D.. Tuberculosis in household contacts of multidrugresistant tuberculosis patients. Eur. Respir. J., 2011; 38(SUPPL. 55):

Grandjean, L; Crossa, A; Gilman, R H; Herrera, C; Bonilla, C; Jave, O; Cabrera, J L; Martin, L; Escombe, A R; Moore, D A J. Tuberculosis in household contacts of multidrug-resistant tuberculosis patients.. Int J Tuberc Lung Dis, 2011; 15(9):1164-i

Guevara Francesa, Giancarlo; Navarro Mora, Monserrat; GonzÃ¡lez Luna, Jennyffer. EpidemiologÃa de la Tuberculosis en el Ã• rea de Salud de Pavas, Costa Rica. Enferm. actual Costa Rica (Online), 2018; (35):85-102

Gupta, Dheeraj; Singh, Navneet; Kumar, Ravinder; Jindal, Surinder K. Manifestations of pulmonary tuberculosis in the elderly: a prospective observational study from north India.. Indian J Chest Dis Allied Sci, 2008; 50(3):263-7

Gupta, H L; Yadav, M; Sundarka, M K; Talwar, V; Saini, M; Garg, P. A study of prevalence of health problems in asymptomatic elderly individuals in Delhi.. , 2002; ():

Gupta, Soham; Shenoy, Vishnu Prasad; Mukhopadhyay, Chiranjay; Bairy, Indira; Muralidharan, Sethumadhavan. Role of risk factors and socio-economic status in pulmonary tuberculosis: a search for the root cause in patients in a tertiary care hospital, South India.. Trop Med Int Health, 2011; 16(1):74-8

Hameed, Sidra; Zuberi, Faisal Faiyaz; Hussain, Sagheer; Ali, Syed Khalid. Risk factors for mortality among inpatients with smear positive pulmonary tuberculosis.. Pak. j. med. sci., 2019; 35(5):1361-1365

Harris, Tashneem; Bardien, Soraya; Schaaf, H Simon; Petersen, Lucretia; De Jong, Greetje; Fagan, Johannes J. Aminoglycoside-induced hearing loss in HIV-positive and HIV-negative multidrug-resistant tuberculosis patients. South African Medical Journal, 2012; 102(6):

He, Yu; Han, Chao; Chang, Kai-Feng; Wang, Mao-Shui; Huang, Tian-Ren. Total delay in treatment among tuberculous meningitis patients in China: a retrospective cohort study.. BMC Infect Dis, 2017; 17(1):341

Hella, Jerry; Cercamondi, Colin I; Mhimbira, Francis; Sasamalo, Mohamed; Stoffel, Nicole; Zwahlen, Marcel; Bodmer, Thomas; Gagneux, Sebastien; Reither, Klaus; Zimmermann, Michael B; Risch, Lorenz; Fenner, Lukas. Anemia in tuberculosis cases and household controls from Tanzania: Contribution of disease, coinfections, and the role of hepcidin.. PLoS ONE, 2018; 13(4):e0195985

Hermosilla, Sabrina; You, Paul; Aifah, Angela; Abildayev, Tleukhan; Akilzhanova, Ainur; Kozhamkulov, Ulan; Muminov, Talgat; Darisheva, Meruert; Zhussupov, Baurzhan; Terlikbayeva, Assel; El-Bassel, Nabila; Schluger, Neil. Identifying risk factors associated with smear positivity of pulmonary tuberculosis in Kazakhstan.. PLoS ONE, 2017; 12(3):e0172942

Hernandez-Guerrero I.A.; Vazquez-Martinez V.H.; Guzman-Lopez F.; Ochoa-Jimenez L.G.; Cervantes-Vazquez D.A.. Social and clinical profile of patients with tuberculosis in a family medicine unit in Reynosa, Tamaulipas, Mexico. Atencion Fam., 2016; 23(1):41487

Huaman, Moises A; Henson, David; Rondan, Paola L; Ticona, Eduardo; Miranda, Gustavo; Kryscio, Richard J; Mugruza, Raquel; Aranda, Ernesto; Ticona, Cesar; Abarca, Susan; Heredia, Paula; Aguirre, Andres; Sterling, Timothy R; Garvy, Beth A; Fichtenbaum, Carl J. Latent tuberculosis infection is associated with increased unstimulated levels of interferon-gamma in Lima, Peru.. PLoS ONE, 2018; 13(9):e0202191

Huangfu, P; Laurence, Y V; Alisjahbana, B; Ugarte-Gil, C; Riza, A-L; Walzl, G; Ruslami, R; Moore, D A J; Ioana, M; McAllister, S; Ronacher, K; Koesoemadinata, R C; Grint, D; Kerry, S; Coronel, J; Malherbe, S T; Griffiths, U; Dockrell, H M; Hill, P C; van Crevel, R; Pearson, F; Critchley, J A. Point of care HbA1c level for diabetes mellitus management and its accuracy among tuberculosis patients: a study in four countries.. Int J Tuberc Lung Dis, 2019; 23(3):283-292

Hussein M.T.; Yousef L.M.; Abusedera M.A.. Pattern of pulmonary tuberculosis in elderly patients in Sohag Governorate: Hospital based study. Egypt. J. Chest Dis. Tuberc., 2013; 62(2):269-274

Irmawati; Ridwan A.; Ansar J.. Risk factors of pulmonary tuberculosis relapse in working population of Makassar City. Indian J. Public Health Res. Dev., 2019; 10(7):1140-1144

Jaber A.A.S.; Ibrahim B.. Outcome of interim multidrug-resistant tuberculosis treatment in Yemen. Trop. J. Pharm. Res., 2019; 18(8):1755-1762

Jaber A.A.S.; Khan A.H.; Sulaiman S.A.S.. Evaluating treatment outcomes and durations among cases of smear-positive pulmonary tuberculosis in Yemen: A prospective follow-up study. J. pharm. policy pract., 2017; 10(1):36

Jaber, Ammar Ali Saleh; Ibrahim, Baharudin. Evaluation of risk factors associated with drug-resistant tuberculosis in Yemen: data from centres with high drug resistance.. BMC Infect Dis, 2019; 19(1):464

Jacobs, Marina Gasino; Pinto Junior, Vitor Laerte. Characterization of drug-resistant tuberculosis in Brazil, 2014. EPIDEMIOLOGIA E SERVICOS DE SAUDE, 2019; 28(3):

Jacobs, Marina Gasino; Pinto Junior, Vitor Laerte. Characterization of drug-resistant tuberculosis in Brazil, 2014.. Epidemiol. serv. saude, 2020; 28(3):e2018294

Jacobs, Marina Gasino; Pinto Junior, Vitor Laerte. CaracterizaÃ§Ã£o da tuberculose drogarresistente no Brasil, 2014. Epidemiol. serv. saÃºde, 2019; 28(3):e2018294-e2018294

Jafari N.J.; Izadi M.; Sarrafzadeh F.; Heidari A.; Ranjbar R.; Saburi A.. Hyponatremia due to pulmonary tuberculosis: Review of 200 cases. Nephro Urol. Mon., 2013; 5(1):687-691

Jain, Ankit; Suma, K. R.. CLINICAL, LABORATORY AND RADIOLOGICAL PROFILE OF RE-TREATMENT TUBERCULOSIS. JOURNAL OF EVOLUTION OF MEDICAL AND DENTAL SCIENCES-JEMDS, 2018; 7(18):2218-2221

Jain, Swapnil; Varudkar, H G; Julka, Arti; Singapurwala, Mustafa; Khosla, Sourav; Shah, Bhavya. Socio-economical and Clinico-Radiological Profile of 474 MDR TB Cases of a Rural Medical College.. J Assoc Physicians India, 2018; 66(12):14-18

Jali M.V.; Mahishale V.K.; Hiremath M.B.; Satyanarayana S.; Kumar A.M.V.; Nagaraja S.B.; Isaakidis P.. Diabetes mellitus and smoking among tuberculosis patients in a tertiary care centre in Karnataka, India. Public Health Action, 2013; 3(SUPPL.1):S51-S53

James S.L.; Abate D.; Abate K.H.; Abay S.M.; Abbafati C.; Abbasi N.; Abbastabar H.; Abd-Allah F.; Abdela J.; Abdelalim A.; Abdollahpour I.; Abdulkader R.S.; Abebe Z.; Abera S.F.; Abil O.Z.; Abraha H.N.; Abu-Raddad L.J.; Abu-Rmeileh N.M.E.; Accrombessi M.M.K.; Acharya D.; Acharya P.; Ackerman I.N.; Adamu A.A.; Adebayo O.M.; Adekanmbi V.; Adetokunboh O.O.; Adib M.G.; Adsuar J.C.; Afanvi K.A.; Afarideh M.; Afshin A.; Agarwal G.; Agesa K.M.; Aggarwal R.; Aghayan S.A.; Agrawal S.; Ahmadi A.; Ahmadi M.; Ahmadieh H.; Ahmed M.B.; Aichour A.N.; Aichour I.; Aichour

M.T.E.; Akinyemiju T.; Akseer N.; Al-Aly Z.; Al-Eyadhy A.; Al-Mekhlafi H.M.; Al-Raddadi R.M.; Alahdab F.; Alam K.; Alam T.; Alashi A.; Alavian S.M.; Alene K.A.; Alijanzadeh M.; Alizadeh-Navaei R.; Aljunid S.M.; Alkerwi A.; Alla F.; Allebeck P.; Alouani M.M.L.; Altirkawi K.; Alvis-Guzman N.; Amare A.T.; Aminde L.N.; Ammar W.; Amoako Y.A.; Anber N.H.; Andrei C.L.; Androudi S.; Animut M.D.; Anjomshoa M.; Ansha M.G.; Antonio C.A.T.; Anwari P.; Arabloo J.; Arauz A.; Aremu O.; Ariani F.; Armoon B.; Arnlov J.; Arora A.; Artaman A.; Aryal K.K.; Asayesh H.; Asghar R.J.; Ataro Z.; Atre S.R.; Ausloos M.; Avila-Burgos L.; Avokpaho E.F.G.A.; Awasthi A.; Ayala Quintanilla B.P.; Ayer R.; Azzopardi P.S.; Babazadeh A.; Badali H.; Badawi A.; Bali A.G.; Ballesteros K.E.; Ballew S.H.; Banach M.; Banoub J.A.M.; Banstola A.; Barac A.; Barboza M.A.; Barker-Collo S.L.; Barnighausen T.W.; Barrero L.H.; Baune B.T.; Bazargan-Hejazi S.; Bedi N.; Beghi E.; Behzadifar M.; Bejot Y.; Belachew A.B.; Belay Y.A.; Bell M.L.; Bello A.K.; Bensenor I.M.; Bernabe E.; Bernstein R.S.; Beuran M.; Beyranvand T.; Bhala N.; Bhattarai S.; Bhaumik S.; Bhutta Z.A.; Biadgo B.; Bijani A.; Bikbov B.; Bilano V.; Bililign N.; Bin Sayeed M.S.; Bisanzio D.; Blacker B.F.; Blyth F.M.; Bou-Orm I.R.; Boufous S.; Bourne R.; Brady O.J.; Brainin M.; Brant L.C.; Brazinova A.; Breitborde N.J.K.; Brenner H.; Briant P.S.; Briggs A.M.; Briko A.N.; Britton G.; Brugha T.; Buchbinder R.; Busse R.; Butt Z.A.; Cahuana-Hurtado L.; Cano J.; Cardenas R.; Carrero J.J.; Carter A.; Carvalho F.; Castaneda-Orjuela C.A.; Castillo Rivas J.; Castro F.; Catala-Lopez F.; Cercy K.M.; Cerin E.; Chaiah Y.; Chang A.R.; Chang H.-Y.; Chang J.-C.; Charlson F.J.; Chattopadhyay A.; Chattu V.K.; Chaturvedi P.; Chiang P.P.-C.; Chin K.L.; Chitheer A.; Choi J.-Y.J.; Chowdhury R.; Christensen H.; Christopher D.J.; Cicuttini F.M.; Ciobanu L.G.; Cirillo M.; Claro R.M.; Collado-Mateo D.; Cooper C.; Coresh J.; Cortesi P.A.; Cortinovis M.; Costa M.; Cousin E.; Criqui M.H.; Cromwell E.A.; Cross M.; Crump J.A.; Dadi A.F.; Dandona L.; Dandona R.; Dargan P.I.; Daryani A.; Das Gupta R.; Das Neves J.; Dasa T.T.; Davey G.; Davis A.C.; Davitoiu D.V.; De Courten B.; De La Hoz F.P.; De Leo D.; De Neve J.-W.; Degefa M.G.; Degenhardt L.; Deiparine S.; Dellavalle R.P.; Demoz G.T.; Deribe K.; Dervenis N.; Des Jarlais D.C.; Dessie G.A.; Dey S.; Dharmaratne S.D.; Dinberu M.T.; Dirac M.A.; Djalalinia S.; Doan L.; Dokova K.; Doku D.T.; Dorsey E.R.; Doyle K.E.; Driscoll T.R.; Dubey M.; Dubljanin E.; Duken E.E.; Duncan B.B.; Duraes A.R.; Ebrahimi H.; Ebrahimpour S.; Echko M.M.; Edvardsson D.; Effiong A.; Ehrlich J.R.; El Bcheraoui C.; El Sayed Zaki M.; El-Khatib Z.; Elkout H.; Elyazar I.R.F.; Enayati A.; Endries A.Y.; Er B.; Erskine H.E.; Eshrati B.; Eskandarieh S.; Esteghamati A.; Esteghamati S.; Fakhim H.; Fallah Omrani V.; Faramarzi M.; Fareed M.; Farhadi F.; Farid T.A.; Farinha C.S.E.; Farioli A.; Faro A.; Farvid M.S.; Farzadfar F.; Feigin V.L.; Fentahun N.; Fereshtehnejad S.-M.; Fernandes E.; Fernandes J.C.; Ferrari A.J.; Feyissa G.T.; Filip I.; Fischer F.; Fitzmaurice C.; Foigt N.A.; Foreman K.J.; Fox J.; Frank T.D.; Fukumoto T.; Fullman N.; Furst T.; Furtado J.M.; Futran N.D.; Gall S.; Ganji M.; Gankpe F.G.; Garcia-Basteiro A.L.; Gardner W.M.; Gebre A.K.; Gebremedhin A.T.; Gebremichael T.G.; Gelano T.F.; Geleijnse J.M.; Genova-Maleras R.; Geramo Y.C.D.; Gething P.W.; Gezae K.E.; Ghadiri K.; Ghasemi Falavarjani K.; Ghasemi-Kasman M.; Ghimire M.; Ghosh R.; Ghoshal A.G.; Giampaoli S.; Gill P.S.; Gill T.K.; Ginawi I.A.; Giussani G.; Gnedovskaya E.V.; Goldberg E.M.; Goli S.; Gomez-Dantes H.; Gona P.N.; Gopalani S.V.; Gorman T.M.; Goulart A.C.; Goulart B.N.G.; Grada A.; Grams M.E.; Grosso G.; Gugnani H.C.; Guo Y.; Gupta P.C.; Gupta R.; Gupta T.; Gyawali B.; Haagsma J.A.; Hachinski V.; Hafezi-Nejad N.; Haghparast Bidgoli H.; Hagos T.B.; Hailu G.B.; Haj-Mirzaian A.; Hamadeh R.R.; Hamidi S.; Handal A.J.; Hankey G.J.; Hao Y.; Harb H.L.; Harikrishnan S.; Haro J.M.; Hasan M.; Hassankhani H.; Hassen H.Y.; Havmoeller R.; Hawley C.N.; Hay R.J.; Hay S.I.; Hedayatizadeh-Omran A.; Heibati B.; Hendrie D.; Henok A.; Herteliu C.; Heydarpour S.; Hibstu D.T.; Hoang H.T.; Hoek H.W.; Hoffman H.J.; Hole M.K.; Homaie Rad E.; Hoogar P.; Hosgood H.D.; Hosseini S.M.; Hosseinzadeh M.; Hostiuc M.; Hostiuc S.; Hotez P.J.; Hoy D.G.; Hsairi M.; Htet A.S.; Hu G.; Huang J.J.; Huynh C.K.; Iburg K.M.; Ikeda C.T.; Ileanu B.; Ilesanmi O.S.; Iqbal U.; Irvani S.S.N.; Irvine C.M.S.; Islam S.; Islami F.; Jacobsen K.H.; Jahangiry L.; Jahanmehr N.; Jain S.K.; Jakovljevic M.; Javanbakht M.; Jayatilleke A.U.; Jeemon P.; Jha R.P.; Jha V.; Ji J.S.; Johnson C.O.; Jonas J.B.; Jozwiak J.J.; Jungari S.B.; Jurisson M.; Kabir Z.; Kadel R.; Kahsay A.; Kalani R.; Kanchan T.; Karami M.; Karami Matin B.; Karch A.; Karema C.; Karimi N.; Karimi S.M.; Kasaeian A.; Kassa D.H.; Kassa G.M.; Kassa T.D.; Kassebaum N.J.; Katikireddi S.V.; Kawakami N.; Kazemi Karyani A.; Keighobadi M.M.; Keiyoro P.N.; Kemmer L.; Kemp G.R.; Kengne A.P.; Keren A.; Khader Y.S.; Khafaei B.; Khafaie M.A.; Khajavi A.; Khalil I.A.; Khan E.A.; Khan M.S.; Khan M.A.; Khang Y.-H.; Khazaei M.; Khoja A.T.; Khosravi A.; Khosravi M.H.; Kiadaliri A.A.; Kiirithio D.N.; Kim C.-I.; Kim D.; Kim P.; Kim Y.-E.; Kim Y.J.; Kimokoti R.W.; Kinfu Y.; Kisa A.; Kissimova-Skarbek K.; Kivimaki M.; Knudsen A.K.S.;

Kocarnik J.M.; Kochhar S.; Kokubo Y.; Kolola T.; Kopec J.A.; Kosen S.; Kotsakis G.A.; Koul P.A.; Koyanagi A.; Kravchenko M.A.; Krishan K.; Krohn K.J.; Kuate Defo B.; Kucuk Bicer B.; Kumar G.A.; Kumar M.; Kyu H.H.; Lad D.P.; Lad S.D.; Lafranconi A.; Lalloo R.; Lallukka T.; Lami F.H.; Lansingh V.C.; Latifi A.; Lau K.M.-M.; Lazarus J.V.; Leasher J.L.; Ledesma J.R.; Lee P.H.; Leigh J.; Leung J.; Levi M.; Lewycka S.; Li S.; Li Y.; Liao Y.; Liben M.L.; Lim L.-L.; Lim S.S.; Liu S.; Lodha R.; Looker K.J.; Lopez A.D.; Lorkowski S.; Lotufo P.A.; Low N.; Lozano R.; Lucas T.C.D.; Lucchesi L.R.; Lunevicius R.; Lyons R.A.; Ma S.; Macarayan E.R.K.; Mackay M.T.; Madotto F.; Magdy Abd El Razek H.; Magdy Abd El Razek M.; Maghavani D.P.; Mahotra N.B.; Mai H.T.; Majdan M.; Majdzadeh R.; Majeed A.; Malekzadeh R.; Malta D.C.; Mamun A.A.; Manda A.-L.; Manguerra H.; Manhertz T.; Mansournia M.A.; Mantovani L.G.; Mapoma C.C.; Maravilla J.C.; Marcenes W.; Marks A.; Martins-Melo F.R.; Martopullo I.; Marz W.; Marzan M.B.; Mashamba-Thompson T.P.; Massenburg B.B.; Mathur M.R.; Matsushita K.; Maulik P.K.; Mazidi M.; McAlinden C.; McGrath J.J.; McKee M.; Mehndiratta M.M.; Mehrotra R.; Mehta K.M.; Mehta V.; Mejia-Rodriguez F.; Mekonen T.; Melese A.; Melku M.; Meltzer M.; Memiah P.T.N.; Memish Z.A.; Mendoza W.; Mengistu D.T.; Mengistu G.; Mensah G.A.; Mereta S.T.; Meretoja A.; Meretoja T.J.; Mestrovic T.; Mezerji N.M.G.; Miazgowski B.; Miazgowski T.; Millear A.I.; Miller T.R.; Miltz B.; Mini G.K.; Mirarefin M.; Mirrakhimov E.M.; Misganaw A.T.; Mitchell P.B.; Mitiku H.; Moazen B.; Mohajer B.; Mohammad K.A.; Mohammadifard N.; Mohammadnia-Afrouzi M.; Mohammed M.A.; Mohammed S.; Mohebi F.; Moitra M.; Mokdad A.H.; Molokhia M.; Monasta L.; Moodley Y.; Moosazadeh M.; Moradi G.; Moradi-Lakeh M.; Moradinazar M.; Moraga P.; Morawska L.; Moreno Velasquez I.; Morgado-Da-Costa J.; Morrison S.D.; Moschos M.M.; Mousavi S.M.; Mruts K.B.; Muche A.A.; Muchie K.F.; Mueller U.O.; Muhammed O.S.; Mukhopadhyay S.; Muller K.; Mumford J.E.; Murhekar M.; Musa J.; Musa K.I.; Mustafa G.; Nabhan A.F.; Nagata C.; Naghavi M.; Naheed A.; Nahvijou A.; Naik G.; Naik N.; Najafi F.; Naldi L.; Nam H.S.; Nangia V.; Nansseu J.R.; Nascimento B.R.; Natarajan G.; Neamati N.; Negoi I.; Negoi R.I.; Neupane S.; Newton C.R.J.; Ngunjiri J.W.; Nguyen A.Q.; Nguyen H.L.T.; Nguyen H.T.; Nguyen L.H.; Nguyen M.; Nguyen N.B.; Nguyen S.H.; Nichols E.; Ningrum D.N.A.; Nixon M.R.; Nolutshungu N.; Nomura S.; Norheim O.F.; Noroozi M.; Norrving B.; Noubiap J.J.; Nouri H.R.; Nourollahpour Shiadeh M.; Nowroozi M.R.; Nsoesie E.O.; Nyasulu P.S.; Odell C.M.; Ofori-Asenso R.; Ogbo F.A.; Oh I.-H.; Oladimeji O.; Olagunju A.T.; Olagunju T.O.; Olivares P.R.; Olsen H.E.; Olusanya B.O.; Ong K.L.; Ong S.K.; Oren E.; Ortiz A.; Ota E.; Otstavnov S.S.; overland S.; Owolabi M.O.; Mahesh P.A.; Pacella R.; Pakpour A.H.; Pana A.; Panda-Jonas S.; Parisi A.; Park E.-K.; Parry C.D.H.; Patel S.; Pati S.; Patil S.T.; Patle A.; Patton G.C.; Paturi V.R.; Paulson K.R.; Pearce N.; Pereira D.M.; Perico N.; Pesudovs K.; Pham H.Q.; Phillips M.R.; Pigott D.M.; Pillay J.D.; Piradov M.A.; Pirsaheb M.; Pishgar F.; Plana-Ripoll O.; Plass D.; Polinder S.; Popova S.; Postma M.J.; Pourshams A.; Poustchi H.; Prabhakaran D.; Prakash S.; Prakash V.; Purcell C.A.; Purwar M.B.; Qorbani M.; Quistberg D.A.; Radfar A.; Rafay A.; Rafiei A.; Rahim F.; Rahimi K.; Rahimi-Movaghar A.; Rahimi-Movaghar V.; Rahman M.; Ur Rahman M.H.; Rahman M.A.; Rahman S.U.; Rai R.K.; Rajati F.; Ram U.; Ranjan P.; Ranta A.; Rao P.C.; Rawaf D.L.; Rawaf S.; Reddy K.S.; Reiner R.C.; Reinig N.; Reitsma M.B.; Remuzzi G.; Renzaho A.M.N.; Resnikoff S.; Rezaei S.; Rezai M.S.; Ribeiro A.L.P.; Robinson S.R.; Roever L.; Ronfani L.; Roshandel G.; Rostami A.; Roth G.A.; Roy A.; Rubagotti E.; Sachdev P.S.; Sadat N.; Saddik B.; Sadeghi E.; Saeedi Moghaddam S.; Safari H.; Safari Y.; Safari-Faramani R.; Safdarian M.; Safi S.; Safiri S.; Sagar R.; Sahebkar A.; Sahraian M.A.; Sajadi H.S.; Salam N.; Salama J.S.; Salamati P.; Saleem K.; Saleem Z.; Salimi Y.; Salomon J.A.; Salvi S.S.; Salz I.; Samy A.M.; Sanabria J.; Sang Y.; Santomauro D.F.; Santos I.S.; Santos J.V.; Santric Milicevic M.M.; Sao Jose B.P.; Sardana M.; Sarker A.R.; Sarrafzadegan N.; Sartorius B.; Sarvi S.; Sathian B.; Satpathy M.; Sawant A.R.; Sawhney M.; Saxena S.; Saylan M.; Schaeffner E.; Schmidt M.I.; Schneider I.J.C.; Schottker B.; Schwebel D.C.; Schwendicke F.; Scott J.G.; Sekerija M.; Sepanlou S.G.; Servan-Mori E.; Seyedmousavi S.; Shabaninejad H.; Shafieesabet A.; Shahbazi M.; Shaheen A.A.; Shaikh M.A.; Shams-Beyranvand M.; Shamsi M.; Shamsizadeh M.; Sharafi H.; Sharafi K.; Sharif M.; Sharif-Alhoseini M.; Sharma M.; Sharma R.; She J.; Sheikh A.; Shi P.; Shibuya K.; Shigematsu M.; Shiri R.; Shirkoohi R.; Shishani K.; Shiue I.; Shokraneh F.; Shoman H.; Shrime M.G.; Si S.; Siabani S.; Siddiqi T.J.; Sigfusdottir I.D.; Sigurvinsdottir R.; Silva J.P.; Silveira D.G.A.; Singam N.S.V.; Singh J.A.; Singh N.P.; Singh V.; Sinha D.N.; Skiadaresi E.; Slepak E.L.N.; Sliwa K.; Smith D.L.; Smith M.; Soares Filho A.M.; Sobaih B.H.; Sobhani S.; Sobngwi E.; Soneji S.S.; Soofi M.; Soosaraei M.; Sorensen R.J.D.; Soriano J.B.; Soyiri I.N.; Sposato L.A.; Sreeramareddy C.T.; Srinivasan V.; Stanaway J.D.; Stein D.J.; Steiner C.; Steiner T.J.; Stokes M.A.;

Stovner L.J.; Subart M.L.; Sudaryanto A.; Sufiyan M.B.; Sunguya B.F.; Sur P.J.; Sutradhar I.; Sykes B.L.; Sylte D.O.; Tabares-Seisdedos R.; Tadakamadla S.K.; Tadesse B.T.; Tandon N.; Tassew S.G.; Tavakkoli M.; Taveira N.; Taylor H.R.; Tehrani-Banihashemi A.; Tekalign T.G.; Tekelemedhin S.W.; Tekle M.G.; Temesgen H.; Temsah M.-H.; Temsah O.; Terkawi A.S.; Teweldemedhin M.; Thankappan K.R.; Thomas N.; Tilahun B.; To Q.G.; Tonelli M.; Topor-Madry R.; Topouzis F.; Torre A.E.; Tortajada-Girbes M.; Touvier M.; Tovani-Palone M.R.; Towbin J.A.; Tran B.X.; Tran K.B.; Troeger C.E.; Truelsen T.C.; Tsilimbaris M.K.; Tsoi D.; Tudor Car L.; Tuzcu E.M.; Ukwaja K.N.; Ullah I.; Undurraga E.A.; Unutzer J.; Updike R.L.; Usman M.S.; Uthman O.A.; Vaduganathan M.; Vaezi A.; Valdez P.R.; Varughese S.; Vasankari T.J.; Venketasubramanian N.; Villafaina S.; Violante F.S.; Vladimirov S.K.; Vlassov V.; Vollset S.E.; Vosoughi K.; Vujcic I.S.; Wagnew F.S.; Waheed Y.; Waller S.G.; Wang Y.; Wang Y.-P.; Weiderpass E.; Weintraub R.G.; Weiss D.J.; Weldegebreal F.; Weldegwergs K.G.; Werdecker A.; West T.E.; Whiteford H.A.; Widecka J.; Wijeratne T.; Wilner L.B.; Wilson S.; Winkler A.S.; Wiyeh A.B.; Wiysonge C.S.; Wolfe C.D.A.; Woolf A.D.; Wu S.; Wu Y.-C.; Wyper G.M.A.; Xavier D.; Xu G.; Yadgir S.; Yadollahpour A.; Yahyazadeh Jabbari S.H.; Yamada T.; Yan L.L.; Yano Y.; Yaseri M.; Yasin Y.J.; Yeshaneh A.; Yimer E.M.; Yip P.; Yisma E.; Yonemoto N.; Yoon S.-J.; Yotebieng M.; Younis M.Z.; Yousefifard M.; Yu C.; Zadnik V.; Zaidi Z.; Zaman S.B.; Zamani M.; Zare Z.; Zeleke A.J.; Zenebe Z.M.; Zhang K.; Zhao Z.; Zhou M.; Zodpey S.; Zucker I.; Vos T.; Murray C.J.L.. Global, regional, and national incidence, prevalence, and years lived with disability for 354 Diseases and Injuries for 195 countries and territories, 1990-2017: A systematic analysis for the Global Burden of Disease Study 2017. Lancet, 2018; 392(10159):1789-1858

Janmeja, A K; Aggarwal, Deepak; Dhillon, Ruchika. Factors predicting treatment success in multi-drug resistant tuberculosis patients treated under programmatic conditions.. INDIAN J. TUBERC., 2018; 65(2):135-139

Jetan, C A; Jamaiah, I; Rohela, M; Nissapatorn, V. Tuberculosis: an eight year (2000-2007) retrospective study at the University of Malaya Medical Centre (UMMC), Kuala Lumpur, Malaysia.. Southeast Asian J Trop Med Public Health, 2010; 41(2):378-85

Jitmuang, Anupop; Munjit, Parnwad; Foongladda, Suporn. PREVALENCE AND FACTORS ASSOCIATED WITH MULTIDRUG-RESISTANT TUBERCULOSIS AT SIRIRAJ HOSPITAL, BANGKOK, THAILAND.. Southeast Asian J Trop Med Public Health, 2015; 46(4):697-706

Joaquim, Andrei Fernandes; Carandina, Luana; Defaveri, Julio. Tuberculose em necropsias realizadas no ServiÃ§o de Anatomia PatolÃ³gica da Faculdade de Medicina de Botucatu. J. bras. patol. med. lab, 2006; 42(3):193-200

Junior R.T.; Loffredo L.C.M.. Epidemiological characterization of patients at a tuberculosis hospital in the state of Sao Paulo, Brazil. Rev. Cienc. Farm. Basica Apl., 2015; 36(1):149-152

Kalyesubula, Robert; Mutyaba, Innocent; Rabin, Tracy; Andia-Biraro, Irene; Alupo, Patricia; Kimuli, Ivan; Nabirye, Stella; Kagimu, Magid; Mayanja-Kizza, Harriet; Rastegar, Asghar; Kamya, Moses R. Trends of admissions and case fatality rates among medical in-patients at a tertiary hospital in Uganda; A four-year retrospective study.. PLoS ONE, 2019; 14(5):e0216060

Kassu A.; Mohammad A.; Fujimaki Y.; Moges F.; Elias D.; Mekonnen F.; Mengistu G.; Yamato M.; Wondmikun Y.; Ota F.. Serum IgE levels of tuberculosis patients in a tropical setup with high prevalence of HIV and intestinal parasitoses. Clin. Exp. Immunol., 2004; 138(1):122-127

Kayhan, Servet. Demographic and clinical characteristics of tuberculosis: A report of 2404 cases at a referral hospital. AFRICAN JOURNAL OF MICROBIOLOGY RESEARCH, 2012; 6(9):2033-2037

Kazempour Dizaji, Mehdi; Kazemnejad, Anoshirvan; Tabarsi, Payam; Zayeri, Farid. Risk Factors Associated with Survival of Pulmonary Tuberculosis.. Iran J Public Health, 2018; 47(7):980-987

Kehbila, Jules; Ekabe, Cyril Jabea; Aminde, Leopold Ndemnge; Noubiap, Jean Jacques N; Fon, Peter Nde; Monekosso, Gottlieb Lobe. Prevalence and correlates of depressive symptoms in adult patients with pulmonary tuberculosis in the Southwest Region of Cameroon.. Infect. dis. poverty, 2016; 5(1):51

Kempker, R R; Mikiashvili, L; Zhao, Y; Benkeser, D; Barbakadze, K; Bablishvili, N; Avaliani, Z; Peloquin, C A; Blumberg, H M; Kipiani, M. Clinical Outcomes Among Patients With Drug-resistant Tuberculosis Receiving Bedaquiline- or Delamanid-Containing Regimens. Clin Infect Dis, 2019; 71(9):2336-2344

Khan A.H.; Sulaiman S.A.S.; Hassali M.A.; Khan K.U.; Ming L.C.; Mateen O.; Ullah M.O.. Effect of smoking on treatment outcome among tuberculosis patients in Malaysia; a multicenter study. BMC Public Health, 2020; 20(1):854

Khan A.H.; Sulaiman S.A.S.; Muttalif A.R.; Hassali M.A.; Abdullah R.. Gender differences in the prevalence of tuberculous lymphadenitis at the state of penang, Malaysia: Findings from a cross-sectional study. Arch. Pharm. Pract., 2010; 1(1):44507

Kikvidze M.; Mikiashvili L.. Impact of diabetes mellitus on drug-resistant tuberculosis treatment outcomes in Georgia-cohort study. Eur. Respir. J., 2013; 42(SUPPL. 57):

Kittikraisak, Wanitchaya; Burapat, Channawong; Nateniyom, Sriprapa; Akksilp, Somsak; Mankatittham, Wiroj; Sirinak, Chawin; Suphanam, Arunee; Kanphukiew, Apiratee; Varma, Jay K. Improvements in physical and mental health among HIV-infected patients treated for TB in Thailand.. Southeast Asian J Trop Med Public Health, 2008; 39(6):1061-71

Kliiman, Kai; Altraja, Alan. Predictors of extensively drug-resistant pulmonary tuberculosis.. Ann Intern Med, 2009; 150(11):766-75

Kocakoglu S.; Simsek Z.; Ceylan E.. Epidemiologic characteristics of the tuberculosis cases followed up at Sanliurfa Central Tuberculosi Control Dispensary between 2001 and 2006 years. Turk Toraks Derg., 2009; 10(1):41883

Kottarath, Manoj; Mavila, Rajani; V., Achuthan; Nair, Smitha. Prevalence of diabetes mellitus in tuberculosis patients: a hospital based study. International Journal of Research in Medical Sciences, 2015; ():

Kumar, Nathella P; Moideen, Kadar; Viswanathan, Vijay; Shruthi, Basavaradhya S; Sivakumar, Shanmugam; Menon, Pradeep A; Kornfeld, Hardy; Babu, Subash. Elevated levels of matrix metalloproteinases reflect severity and extent of disease in tuberculosis-diabetes co-morbidity and are predominantly reversed following standard anti-tuberculosis or metformin treatment.. BMC Infect Dis, 2018; 18(1):345

LÃ³pez, Jenny; Hermoso, Roland; Loreto, Andrea; Ovalles, Valentina; Quintini, JosÃ©. CaracterÃsticas clÃnicas, paraclÃnicas y sociodemogrÃ¡ficas en pacientes con tuberculosis en la regiÃ³n de Barlovento entre 1998-2002. Centro mÃ©d, 2003; 48(2):62-65

Lagonegro, Eduardo Ronner. Co-infecÃ§Ã£o tuberculose HIV/AIDS. Analise do momento do diagnÃ³stico e prognÃ³stico. , 2000; ():93-93

Lakadamyali H.; Ergun T.; Ozkara S.. Factors influencing sputum smear and culture positivity in the diagnosis of pulmonary tuberculosis: How many specimens do we need for diagnosis?. J. Clin. Anal. Med., 2011; 2(1):25-28

Lalkhen, Hoosain; Mash, Robert. Multimorbidity in non-communicable diseases in South African primary healthcare.. SAMJ, S. Afr. med. j., 2015; 105(2):134-8

Laxmeshwar, C; Stewart, A G; Dalal, A; Kumar, A M V; Kalaiselvi, S; Das, M; Gawde, N; Thi, S S; Isaakidis, P. Beyond 'cure' and 'treatment success': quality of life of patients with multidrug-resistant tuberculosis.. Int J Tuberc Lung Dis, 2019; 23(1):73-81

Le, Huy Ngoc; Sriplung, Hutcha; Chongsuvivatwong, Virasakdi; Nguyen, Nhung Viet; Nguyen, Tri Huu. The accuracy of tuberculous meningitis diagnostic tests using Bayesian latent class analysis. The Journal of Infection in Developing Countries, 2020; 14(5):479-487

Lee, Gwenyth O; Comina, German; Hernandez-Cordova, Gustavo; Naik, Nehal; Gayoso, Oscar; Ticona, Eduardo; Coronel, Jorge; Evans, Carlton A; Zimic, Mirko; Paz-Soldan, Valerie A; Gilman,

Robert H; Oberhelman, Richard. Cough dynamics in adults receiving tuberculosis treatment.. PLoS ONE, 2020; 15(6):e0231167

Leite, Ricardo Costa. Intervalo do tempo decorrido entre a investigaÃ§Ã£o diagnÃ³stica laboratorial e o inÃcio do tratamento em casos de tuberculose pulmonar em um distrito da atenÃ§Ã£o primÃ¡ria de saÃºde em Recife - PE. , 2016; ():87-87

Lesnic E.; Caraiani O.; Zlepca V.. Features and treatment outcome in caseous pneumonia. Eur. Respir. J., 2012; 40(SUPPL. 56):

Lindoso, Ana AngÃ©lica BulcÃ£o Portela; Waldman, Eliseu Alves; Komatsu, Naomi Kawaoka; Figueiredto, Sumie Matai de; Taniguchi, Mauro; Rodrigues, Laura C. Perfil de pacientes que evoluem para Ã³bito por tuberculose no municÃpio de SÃ£o Paulo, 2002. Rev. saÃºde pÃºblica, 2008; 42(5):805-812

Lindoso, Ana Angelica Bulcao Portela; Waldman, Eliseu Alves; Komatsu, Naomi Kawaoka; Figueiredo, Sumie Matai de; Taniguchi, Mauro; Rodrigues, Laura C. Profile of tuberculosis patients progressing to death, city of Sao Paulo, Brazil, 2002.. Rev Saude Publica, 2008; 42(5):805-12

Lins, Tatiana Bacelar Acioli; Soares, Eldom de Medeiros; dos Santos, Felipe Macedo; Mandacaru, Polyana Maria Pimenta; Pina, Tatiana; de Araujo Filho, Joao Alves. Mycobacterium tuberculosis and human immunodeficiency virus coinfection in a tertiary care hospital in Midwestern Brazil.. Infez Med, 2012; 20(2):108-16

Lisha, P V; James, P T; Ravindran, C. Morbidity and mortality at five years after initiating Category I treatment among patients with new sputum smear positive pulmonary tuberculosis.. INDIAN J. TUBERC., 2012; 59(2):83-91

Liu, Y; Zheng, Y; Chen, J; Shi, Y; Shan, L-Y; Wang, S; Wang, W-B; Shen, X; Zhang, Y. Tuberculosis-associated mortality and its risk factors in a district of Shanghai, China: a retrospective cohort study.. Int J Tuberc Lung Dis, 2018; 22(6):655-660

Loua, C. L.; Millogo, T.; Baguiya, A.; MÃ©da, B.; Coulibaly, A.; Kouanda, S.. Facteurs associÃ©s aux dÃ©cÃ¨s des patients atteints de tuberculose pulmonaire au service de pneumo phtisiologie du Centre hospitalier universitaire Yalgado OuÃ©draogo de Ouagadougou, Burkina Faso : Ã©tude cas tÃ©moin. Science et Technique, Sciences de la SantÃ©, 2017; ():

Louw, Julia; Peltzer, Karl; Naidoo, Pamela; Matseke, Gladys; Mchunu, Gugu; Tutshana, Bomkazi. Quality of life among tuberculosis (TB), TB retreatment and/or TB-HIV co-infected primary public health care patients in three districts in South Africa.. Health Qual Life Outcomes, 2012; 10(101153626):77

Machado, N; Grant, C S; Scrimgeour, E. Abdominal tuberculosis--experience of a University hospital in Oman.. Acta Trop, 2001; 80(2):187-90

Maharaj, B; Leary, W P; Pudifin, D J. A prospective study of hepatic tuberculosis in 41 black patients.. Q J Med, 1987; 63(242):517-22

Mahshid Talebi, Taher; Seyed Ali Javad, Moosavi; Mehdi, Pourghasemian. [Comparing pulmonary tuberculosis between elderly patients and young adults]. Razi J. Med. Sci., 2011; 18(88):30-35

Makhlouf H.A.; Helmy A.; Fawzy E.; El-Attar M.; Rashed H.A.G.. A prospective study of antituberculous drug-induced hepatotoxicity in an area endemic for liver diseases. Hepatol. Int., 2008; 2(3):353-360

Marak, Bibha; Kaur, Prabhdeep; Rao, Sudha R; Selvaraju, Sriram. Non-communicable disease comorbidities and risk factors among tuberculosis patients, Meghalaya, India.. INDIAN J. TUBERC., 2016; 63(2):123-5

Maria N.; Radji M.; Burhan E.. The impact of antituberculosis drug-induced hepatotoxicity to succesful tuberculosis treatment in Indonesia. Asian J. Pharm. Clin. Res., 2017; 10(11):194-198

Marjani M.; Tabarsi P.; Baghaei P.; Shamaei M.; Biani P.G.; Mansouri D.; Masjedi M.R.; Velayati A.A.. Incidence of thromboembolism in hospitalized patients with tuberculosis and associated risk factors. Arch. Clin. Infect. Dis., 2012; 7(2):56-59

Marjani, Majid; Baghaei, Parvaneh; Malekmohammad, Majid; Tabarsi, Payam; Sharif-Kashani, Babak; Behzadnia, Neda; Mansouri, Davood; Masjedi, Mohammad Reza; Velayati, Ali Akbar. Effect of pulmonary hypertension on outcome of pulmonary tuberculosis. Braz. j. infect. dis, 2014; 18(5):487-490

MartÃnez de Cuellar, Celia; Lovera, Dolores; Gatti, Luis; Ojeda, Limpia; Apodaca, Silvio; Zarate, Claudia; Arbo, Antonio. Tuberculosis: Factores de riesgo asociados a mortalidad en pacientes ≤19 aÃ±os hospitalizados en el Instituto de Medicina Tropical. Pediatr. (AsunciÃ³n), 2019; 46(2):77-81

Matias CoÃªlho, Danieli Maria; Machado Moita Neto, JosÃ©; Campelo, Viriato. Comorbidades e estilo de vida de idosos com tuberculose. Rev. bras. promoÃ§. saÃºde (Impr.), 2014; 27(3):

Matos, E D; Moreira Lemos, A C. Association between serum albumin levels and in-hospital deaths due to tuberculosis.. Int J Tuberc Lung Dis, 2006; 10(12):1360-6

Matos, Eliana Dias; Lemos, AntÃ´nio Carlos Moreira; Bittencourt, Carolina; Mesquita, Cristiane Leite. Anti-tuberculosis drug resistance in strains of Mycobacterium tuberculosis isolated from patients in a tertiary hospital in Bahia. Braz. j. infect. dis, 2007; 11(3):331-338

Mbatchou Ngahane, Bertrand Hugo; Nouyep, Junior; Nganda Motto, Malea; Mapoure Njankouo, Yacouba; Wandji, Adeline; Endale, Mireille; Afane Ze, Emmanuel. Post-tuberculous lung function impairment in a tuberculosis reference clinic in Cameroon.. Respir Med, 2016; 114(8908438, rme):67-71

McLachlan, Irene; Visser, Willem I; Jordaan, H Francois. Skin conditions in a South African tuberculosis hospital: Prevalence, description, and possible associations.. Int J Dermatol, 2016; 55(11):1234-1241

Mekonnen, Habtamu Sewunet; Azagew, Abere Woretaw. Non-adherence to anti-tuberculosis treatment, reasons and associated factors among TB patients attending at Gondar town health centers, Northwest Ethiopia.. BMC Res Notes, 2018; 11(1):691

Merid, Mehari Woldemariam; Gezie, Lemma Derseh; Kassa, Getahun Molla; Muluneh, Atalay Goshu; Akalu, Temesgen Yihunie; Yenit, Melaku Kindie. Incidence and predictors of major adverse drug events among drug-resistant tuberculosis patients on second-line anti-tuberculosis treatment in Amhara regional state public hospitals; Ethiopia: a retrospective cohort study.. BMC Infect Dis, 2019; 19(1):286

Micheletti, Vania Celina Dezoti; Kritski, Afranio Lineu; Braga, Jose Ueleres. Clinical Features and Treatment Outcomes of Patients with Drug-Resistant and Drug-Sensitive Tuberculosis: A Historical Cohort Study in Porto Alegre, Brazil.. PLoS ONE, 2016; 11(8):e0160109

Miklausic B.; Dusek D.; Civljak R.; Oljaca Pribanic M.; Copois M.; Skocibusic S.; Zmak L.; Katalinic-Jankovic V.; Barsic B.. Tuberculous meningitis in adults: Results from a 12-year retrospective study. Infektol. Glas., 2013; 33(2):73-78

Milian, F; Sanchez, L M; Toledo, P; Ramirez, C; Santillan, M A. Descriptive study of human and bovine tuberculosis in Queretaro, Mexico.. Rev Latinoam Microbiol, 2000; 42(1):44452

Misra, Usha K; Kalita, Jayantee; Bhoi, Sanjeev K; Singh, Rajesh K. A study of hyponatremia in tuberculous meningitis.. J Neurol Sci, 2016; 367(jbj, 0375403):152-7

Mitnick, Carole D; Franke, Molly F; Rich, Michael L; Alcantara Viru, Felix A; Appleton, Sasha C; Atwood, Sidney S; Bayona, Jaime N; Bonilla, Cesar A; Chalco, Katiuska; Fraser, Hamish S F; Furin, Jennifer J; Guerra, Dalia; Hurtado, Rocio M; Joseph, Keith; Llaro, Karim; Mestanza, Lorena; Mukherjee, Joia S; Munoz, Maribel; Palacios, Eda; Sanchez, Epifanio; Seung, Kwonjune J; Shin, Sonya S; Sloutsky, Alexander; Tolman, Arielle W; Becerra, Mercedes C. Aggressive regimens for multidrug-resistant tuberculosis decrease all-cause mortality.. PLoS ONE, 2013; 8(3):e58664

Modongo, Chawangwa; Pasipanodya, Jotam G; Magazi, Beki T; Srivastava, Shashikant; Zetola, Nicola M; Williams, Scott M; Sirugo, Giorgio; Gumbo, Tawanda. Artificial Intelligence and Amikacin

Exposures Predictive of Outcomes in Multidrug-Resistant Tuberculosis Patients.. Antimicrob Agents Chemother, 2016; 60(10):5928-32

Molla, Alemayehu; Mengesha, Atikilt; Derajew, Habtamu; Kerebih, Habtamu. Suicidal Ideation, Attempt, and Associated Factors among Patients with Tuberculosis in Ethiopia: A Cross-Sectional Study.. psychiatry j., 2019; 2019(101607838):4149806

Mona, Ahmad; Hossam, Abdel Hamid; Marwa, AL Makawy. <The> impact of comorbidities on the outcome of tuberculous patient in respiratory intensive care unit. Egypt. J. Hosp. Med., 2017; 68(3):1533-1540

Monteiro, Michele Carreira Miranda; Neves, Denise Duprat; Signorini, Dario JosÃ© Hart Pontes; Silva, Isabelle Beatriz Dolavale; Dias, Maria da ConceiÃ§Ã£o. CaracterÃsticas sÃ³cio-demogrÃ¡ficas dos pacientes com tuberculose atendidos em um hospital universitÃ¡rio. PulmÃ¤o RJ, 2006; 15(4):228-232

Montiel, Ivonne; Alarcon, Edith; Aguirre, Sarita; Sequera, Guillermo; Marin, Diana. [Factors associated with unsuccessful treatment of patients with drug-sensitive tuberculosis in Paraguay].. Rev Panam Salud Publica, 2020; 44(csl, 9705400):e89

Montufar Andrade, Franco E; Aguilar LondoÃ±o, Carolina; Saldarriaga Acevedo, Carolina; Quiroga Echeverri, Alicia; Builes MontaÃ±o, Carlos E; Mesa Navas, Miguel A; Molina UpegÃ¼i, Olga L; Zuleta TobÃ³n, John J. CaracterÃsticas clÃnicas, factores de riesgo y perfil de susceptibilidad de las infecciones por micobacterias documentadas por cultivo, en un hospital universitario de alta complejidad en MedellÃn (Colombia). Rev. chil. infectol, 2014; 31(6):735-742

Montufar Andrade, Franco E; Aguilar Londono, Carolina; Saldarriaga Acevedo, Carolina; Quiroga Echeverri, Alicia; Builes Montano, Carlos E; Mesa Navas, Miguel A; Molina Upegui, Olga L; Zuleta Tobon, John J. [Clinical features, risk factors and susceptibility profile of mycobacterial infections documented by culture in a university hospital of high complexity in Medellin (Colombia)].. Rev. chil. infectol., 2014; 31(6):735-42

Moreira, Jose; Fochesatto, Jamila Belicanta; Moreira, Ana L; Pereira, Marisa; Porto, Nelson; Hochhegger, Bruno. Pneumonia tuberculosa: um estudo de 59 casos confirmados microbiologicamente. J. bras. pneumol, 2011; 37(2):232-237

Moreira, Jose; Fochesatto, Jamila Belicanta; Moreira, Ana L; Pereira, Marisa; Porto, Nelson; Hochhegger, Bruno. Tuberculous pneumonia: a study of 59 microbiologically confirmed cases.. J Bras Pneumol, 2011; 37(2):232-7

Mounika, K. L.; Swapna, Katla; Jyothirmai, J.; Firdous, Nisa; Ramidi, Prasanna; Sucharitha, Karra. A STUDY OF PERVASIVENESS OF HEPATOTOXICITY AND OTHER SIDE EFFECTS OF ANTI-TUBERCULOSIS DRUGS ON PULMONARY KOCH PATIENTS. INDO AMERICAN JOURNAL OF PHARMACEUTICAL SCIENCES, 2018; 5(5):3954-3959

Mugusi, Ferdinand M; Mehta, Saurabh; Villamor, Eduardo; Urassa, Willy; Saathoff, Elmar; Bosch, Ronald J; Fawzi, Wafaie W. Factors associated with mortality in HIV-infected and uninfected patients with pulmonary tuberculosis.. BMC Public Health, 2009; 9(100968562):409

Musellim, B; Erturan, S; Sonmez Duman, E; Ongen, G. Comparison of extra-pulmonary and pulmonary tuberculosis cases: factors influencing the site of reactivation.. Int J Tuberc Lung Dis, 2005; 9(11):1220-3

Musuenge, Beatrice B; Poda, Ghislain G; Chen, Pei-Chun. Nutritional Status of Patients with Tuberculosis and Associated Factors in the Health Centre Region of Burkina Faso.. Nutrients, 2020; 12(9):

Nagu T.; Ray R.; Munseri P.; Moshiro C.; Shayo G.; Kazema R.; Mugusi F.; Pallangyo K.. Tuberculosis among the elderly in Tanzania: Disease presentation and initial response to treatment. Int. J. Tuberc. Lung Dis., 2017; 21(12):1251-1257

Naidoo, Pamela; Peltzer, Karl; Louw, Julia; Matseke, Gladys; McHunu, Gugu; Tutshana, Bomkazi. Predictors of tuberculosis (TB) and antiretroviral (ARV) medication non-adherence in public primary care patients in South Africa: a cross sectional study.. BMC Public Health, 2013; 13(100968562):396

Nansera D.; Bajunirwe F.; Elyanu P.; Asiimwe C.; Amanyire G.; Graziano F.M.. Mortality and loss to follow-up among tuberculosis and HIV co-infected patients in rural southwestern Uganda. Int. J. Tuberc. Lung Dis., 2012; 16(10):1371-1376

Ncube, R T; Dube, S A; Machekera, S M; Timire, C; Zishiri, C; Charambira, K; Mapuranga, T; Duri, C; Sandy, C; Dlodlo, R A; Lin, Y. Feasibility and yield of screening for diabetes mellitus among tuberculosis patients in Harare, Zimbabwe.. Public health action, 2019; 9(2):72-77

Nik Nor Ronaidi NM; Mohd NS; Wan Mohammad Z; Sharina D; Nik Rosmawati NH. Factors Associated with Unsuccessful Treatment Outcome of Pulmonary Tuberculosis in Kota Bharu, Kelantan. Malaysian Journal of Public Health Medicine, 2011; ():42156

No authorship indicated. Niger 2012 DHS.. Studies in Family Planning, 2014; 45(3):389-398

Oliveira, Hedi Marinho de Melo Guedes de; Brito, Rossana Coimbra; Kritski, Afranio Lineu; Ruffino-Netto, Antonio. Perfil epidemiolÃ³gico de pacientes portadores de TB internados em um hospital de referÃªncia na cidade do Rio de Janeiro. J. bras. pneumol, 2009; 35(8):780-787

Oliveira, Hedi Marinho de Melo Guedes de; Brito, Rossana Coimbra; Kritski, Afranio Lineu; Ruffino-Netto, Antonio. Epidemiological profile of hospitalized patients with TB at a referral hospital in the city of Rio de Janeiro, Brazil.. J Bras Pneumol, 2009; 35(8):780-7

Oliveira, Marina Gribel; Delogo, Karina Neves; Oliveira, Hedi Marinho de Melo Gomes de; Ruffino-Netto, Antonio; Kritski, Afranio Lineu; Oliveira, Martha Maria. Anemia in hospitalized patients with pulmonary tuberculosis. J. bras. pneumol, 2014; 40(4):403-410

Oliveira, Simoni Pimenta de; Silveira, Juliana Taques Pessoa da; Beraldi-Magalhaes, Francisco; Oliveira, Rosana Rosseto de; Andrade, Luciano de; Cardoso, Rosilene Fressatti. Early death by tuberculosis as the underlying cause in a state of Southern Brazil: Profile, comorbidities and associated vulnerabilities.. Int J Infect Dis, 2019; 80S(c3r, 9610933):S50-S57

Olle-Goig J.E.. Tuberculosis in rural Uganda. Afr. Health Sci., 2010; 10(3):226-229

Orofino, Renata de Lima; Brasil, Pedro Emmanuel Americano do; Trajman, Anete; Schmaltz, Carolina Arana Stanis; Dalcolmo, Margareth; Rolla, ValÃ©ria Cavalcanti. Preditores dos desfechos do tratamento da tuberculose. J. bras. pneumol, 2012; 38(1):88-97

PÃ©rez Navarro LM. CaracterizaciÃ³n y estimaciÃ³n de factores asociados a tuberculosis pulmonar en pacientes con y sin diabetes mellitus del Estado de Veracruz. , 2009; ():

Pablo MartÃn Fescina; Evangelina Membriani; Leticia Limongi; Ana Putruele. Incidencia de la resistencia a drogas en tuberculosis y su asociaciÃ³n a comorbilidades en pacientes tratados en un hospital universitario. Rev Am Med Resp, 2013; 2():64-70

Palma I.V.; Fescina P.; Aguirre R.; Rolando L.; Giovini V.; Kempf N.; Limongi L.; Putruele A.. Current antituberculosis drugs resistance. Am. J. Respir. Crit. Care Med., 2011; 183(1 MeetingAbstracts):

Palmero, Domingo; Ritacco, Viviana; Ambroggi, Marta; Poggi, Susana; Guemes Gurtubay, Jose; Alberti, Federico; Waisman, Jaime. [Multidrug-resistant tuberculosis in AIDS patients at the beginning of the millennium].. Medicina (B Aires), 2006; 66(5):399-404

Pang Y.; Jing W.; Chen W.; Guo R.; Han X.; Wu L.; Yang G.; Yang K.; Chen C.; Jiang L.; Cai C.; Dou Z.; Diao L.; Pan H.; Wang J.; Du F.; Xu T.; Wang L.; Li R.; Chu N.. Efficacy and safety of cycloserine-containing regimens in the treatment of multidrug-resistant tuberculosis: A nationwide retrospective cohort study in China. Infect. Drug Resist., 2019; 12((Wang, Jing, Chen, Guo, Han, Chu) Department of Tuberculosis, Beijing Chest Hospital, Capital Medical University Beijing Tuberculosis & Thoracic Tumor Research Institute, Beijing 101149, China):763-770

Parvaneh, Baghaei; Payam, Tabarsi; Zoha, Abrishami; Mehdi, Mirsaeidi; Yazdan Ali, Faghani; Seyed Davood, Mansouri; Mohammad Reza, Masjedi. Comparison of pulmonary TB patients with and without diabetes mellitus type II. Tanaffos, 2010; 9(2):13-20

Patil, Shital; Narwade, Swati; Mirza, Mazhar. Bronchial Wash Gene Xpert MTB/RIF in Lower Lung Field Tuberculosis: Sensitive, Superior, and Rapid in Comparison with Conventional Diagnostic Techniques.. J. transl. int. med., 2017; 5(3):174-181

Peltzer, Karl. Decline of common mental disorders over time in public primary care tuberculosis patients in South Africa.. Int J Psychiatry Med, 2016; 51(3):236-45

Peltzer, Karl. Correlates of tobacco use among tuberculosis patients in South Africa: A brief report.. Journal of Psychology in Africa, 2016; 26(5):473-476

Peltzer, Karl. Tuberculosis non-communicable disease comorbidity and multimorbidity in public primary care patients in South Africa.. Afr. j. prim. health care fam. med., 2018; 10(1):e1-e6

Peltzer, Karl; Louw, Julia. Prevalence of suicidal behaviour & associated factors among tuberculosis patients in public primary care in South Africa.. Indian J Med Res, 2013; 138(gjf, 0374701):194-200

Peltzer, Karl; Naidoo, Pamela; Matseke, Gladys; Louw, Julia; McHunu, Gugu; Tutshana, Bomkazi. Prevalence of psychological distress and associated factors in tuberculosis patients in public primary care clinics in South Africa.. BMC Psychiatry, 2012; 12(100968559):89

Pengpid, Supa; Peltzer, Karl. Mental morbidity and its associations with socio-behavioural factors and chronic conditions in rural middle- and older-aged adults in South Africa. JOURNAL OF PSYCHOLOGY IN AFRICA, 2020; 30(3):257-263

Penuelas-Urquides, Katia; Martinez-Rodriguez, Herminia Guadalupe; Enciso-Moreno, Jose Antonio; Molina-Salinas, Gloria Maria; Silva-Ramirez, Beatriz; Padilla-Rivas, Gerardo Raymundo; Vera-Cabrera, Lucio; Torres-de-la-Cruz, Victor Manuel; Martinez-Martinez, Yazmin Berenice; Ortega-Garcia, Jorge Luis; Garza-Trevino, Elsa Nancy; Enciso-Moreno, Leonor; Saucedo-Cardenas, Odila; Becerril-Montes, Pola; Said-Fernandez, Salvador. Correlations between major risk factors and closely related Mycobacterium tuberculosis isolates grouped by three current genotyping procedures: a population-based study in northeast Mexico.. Mem Inst Oswaldo Cruz, 2014; 109(6):814-9

Pepper, Dominique J; Rebe, Kevin; Morroni, Chelsea; Wilkinson, Robert J; Meintjes, Graeme. Clinical deterioration during antitubercular treatment at a district hospital in South Africa: the importance of drug resistance and AIDS defining illnesses.. PLoS ONE, 2009; 4(2):e4520

Perez-Navarro LM; Fuentes-Dominguez F; Morales-Romero J; Zenteno-Cuevas R. Factores asociados a tuberculosis pulmonar en pacientes con diabetes mellitus de Veracruz, Mâ€‚xico. Gaceta Mâ€‚dica de Mâ€‚xico., 2011; 147(3):219-225

Perez-Navarro, Lucia Monserrat; Fuentes-Dominguez, Francisco Javier; Zenteno-Cuevas, Roberto. Type 2 diabetes mellitus and its influence in the development of multidrug resistance tuberculosis in patients from southeastern Mexico.. J Diabetes Complications, 2015; 29(1):77-82

Perez-Navarro, Lucia Monserrat; Restrepo, Blanca I; Fuentes-Dominguez, Francisco Javier; Duggirala, Ravindranath; Morales-Romero, Jaime; Lopez-Alvarenga, Juan Carlos; Comas, Inaki; Zenteno-Cuevas, Roberto. The effect size of type 2 diabetes mellitus on tuberculosis drug resistance and adverse treatment outcomes.. Tuberculosis (Edinb), 2017; 103(d4x, 100971555):83-91

Picon, Pedro Dornelles; Bassanesi, Sergio Luiz; Caramori, Maria Luiza Avancini; Ferreira, Roberto Luiz Targa; Jarczewski, Carla Adriane; Vieira, PatrÃcia Rodrigues de Borba. Fatores de risco para a recidiva da tuberculose. J. bras. pneumol, 2007; 33(5):572-578

Pontino M.V.. Risk factors for defaults in tuberculosis treatment. Am. J. Respir. Crit. Care Med., 2010; 181(1 MeetingAbstracts):

Pontino, Monica V.; Brian, C; Ambrosino, N; Feldman, A; Iglesias, M; Doro, A; Garcia, M; Sancineto, A. Factores asociados al abandono del tratamiento antituberculoso. 8vo Congreso ALAT, 2012; ():

Porwal C.; Kaushik A.; Makkar N.; Banavaliker J.N.; Hanif M.; Singla R.; Bhatnagar A.K.; Behera D.; Pande J.N.; Singh U.B.. Incidence and Risk Factors for Extensively Drug-Resistant Tuberculosis in Delhi Region. PLoS ONE, 2013; 8(2):e55299

Poudyal, Indra Prasad; Khanal, Pratik; Mishra, Shiva Raj; Malla, Milan; Poudel, Prakash; Jha, Raj Kumar; Phuyal, Anil; Barakoti, Abiral; Adhikari, Bipin. Cardiometabolic risk factors among patients with tuberculosis attending tuberculosis treatment centers in Nepal.. BMC Public Health, 2020; 20(1):1364

Prado, Thiago Nascimento do; Caus, Antonio Luiz; Marques, Murilo; Maciel, Ethel Leonor; Golub, Jonathan E; Miranda, Angelica Espinosa. Epidemiological profile of adult patients with tuberculosis and AIDS in the state of Espirito Santo, Brazil: cross-referencing tuberculosis and AIDS databases.. J Bras Pneumol, 2011; 37(1):93-9

Prince, L; Andrews, J R; Basu, S; Goldhaber-Fiebert, J D. Risk of self-reported symptoms or diagnosis of active tuberculosis in relationship to low body mass index, diabetes and their co-occurrence.. Trop Med Int Health, 2016; 21(10):1272-1281

Qu, Junyan; Zhou, Taoyou; Zhong, Cejun; Deng, Rong; Lu, Xiaoju. Comparison of clinical features and prognostic factors in HIV-negative adults with cryptococcal meningitis and tuberculous meningitis: a retrospective study.. BMC Infect Dis, 2017; 17(1):51

Ranzani, Otavio T; Carvalho, Carlos R R; Waldman, Eliseu A; Rodrigues, Laura C. The impact of being homeless on the unsuccessful outcome of treatment of pulmonary TB in Sao Paulo State, Brazil.. BMC Med, 2016; 14(101190723):41

Ranzani, Otavio T; Rodrigues, Laura C; Bombarda, Sidney; Minto, CÃ¡tia M; Waldman, Eliseu A; Carvalho, Carlos R R. Long-term survival and cause-specific mortality of patients newly diagnosed with tuberculosis in SÃ£o Paulo state, Brazil, 2010Â15: a population-based, longitudinal study. The lancet, 2019; 2019():44470

Ranzani, Otavio T; Rodrigues, Laura C; Bombarda, Sidney; Minto, Catia M; Waldman, Eliseu A; Carvalho, Carlos R R. Long-term survival and cause-specific mortality of patients newly diagnosed with tuberculosis in Sao Paulo state, Brazil, 2010-15: a population-based, longitudinal study.. Lancet Infect Dis, 2020; 20(1):123-132

Rawat, Jagdish; Sindhwani, Girish; Juyal, Ruchi. Clinico-radiological profile of new smear positive pulmonary tuberculosis cases among young adult and elderly people in a tertiary care hospital at Deheradun (Uttarakhand).. INDIAN J. TUBERC., 2008; 55(2):84-90

Reechaipichitkul W.; So-Ngern A.; Chaimanee P.. Treatment outcomes of new and previously-treated smear positive pulmonary tuberculosis at Srinagarind Hospital, a tertiary care center in Northeast Thailand. J. Med. Assoc. Thailand, 2014; 97(5):490-499

Reechaipichitkul, Wipa. Multidrug-resistant tuberculosis at Srinagarind Hospital, Khon Kaen, Thailand.. Southeast Asian J Trop Med Public Health, 2002; 33(3):570-4

Requena-Mendez, Ana; Davies, Geraint; Ardrey, Alison; Jave, Oswaldo; Lopez-Romero, Sonia L; Ward, Stephen A; Moore, David A J. Pharmacokinetics of rifampin in Peruvian tuberculosis patients with and without comorbid diabetes or HIV.. Antimicrob Agents Chemother, 2012; 56(5):2357-63

Requena-Mendez, Ana; Davies, Geraint; Waterhouse, David; Ardrey, Alison; Jave, Oswaldo; Lopez-Romero, Sonia Llanet; Ward, Stephen A; Moore, David A J. Effects of dosage, comorbidities, and food on isoniazid pharmacokinetics in Peruvian tuberculosis patients.. Antimicrob Agents Chemother, 2014; 58(12):7164-70

Resende, MariÃ¢ngela Ribeiro; Sinkoc, VerÃ´nica Maria; Garcia, MÃ¡rcia Teixeira; Moraes, Eliane Oliveira de; Kritski, AfrÃ¢nio Lineu; Papaiordanou, Priscila Maria de Oliveira. Indicadores relacionados ao retardo no diagnÃ³stico e na instituiÃ§Ã£o das precauÃ§Ãµes para aerossÃ³is entre pacientes com tuberculose pulmonar bacilÃfera em um hospital terciÃ¡rio. J. bras. pneumol, 2005; 31(3):225-230

RESTREPO, B. I.; FISHER-HOCH, S. P.; CRESPO, J. G.; WHITNEY, E.; PEREZ, A.; SMITH, B.; McCORMICK, J. B.. Type 2 diabetes and tuberculosis in a dynamic bi-national border population. Epidemiology and Infection, 2007; 135(3):

Rocha, Marli Souza; Oliveira, Gisele Pinto de; Aguiar, Fernanda Pinheiro; Saraceni, ValÃ©ria; Pinheiro, Rejane Sobrino. Do que morrem os pacientes com tuberculose: causas mÃºltiplas de morte de uma coorte de casos notificados e uma proposta de investigaÃ§Ã£o de causas presumÃ-veis. Cad. saÃºde pÃºblica, 2015; 31(4):709-721

Roche, S; De Vries, E. Multimorbidity in a large district hospital: A descriptive cross-sectional study.. SAMJ, S. Afr. med. j., 2017; 107(12):1110-1115

Rubeen R.; Zareen N.; Zameer S.; Rasool A.G.; Naqvi S.S.N.; Iqbal J.. Anxiety and depression in tuberculosis can create impact on quality of life of patient. Acta Med. Int., 2014; 1(2):93-98

Sadiq, Shamiya; Khajuria, Vijay; Tandon, Vishal R; Mahajan, Annil; Singh, Jang B. Adverse Drug Reaction Profile in Patients on Anti-tubercular Treatment Alone and in Combination with Highly Active Antiretroviral Therapy.. J Clin Diagn Res, 2015; 9(10):FC01-4

Saha A.; Vaidya P.J.; Chavhan V.B.; Pandey K.V.; Kate A.H.; Leuppi J.D.; Tamm M.; Chhajed P.N.. Factors affecting outcomes of individualised treatment for drug resistant tuberculosis in an endemic region>. Eur. Respir. J., 2017; 50(Supplement 61):

Saifodine, Abuchahama; Gudo, Paula Samo; Sidat, Mohsin; Black, James. Patient and health system delay among patients with pulmonary tuberculosis in Beira city, Mozambique. BMC PUBLIC HEALTH, 2013; 13():

Saktiawati A.; Stienstra Y.; Subronto Y.W.; Sumardi S.; Gerritsen J.; Oord H.; Akkerman O.W.; Van Der Werf T.S.. Sensitivity and specificity of an electronic nose in diagnosing pulmonary tuberculosis among patients with suspected tuberculosis in Indonesia. Am. J. Respir. Crit. Care Med., 2019; 199(9):

Saktiawati, Antonia M I; Stienstra, Ymkje; Subronto, Yanri W; Rintiswati, Ning; Sumardi; Gerritsen, Jan-Willem; Oord, Henny; Akkerman, Onno W; van der Werf, Tjip S. Sensitivity and specificity of an electronic nose in diagnosing pulmonary tuberculosis among patients with suspected tuberculosis.. PLoS ONE, 2019; 14(6):e0217963

Saleh Jaber A.A.; Khan A.H.; Syed Sulaiman S.A.. Evaluation of tuberculosis defaulters in Yemen from the perspective of health care service. J. Pharm. Health. Serv. Res., 2018; 9(4):381-392

Santo, AH; Pinheiro, CE; Jordani, MS. Multiple-causes-of-death related to tuberculosis in the state of Sao Paulo, Brazil, 1998. REVISTA DE SAUDE PUBLICA, 2003; 37(6):714-721

Santo, Augusto Hasiak. Causas mÃºltiplas de morte relacionadas Ã  tuberculose no Estado do Rio de Janeiro entre 1999 e 2001. J. bras. pneumol, 2006; 32(6):544-552

Santos, Dinamara Barreto dos. Diabetes mellitus referido e fatores sociodemogrÃ¡ficos, clÃnicos e epidemiolÃ³gicos em pacientes adultos com tuberculose. , 2013; ():91-91

Sattar, S; Van Schalkwyk, C; Claassens, M; Dunbar, R; Floyd, S; Enarson, D A; Godfrey-Faussett, P; Ayles, H; Beyers, N. Symptom reporting among prevalent tuberculosis cases who smoke, are HIV-positive or have hyperglycaemia.. Public health action, 2014; 4(4):222-5

Sawadogo, Bernard; Tint, Khin San; Tshimanga, Mufuta; Kuonza, Lazarus; Ouedraogo, Laurent. Risk factors for tuberculosis treatment failure among pulmonary tuberculosis patients in four health regions of Burkina Faso, 2009: case control study.. Pan Afr Med J, 2015; 21(101517926):152

Seiscento, MÃ¡rcia; Vargas, Francisco Suso; Rujula, Maria Josefa Penon; Bombarda, Sidney; Uip, David Everson; Galesi, Vera Maria Nedes. Aspectos epidemiolÃ³gicos da tuberculose pleural no estado de SÃ£o Paulo (1998-2005). J. bras. pneumol, 2009; 35(6):548-554

Seiscento, Marcia; Vargas, Francisco Suso; Rujula, Maria Josefa Penon; Bombarda, Sidney; Uip, David Everson; Galesi, Vera Maria Nedes. Epidemiological aspects of pleural tuberculosis in the state of Sao Paulo, Brazil (1998-2005).. J Bras Pneumol, 2009; 35(6):548-54

Sengul, Aysun; Akturk, Ulku Aka; Aydemir, Yusuf; Kaya, Nurullah; Kocak, Nagihan Durmus; Tasolar, Fatma Turan. Factors affecting successful treatment outcomes in pulmonary tuberculosis: a single-center experience in Turkey, 2005-2011.. J. infect. dev. ctries., 2015; 9(8):821-8

Seni J.; Kidenya B.R.; Obassy E.; Mirambo M.; Burushi V.; Mazigo H.D.; Kapesa A.; Majigo M.; Mshana S.E.. Low sputum smear positive tuberculosis among pulmonary tuberculosis suspects in a tertiary hospital in Mwanza, Tanzania. Tanzan. J. Health Res., 2012; 14(2):44440

Seyed Mohammad, Alavi; Nejad, Salami. <The> causes of death among patients with tuberculosis in Khuzestan, Iran. Pak. J. Med. Sci., 2008; 24(2):217-220

Shah, Arti D; Akkara, Stani A; Adalja, Mayur; Akkara, Ajay G; Shah, Sejal. Association of pulmonary tuberculosis and dermatological conditions among patients of a rural medical college hospital.. Indian J Chest Dis Allied Sci, 2013; 55(4):201-4

Shamaei, Masoud; Samiei-Nejad, Mozhgan; Nadernejad, Masoumeh; Baghaei, Parvaneh. Risk factors for readmission to hospital in patients with tuberculosis in Tehran, Iran: three-year surveillance.. Int J STD AIDS, 2017; 28(12):1169-1174

Shanmuganathan, Aruna; R, Srinivasan; G, Thilagavathy; D, Satishkumar; C, Sidduraj; James, Bonny. Determination of Sites Involved, HIV Co-Infection & Utility of Diagnostic Modalities in EPTB.. J Clin Diagn Res, 2013; 7(8):1644-6

Shanmuganathan, Rohan; Subramaniam, Indra Devi. Clinical manifestation and risk factors of tuberculosis infection in Malaysia: case study of a community clinic.. Glob J Health Sci, 2015; 7(4):110-20

Sharma, A; Chhabra, H S; Chabra, T; Mahajan, R; Batra, S; Sangondimath, G. Demographics of tuberculosis of spine and factors affecting neurological improvement in patients suffering from tuberculosis of spine: a retrospective analysis of 312 cases.. Spinal Cord, 2017; 55(1):59-63

Sharma, M K; Kalia, Meenu; Walia, Dinesh; Goel, N K; Swami, H M. Surveillance of communicable diseases in tertiary health care system in Chandigarh, UT.. Indian J Med Sci, 2007; ():

Sharma, Parag; Lalwani, Jaya; Pandey, Pavan; Thakur, Avinash. Factors Associated with the Development of Secondary Multidrug-resistant Tuberculosis.. Int J Prev Med, 2019; 10(101535380):67

Shen, Xin; Xia, Zhen; Li, Xiangqun; Wu, Jie; Wang, Lili; Li, Jing; Jiang, Yuan; Guo, Juntao; Chen, Jing; Hong, Jianjun; Yuan, Zheng'an; Pan, Qichao; DeRiemer, Kathryn; Sun, Guomei; Gao, Qian; Mei, Jian. Tuberculosis in an urban area in China: differences between urban migrants and local residents.. PLoS ONE, 2012; 7(11):e51133

Shimazaki, T; Marte, S D; Saludar, N R D; Dimaano, E M; Salva, E P; Ariyoshi, K; Villarama, J B; Suzuki, M. Risk factors for death among hospitalised tuberculosis patients in poor urban areas in Manila, The Philippines.. Int J Tuberc Lung Dis, 2013; 17(11):1420-6

Sikalengo, George; Hella, Jerry; Mhimbira, Francis; Rutaihwa, Liliana K; Bani, Farida; Ndege, Robert; Sasamalo, Mohamed; Kamwela, Lujeko; Said, Khadija; Mhalu, Grace; Mlacha, Yeromin; Hatz, Christoph; Knopp, Stefanie; Gagneux, Sebastien; Reither, Klaus; Utzinger, Jurg; Tanner, Marcel; Letang, Emilio; Weisser, Maja; Fenner, Lukas. Distinct clinical characteristics and helminth co-infections in adult tuberculosis patients from urban compared to rural Tanzania.. Infect. dis. poverty, 2018; 7(1):24

Silva, Denise R; Menegotto, Diego M; Schulz, Luis F; Gazzana, Marcelo B; Dalcin, Paulo Tr. Mortality among patients with tuberculosis requiring intensive care: a retrospective cohort study.. BMC Infect Dis, 2010; 10(100968551):54

Smiljic S.; Stanisavljevic D.; Radovic B.; Mijovic M.; Savic S.; Ristic S.; Mandic P.. The sociodemographic characteristics and risk factors for tuberculosis morbidity between two decades at the beginning of the 21st century at the north of kosovo, Serbia. Vojnosanit. Pregl., 2018; 75(5):461-467

Soares, Luciana Nunes; Spagnolo, Lilian Moura de Lima; Tomberg, Jessica Oliveira; Zanatti, Christian Loret de Mola; Cardozo-Gonzales, Roxana Isabel. Relationship between multimorbidity and the outcome of the treatment for pulmonary tuberculosis.. Rev Gaucha Enferm, 2020; 41(8504882, rev):e20190373

Soares, Valeria Martins; Almeida, Isabela Neves de; Figueredo, Lida Jouca de Assis; Haddad, Joao Paulo Amaral; Oliveira, Camila Stefanie Fonseca de; Carvalho, Wania da Silva; Miranda, Silvana Spindola de. Factors associated with tuberculosis and multidrug-resistant tuberculosis in patients treated at a tertiary referral hospital in the state of Minas Gerais, Brazil.. J Bras Pneumol, 2020; 46(2):e20180386

Sogebi, Olusola Ayodele; Fadeyi, Muse Olatunbosun; Adefuye, Bolanle Olufunlola; Soyinka, Festus Olukayode. Hearing thresholds in patients with drug-resistant tuberculosis: baseline audiogram configurations and associations. J. bras. pneumol, 2017; 43(3):195-201

Sousa, Erivelton de Oliveira. CaracterizaÃ§Ã£o dos perfis genÃ©ticos e de resistÃªncia a fÃ¡rmacos de isolados de mycobacterium tuberculosis associados com casos de tuberculose multirresistente na Bahia, Brasil. , 2012; ():94-94

Srikantiah, P; Lin, R; Walusimbi, M; Okwera, A; Luzze, H; Whalen, C C; Boom, W H; Havlir, D V; Charlebois, E D. Elevated HIV seroprevalence and risk behavior among Ugandan TB suspects: implications for HIV testing and prevention.. Int J Tuberc Lung Dis, 2007; 11(2):168-74

Stoffel, Carina; Lorenz, Rosana; Arce, Marta; Rico, Marina; FernÃ¡ndez, Liliana; Imaz, MarÃa S.. Tratamiento de la tuberculosis pulmonar en un Ã¡rea urbana de baja prevalencia: Cumplimiento y negativizaciÃ³n bacteriolÃ³gica. Medicina (B.Aires), 2014; 74(1):43344

Stoffel, Carina; Lorenz, Rosana; Arce, Marta; Rico, Marina; Fernandez, Liliana; Imaz, Maria S. [Treatment of pulmonary tuberculosis in a low-prevalence urban area. Compliance and sputum conversion].. Medicina (B Aires), 2014; 74(1):43344

Sunnetcioglu, Mahmut; Baran, Ali Irfan; Binici, Irfan; Esmer, Fatih; Gultepe, Bilge. Evaluation of 257 extra pulmonary tuberculosis cases at the Tuberculosis Control Dispensary, Van, Turkey.. JPMA J Pak Med Assoc, 2018; 68(5):764-767

Tabassum, Masood Nizam; Khan, Muhammad Ather; Afzal, Saira; Gilani, Syed Amir; Gureja, Abdul Wahab; Tabassum, Shafaq. Determination of Risk Factors among Multi-Drug Resistant Tuberculosis Patients. ANNALS OF KING EDWARD MEDICAL UNIVERSITY LAHORE PAKISTAN, 2018; 24(2):787-796

Tag El Din M.A.; El Maraghy A.A.; Abdel Hay A.H.R.. Adverse reactions among patients being treated for multi-drug resistant tuberculosis at Abbassia Chest Hospital. Egypt. J. Chest Dis. Tuberc., 2015; 64(4):939-952

Tahir, Zarfishan; Ahmad, Mansur-Ud-Din; Akhtar, Abdul Majeed; Yaqub, Tahir; Mushtaq, Muhammad Hassan; Javed, Hasnain. Diabetes mellitus among tuberculosis patients: a cross sectional study from Pakistan.. Afr Health Sci, 2016; 16(3):671-676

Tan T.L.; Lee L.Y.; Yong K.T.; Rohimi M.A.B.; Chiew S.C.; Cheng S.H.; Haniba H.B.M.; Ding M.T.. Pre-existing chronic medical illnesses and follow up status among active pulmonary tuberculosis cases in a district population. Med. J. Malays., 2020; 75(3):204-208

Tang S.; Tan S.; Yao L.; Li F.; Li L.; Guo Z.; Liu Y.; Hao X.; Li Y.; Ding X.; Zhang Z.; Tong L.; Huang J.. Risk factors for poor treatment outcomes in patients with MDR-TB and XDR-TB in China: Retrospective multicenter investigation. Chest, 2014; 145(3 MEETING ABSTRACT):

Taskin F.; Olgun N.. Quality of life in patients with pulmonary tuberculosis. Turk Toraks Derg., 2010; 11(1):19-25

Tessema T.A.; Bjune G.; Assefa G.; Svenson S.; Hamasur B.; Bjorvatn B.. Clinical and radiological features in relation to urinary excretion of lipoarabinomannan in Ethiopian tuberculosis patients. Scand. J. Infect. Dis., 2002; 34(3):167-171

Thai, L T; Li, Y L; Kig, T Y; Muhammad Afiq, R; Shoen, C C; Sing, H C; Hafizah, M H; Min, T D. Pre-existing chronic medical illnesses and follow up status among active pulmonary tuberculosis cases in a district population.. Med J Malaysia, 2020; 75(3):204-208

Tomita, Andrew; Ramlall, Suvira; Naidu, Thirusha; Mthembu, Sbusisiwe Sandra; Padayatchi, Nesri; Burns, Jonathan K. Neurocognitive Impairment Risk Among Individuals With Multiple Drug-Resistant Tuberculosis and Human Immunodeficiency Virus Coinfection: Implications for Systematic Linkage to and Retention of Care in Tuberculosis/Human Immunodeficiency Virus Treatment.. J Nerv Ment Dis, 2019; 207(4):307-310

Torres-Garcia, Diana; Cruz-Lagunas, Alfredo; Garcia-Sancho Figueroa, Ma Cecilia; Fernandez-Plata, Rosario; Baez-Saldana, Renata; Mendoza-Milla, Criselda; Barquera, Rodrigo; Carrera-Eusebio, Aida; Ramirez-Bravo, Salomon; Campos, Lizeth; Angeles, Javier; Vargas-Alarcon, Gilberto; Granados, Julio; Gopal, Radha; Khader, Shabaana A; Yunis, Edmond J; Zuniga, Joaquin. Variants in toll-like receptor 9 gene influence susceptibility to tuberculosis in a Mexican population.. J. transl. med., 2013; 11(101190741):220

Trinnawoottipong K.; Suggaravetsiri P.; Tesana N.; Chaiklieng S.. Factors associated with multidrug-resistant tuberculosis patients in the Upper Northeast Thailand. Res. J. Med. Sci., 2012; 6(4):208-213

Ugoeze, F. C.; Ele, P. U.; Anyabolu, A. E.; Enemuo, E. H.; Okonkwo, R. C.; Umeh, E. O.; Ezeani, I. U.; Onyeonoro, U. U.. Pulmonary tuberculosis and diabetes mellitus co-morbidity in a nigerian tertiary hospital. AFRICAN JOURNAL OF RESPIRATORY MEDICINE, 2019; 14(2):13-17

Vallejos-Paras, Alfonso; Cabrera-Gaytan, David Alejandro; Padilla-Velazquez, Rosario; Arriaga-Nieto, Lumumba; Valle-Alvarado, Gabriel; Grajales-Muniz, Concepcion; Rojas-Mendoza, Teresita; Niebla-Fuentes, Maria Del Rosario. [Association of Notified Cases and Treatment Success Rates in Pulmonary Tuberculosis].. Rev Invest Clin, 2018; 70(4):198-202

Van der Plas, Helen; Mendelson, Marc. High prevalence of comorbidity and need for up-referral among inpatients at a district-level hospital with specialist tuberculosis services in South Africa: the need for specialist support.. SAMJ, S. Afr. med. j., 2011; 101(8):529-32

VanSteelandt, Amanda. Cultural transmission and the disease ecology of tuberculosis in indigenous communities of the Paraguayan Chaco.. Dissertation Abstracts International: Section B: The Sciences and Engineering, 2015; 76(Health & Mental Health Treatment & Prevention [3300]):No-Specified

Vieira, Rafael da Cruz AraÃºjo; Fregona, Geisa; Palaci, MoisÃ©s; Dietze, Reynaldo; Maciel, Ethel Leonor Noia; Sassi, BÃ¡rbara Tomasi. Perfil epidemiolÃ³gico dos casos de tuberculose multirresistente do EspÃrito Santo. Rev. bras. epidemiol, 2007; 10(1):56-65

Villa, Liliana; Trompa, Ivan Mauricio; Montes, Fernando Nicolas; Gomez, Joaquin Guillermo; Restrepo, Carlos Andres. [Analysis of mortality caused by tuberculosis in Medellin, Colombia, 2012].. Biomedica (Bogota), 2014; 34(3):425-32

Waghmare, Manoj Ashok; Utpat, Ketaki; Joshi, Jyotsna M.. Treatment outcomes of drug-resistant pulmonary tuberculosis under programmatic management of multidrug-resistant tuberculosis, at tertiary care center in Mumbai. Medical Journal of Dr. D.Y. Patil University, 2017; 10(1):41

Wang Y.; Chen H.; Huang Z.; McNeil E.B.; Lu X.; Chongsuvivatwong V.. Drug non-adherence and reasons among multidrug-resistant tuberculosis patients in Guizhou, China: A cross-sectional study. Patient Preference Adherence, 2019; 13((Wang, Lu) School of Medicine and Health Management, Guizhou Medical University, Guiyang, Guizhou, China):1641-1653

Wang, J-Y; Hsueh, P-R; Jan, I-S; Lee, L-N; Liaw, Y-S; Yang, P-C; Luh, K-T. Empirical treatment with a fluoroquinolone delays the treatment for tuberculosis and is associated with a poor prognosis in endemic areas.. Thorax, 2006; 61(10):903-8

Wang, Qiuzhen; Ma, Aiguo; Han, Xiuxia; Zhao, Shanliang; Cai, Jing; Ma, Yunbo; Zhao, Jie; Wang, Yuwen; Dong, Huaifeng; Zhao, Zhenlei; Wei, Lai; Yu, Tao; Chen, Peixue; Schouten, Evert G; Kok,

Frans J; Kapur, Anil. Prevalence of type 2 diabetes among newly detected pulmonary tuberculosis patients in China: a community based cohort study.. PLoS ONE, 2013; 8(12):e82660

Wang, W B; Zhao, Q; Yuan, Z A; Jiang, W L; Liu, M L; Xu, B. Deaths of tuberculosis patients in urban China: a retrospective cohort study.. Int J Tuberc Lung Dis, 2013; 17(4):493-8

Wijayanto M.A.; Arnanda R.A.; Thamrin E.P.. Risk factors for development of multidrug-resistant tuberculosis among relapsed patients in west Papua, Indonesia: A descriptive and analytical study. Int. J. Appl. Pharm., 2019; 11(Special Issue 6):50-55

William, Timothy; Parameswaran, Uma; Lee, Wai Khew; Yeo, Tsin Wen; Anstey, Nicholas M; Ralph, Anna P. Pulmonary tuberculosis in outpatients in Sabah, Malaysia: advanced disease but low incidence of HIV co-infection.. BMC Infect Dis, 2015; 15(100968551):32

Yang T.; Chen T.; Che Y.; Chen Q.; Bo D.. Factors associated with catastrophic total costs due to tuberculosis under a designated hospital service model: a cross-sectional study in China. BMC Public Health, 2020; 20(1):1009

Yasar, K K; Pehlivanoglu, F; Sengoz, G. Predictors of mortality in tuberculous meningitis: a multivariate analysis of 160 cases.. Int J Tuberc Lung Dis, 2010; 14(10):1330-5

Yoon H.J.; Song Y.G.; Park W.I.; Choi J.P.; Chang K.H.; Kim J.-M.. Clinical manifestations and diagnosis of extrapulmonary tubercolosis. Yonsei Med. J., 2004; 45(3):453-461

Yorke, Ernest; Boima, Vincent; Dey, Ida Dzifa; Ganu, Vincent; Nkornu, Norah; Acquaye, Kelvin Samuel; Mate-Kole, C. Charles. Comparison of neurocognitive changes among newly diagnosed tuberculosis patients with and without dysglycaemia.. BMC Psychiatry, 2020; 20(Alberti, G., Zimmet, P., Shaw, J., and Grundy, S.M. (2006). The IDF consensus worldwide definition of the metabolic syndrome. Brussels: Int Diabetes Federation, 23(5), 469-480.):

Young, Bonnie N; Burgos, Marcos; Handal, Alexis J; Baker, Jack; Rendon, Adrian; Rosas-Taraco, Adrian; Long, Jeffrey; Hunley, Keith. Social and clinical predictors of drug-resistant tuberculosis in a public hospital, Monterrey, Mexico.. Ann Epidemiol, 2014; 24(10):771-5

Yu Y.-H.; Liao C.-C.; Hsu W.-H.; Chen H.-J.; Liao W.-C.; Muo C.-H.; Sung F.-C.; Chen C.-Y.. Increased lung cancer risk among patients with pulmonary tuberculosis: A population cohort study. J. Thorac. Oncol., 2011; 6(1):32-37

Yuan, Xiaoliang; Zhang, Tiantuo; Kawakami, Kazuyoshi; Zhu, Jiaxin; Zheng, Wenzheng; Li, Hongtao; Deng, Guofang; Tu, Shaohua; Liu, Weiyou. Genotyping and clinical characteristics of multidrug and extensively drug-resistant tuberculosis in a tertiary care tuberculosis hospital in China.. BMC Infect Dis, 2013; 13(100968551):315

Zaikov, S.; Bogomolov, A.; Grishilo, A.. Bronchial asthma and pulmonary tuberculosis as comorbid diseases. ALLERGY, 2019; 74(106, SI):432-433

Zangana, Abdulqadir Maghded; Al-Hadithi, Tariq S.; Ismail, Sherzad Ali. Prevalence of tuberculous peritonitis in the North of Iraq and sociodemographic comparison with pulmonary tuberculosis. ASIAN PACIFIC JOURNAL OF TROPICAL MEDICINE, 2009; 2(2):58-63

Zhan, X; Wang, Z; Zhang, L; Jin, M-L; Liu, M; Chen, W-H; Dai, H-P. Clinical and pathological features of adult pulmonary tuberculosis with reversed halo sign.. Int J Tuberc Lung Dis, 2013; 17(12):1621-5

Zhao, Q; Xiao, X; Lu, W; Qiu, L-X; Zhou, C-M; Jiang, W-L; Xu, B; Diwan, V. Screening diabetes in tuberculosis patients in eastern rural China: a community-based cross-sectional study.. Int J Tuberc Lung Dis, 2016; 20(10):1370-1376

**- No outcomes of interest**

Ade, S.; Affolabi, D.; Agodokpessi, G.; Wachinou, P.; FaÃ¯hun, F.; Toundoh, N.; BÃ©kou, W.; Makpenon, A.; Ade, G.; Anagonou, S.; Harries, A. D.. Low prevalence of diabetes mellitus in patients with tuberculosis in Cotonou, Benin. Public Health Action, 2015; 5(2):

Adewole, Olufemi O; Erhabor, Greg E; Ogunrombi, Akinwumi B; Awopeju, Fehintola A. Prevalence and patient characteristics associated with pleural tuberculosis in Nigeria.. J. infect. dev. ctries., 2010; 4(4):213-7

Aggarwal D.; Mohapatra P.R.. Sputum conversion at the end of intensive phase treatment of pulmonary tuberculosis patients with diabetes mellitus or HIV infection. Indian J. Med. Res., 2008; 127(4):408

Al-Jahdali, H; Al-Zahrani, K; Amene, P; Memish, Z; Al-Shimemeri, A; Moamary, M; Alduhaim, A. Clinical aspects of miliary tuberculosis in Saudi adults.. Int J Tuberc Lung Dis, 2000; 4(3):252-5

Ambaw, Fentie; Mayston, Rosie; Hanlon, Charlotte; Alem, Atalay. Incidence of depression in people with newly diagnosed tuberculosis in Ethiopia: a cohort study.. Glob Ment Health (Camb), 2020; 7(101659641):e1

Aweis, Daud M I; Suleiman, Syed Azhar Syed. Economic Burden of Diabetic Tuberculosis Patients.. Br J Med Med Res, 2012; ():

Balde, N M; Camara, A; Camara, L M; Diallo, M M; Kake, A; Bah-Sow, O Y. Associated tuberculosis and diabetes in Conakry, Guinea: prevalence and clinical characteristics.. Int J Tuberc Lung Dis, 2006; 10(9):1036-40

Bao X.-Y.; Xie Y.-X.; Zhang X.-X.; Peng X.; Huang J.-X.; Du Q.-F.; Wang P.-X.. The association between multimorbidity and health-related quality of life: A cross-sectional survey among community middle-aged and elderly residents in southern China. Health Qual. Life Outcomes, 2019; 17(1):107

Dawra S.; Mandavdhare H.S.; Singh H.; Prasad K.K.; Dutta U.; Sharma V.. Extra-abdominal involvement is associated with antitubercular therapy-related hepatitis in patients treated for abdominal tuberculosis. Clin. Exp. Hepatol., 2019; 5(1):60-64

Gugnani, H C; Denning, D W; Rahim, R; Sadat, A; Belal, M; Mahbub, M S. Burden of serious fungal infections in Bangladesh.. Eur J Clin Microbiol Infect Dis, 2017; 36(6):993-997

H.H, Sara; M.A.B, Chowdhury; M.A, Haque. Multimorbidity among elderly in Bangladesh. Aging Medicine, 2018; 1(3):267-275

Hu, Hongyan; Chen, Jiaying; Sato, Kaori D; Zhou, Yang; Jiang, Hui; Wu, Pingbo; Wang, Hong. Factors that associated with TB patient admission rate and TB inpatient service cost: a cross-sectional study in China.. Infect. dis. poverty, 2016; 5(101606645):4

J.S, Zifodya; S.M, LaCourse; T, Temu; E, Attia; S, Masyuko; G, Nyale; J, Nyabiage; D, Onyango; J, Kinuthia; S, Page; C, Farquhar; K.A, Crothers. Latent TB infection and quantiferon-TB gold plus in HIV-infected and uninfected Kenyan adults. American Journal of Respiratory and Critical Care Medicine, 2020; 201(1):

Kubjane, Mmamapudi; Berkowitz, Natacha; Goliath, Rene; Levitt, Naomi S; Wilkinson, Robert J; Oni, Tolu. Tuberculosis, Human Immunodeficiency Virus, and the Association With Transient Hyperglycemia in Periurban South Africa.. Clin Infect Dis, 2020; 71(4):1080-1088

Lin, Fei-Shen; Wu, Mei-Ying; Tu, Wen-Jun; Pan, Hong-Qiu; Zheng, Jian; Shi, Jun-Wei; Fei, Zhong-Ting; Zhang, Rui-Mei; Yan, Wei-Guo; Shang, Ming-Qun; Zheng, Qiang; Wang, Meng-Jie; Zhang, Xia. A cross-sectional and follow-up study of leukopenia in tuberculosis patients: prevalence, risk factors and impact of anti-tuberculosis treatment.. J. thorac. dis., 2015; 7(12):2234-42

MACGEE, W. CAUSES OF DEATH IN A HOSPITALIZED GERIATRIC POPULATION - AN AUTOPSY STUDY OF 3000 PATIENTS. VIRCHOWS ARCHIV A-PATHOLOGICAL ANATOMY AND HISTOPATHOLOGY, 1993; 423(5):343-349

Mahato R.K.; Laohasiriwong W.; Koju R.. The role of type 2 diabetes mellitus on the clinical manifestation of pulmonary tuberculosis: A study from Nepal. J. Clin. Diagn. Res., 2019; 13(9):LC09-LC14

Marzuki O.A.; Fauzi A.R.M.; Ayoub S.; Kamarul Imran M.. Prevalence and risk factors of anti-tuberculosis drug-induced hepatitis in Malaysia. Singapore Med. J., 2008; 49(9):688-693

Miranda, Angelica Espinosa; Golub, Jonathan E; Lucena, Francisca de Fatima; Maciel, Ethel Noia; Gurgel, Maria de Fatima; Dietze, Reynaldo. Tuberculosis and AIDS co-morbidity in Brazil: linkage of the tuberculosis and AIDS databases.. Braz J Infect Dis, 2009; 13(2):137-41

Moussas, Georgios; Tselebis, Athanasios; Karkanias, Athanasios; Stamouli, Dimitra; Ilias, Ioannis; Bratis, Dionisios; Vassila-Demi, Kalliopi. A comparative study of anxiety and depression in patients with bronchial asthma, chronic obstructive pulmonary disease and tuberculosis in a general hospital of chest diseases.. Ann Gen Psychiatry, 2008; 7(101236515):7

Rashak, H A; Sanchez-Perez, H J; Abdelbary, B E; Bencomo-Alerm, A; Enriquez-Rios, N; Gomez-Velasco, A; Colorado, A; Castellanos-Joya, M; Rahbar, M H; Restrepo, B I. Diabetes, undernutrition, migration and indigenous communities: tuberculosis in Chiapas, Mexico.. Epidemiol Infect, 2019; 147(epi, 8703737):e71

Russom, Mulugeta; Tesfaselassie, Hager; Goitom, Rozina; Ghirmai, Tadese; Weldedhawariat, Freweini; Berhe, Abiel; Tesfai, Dawit; Debesai, Merhawi; Berhane, Tesfit; Woldu, Henok G. Risk Factors of Gout in MDR-TB Patients in Eritrea: A Case-Control Study.. tuberc. res. treat. (print), 2019; 2019(101576351):9429213

Sarker, Malabika; Barua, Mrittika; Guerra, Fiona; Saha, Avijit; Aftab, Afzal; Latif, A H M Mahbub; Islam, Shayla; Islam, Akramul. Double Trouble: Prevalence and Factors Associated with Tuberculosis and Diabetes Comorbidity in Bangladesh.. PLoS ONE, 2016; 11(10):e0165396

Segafredo, Giulia; Kapur, Anil; Robbiati, Claudia; Joseph, Nsuka; de Sousa, Joseth Rita; Putoto, Giovanni; Manenti, Fabio; Atzori, Andrea; Fedeli, Ugo. Integrating TB and non-communicable diseases services: Pilot experience of screening for diabetes and hypertension in patients with Tuberculosis in Luanda, Angola.. PLoS ONE, 2019; 14(7):e0218052

Sharma, Vishal; Bhagat, Sanjeev; Verma, Bhimsain; Singh, Ravinder; Singh, Surinderpal. Audiological Evaluation of Patients Taking Kanamycin for Multidrug Resistant Tuberculosis.. Iran. j. otorhinolyngol., 2016; 28(86):203-8

Souza, Debora Christina Santos; Oliveira, Keurilene Sutil de; Andrade, Rubia Laine de Paula; Scatena, Lucia Marina; Silva-Sobrinho, Reinaldo Antonio. Aspects related to the outcomes of the treatment, in international borders, of cases of tuberculosis as associated to comorbidities.. Rev Gaucha Enferm, 2019; 40(8504882, rev):e20190050

Sutar R.. Psycho-pulmonology: An observational study to screen psychiatric comorbidities in respiratory disorders. Indian J. Psychiatry, 2019; 61(9 Supplement 3):S463-S464

Tabassum, Masood Nizam; Gureja, Abdul Wahab; Tabassum, Shafaq; Sheikh, Muhammad Usman; Noor, Sana; Ikram, Nasir. Determination of risk factors among Tuberculosis patients at public sector Hospital Lahore. PAKISTAN JOURNAL OF MEDICAL & HEALTH SCIENCES, 2019; 13(4):792-795

Tatar, Dursun; Senol, Gunes; Kirakli, Cenk; Edipoglu, Ozlem; Cimen, Pinar. Contributing factors to mortality rates of pulmonary tuberculosis in intensive care units.. J Chin Med Assoc, 2018; 81(7):605-610

Toure, N O; Dia Kane, Y; Diatta, A; Ba Diop, S; Niang, A; Ndiaye, E M; Thiam, K; Mbaye, F B R; Badiane, M; Hane, A A. [Tuberculosis in elderly persons].. Rev Mal Respir, 2010; 27(9):1062-8

Tripathi SB; Kapadia VK. Treatment Outcome of Tuberculosis in HIV Seropositive Patients: An Experience of Southeast Region of Ahmedabad .. Ntl J of Community Med, 2015; 6(4):462-465

Umanah, Teye; Ncayiyana, Jabulani; Padanilam, Xavier; Nyasulu, Peter S. Treatment outcomes in multidrug resistant tuberculosis-human immunodeficiency virus Co-infected patients on anti-retroviral therapy at Sizwe Tropical Disease Hospital Johannesburg, South Africa.. BMC Infect Dis, 2015; 15(100968551):478

Valenzuela-Jimenez, Hiram; Manrique-Hernandez, Edgar Fabian; Idrovo, Alvaro Javier. Association of tuberculosis with multimorbidity and social networks.. J Bras Pneumol, 2017; 43(1):51-53 van der Plas, Helen; Meintjes, Graeme; Schutz, Charlotte; Goliath, Rene; Myer, Landon; Baatjie, Dorothea; Wilkinson, Robert J; Maartens, Gary; Mendelson, Marc. Complications of antiretroviral therapy initiation in hospitalised patients with HIV-associated tuberculosis.. PLoS ONE, 2013; 8(2):e54145

Vasudevan, KavitaP; Govindarajan, S; Chinnakali, Palanivel; Panigrahi, KrishnaChandra; Raghuraman, Soundararajan. Prevalence of diabetes mellitus among tuberculosis patients in Urban Puducherry. North American Journal of Medical Sciences, 2014; 6(1):

Watanabe, Arthur; Ruffino-Netto, Antonio. Aspectos epidemiolÃ³gicos da co-infecÃ§Ã¤o tuberculose-HIV - RibeirÃ¤o Preto/SP. Medicina (RibeirÃ¤o Preto), 1995; 28(4):856-65

Wu H.; Xu J.; Wen Y.; Dan X.; Liu M.; Ma J.; Wu Z.; Dong H.. Association between cystatin c and the interaction of pulmonary tuberculosis with chronic diseases. Trop. J. Pharm. Res., 2017; 16(8):2007-2012

Yu, Elaine A; Finkelstein, Julia L; Brannon, Patsy M; Bonam, Wesley; Russell, David G; Glesby, Marshall J; Mehta, Saurabh. Nutritional assessment among adult patients with suspected or confirmed active tuberculosis disease in rural India.. PLoS ONE, 2020; 15(5):e0233306

Zhovanyk, Natalia V; Tovt-Korshynska, Mariana I. Interaction between clinical and psychological changes among patients with chronic obstructive pulmonary disease and pulmonary tuberculosis co-morbidity.. Wiad Lek, 2019; 72(4):635-638

**- Wrong population**

Abate, Ebba; Belayneh, Meseret; Gelaw, Aschalew; Idh, Jonna; Getachew, Assefa; Alemu, Shitaye; Diro, Ermias; Fikre, Nigussu; Britton, Sven; Elias, Daniel; Aseffa, Abraham; Stendahl, Olle; Schon, Thomas. The impact of asymptomatic helminth co-infection in patients with newly diagnosed tuberculosis in north-west Ethiopia.. PLoS ONE, 2012; 7(8):e42901

Bates, Matthew; Mudenda, Victor; Shibemba, Aaron; Kaluwaji, Jonas; Tembo, John; Kabwe, Mwila; Chimoga, Charles; Chilukutu, Lophina; Chilufya, Moses; Kapata, Nathan; Hoelscher, Michael; Maeurer, Markus; Mwaba, Peter; Zumla, Alimuddin. Burden of tuberculosis at post mortem in inpatients at a tertiary referral centre in sub-Saharan Africa: a prospective descriptive autopsy study.. Lancet Infect Dis, 2015; 15(5):544-51

Bigna, Jean Joel R; Noubiap, Jean Jacques N; Agbor, Ako A; Plottel, Claudia S; Billong, Serge Clotaire; Ayong, Andre Patrick R; Koulla-Shiro, Sinata. Early Mortality during Initial Treatment of Tuberculosis in Patients Co-Infected with HIV at the Yaounde Central Hospital, Cameroon: An 8-Year Retrospective Cohort Study (2006-2013).. PLoS ONE, 2015; 10(7):e0132394

Chebrolu P.; Laux T.; Chowdhury S.; Seth B.; Ranade P.; Goswami J.; Chatterjee S.. The risk of refeeding syndrome among severely malnourished tuberculosis patients in Chhattisgarh, India. Indian J. Tuberc., 2020; 67(2):152-158

do Prado, Thiago Nascimento; Galavote, Heleticia Scabelo; Brioshi, Ana Paula; Lacerda, Thamy; Fregona, Geisa; Detoni, Valderio do Valle; Lima, Rita de Cassia Duarte; Dietze, Reynaldo; Maciel, Ethel Leonor Noia. Epidemiological profile of tuberculosis cases reported among health care workers at the University Hospital in Vitoria, Brazil.. J Bras Pneumol, 2008; 34(8):607-13

Erawati M.; Andriany M.. The prevalence and demographic risk factors for latent tuberculosis infection (LTBI) among healthcare workers in Semarang, Indonesia. J. Multidiscip.Healthc., 2020; 13((Erawati, Andriany) Department of Nursing, Faculty of Medicine, Universitas Diponegoro, Semarang, Indonesia):197-206

Faurholt-Jepsen D.; Range N.; Praygod G.; Jeremiah K.; Faurholt-Jepsen M.; Aabye M.G.; Changalucha J.; Christensen D.L.; Grewal H.M.S.; Martinussen T.; Krarup H.; Witte D.R.; Andersen A.B.; Friis H.. Diabetes is a strong predictor of mortality during tuberculosis treatment: A prospective cohort study among tuberculosis patients from Mwanza, Tanzania. Trop. Med. Int. Health, 2013; 18(7):822-829

John, G T; Shankar, V; Abraham, A M; Mukundan, U; Thomas, P P; Jacob, C K. Risk factors for post-transplant tuberculosis.. Kidney Int, 2001; 60(3):1148-53

K, Shah; M, Kothari; A, Nene. Role of Frailty Scoring in the Assessment of Perioperative Mortality in Surgical Management of Tuberculous Spondylodiscitis in the Elderly. Global Spine Journal, 2018; 8(7):698-702

Karthika, M; Philip, Sairu; Prathibha, M T; Varghese, Anu; Rakesh, P S. Why are people dying due to tuberculosis? A study from Alappuzha District, Kerala, India.. INDIAN J. TUBERC., 2019; 66(4):443-447

Kerketta, A S; Bulliyya, G; Babu, B V; Mohapatra, S S S; Nayak, R N. Health status of the elderly population among four primitive tribes of Orissa, India: a clinico-epidemiological study.. Z Gerontol Geriatr, 2009; 42(1):53-9

Mantilla-Hernandez J.C.; Diaz-Perez J.A.; Herrera L.P.; Melo-Uribe M.A.. Autopsy findings in patients with AIDS/tuberculosis coinfection. Lab. Invest., 2009; 89(Suppl. 1):7A

Menon, K Venugopal; Sorour, Tamer Malak Moawad. Epidemiologic and Demographic Attributes of Primary Spondylodiscitis in a Middle Eastern Population Sample.. World Neurosurg, 2016; 95(101528275):31-39

Motta, I; Centis, R; D'Ambrosio, L; Garcia-Garcia, J-M; Goletti, D; Gualano, G; Lipani, F; Palmieri, F; Sanchez-Montalva, A; Pontali, E; Sotgiu, G; Spanevello, A; Stochino, C; Tabernero, E; Tadolini, M; van den Boom, M; Villa, S; Visca, D; Migliori, G B. Tuberculosis, COVID-19 and migrants: Preliminary analysis of deaths occurring in 69 patients from two cohorts.. Pulmonology, 2020; 26(4):233-240

Prado, Thiago Nascimento do; Galavote, Heleticia Scabelo; Brioshi, Ana Paula; Lacerda, Thamy; Fregona, Geisa; Detoni, ValdÃ©rio do Valle; Lima, Rita de CÃ¡ssia Duarte; Dietze, Reynaldo; Maciel, Ethel Leonor Noia. Perfil epidemiolÃ³gico dos casos notificados de tuberculose entre os profissionais de saÃºde no Hospital UniversitÃ¡rio em VitÃ³ria (ES) Brasil. J. bras. pneumol, 2008; 34(8):607-613

Ramchandra, Tagaram; Jha, Punam Kumari; Bhavani, K.; Kumar, N. Pragathi. FACTORS RELATED TO DEFAULTERS OF TUBERCULOSIS PATIENTS IN DOTS PROGRAM IN WARANGAL DISTRICT. JOURNAL OF EVOLUTION OF MEDICAL AND DENTAL SCIENCES-JEMDS, 2015; 4(101):16617-16621

Rogerio, Wesley Pereira; Prado, Thiago Nascimento do; Souza, Fernanda Mattos de; Pinheiro, Jair dos Santos; Rodrigues, PatrÃcia Marques; Sant’anna, Amanda Pissinate do Nascimento; Jesus, KÃ¡ssia Gomes de; Cerutti Junior, Crispim; Lima, Rita de CÃ¡ssia Duarte; Maciel, Ethel Leonor Noia. PrevalÃªncia e fatores associados Ã  infecÃ§Ã£o pelo Mycobacterium tuberculosis entre agentes comunitÃ¡rios de saÃºde no Brasil, usando-se a prova tuberculÃnica. Cad. saÃºde pÃºblica, 2015; 31(10):2199-2210

Saiman, L; Aronson, J; Zhou, J; Gomez-Duarte, C; Gabriel, P S; Alonso, M; Maloney, S; Schulte, J. Prevalence of infectious diseases among internationally adopted children.. Pediatrics, 2001; 108(3):608-12

Seedat, Faheem; Martinson, Neil; Motlhaoleng, Katlego; Abraham, Pattamukkil; Mancama, Dalu; Naicker, Saraladevi; Variava, Ebrahim. Acute Kidney Injury, Risk Factors, and Prognosis in Hospitalized HIV-Infected Adults in South Africa, Compared by Tenofovir Exposure.. AIDS Res Hum Retroviruses, 2017; 33(1):33-40

Shidam, U. G.; Roy, G.; Sahu, S. K.; Kumar, S. V.; Ananthanarayanan, P. H.. Screening for diabetes among presumptive tuberculosis patients at a tertiary care centre in Pondicherry, India. The International Journal of Tuberculosis and Lung Disease, 2015; 19(10):

Teixeira, E G; Menzies, D; Comstock, G W; Cunha, A J L A; Kritski, A L; Soares, L C; Bethlem, E; Zanetti, G; Ruffino-Netto, A; Belo, M T C T; Selig, L; Branco, M M Castello; Cherri, D; Maia, S; Marandino, R; Luiz, R R; Chaisson, R E; Trajman, A. Latent tuberculosis infection among undergraduate medical students in Rio de Janeiro State, Brazil.. Int J Tuberc Lung Dis, 2005; 9(8):841-7

Tiwari, Bishnu R; Karki, Surendra; Ghimire, Prakash; Sharma, Bimala; Malla, Sarala. Factors associated with high prevalence of pulmonary tuberculosis in HIV-infected people visiting for assessment of eligibility for highly active antiretroviral therapy in Kathmandu, Nepal.. WHO South-East Asia j. public health, 2012; 1(4):404-411

**- Not LMIC**

Altet, M N; Vidal, R; Mila, C; Rodrigo, T; Casals, M; Mir, I; Ruiz-Manzano, J; Jimenez-Fuentes, M A; Sanchez, F; Maldonado, J; Blanquer, R; de Souza-Galvao, M L; Solsona, J; Azlor, E; Diaz, D; Calpe, J L; Cayla, J A. Monitoring changes in anti-tuberculosis treatment: associated factors determined at the time of diagnosis.. Int J Tuberc Lung Dis, 2013; 17(11):1435-41

Chang, Chia-Hao; Chen, Yen-Fu; Wu, Vin-Cent; Shu, Chin-Chung; Lee, Chih-Hsin; Wang, Jann-Yuan; Lee, Li-Na; Yu, Chong-Jen. Acute kidney injury due to anti-tuberculosis drugs: a five-year experience in an aging population.. BMC Infect Dis, 2014; 14(100968551):23

Deng, Wang; Yu, Min; Ma, Hilary; Hu, Liang An; Chen, Gang; Wang, Yong; Deng, Jia; Li, ChangYi; Tong, Jin; Wang, Dao Xin. Predictors and outcome of patients with acute respiratory distress syndrome caused by miliary tuberculosis: a retrospective study in Chongqing, China.. BMC Infect Dis, 2012; 12(100968551):121

Hernandez A.. Chest computed tomography characteristics and time to culture conversion among patients with pulmonary tuberculosis. Open Forum Infect. Dis., 2017; 4(Supplement 1):S720

Hsing S.-C.; Weng S.-F.; Cheng K.-C.; Shieh J.-M.; Chen C.-H.; Chiang S.-R.; Wang J.-J.. Increased risk of pulmonary tuberculosis in patients with previous non-tuberculous mycobacterial disease. Int. J. Tuberc. Lung Dis., 2013; 17(7):928-933

Kang, Young Ae; Kim, Song Yee; Jo, Kyung-Wook; Kim, Hee Jin; Park, Seung-Kyu; Kim, Tae-Hyung; Kim, Eun Kyung; Lee, Ki Man; Lee, Sung Soon; Park, Jae Seuk; Koh, Won-Jung; Kim, Dae Yun; Shim, Tae Sun. Impact of diabetes on treatment outcomes and long-term survival in multidrug-resistant tuberculosis.. Respiration, 2013; 86(6):472-8

Lin, Chou-Han; Lin, Chou-Jui; Kuo, Yao-Wen; Wang, Jann-Yuan; Hsu, Chia-Lin; Chen, Jong-Min; Cheng, Wern-Cherng; Lee, Li-Na. Tuberculosis mortality: patient characteristics and causes.. BMC Infect Dis, 2014; 14(100968551):5

Lin, Pei-Ying; Wang, Jann-Yuan; Hsueh, Po-Ren; Lee, Li-Na; Hsiao, Cheng-Hsiang; Yu, Chong-Jen; Yang, Pan-Chyr. Lower gastrointestinal tract tuberculosis: an important but neglected disease.. Int J Colorectal Dis, 2009; 24(10):1175-80

Matsumura, T; Watanabe, K. [Management of mycobacteriosis in general hospital without isolation ward for tuberculosis patients. 1. Diagnosis and treatment of mycobacterial diseases in a community general hospital].. Kekkaku, 1999; 74(2):129-31

Meira, L.; Chaves, C.; AraÃºjo, D.; Almeida, L.; Boaventura, R.; Ramos, A.; Carvalho, T.; OsÃ³rio, N. S.; Castro, A. G.; Rodrigues, F.; GuimarÃ£es, J. T.; Saraiva, M.; Bastos, H. N.. Predictors and outcomes of disseminated tuberculosis in an intermediate burden setting. Pulmonology, 2019; 25(6):320-327

Moran, Ana; Harbour, Deborah V.; Teeter, Larry D.; Musser, James M.; Graviss, Edward A.. Is alcohol use associated with cavitary disease in tuberculosis?. ALCOHOLISM-CLINICAL AND EXPERIMENTAL RESEARCH, 2007; 31(1):33-38

Riquelme D, Javier; Morales V, Julio; Aguilera V, Rodrigo; Espinoza O, Mauricio; Vidal A, Alexis; Riquelme O, RaÃºl. Impacto de la tuberculosis en el hospital de Puerto Montt. Rev. chil. enferm. respir, 2018; 34(3):165-170

Shen, Te-Chun; Wang, Chien-Yu; Lin, Cheng-Li; Liao, Wei-Chih; Chen, Chia-Hung; Tu, Chih-Yen; Hsia, Te-Chun; Shih, Chuen-Ming; Hsu, Wu-Huei; Chung, Chi-Jung. People with tuberculosis are associated with a subsequent risk of depression.. EUR. J. INTERN. MED., 2014; 25(10):936-40

Sheu, Jau-Jiuan; Chiou, Hung-Yi; Kang, Jiunn-Horng; Chen, Yi-Hua; Lin, Herng-Ching. Tuberculosis and the risk of ischemic stroke: a 3-year follow-up study.. Stroke, 2010; 41(2):244-9

Su Z.; Cheng Y.; Wu Z.; Zhang P.; Chen W.; Zhou Z.; Zhong M.; Luo W.; Guo W.; Li S.. Incidence and predictors of tracheobronchial tuberculosis in pulmonary tuberculosis: A multicentre, large-scale and prospective study in Southern China. Respiration, 2019; 97(2):153-159

Tazawa, S; Marumo, K; Nakamura, Y. [Epidemiological evaluation of mycobacteria isolates in one city hospital: reports from the hospital microbiology laboratory].. Kekkaku, 1997; 72(7):435-42

Tonko, S.; Baty, F.; Brutsche, M. H.; Schoch, O. D.. Length of hospital stay for TB varies with comorbidity and hospital location. The International Journal of Tuberculosis and Lung Disease, 2020; 24(9):948-955

Wu, Huang-Pin; Pan, Yu-Huei; Hua, Chung-Ching; Shieh, Wen-Bin; Jiang, Bor-Yiing; Yu, Teng-Jen. Pneumoconiosis and liver cirrhosis are not risk factors for tuberculosis in patients with pulmonary infection.. Respirology, 2007; 12(3):416-9

**- Wrong publication type**

A.R, Sayo; E.G.M, Balinas; J.A, Verona; A.M.G, Villanueva; S.M, Han; J, Suzuki; K, Ariyoshi; C, Smith; R.M, Solante. COVID-19 screening on a tuberculosis ward in Manila, the Philippines. Journal of Clinical Tuberculosis and Other Mycobacterial Diseases, 2020; 20():100167

Atif M.; Sulaiman S.A.S.; Shafie A.A.; Zaman M.Q.U.; Asif M.. Tuberculosis deaths: Are we measuring accurately?. J. pharm. policy pract., 2014; 7(1):44256

Burki, Talha. MULTIMORBIDITY Why India should worry about a coepidemic of diabetes and tuberculosis. BMJ-BRITISH MEDICAL JOURNAL, 2015; 350():

M.S, Schepisi; J, Miah; A, Kaluzhenina; K.M, Manika; E, Pontali; M.R, Prego; M.N.A, Altet; D.S.M.S, Mellado; E, Girardi. The diabetes-tuberculosis comorbidity among persons with HIV. Topics in Antiviral Medicine, 2017; 25(1):314s

Vilaichone R.-K.; Tumwasorn S.; Wilde H.; Vilaichone W.; Suwanagool P.; Mahachai V.. Clinical spectrum of hepatic tuberculosis: Comparison between immunocompetent and immunocompromised hosts. J. Med. Assoc. Thailand, 2003; 86(SUPPL. 2):S432-S438

**- Wrong study design**

Aibar-Arregui, M A; de Escalante-Yanguela, B; Tejero-Juste, C; Martin-Fortea, M P. [Mixed meningoencephalitis caused by Mycobacterium tuberculosis and varicella zoster virus].. Rev Neurol, 2009; 48(2):91-3

Faurholt-Jepsen D.. The double burden: The role of diabetes for tuberculosis risk, manifestations, treatment outcomes and survival. Dan. Med. J., 2013; 60(7):18

Oscanoa T.; Moscol S.; Fujita R.; Carvajal A.. Hepatotoxicity associated with anti-tuberculosis drugs. A case series from hospitalized patients in Lima, Peru. Drug Saf., 2017; 40(10):982

Tofts R.P.; Alvarez M.; Cardona L.; Smolley L.; Oliveira E.; Ferrer G.. Fast growers vs Slow Growers: Who is at risk for an atypical microbacterial infection?. Chest, 2010; 138(4):

Wang, Qiuzhen; Ma, Aiguo; Schouten, Evert G; Kok, Frans J. A double burden of tuberculosis and diabetes mellitus and the possible role of vitamin D deficiency.. Clin Nutr, 2020; (c3x, 8309603):

Yablonskii P.; Kudriashov G.; Vasiliev I.; Avetisian A.. Robot-assisted lobectomy for multidrug-resistantor extradrug-resistant pulmonary tuberculosis. Interact. Cardiovasc. Thorac. Surg., 2016; 23(Supplement 1):

**- Overlap with another study**

Ponce-De-Leon, Alfredo; Garcia-Garcia Md, Ma de Lourdes; Garcia-Sancho, Ma Cecilia; Gomez-Perez, Francisco J; Valdespino-Gomez, Jose Luis; Olaiz-Fernandez, Gustavo; Rojas, Rosalba; Ferreyra-Reyes, Leticia; Cano-Arellano, Bulmaro; Bobadilla, Miriam; Small, Peter M; Sifuentes-Osornio, Jose. Tuberculosis and diabetes in southern Mexico.. Diabetes Care, 2004; 27(7):1584-90 *- Overlaps with Jimenez-Corona 2013, but covering less years (1995-2003 instead of 1995-2010) and more restrictive age (>19 vs >15)*

Mukhtar, Fatima; Butt, Zahid A. Cohort profile: the diabetes-tuberculosis treatment outcome (DITTO) study in Pakistan. BMJ Open, 2016; 6(12): *- Overlaps with Mukhtar 2018, which reports data on more clusters.*

Workneh, Mahteme Haile; Bjune, Gunnar Aksel; Yimer, Solomon Abebe. Diabetes mellitus is associated with increased mortality during tuberculosis treatment: a prospective cohort study among tuberculosis patients in South-Eastern Amahra Region, Ethiopia.. Infect. dis. poverty, 2016; 5(101606645):22 *- Overlaps with Workneh 2016, with identical results.*

Faurholt-Jepsen, Daniel; Range, Nyagosya; PrayGod, George; Jeremiah, Kidola; Faurholt-Jepsen, Maria; Aabye, Martine G.; Changalucha, John; Christensen, Dirk L.; Krarup, Henrik; Witte, Daniel R.; Andersen, Aase B.; Friis, Henrik. The role of diabetes on the clinical manifestations of pulmonary tuberculosis. Tropical Medicine & International Health, 2012; 17(7): *- Overlaps with Faurholt-Jepsen 2012 with almost identical data.*

Ugarte-Gil, Cesar; Alisjahbana, Bachti; Ronacher, Katharina; Riza, Anca Lelia; Koesoemadinata, Raspati C; Malherbe, Stephanus T; Cioboata, Ramona; Llontop, Juan Carlos; Kleynhans, Leanie; Lopez, Sonia; Santoso, Prayudi; Marius, Ciontea; Villaizan, Katerine; Ruslami, Rovina; Walzl, Gerhard; Panduru, Nicolae Mircea; Dockrell, Hazel M; Hill, Philip C; Mc Allister, Susan; Pearson, Fiona; Moore, David A J; Critchley, Julia A; van Crevel, Reinout. Diabetes Mellitus Among Pulmonary Tuberculosis Patients From 4 Tuberculosis-endemic Countries: The TANDEM Study. Clinical Infectious Diseases, 2019; (): *- Same sample as Ugarte-Gil 2020 but reporting less outcomes of interest.*

Peltzer K.. Conjoint alcohol and tobacco use among tuberculosis patients in public primary healthcare in South Africa. Afr. J. Psychiatry, 2014; 20(1):21-26 *- Overlaps with Peltzer 2013 and Peltzer 2012, but reporting less outcomes.*

Abreu, Ricardo Gadelha de; Sousa, Artur Iuri Alves de; Oliveira, Maria Regina Fernandes de; Sanchez, Mauro Niskier. Tuberculosis and diabetes: probabilistic linkage of databases to study the association between both diseases.. Epidemiol. serv. saude, 2017; 26(2):359-368 *- Partially overlaps with Reis-Santos 2013, but with less outcomes and treatment outcomes data for TB+HIV+DM.*

Reis-Santos, Barbara; Locatelli, Rodrigo; Horta, Bernardo L.; Faerstein, Eduardo; Sanchez, Mauro N.; Riley, Lee W.; Maciel, Ethel Leonor. Socio-Demographic and Clinical Differences in Subjects with Tuberculosis with and without Diabetes Mellitus in Brazil â€“ A Multivariate Analysis. PLoS ONE, 2013; 8(4): *- Uses SINAN, like Reis-Santos 2013, but years 2001-2009, instead of 2001-2011.*

Moreira, Tiago Ricardo; GonÃ§alves, Evelyn Soares Medina; Colodette, Renata Maria; Fernandes, Maiane da Silva; Prado, Mara Rubia Maciel Cardoso; Oliveira, DeÃse Moura. Fatores associados a HIV/AIDS em pacientes com tuberculose em Minas Gerais entre os anos de 2006 e 2015. REME rev. min. enferm, 2019; 23():e-1211 *- Uses SINAN, like Reis-Santos 2013, but years 2006-2015, instead of 2001-2011. We used Reis-Santos 2013 because it includes more participants and we have more detailed data.*

Reis-Santos, Barbara; Gomes, Teresa; Locatelli, Rodrigo; de Oliveira, Elizabete R; Sanchez, Mauro N; Horta, Bernardo L; Riley, Lee W; Maciel, Ethel L. Treatment outcomes in tuberculosis patients with diabetes: a polytomous analysis using Brazilian surveillance system.. PLoS ONE, 2014; 9(7):e100082 *- Uses SINAN, like Reis-Santos 2013, same years (2001-2011), but more restrictive inclusion criteria.*

White L.V.; Edwards T.; Lee N.; Castro M.C.; Saludar N.R.; Calapis R.W.; Faguer B.N.; Garfin C.; Solon J.A.; Cox S.E.. Co-morbidities in filipino persons with tuberculosis: A cross-sectional study in urban and rural public TBDOTS facilities. Trans. R. Soc. Trop. Med. Hyg., 2019; 113(Supplement 1):S210 - *Same sample as White 2020, but reporting less outcomes.*
