## Appendix 3 for "Prevalence, clusters, and burden of complex tuberculosis multimorbidity in low- and middle-income countries: a systematic review and meta-analysis"

**APPENDIX 3: References of included studies**


S1 Dos Santos APC, Lazzari TK, Silva DR. Health-Related Quality of Life, Depression and Anxiety in Hospitalized Patients with Tuberculosis. *Tuberc Respir Dis* 2017;**80**:69–76. doi:10.4046/trd.2017.80.1.69

S2 Yaneth-Giovanetti MC, Morales Parra GI, Herrera C N, *et al.* Frecuencia de diabetes mellitus en pacientes con tratamiento para tuberculosis en Colombia. *Rev Habanera Cienc Méd* 2019;**18**:477–86.

S3 Castro S de S, Scatena LM, Miranzi A, *et al.* Characteristics of cases of tuberculosis coinfected with HIV in Minas Gerais State in 2016. *Rev Inst Med Trop SAO PAULO* 2019;**61**. doi:10.1590/S1678-9946201961021

S4 Kelly AM. Adverse drug reactions and resultant health-related quality of life during multidrug-resistant tuberculosis treatment in South Africa. *Diss Abstr Int Sect B Sci Eng* 2016;**76**:No-Specified.

S5 Carvalho A.C., Nunes Z.B., Martins M., *et al.* Clinical presentation and survival of smear-positive pulmonary tuberculosis patients of a university general hospital in a developing country. *Mem Inst Oswaldo Cruz* 2002;**97**:1225–30.

S6 Weimann A., Dai D., Oni T. A cross-sectional and spatial analysis of the prevalence of multimorbidity and its association with socioeconomic disadvantage in South Africa: A comparison between 2008 and 2012. *Soc Sci Med* 2016;**163**:144–56. doi:10.1016/j.socscimed.2016.06.055

S7 Munoz-Torrico M, Caminero-Luna J, Migliori GB, *et al.* Diabetes is Associated with Severe Adverse Events in Multidrug-Resistant Tuberculosis. *Diabetes Se Asoc Con Reacciones Advers Graves En Tuberc Multirresistente* 2017;**53**:245–50. doi:10.1016/j.arbres.2016.10.021

S8 Azovtzeva O.V., Panteleev A.M., Karpov A.V., *et al.* Analysis of medical and social factors affecting the formation and course of co-infection HIV, tuberculosis and viral hepatitis. *Russ J Infect Immun* 2019;**9**:787–99. doi:10.15789/2220-7619-2019-5-6-787-799

S9 Blal CA, Passos SRL, Horn C, *et al.* High prevalence of hepatitis B virus infection among tuberculosis patients with and without HIV in Rio de Janeiro, Brazil. *Eur J Clin Microbiol Infect Dis Off Publ Eur Soc Clin Microbiol* 2005;**24**:41–3.

S10 Peltzer K, Louw J, McHunu G, *et al.* Hazardous and harmful alcohol use and associated factors in tuberculosis public primary care patients in South Africa. *Int J Environ Res Public Health* 2012;**9**:3245–57. doi:10.3390/ijerph9093245

S11 Jimenez-Corona ME, Cruz-Hervert LP, Garcia-Garcia L, *et al.* Association of diabetes and tuberculosis: impact on treatment and post-treatment outcomes. *Thorax* 2013;**68**:214–20. doi:10.1136/thoraxjnl-2012-201756

S12 Pepper DJ, Marais S, Wilkinson RJ, *et al.* Clinical deterioration during antituberculosis treatment in Africa: incidence, causes and risk factors. *BMC Infect Dis* 2010;**10**:83. doi:10.1186/1471-2334-10-83

S13 Azeez A, Ndege J, Mutambayi R. Associated factors with unsuccessful tuberculosis treatment outcomes among tuberculosis/HIV coinfected patients with drug-resistant tuberculosis. *Int J Mycobacteriology* 2018;**7**:347–54. doi:10.4103/ijmy.ijmy_140_18

S14 Whitehouse ER, Perrin N, Levitt N, *et al.* Cardiovascular risk prevalence in South Africans with drug-resistant tuberculosis: a cross-sectional study. *Int J Tuberc Lung Dis Off J Int Union Tuberc Lung Dis* 2019;**23**:587–93. doi:10.5588/ijtld.18.0374

S15 de Oliveira NF, Goncalves MJF. Social and environmental factors associated with the hospitalization of tuberculosis patients. *Rev Lat Am Enfermagem* 2013;**21**:507–14. doi:10.1590/S0104-11692013000200006

S16 Peltzer K, Naidoo P, Matseke G, *et al.* Prevalence of post-traumatic stress symptoms and associated factors in tuberculosis (TB), TB retreatment and/or TB-HIV co-infected primary public health-care patients in three districts in South Africa. *Psychol Health Med* 2013;**18**:387–97. doi:10.1080/13548506.2012.726364

S17 Heredia N.S., Lezama M.A.S. Tuberculosis and diabetes mellitus in the Sanitary District No. 2 of the state of Guerrero. A brief report of a descriptive study. *Rev Inst Nac Enfermedades Respir* 2011;**70**:152–6.

S18 Alavi SM, Khoshkhoy MM. Pulmonary tuberculosis and diabetes mellitus: Co-existence of both diseases in patients admitted in a teaching hospital in the southwest of Iran. *Casp J Intern Med* 2012;**3**:421–4.

S19 Aires R.S., Matos M.A.D., Lopes C.L.R., *et al.* Prevalence of hepatitis B virus infection among tuberculosis patients with or without HIV in Goiania City, Brazil. *J Clin Virol* 2012;**54**:327–31. doi:10.1016/j.jcv.2012.04.006

S20 Shokri M., Najafi R., Niromand J., *et al.* The frequency of risk factors for pulmonary tuberculosis in tuberculosis patients in Babol, Northern Iran, during 2008-2015. *Curr Issues Pharm Med Sci* 2018;**31**:144–7. doi:10.1515/cipms-2018-0028

S21 Reis-Santos B, Gomes T, Macedo LR, *et al.* Prevalence and patterns of multimorbidity among tuberculosis patients in Brazil: a cross-sectional study. *Int J Equity Health* 2013;**12**:61. doi:10.1186/1475-9276-12-61

S22 Oni T, Youngblood E, Boulle A, *et al.* Patterns of HIV, TB, and non-communicable disease multi-morbidity in peri-urban South Africa- a cross sectional study. *BMC Infect Dis* 2015;**15**:20. doi:10.1186/s12879-015-0750-1

S23 Jalal TMT, Abdullah S, Wahab FA, *et al.* Prevalence and Factors Associated with Tuberculosis Treatment Success among TB/HIV Co-Infection in North-East Malaysia. *Malays J Med Sci MJMS* 2017;**24**:75–82. doi:10.21315/mjms2017.24.6.9

S24 Echazarreta A, Zerbini E, De Sandro J, *et al.* [Tuberculosis and comorbidities in urban areas in Argentina. A gender and age perspective]. *Biomed Rev Inst Nac Salud* 2018;**38**:180–8. doi:10.7705/biomedica.v38i0.3904

S25 Khan AH, Sulaiman SAS, Laghari M, *et al.* Treatment outcomes and risk factors of extra-pulmonary tuberculosis in patients with co-morbidities. *BMC Infect Dis* 2019;**19**:691. doi:10.1186/s12879-019-4312-9

S26 Sharman M, Bachmann M. Prevalence and health effects of communicable and non-communicable disease comorbidity in rural KwaZulu-Natal, South Africa. *Trop Med Int Health TM IH* 2019;**24**:1198–207. doi:10.1111/tmi.13297

S27 Cespedes C, Lopez L, Aguirre S, *et al.* [Prevalence of comorbidity tuberculosis and diabetes mellitus in Paraguay, 2016 and 2017Prevalencia de comorbidade tuberculose-diabetes mellitus no Paraguai, 2016 e 2017]. *Prevalencia Comorbilidad Tuberc Diabetes Mellit En Parag 2016 2017* 2019;**43**:e105. doi:10.26633/RPSP.2019.105

S28 Modongo C, Zetola NM. Prevalence of hypothyroidism among MDR-TB patients in Botswana [Correspondence]. *Int J Tuberc Lung Dis* 2012;**16**. doi:10.5588/ijtld.12.0403

S29 Appana D, Joseph L, Paken J. An audiological profile of patients infected with multi-drug resistant tuberculosis at a district hospital in KwaZulu-Natal. *S Afr J Commun Disord* 2016;**63**. doi:10.4102/sajcd.v63i1.154

S30 Pando MA, De Salvo C, Bautista CT, *et al.* Human immunodeficiency virus and tuberculosis in Argentina: prevalence, genotypes and risk factors. *J Med Microbiol* 2008;**57**. doi:10.1099/jmm.0.47492-0

S31 Reis NRS, Lopes CLR, Teles SA, *et al.* Hepatitis C virus infection in patients with tuberculosis in Central Brazil. *Int J Tuberc Lung Dis* 2011;**15**. doi:10.5588/ijtld.10.0636

S32 Wang X, Li X, Zhang Q, *et al.* A Survey of Anxiety and Depressive Symptoms in Pulmonary Tuberculosis Patients With and Without Tracheobronchial Tuberculosis. *Front Psychiatry* 2018;**9**. doi:10.3389/fpsyt.2018.00308

S33 do Prado TN, Miranda AE, de Souza FM, *et al.* Factors associated with tuberculosis by HIV status in the Brazilian national surveillance system: a cross sectional study. *BMC Infect Dis* 2014;**14**. doi:10.1186/1471-2334-14-415

S34 Delgado-Sánchez G, García-García L, Castellanos-Joya M, *et al.* Association of Pulmonary Tuberculosis and Diabetes in Mexico: Analysis of the National Tuberculosis Registry 2000–2012. *PLOS ONE* 2015;**10**. doi:10.1371/journal.pone.0129312

S35 Oni T, Berkowitz N, Kubjane M, *et al.* Trilateral overlap of tuberculosis, diabetes and HIV-1 in a high-burden African setting: implications for TB control. *Eur Respir J* 2017;**50**. doi:10.1183/13993003.00004-2017

S36 Tabarsi P, Baghaei P, Marjani M, *et al.* Changes in glycosylated haemoglobin and treatment outcomes in patients with tuberculosis in Iran: a cohort study. *J Diabetes Metab Disord* 2014;**13**. doi:10.1186/s40200-014-0123-0

S37 Gupta S, Shenoy VP, Bairy I, *et al.* Diabetes mellitus and HIV as co-morbidities in tuberculosis patients of rural south India. *J Infect Public Health* 2011;**4**:140–4. doi:10.1016/j.jiph.2011.03.005

S38 Naik B., Kumar A.M.V., Satyanarayana S., *et al.* Is screening for diabetes among tuberculosis patients feasible at the field level? *Public Health Action* 2013;**3**:S34–7. doi:10.5588/pha.13.0022

S39 Khanna A., Lohya S., Sharath B.N., *et al.* Characteristics and treatment response in patients with tuberculosis and diabetes mellitus in New Delhi, India. *Public Health Action* 2013;**3**:S48–50. doi:10.5588/pha.13.0025

S40 Shyamala KK, Naveen RS, Khatri B. Depression: A Neglected Comorbidity in Patients with Tuberculosis. *J Assoc Physicians India* 2018;**66**:18–21.

S41 Achanta S, Tekumalla RR, Jaju J, *et al.* Screening tuberculosis patients for diabetes in a tribal area in South India. *Public Health Action* 2013;**3**. doi:10.5588/pha.13.0033

S42 Piparva K.G. Treatment outcome of tuberculosis patients on dots therapy for category 1 and 2 at district tuberculosis centre. *Int J Pharm Sci Res* 2017;**8**:207–12. doi:10.13040/IJPSR.0975-8232.8%281%29.207-12

S43 Dave P, Shah A, Chauhan M, *et al.* Screening patients with tuberculosis for diabetes mellitus in Gujarat, India. *Public Health Action* 2013;**3**:S29-33. doi:10.5588/pha.13.0027

S44 Pande T, Huddart S, Xavier W, *et al.* Prevalence of diabetes mellitus amongst hospitalized tuberculosis patients at an Indian tertiary care center: A descriptive analysis. *PLOS ONE* 2018;**13**. doi:10.1371/journal.pone.0200838

S45 Viswanathan V, Vigneswari A, Selvan K, *et al.* Effect of diabetes on treatment outcome of smear-positive pulmonary tuberculosis—A report from South India. *J Diabetes Complications* 2014;**28**. doi:10.1016/j.jdiacomp.2013.12.003

S46 Singh Pawar DK. Prevalence of Co morbidities among patients having Multi Drug Resistant Tuberculosis: A Retrospective Analysis. *J Med Sci Clin Res* 2018;**6**:2019–1032. doi:10.18535/jmscr/v6i4.168

S47 KV N, Duraisamy K, Balakrishnan S, *et al.* Outcome of Tuberculosis Treatment in Patients with Diabetes Mellitus Treated in the Revised National Tuberculosis Control Programme in Malappuram District, Kerala, India. *PLoS ONE* 2013;**8**. doi:10.1371/journal.pone.0076275

S48 Ambaw F, Mayston R, Hanlon C, *et al.* Burden and presentation of depression among newly diagnosed individuals with TB in primary care settings in Ethiopia. *BMC Psychiatry* 2017;**17**:57. doi:10.1186/s12888-017-1231-4

S49 Deribew A, Tesfaye M, Hailmichael Y, *et al.* Tuberculosis and HIV co-infection: its impact on quality of life. *Health Qual Life Outcomes* 2009;**7**:105. doi:10.1186/1477-7525-7-105

S50 Duko B, Gebeyehu A, Ayano G. Prevalence and correlates of depression and anxiety among patients with tuberculosis at WolaitaSodo University Hospital and Sodo Health Center, WolaitaSodo, South Ethiopia, Cross sectional study. *BMC Psychiatry* 2015;**15**:214. doi:10.1186/s12888-015-0598-3

S51 Dasa TT, Roba AA, Weldegebreal F, *et al.* Prevalence and associated factors of depression among tuberculosis patients in Eastern Ethiopia. *BMC Psychiatry* 2019;**19**:82. doi:10.1186/s12888-019-2042-6

S52 Molla A, Mekuriaw B, Kerebih H. Depression and associated factors among patients with tuberculosis in Ethiopia: a cross-sectional study. *Neuropsychiatr Dis Treat* 2019;**Volume 15**. doi:10.2147/NDT.S208361

S53 Adem A, Tesfaye M, Mohammed M. The Prevalence and Pattern of Depression in Patients with Tuberculosis on Follow up at Jimma University Specialized Hospital and Jimma Health Center. *Med Sci Int Med J* 2014;**3**. doi:10.5455/medscience.2013.02.8097

S54 Getachew A, Mekonnen S, Alemu S. High magnitude of diabetes mellitus among active pulmonary tuberculosis patients in Ethiopia. *J Adv Med Med Res* 2014;:862–72.

S55 Workneh MH, Bjune GA, Yimer SA. Prevalence and Associated Factors of Diabetes Mellitus among Tuberculosis Patients in South-Eastern Amhara Region, Ethiopia: A Cross Sectional Study. *PloS One* 2016;**11**:e0147621. doi:10.1371/journal.pone.0147621

S56 Mohammedhussein M, Alenko A, Tessema W, *et al.* Prevalence and Associated Factors of Depression and Anxiety Among Patients with Pulmonary Tuberculosis Attending Treatment at Public Health Facilities in Southwest Ethiopia. *Neuropsychiatr Dis Treat* 2020;**16**:1095–104. doi:10.2147/NDT.S249431

S57 Damtew E, Ali I, Meressa D. Prevalence of Diabetes Mellitus among Active Pulmonary Tuberculosis Patients at St. Peter Specialized Hospital, Addis Ababa, Ethiopia. *World J Med Sci* 2014;**11**:389–96.

S58 Richards DC, Mikiashvili T, Parris JJ, *et al.* High prevalence of hepatitis C virus but not HIV co-infection among patients with tuberculosis in Georgia. *Int J Tuberc Lung Dis* 2006;**10**:396–401.

S59 Kuniholm MH, Mark J, Aladashvili M, *et al.* Risk factors and algorithms to identify hepatitis C, hepatitis B, and HIV among Georgian tuberculosis patients. *Int J Infect Dis* 2008;**12**. doi:10.1016/j.ijid.2007.04.015

S60 Ugarte-Gil C, Alisjahbana B, Ronacher K, *et al.* Diabetes Mellitus Among Pulmonary Tuberculosis Patients From 4 Tuberculosis-endemic Countries: The TANDEM Study. *Clin Infect Dis Off Publ Infect Dis Soc Am* 2020;**70**:780–8. doi:10.1093/cid/ciz284

S61 Owiti P, Keter A, Harries AD, *et al.* Diabetes and pre-diabetes in tuberculosis patients in western Kenya using point-of-care glycated haemoglobin. *Public Health Action* 2017;**7**. doi:10.5588/pha.16.0114

S62 Adejumo OA, Olusola-Faleye B, Adepoju VA, *et al.* The pattern of comorbidity and its prevalence among drug-resistant tuberculosis patients at treatment initiation in Lagos, Nigeria. *Trans R Soc Trop Med Hyg* 2020;**114**:415–23. doi:10.1093/trstmh/trz126

S63 Ekeke N, Ukwaja KN, Chukwu JN, *et al.* Screening for diabetes mellitus among tuberculosis patients in Southern Nigeria: a multi-centre implementation study under programme settings. *Sci Rep* 2017;**7**. doi:10.1038/srep44205

S64 Javed I., Javed M.T., Mahmood Z., *et al.* Hematological profiling of tuberculosis-infected and co-morbid patients: A study carried out in central Punjab, Pakistan. *Eur J Inflamm* 2018;**16**. doi:10.1177/2058739218818684

S65 White LV, Edwards T, Lee N, *et al.* Patterns and predictors of co-morbidities in Tuberculosis: A cross-sectional study in the Philippines. *Sci Rep* 2020;**10**:4100. doi:10.1038/s41598-020-60942-2

S66 Abdelsalam M. N, Nazar E. A, Mohammed O. E. G. Seroprevalence of hepatitis B and C viruses among tuberculosis patients. *Sudan J Med Sci* 2013;**8**:17–22.

S67 Munseri P.J., Kimambo H., Pallangyo K. Diabetes mellitus among patients attending TB clinics in Dar es Salaam: A descriptive cross-sectional study. *BMC Infect Dis* 2019;**19**:915. doi:10.1186/s12879-019-4539-5

S68 Kibirige D., Ssekitoleko R., Mutebi E., *et al.* Overt diabetes mellitus among newly diagnosed Ugandan tuberculosis patients: A cross sectional study. *BMC Infect Dis* 2013;**13**:122. doi:10.1186/1471-2334-13-122

S69 Alinaitwe R. Prevalence and factors associated with depressive illness in patients with tuberculosis in Mulago Hospital. 2018.

S70 van den Heuvel L, Chishinga N, Kinyanda E, *et al.* Frequency and correlates of anxiety and mood disorders among TB- and HIV-infected Zambians. *AIDS Care* 2013;**25**:1527–35. doi:10.1080/09540121.2013.793263

S71 Boillat-Blanco N, Ramaiya KL, Mganga M, *et al.* Transient Hyperglycemia in Patients With Tuberculosis in Tanzania: Implications for Diabetes Screening Algorithms. *J Infect Dis* 2016;**213**. doi:10.1093/infdis/jiv568

S72 Wu Z, Guo J, Huang Y, *et al.* Diabetes mellitus in patients with pulmonary tuberculosis in an aging population in Shanghai, China: Prevalence, clinical characteristics and outcomes. *J Diabetes Complications* 2016;**30**. doi:10.1016/j.jdiacomp.2015.11.014

S73 Magee MJ, Kempker RR, Kipiani M, *et al.* Diabetes mellitus is associated with cavities, smear grade, and multidrug-resistant tuberculosis in Georgia. *Int J Tuberc Lung Dis* 2015;**19**. doi:10.5588/ijtld.14.0811

S74 Vashakidze S, Despuig A, Gogishvili S, *et al.* Retrospective study of clinical and lesion characteristics of patients undergoing surgical treatment for Pulmonary Tuberculosis in Georgia. *Int J Infect Dis IJID Off Publ Int Soc Infect Dis* 2017;**56**:200–7. doi:10.1016/j.ijid.2016.12.009

S75 Walker IF, Kanal S, Baral SC, *et al.* Depression and anxiety in patients with multidrug-resistant tuberculosis in Nepal: an observational study. *Public Health Action* 2019;**9**:42–8. doi:10.5588/pha.18.0047

S76 Mukhtar F, Butt ZA. Risk of adverse treatment outcomes among new pulmonary TB patients co-infected with diabetes in Pakistan: A prospective cohort study. *PLOS ONE* 2018;**13**. doi:10.1371/journal.pone.0207148

S77 Velásquez GE, Cegielski JP, Murray MB, *et al.* Impact of HIV on mortality among patients treated for tuberculosis in Lima, Peru: a prospective cohort study. *BMC Infect Dis* 2015;**16**. doi:10.1186/s12879-016-1375-8

S78 Krupitsky EM, Zvartau EE, Lioznov DA, *et al.* Co-morbidity of infectious and addictive diseases in St. Petersburg and the Leningrad Region, Russia. *Eur Addict Res* 2006;**12**:12–9.

S79 Faurholt-Jepsen D, Range N, Praygod G, *et al.* The role of diabetes co-morbidity for tuberculosis treatment outcomes: a prospective cohort study from Mwanza, Tanzania. *BMC Infect Dis* 2012;**12**:165. doi:10.1186/1471-2334-12-165

S80 Manosuthi W, Kawkitinarong K, Suwanpimolkul G, *et al.* Clinical characteristics and treatment outcomes among patients with tuberculosis in Bangkok and Nonthaburi, Thailand. *Southeast Asian J Trop Med Public Health* 2012;**43**:1426–36.

S81 Sirinak C, Kittikraisak W, Pinjeesekikul D, *et al.* Viral hepatitis and HIV-associated tuberculosis: Risk factors and TB treatment outcomes in Thailand. *BMC Public Health* 2008;**8**. doi:10.1186/1471-2458-8-245

S82 Chin A.T., Rylance J., Makumbirofa S., *et al.* Chronic lung disease in adult recurrent tuberculosis survivors in Zimbabwe: A cohort study. *Int J Tuberc Lung Dis* 2019;**23**:203–11. doi:10.5588/ijtld.18.0313
