## Supplemental file for "Prevalence, clusters, and burden of complex tuberculosis multimorbidity in low- and middle-income countries: a systematic review and meta-analysis"

#### Medline (October 1st 2020)

| # ▲ | Searches | Results |
|---|---|---|
| 1 | exp Noncommunicable Diseases/ or ((Non-communicable or Noncommunicable or Non-infectious) adj (disease* or condition* or illness*)).mp. | 11623 |
| 2 | exp Chronic Disease/ or ((chronic or long-term) adj (disease* or condition* or illness*)).mp. | 342329 |
| 3 | exp Heart Diseases/ or (heart adj (disease* or disorder* or failure)).mp. or (cardiac adj (disease* or disorder* or failure)).mp. | 1249276 |
| 4 | exp Cardiovascular Diseases/ or (cardiovascular adj (disease* or disorder* or failure)).mp. | 2475828 |
| 5 | exp Coronary Disease/ or (coronary adj (disease* or disorder* or failure)).mp. | 224195 |
| 6 | exp Cerebrovascular Disorders/ or (cerebrovascular adj (disease* or disorder* or insufficienc* or occlusion*)).mp. or (vascular adj (disease* or disorder*)).mp. or (carotid* adj (disease* or disorder*)).mp. | 459328 |
| 7 | exp Peripheral Arterial Disease/ or (arter* adj (disease* or disorder*)).mp. | 172280 |
| 8 | exp Rheumatic Heart Disease/ or exp Heart Defects, Congenital/ or (heart adj3 (malform* or defect* or congeni*)).mp. | 181794 |
| 9 | exp Venous Thrombosis/ or ((deep vein or deep venous) adj thrombos*).mp. or phlebothrombos*.mp. | 68279 |
| 10 | exp Pulmonary Embolism/ or (pulmonar* adj (thromboembolism* or embolism* or disease* or disorder*)).mp. | 137734 |
| 11 | exp Stroke/ or stroke.mp. | 319373 |
| 12 | exp Neoplasms/ or Cancer*.mp. or neoplas*.mp. or tumor*.mp. | 4407774 |
| 13 | exp Lung Diseases/ or exp Respiratory Tract Diseases/ or exp Lung Diseases, Obstructive/ or ((lung* or respiratory or pulmonar* or airflow or airway) adj2 (disease* or obstruct* or hypersensitiv*)).mp. or exp Asthma/ or asthma*.mp. or exp Pulmonary Disease, Chronic Obstructive/ or exp Respiratory Hypersensitivity/ | 1466914 |
| 14 | exp Diabetes Mellitus/ or diabet*.mp. | 708447 |
| 15 | exp Autoimmune Diseases/ or ((autoimmun* or auto immun* or autoaggress* or auto aggress*) adj (disorder* or disease*)).mp. | 515503 |
| 16 | exp Metabolic Syndrome/ or exp Metabolic Diseases/ or ((metabolic or insulin resistance) adj (disorder* or disease* or syndrome*)).mp. | 1064607 |
| 17 | exp Obesity/ or obes*.mp. | 364350 |
| 18 | exp Osteoporosis/ or osteoporo*.mp. or bone loss.mp. or exp osteolysis/ or osteolysis.mp. or bone resorption.mp. | 153182 |

| | | |
|---|---|---|
| 19 | exp Parkinson disease/ or parkinson*.mp. or paralysis agitans.mp. | 131069 |
| 20 | exp Arthritis/ or arthriti*.mp. or polyarthriti*.mp. or rheumarthriti*.mp. | 320672 |
| 21 | exp Kidney Diseases/ or (kidney adj (disease* or disorder*)).mp. | 543248 |
| 22 | exp Liver Diseases/ or (liver adj (disease* or disorder* or dysfunction*)).mp. | 590473 |
| 23 | exp Hypertension/ or high blood pressure*.mp. or hypertens*.mp. | 522163 |
| 24 | exp Hyperlipidemias/ or hyperlipem*.mp. or hyperlipidem*.mp. or lipem*.mp. or lipidem*.mp. | 84639 |
| 25 | exp Hypercholesterolemia/ or ((high* or elevat*) adj cholesterol*).mp. or hypercholesterem*.mp. or hypercholesterolem*.mp. | 50852 |
| 26 | exp Hypertriglyceridemia/ or hypertriglyceridem*.mp. | 15240 |
| 27 | exp Thyroid Diseases/ or (thyroid adj (disease* or disorder*)).mp. or exp Hyperthyroidism/ or hyperthyroid*.mp. or exp Hypothyroidism/ or hypothyroid*.mp. or ((thyroid-stimulating hormone* or tsh) adj deficien*).mp. | 166261 |
| 28 | exp Motor Neuron Disease/ or motor neuron* disease*.mp. or lateral scleros*.mp. or motor system disease*.mp. | 38287 |
| 29 | exp Multiple Sclerosis/ or multiple sclerosis.mp. or disseminated sclerosis.mp. | 84526 |
| 30 | exp Emphysema/ or emphysema*.mp. | 38477 |
| 31 | exp Bronchitis/ or bronchit*.mp. | 41164 |
| 32 | 1 or 2 or 3 or 4 or 5 or 6 or 7 or 8 or 9 or 10 or 11 or 12 or 13 or 14 or 15 or 16 or 17 or 18 or 19 or 20 or 21 or 22 or 23 or 24 or 25 or 26 or 27 or 28 or 29 or 30 or 31 | 10268254 |
| 33 | exp Mental Disorders/ or exp Psychotic Disorders/ or ((mental* or psychiatr* or psycho*) adj (disorder* or disease* or illness*)).mp. | 1310446 |
| 34 | exp Depressive Disorder, Major/ or exp Depression/ or Depress*.mp. or MDD.mp. | 551557 |
| 35 | exp Anxiety Disorders/ or exp Anxiety/ or anxi*.mp. | 288539 |
| 36 | exp Phobic Disorders/ or phobi*.mp. | 17668 |
| 37 | exp Schizophrenia/ or schizophreni*.mp. or hebephreni*.mp. | 150317 |
| 38 | exp Somatoform Disorders/ or ((somatoform* or somati* or medically unexplained or briquet or pain) adj (disorder* or syndrome* or symptom*)).mp. or exp Medically Unexplained Symptoms/ | 49666 |
| 39 | exp Dissociative Disorders/ or (dissociative adj (disorder* or hysteri* or reaction*)).mp. or dissociation*.mp. | 115502 |
| 40 | exp Hysteria/ or hysteri*.mp. | 6087 |
| 41 | exp Mood Disorders/ or ((affective* or mood*) adj (disorder* or disease* or illness* or symptom*)).mp. | 158786 |

| | | |
|---|---|---|
| 42 | exp Stress Disorders, Post-Traumatic/ or PTSD.mp. or ((post trauma* or posttrauma*) adj (stress* or neurose*)).mp. or combat disorder*.mp. or war disorder*.mp. | 48182 |
| 43 | exp Cognition Disorders/ or ((cognitive or cognition or mental or neurocognitive) adj (dysfunction* or decline* or impairment* or deterioration* or disorder* or illness* or disease*)).mp. | 376563 |
| 44 | exp Personality Disorders/ or personality disorder*.mp. | 48806 |
| 45 | exp "Disruptive, Impulse Control, and Conduct Disorders"/ or impulse control disorder*.mp. or intermittent explosive disorder*.mp. | 9750 |
| 46 | exp "Feeding and Eating Disorders"/ or ((eating or appetite or feeding) adj disorder*).mp. | 38291 |
| 47 | exp Bipolar Disorder/ or ((bipolar or mani*) adj (disorder* or illness* or disease*)).mp. | 51981 |
| 48 | exp Obsessive-Compulsive Disorder/ or OCD*.mp. or ((obsess*-compulsi* or obsess* or compulsi*) adj (disorder* or illness* or disease* or neuros*)).mp. | 22104 |
| 49 | exp Panic Disorder/ or (panic adj (attack* or disorder*)).mp. | 12661 |
| 50 | exp Agoraphobia/ or agoraphobi*.mp. | 4210 |
| 51 | exp Neurotic Disorders/ or neuros*.mp. or neurotic disorder*.mp. or psychoneuros*.mp. | 190586 |
| 52 | 33 or 34 or 35 or 36 or 37 or 38 or 39 or 40 or 41 or 42 or 43 or 44 or 45 or 46 or 47 or 48 or 49 or 50 or 51 | 2104181 |
| 53 | exp Communicable Diseases/ or ((communic* or contag* or transmi* or infect*) adj (disease* or infection* or illness*)).mp. | 212116 |
| 54 | exp Bacterial Infections/ or bacteri* infection*.mp. | 914677 |
| 55 | exp Conjunctivitis/ or conjunctivitis.mp. | 24660 |
| 56 | exp HIV/ or hiv.mp. or Human immuno deficiency virus.mp. | 362241 |
| 57 | exp Acquired Immunodeficiency Syndrome/ or AIDS.mp. or immunodeficiency associated virus.mp. or immun* deficiency associated virus.mp. or acquired immunodeficiency syndrome*.mp. or acquired immun* deficiency syndrome*.mp. | 226631 |
| 58 | exp Buruli Ulcer/ or Bairnsdale.mp. or Buruli.mp. | 1103 |
| 59 | exp Onchocerciasis/ or onchocer*.mp. | 6013 |
| 60 | hepatitis.mp. or exp Hepatitis B/ or exp Hepatitis C/ | 256737 |
| 61 | exp Leishmaniasis/ or leishmania*.mp. | 37900 |
| 62 | exp Leprosy/ or lepros*.mp. or hansen*.mp. | 31195 |
| 63 | exp Elephantiasis, Filarial/ or elephantias*.mp. or filaria*.mp. | 14422 |
| 64 | exp Trachoma/ or egyptian ophthalmia*.mp. or trachoma*.mp. | 20426 |
| 65 | exp Chikungunya Fever/ or chickungunya.mp. or chikungunya.mp. | 5752 |

| | | |
|---|---|---|
| 66 | exp Taeniasis/ or taenia*.mp. | 12356 |
| 67 | exp Cysticercosis/ or cysticercos*.mp. | 7142 |
| 68 | exp Echinococcosis/ or hydatid*.mp. or echinococc*.mp. | 29881 |
| 69 | exp Chagas Disease/ or trypanosom*.mp. or chagas.mp. | 44224 |
| 70 | exp Trypanosomiasis/ or sleeping sickness.mp. | 23384 |
| 71 | exp Encephalitis, Japanese/ or (japanese adj3 encephalitis).mp. | 5840 |
| 72 | exp Syphilis/ or syphilis.mp. | 37541 |
| 73 | 53 or 54 or 55 or 56 or 57 or 58 or 59 or 60 or 61 or 62 or 63 or 64 or 65 or 66 or 67 or 68 or 69 or 70 or 71 or 72 | 1870766 |
| 74 | exp Tuberculosis/ | 192575 |
| 75 | Tuberculos*.mp. | 256277 |
| 76 | TB.mp. | 56723 |
| 77 | koch*.mp. | 9432 |
| 78 | exp Tuberculosis/ or Tuberculos*.mp. or TB.mp. or koch*.mp. | 283267 |
| 79 | (multiple adj (ill* or disease* or condition* or syndrom* or disorder*)).mp. | 5502 |
| 80 | ((Cooccur* or co-occur* or coexist* or co-exist* or multipl* or concord* or discord* or long-term or physical*) adj3 (disease* or ill* or care or condition* or disorder* or health* or medication* or symptom* or syndrom*)).mp. | 268878 |
| 81 | (comorbid* or multimorbid* or co-occurren* or co-morbid* or Multidisease* or multi-disease*).mp. | 267040 |
| 82 | (comorbid* or multimorbid* or co-occurren* or co-morbid* or multi-morbid* or Multidisease* or multi-disease*).mp. | 267538 |
| 83 | exp Comorbidity/ or exp Multimorbidity/ or exp Multiple Chronic Conditions/ | 110592 |
| 84 | 79 or 80 or 81 or 82 or 83 | 517305 |
| 85 | exp "Systematic Review"/ | 135796 |
| 86 | "systematic review*".m_titl. | 133397 |
| 87 | exp Meta-Analysis/ | 120248 |
| 88 | "meta-analys*".m_titl. | 114268 |
| 89 | exp "Systematic Review"/ or "systematic review*".m_titl. or exp Meta-Analysis/ or "meta-analys*".m_titl. | 258179 |
| 90 | 32 or 52 or 73 | 13034344 |
| 91 | (32 or 52 or 73) and 78 | 228625 |
| 92 | (32 or 52 or 73) and 78 and 84 | 4052 |

| | | |
|---|---|---|
| 93 | (32 or 52 or 73) and 78 and 84 and 89 | 89 |
| 94 | exp Animals/ not exp Humans/ | 4738878 |
| **95** | **((32 or 52 or 73) and 78 and 84) not 94** | **3984** |

#### Embase (October 1st 2020)

| # ▲ | Searches | Results |
|---|---|---|
| 1 | exp non communicable disease/ or ((Non-communicable or Noncommunicable or Non-infectious) adj (disease* or condition* or illness*)).mp. | 16262 |
| 2 | exp chronic disease/ or ((chronic or long-term) adj (disease* or condition* or illness*)).mp. | 267208 |
| 3 | exp heart disease/ or (heart adj (disease* or disorder* or failure)).mp. or (cardiac adj (disease* or disorder* or failure)).mp. | 1924675 |
| 4 | exp cardiovascular disease/ or (cardiovascular adj (disease* or disorder* or failure)).mp. | 4148975 |
| 5 | exp coronary artery disease/ or (coronary adj (disease* or disorder* or failure)).mp. | 336818 |
| 6 | exp cerebrovascular disease/ or (cerebrovascular adj (disease* or disorder* or insufficienc* or occlusion*)).mp. or (vascular adj (disease* or disorder*)).mp. or (carotid* adj (disease* or disorder*)).mp. | 754592 |
| 7 | exp peripheral occlusive artery disease/ or (arter* adj (disease* or disorder*)).mp. | 425386 |
| 8 | exp rheumatic heart disease/ or exp congenital heart malformation/ or (heart adj3 (malform* or defect* or congeni*)).mp. | 186148 |
| 9 | exp vein thrombosis/ or ((deep vein or deep venous) adj thrombos*).mp. or phlebothrombos*.mp. | 136389 |
| 10 | exp lung embolism/ or (pulmonar* adj (thromboembolism* or embolism* or disease* or disorder*)).mp. | 203846 |
| 11 | exp cerebrovascular accident/ or stroke.mp. | 509268 |
| 12 | exp neoplasm/ or Cancer*.mp. or neoplas*.mp. or tumor*.mp. | 5775772 |
| 13 | exp lung disease/ or exp respiratory tract disease/ or exp chronic obstructive lung disease/ or ((lung* or respiratory or pulmonar* or airflow or airway) adj2 (disease* or obstruct* or hypersensitiv*)).mp. or exp asthma/ or asthma*.mp. or exp chronic obstructive lung disease/ or exp respiratory tract allergy/ | 2513711 |
| 14 | exp diabetes mellitus/ or diabet*.mp. | 1158554 |
| 15 | exp autoimmune disease/ or ((autoimmun* or auto immun* or autoaggress* or auto aggress*) adj (disorder* or disease*)).mp. | 623414 |
| 16 | exp metabolic syndrome X/ or exp metabolic disorder/ or ((metabolic or insulin resistance) adj (disorder* or disease* or syndrome*)).mp. | 2713640 |
| 17 | exp obesity/ or obes*.mp. | 628127 |
| 18 | exp osteoporosis/ or osteoporo*.mp. or bone loss.mp. or exp osteolysis/ or osteolysis.mp. or bone resorption.mp. | 236458 |
| 19 | exp Parkinson disease/ or parkinson*.mp. or paralysis agitans.mp. | 211005 |

| | | |
|---|---|---|
| 20 | exp arthritis/ or arthriti*.mp. or polyarthriti*.mp. or rheumarthriti*.mp. | 506546 |
| 21 | exp kidney disease/ or (kidney adj (disease* or disorder*)).mp. | 968536 |
| 22 | exp liver disease/ or (liver adj (disease* or disorder* or dysfunction*)).mp. | 1016005 |
| 23 | exp hypertension/ or high blood pressure*.mp. or hypertens*.mp. | 1004479 |
| 24 | exp hyperlipidemia/ or hyperlipem*.mp. or hyperlipidem*.mp. or lipem*.mp. or lipidem*.mp. | 169810 |
| 25 | exp hypercholesterolemia/ or ((high* or elevat*) adj cholesterol*).mp. or hypercholesterem*.mp. or hypercholesterolem*.mp. | 90475 |
| 26 | exp hypertriglyceridemia/ or hypertriglyceridem*.mp. | 31544 |
| 27 | exp thyroid disease/ or (thyroid adj (disease* or disorder*)).mp. or exp hyperthyroidism/ or hyperthyroid*.mp. or exp hypothyroidism/ or hypothyroid*.mp. or ((thyroid-stimulating hormone* or tsh) adj deficien*).mp. | 237331 |
| 28 | exp motor neuron disease/ or motor neuron* disease*.mp. or lateral scleros*.mp. or motor system disease*.mp. | 52222 |
| 29 | exp multiple sclerosis/ or multiple sclerosis.mp. or disseminated sclerosis.mp. | 142570 |
| 30 | exp emphysema/ or emphysema*.mp. | 52255 |
| 31 | exp bronchitis/ or bronchit*.mp. | 69877 |
| 32 | 1 or 2 or 3 or 4 or 5 or 6 or 7 or 8 or 9 or 10 or 11 or 12 or 13 or 14 or 15 or 16 or 17 or 18 or 19 or 20 or 21 or 22 or 23 or 24 or 25 or 26 or 27 or 28 or 29 or 30 or 31 | 13854582 |
| 33 | exp mental disease/ or exp psychosis/ or ((mental* or psychiatr* or psycho*) adj (disorder* or disease* or illness*)).mp. | 2238167 |
| 34 | exp major depression/ or exp depression/ or Depress*.mp. or MDD.mp. | 832774 |
| 35 | exp anxiety disorder/ or exp anxiety/ or anxi*.mp. | 514236 |
| 36 | exp phobia/ or phobi*.mp. | 35468 |
| 37 | exp schizophrenia/ or schizophreni*.mp. or hebephreni*.mp. | 208951 |
| 38 | exp somatoform disorder/ or ((somatoform* or somati* or medically unexplained or briquet or pain) adj (disorder* or syndrome* or symptom*)).mp. or exp medically unexplained symptom/ | 82099 |
| 39 | exp dissociative disorder/ or (dissociative adj (disorder* or hysteri* or reaction*)).mp. or dissociation*.mp. | 141516 |
| 40 | exp hysteria/ or hysteri*.mp. | 7284 |
| 41 | exp mood disorder/ or ((affective* or mood*) adj (disorder* or disease* or illness* or symptom*)).mp. | 536136 |
| 42 | exp posttraumatic stress disorder/ or PTSD.mp. or ((post trauma* or posttrauma*) adj (stress* or neurose*)).mp. or combat disorder*.mp. or war disorder*.mp. | 68163 |

| | | |
|---|---|---|
| 43 | exp cognitive defect/ or ((cognitive or cognition or mental or neurocognitive) adj (dysfunction* or decline* or impairment* or deterioration* or disorder* or illness* or disease*)).mp. | 796956 |
| 44 | exp personality disorder/ or personality disorder*.mp. | 64573 |
| 45 | exp impulse control disorder/ or impulse control disorder*.mp. or intermittent explosive disorder*.mp. | 12430 |
| 46 | exp eating disorder/ or ((eating or appetite or feeding) adj disorder*).mp. | 64272 |
| 47 | exp bipolar disorder/ or ((bipolar or mani*) adj (disorder* or illness* or disease*)).mp. | 73395 |
| 48 | exp obsessive compulsive disorder/ or OCD*.mp. or ((obsess*-compulsi* or obsess* or compulsi*) adj (disorder* or illness* or disease* or neuros*)).mp. | 45783 |
| 49 | exp panic/ or (panic adj (attack* or disorder*)).mp. | 25510 |
| 50 | exp agoraphobia/ or agoraphobi*.mp. | 7090 |
| 51 | exp neurosis/ or neuros*.mp. or neurotic disorder*.mp. or psychoneuros*.mp. | 283636 |
| 52 | 33 or 34 or 35 or 36 or 37 or 38 or 39 or 40 or 41 or 42 or 43 or 44 or 45 or 46 or 47 or 48 or 49 or 50 or 51 | 3014531 |
| 53 | exp communicable disease/ or ((communic* or contag* or transmi* or infect*) adj (disease* or infection* or illness*)).mp. | 225239 |
| 54 | exp bacterial infection/ or bacteri* infection*.mp. | 871943 |
| 55 | exp conjunctivitis/ or conjunctivitis.mp. | 41971 |
| 56 | exp Human immunodeficiency virus/ or hiv.mp. or Human immuno deficiency virus.mp. | 438352 |
| 57 | exp acquired immune deficiency syndrome/ or AIDS.mp. or immunodeficiency associated virus.mp. or immun* deficiency associated virus.mp. or acquired immunodeficiency syndrome*.mp. or acquired immun* deficiency syndrome*.mp. | 244552 |
| 58 | exp Buruli ulcer/ or Bairnsdale.mp. or Buruli.mp. | 1384 |
| 59 | exp onchocerciasis/ or onchocer*.mp. | 7068 |
| 60 | hepatitis.mp. or exp hepatitis B/ or exp hepatitis C/ | 400316 |
| 61 | exp leishmaniasis/ or leishmania*.mp. | 44867 |
| 62 | exp leprosy/ or lepros*.mp. or hansen*.mp. | 32624 |
| 63 | exp elephantiasis/ or elephantias*.mp. or filaria*.mp. | 17263 |
| 64 | exp trachoma/ or egyptian ophthalmia*.mp. or trachoma*.mp. | 25279 |
| 65 | exp chikungunya/ or chickungunya.mp. or chikungunya.mp. | 8055 |
| 66 | exp taeniasis/ or taenia*.mp. | 13756 |
| 67 | exp Cysticercosis/ or cysticercos*.mp. | 5967 |

| | | |
|---|---|---|
| 68 | exp echinococcosis/ or hydatid*.mp. or echinococc*.mp. | 30248 |
| 69 | exp Chagas disease/ or trypanosom*.mp. or chagas.mp. | 47561 |
| 70 | exp trypanosomiasis/ or sleeping sickness.mp. | 25121 |
| 71 | exp Japanese encephalitis/ or (japanese adj3 encephalitis).mp. | 7011 |
| 72 | exp syphilis/ or syphilis.mp. | 32557 |
| 73 | 53 or 54 or 55 or 56 or 57 or 58 or 59 or 60 or 61 or 62 or 63 or 64 or 65 or 66 or 67 or 68 or 69 or 70 or 71 or 72 | 2031029 |
| 74 | exp tuberculosis/ | 192070 |
| 75 | Tuberculos*.mp. | 243944 |
| 76 | TB.mp. | 71895 |
| 77 | koch*.mp. | 12352 |
| 78 | exp tuberculosis/ or Tuberculos*.mp. or TB.mp. or koch*.mp. | 282378 |
| 79 | (multiple adj (ill* or disease* or condition* or syndrom* or disorder*)).mp. | 7418 |
| 80 | ((Cooccur* or co-occur* or coexist* or co-exist* or multipl* or concord* or discord* or long-term or physical*) adj3 (disease* or ill* or care or condition* or disorder* or health* or medication* or symptom* or syndrom*)).mp. | 464891 |
| 81 | (comorbid* or multimorbid* or co-occurren* or co-morbid* or Multidisease* or multi-disease*).mp. | 456607 |
| 82 | (comorbid* or multimorbid* or co-occurren* or co-morbid* or multi-morbid* or Multidisease* or multi-disease*).mp. | 457404 |
| 83 | exp Comorbidity/ or exp Multimorbidity/ or exp Multiple Chronic Conditions/ | 277616 |
| 84 | 79 or 80 or 81 or 82 or 83 | 886879 |
| 85 | exp "systematic review"/ | 263121 |
| 86 | "systematic review* ".m_titl. | 160154 |
| 87 | exp meta analysis/ | 197766 |
| 88 | "meta-analys* ".m_titl. | 142353 |
| 89 | exp "systematic review"/ or "systematic review* ".m_titl. or exp meta analysis/ or "meta-analys* ".m_titl. | 405316 |
| 90 | 32 or 52 or 73 | 16863583 |
| 91 | (32 or 52 or 73) and 78 | 236355 |
| 92 | (32 or 52 or 73) and 78 and 84 | 9136 |
| 93 | (32 or 52 or 73) and 78 and 84 and 89 | 269 |
| 94 | exp animal/ not exp human/ | 4692708 |

| | | |
|---|---|---|
| **95** | **((32 or 52 or 73) and 78 and 84) not 94** | **9038** |

#### PsycINFO (October 1st 2020)

| # ▲ | Searches | Results |
|---|---|---|
| 1 | exp non communicable disease/ or ((Non-communicable or Noncommunicable or Non-infectious) adj (disease* or condition* or illness*)).mp. | 1045 |
| 2 | exp chronic disease/ or ((chronic or long-term) adj (disease* or condition* or illness*)).mp. | 31655 |
| 3 | exp heart disease/ or (heart adj (disease* or disorder* or failure)).mp. or (cardiac adj (disease* or disorder* or failure)).mp. | 18203 |
| 4 | exp cardiovascular disease/ or (cardiovascular adj (disease* or disorder* or failure)).mp. | 17355 |
| 5 | exp coronary artery disease/ or (coronary adj (disease* or disorder* or failure)).mp. | 450 |
| 6 | exp cerebrovascular disease/ or (cerebrovascular adj (disease* or disorder* or insufficienc* or occlusion*)).mp. or (vascular adj (disease* or disorder*)).mp. or (carotid* adj (disease* or disorder*)).mp. | 6348 |
| 7 | exp peripheral occlusive artery disease/ or (arter* adj (disease* or disorder*)).mp. | 2600 |
| 8 | exp rheumatic heart disease/ or exp congenital heart malformation/ or (heart adj3 (malform* or defect* or congeni*)).mp. | 1005 |
| 9 | exp vein thrombosis/ or ((deep vein or deep venous) adj thrombos*).mp. or phlebothrombos*.mp. | 283 |
| 10 | exp lung embolism/ or (pulmonar* adj (thromboembolism* or embolism* or disease* or disorder*)).mp. | 3167 |
| 11 | exp cerebrovascular accident/ or stroke.mp. | 36654 |
| 12 | exp neoplasm/ or Cancer*.mp. or neoplas*.mp. or tumor*.mp. | 83597 |
| 13 | exp lung disease/ or exp respiratory tract disease/ or exp chronic obstructive lung disease/ or ((lung* or respiratory or pulmonar* or airflow or airway) adj2 (disease* or obstruct* or hypersensitiv*)).mp. or exp asthma/ or asthma*.mp. or exp chronic obstructive lung disease/ or exp respiratory tract allergy/ | 12588 |
| 14 | exp diabetes mellitus/ or diabet*.mp. | 33429 |
| 15 | exp autoimmune disease/ or ((autoimmun* or auto immun* or autoaggress* or auto aggress*) adj (disorder* or disease*)).mp. | 2450 |
| 16 | exp metabolic syndrome X/ or exp metabolic disorder/ or ((metabolic or insulin resistance) adj (disorder* or disease* or syndrome*)).mp. | 5832 |

| | | |
|---|---|---|
| 17 | exp obesity/ or obes*.mp. | 45008 |
| 18 | exp osteoporosis/ or osteoporo*.mp. or bone loss.mp. or exp osteolysis/ or osteolysis.mp. or bone resorption.mp. | 2582 |
| 19 | exp Parkinson disease/ or parkinson*.mp. or paralysis agitans.mp. | 36156 |
| 20 | exp arthritis/ or arthriti*.mp. or polyarthriti*.mp. or rheumarthriti*.mp. | 7166 |
| 21 | exp kidney disease/ or (kidney adj (disease* or disorder*)).mp. | 2902 |
| 22 | exp liver disease/ or (liver adj (disease* or disorder* or dysfunction*)).mp. | 2533 |
| 23 | exp hypertension/ or high blood pressure*.mp. or hypertens*.mp. | 20303 |
| 24 | exp hyperlipidemia/ or hyperlipem*.mp. or hyperlipidem*.mp. or lipem*.mp. or lipidem*.mp. | 1309 |
| 25 | exp hypercholesterolemia/ or ((high* or elevat*) adj cholesterol*).mp. or hypercholesterem*.mp. or hypercholesterolem*.mp. | 23463 |
| 26 | exp hypertriglyceridemia/ or hypertriglyceridem*.mp. | 293 |
| 27 | exp thyroid disease/ or (thyroid adj (disease* or disorder*)).mp. or exp hyperthyroidism/ or hyperthyroid*.mp. or exp hypothyroidism/ or hypothyroid*.mp. or ((thyroid-stimulating hormone* or tsh) adj deficien*).mp. | 2876 |
| 28 | exp motor neuron disease/ or motor neuron* disease*.mp. or lateral scleros*.mp. or motor system disease*.mp. | 6309 |
| 29 | exp multiple sclerosis/ or multiple sclerosis.mp. or disseminated sclerosis.mp. | 16467 |
| 30 | exp emphysema/ or emphysema*.mp. | 284 |
| 31 | exp bronchitis/ or bronchit*.mp. | 376 |
| 32 | 1 or 2 or 3 or 4 or 5 or 6 or 7 or 8 or 9 or 10 or 11 or 12 or 13 or 14 or 15 or 16 or 17 or 18 or 19 or 20 or 21 or 22 or 23 or 24 or 25 or 26 or 27 or 28 or 29 or 30 or 31 | 320242 |
| 33 | exp mental disease/ or exp psychosis/ or ((mental* or psychiatr* or psycho*) adj (disorder* or disease* or illness*)).mp. | 293988 |
| 34 | exp major depression/ or exp depression/ or Depress*.mp. or MDD.mp. | 372735 |
| 35 | exp anxiety disorder/ or exp anxiety/ or anxi*.mp. | 272212 |
| 36 | exp phobia/ or phobi*.mp. | 24164 |
| 37 | exp schizophrenia/ or schizophreni*.mp. or hebephreni*.mp. | 140086 |

| | | |
|---|---|---|
| 38 | exp somatoform disorder/ or ((somatoform* or somati* or medically unexplained or briquet or pain) adj (disorder* or syndrome* or symptom*)).mp. or exp medically unexplained symptom/ | 27266 |
| 39 | exp dissociative disorder/ or (dissociative adj (disorder* or hysteri* or reaction*)).mp. or dissociation*.mp. | 24719 |
| 40 | exp hysteria/ or hysteri*.mp. | 8248 |
| 41 | exp mood disorder/ or ((affective* or mood*) adj (disorder* or disease* or illness* or symptom*)).mp. | 170186 |
| 42 | exp posttraumatic stress disorder/ or PTSD.mp. or ((post trauma* or posttrauma*) adj (stress* or neurose*)).mp. or combat disorder*.mp. or war disorder*.mp. | 51301 |
| 43 | exp cognitive defect/ or ((cognitive or cognition or mental or neurocognitive) adj (dysfunction* or decline* or impairment* or deterioration* or disorder* or illness* or disease*)).mp. | 220755 |
| 44 | exp personality disorder/ or personality disorder*.mp. | 49496 |
| 45 | exp impulse control disorder/ or impulse control disorder*.mp. or intermittent explosive disorder*.mp. | 2226 |
| 46 | exp eating disorder/ or ((eating or appetite or feeding) adj disorder*).mp. | 38899 |
| 47 | exp bipolar disorder/ or ((bipolar or mani*) adj (disorder* or illness* or disease*)).mp. | 38226 |
| 48 | exp obsessive compulsive disorder/ or OCD*.mp. or ((obsess*-compulsi* or obsess* or compulsi*) adj (disorder* or illness* or disease* or neuros*)).mp. | 21395 |
| 49 | exp panic/ or (panic adj (attack* or disorder*)).mp. | 14398 |
| 50 | exp agoraphobia/ or agoraphobi*.mp. | 6088 |
| 51 | exp neurosis/ or neuros*.mp. or neurotic disorder*.mp. or psychoneuros*.mp. | 80933 |
| 52 | 33 or 34 or 35 or 36 or 37 or 38 or 39 or 40 or 41 or 42 or 43 or 44 or 45 or 46 or 47 or 48 or 49 or 50 or 51 | 980583 |
| 53 | exp communicable disease/ or ((communic* or contag* or transmi* or infect*) adj (disease* or infection* or illness*)).mp. | 69749 |
| 54 | exp bacterial infection/ or bacteri* infection*.mp. | 604 |
| 55 | exp conjunctivitis/ or conjunctivitis.mp. | 87 |

| | | |
|---|---|---|
| 56 | exp Human immunodeficiency virus/ or hiv.mp. or Human immuno deficiency virus.mp. | 57258 |
| 57 | exp acquired immune deficiency syndrome/ or AIDS.mp. or immunodeficiency associated virus.mp. or immun* deficiency associated virus.mp. or acquired immunodeficiency syndrome*.mp. or acquired immun* deficiency syndrome*.mp. | 48618 |
| 58 | exp Buruli ulcer/ or Bairnsdale.mp. or Buruli.mp. | 7 |
| 59 | exp onchocerciasis/ or onchocer*.mp. | 47 |
| 60 | hepatitis.mp. or exp hepatitis B/ or exp hepatitis C/ | 4725 |
| 61 | exp leishmaniasis/ or leishmania*.mp. | 61 |
| 62 | exp leprosy/ or lepros*.mp. or hansen*.mp. | 1152 |
| 63 | exp elephantiasis/ or elephantias*.mp. or filaria*.mp. | 75 |
| 64 | exp trachoma/ or egyptian ophthalmia*.mp. or trachoma*.mp. | 309 |
| 65 | exp chikungunya/ or chickungunya.mp. or chikungunya.mp. | 39 |
| 66 | exp taeniasis/ or taenia*.mp. | 160 |
| 67 | exp Cysticercosis/ or cysticercos*.mp. | 102 |
| 68 | exp echinococcosis/ or hydatid*.mp. or echinococc*.mp. | 39 |
| 69 | exp Chagas disease/ or trypanosom*.mp. or chagas.mp. | 217 |
| 70 | exp trypanosomiasis/ or sleeping sickness.mp. | 50 |
| 71 | exp Japanese encephalitis/ or (japanese adj3 encephalitis).mp. | 72 |
| 72 | exp syphilis/ or syphilis.mp. | 1866 |
| 73 | 53 or 54 or 55 or 56 or 57 or 58 or 59 or 60 or 61 or 62 or 63 or 64 or 65 or 66 or 67 or 68 or 69 or 70 or 71 or 72 | 102680 |
| 74 | exp tuberculosis/ | 1220 |
| 75 | Tuberculos*.mp. | 2791 |
| 76 | TB.mp. | 1351 |
| 77 | koch*.mp. | 1045 |
| 78 | exp tuberculosis/ or Tuberculos*.mp. or TB.mp. or koch*.mp. | 4445 |
| 79 | (multiple adj (ill* or disease* or condition* or syndrom* or disorder*)).mp. | 909 |

| | | |
|---|---|---|
| 80 | ((Cooccur* or co-occur* or coexist* or co-exist* or multipl* or concord* or discord* or long-term or physical*) adj3 (disease* or ill* or care or condition* or disorder* or health* or medication* or symptom* or syndrom*)).mp. | 115733 |
| 81 | (comorbid* or multimorbid* or co-occurren* or co-morbid* or Multidisease* or multi-disease*).mp. | 87651 |
| 82 | (comorbid* or multimorbid* or co-occurren* or co-morbid* or multi-morbid* or Multidisease* or multi-disease*).mp. | 87752 |
| 83 | exp Comorbidity/ or exp Multimorbidity/ or exp Multiple Chronic Conditions/ | 32938 |
| 84 | 79 or 80 or 81 or 82 or 83 | 191815 |
| 85 | exp "systematic review"/ | 442 |
| 86 | "systematic review* ".m_titl. | 21369 |
| 87 | exp meta analysis/ | 4803 |
| 88 | "meta-analys* ".m_titl. | 18471 |
| 89 | exp "systematic review"/ or "systematic review* ".m_titl. or exp meta analysis/ or "meta-analys* ".m_titl. | 36905 |
| 90 | 32 or 52 or 73 | 1281933 |
| 91 | (32 or 52 or 73) and 78 | 2900 |
| 92 | (32 or 52 or 73) and 78 and 84 | 354 |
| 93 | (32 or 52 or 73) and 78 and 84 and 89 | 6 |
| 94 | exp animal/ not exp human/ | 0 |
| 95 | ((32 or 52 or 73) and 78 and 84) not 94 | 354 |
| 96 | ((Non-communicable or Noncommunicable or Non-infectious) adj (disease* or condition* or illness*)).mp. | 1045 |
| 97 | exp Chronic Illness/ or ((chronic or long-term) adj (disease* or condition* or illness*)).mp. | 48780 |
| 98 | exp Heart Disorders/ or (heart adj (disease* or disorder* or failure)).mp. or (cardiac adj (disease* or disorder* or failure)).mp. | 22412 |
| 99 | exp Cardiovascular Disorders/ or (cardiovascular adj (disease* or disorder* or failure)).mp. | 68787 |
| 100 | (coronary adj (disease* or disorder* or failure)).mp. | 450 |

| | | |
|---|---|---|
| 101 | exp Cerebrovascular Disorders/ or (cerebrovascular adj (disease* or disorder* or insufficienc* or occlusion*)).mp. or (vascular adj (disease* or disorder*)).mp. or (carotid* adj (disease* or disorder*)).mp. | 31463 |
| 102 | (arter* adj (disease* or disorder*)).mp. | 2600 |
| 103 | (heart adj3 (malform* or defect* or congeni*)).mp. | 1005 |
| 104 | exp Thromboses/ or ((deep vein or deep venous) adj thrombos*).mp. or phlebothrombos*.mp. | 1069 |
| 105 | exp Embolisms/ or (pulmonar* adj (thromboembolism* or embolism* or disease* or disorder*)).mp. | 3497 |
| 106 | exp Cerebrovascular Accidents/ or stroke.mp. | 36654 |
| 107 | exp Neoplasms/ or Cancer*.mp. or neoplas*.mp. or tumor*.mp. | 83597 |
| 108 | exp Lung Disorders/ or exp Respiratory Tract Disorders/ or ((lung* or respiratory or pulmonar* or airflow or airway) adj2 (disease* or obstruct* or hypersensitiv*)).mp. or exp Asthma/ or asthma*.mp. or exp Chronic Obstructive Pulmonary Disease/ | 20385 |
| 109 | exp Diabetes Mellitus/ or diabet*.mp. | 33429 |
| 110 | ((autoimmun* or auto immun* or autoaggress* or auto aggress*) adj (disorder* or disease*)).mp. | 2450 |
| 111 | exp Metabolic Syndrome/ or ((metabolic or insulin resistance) adj (disorder* or disease* or syndrome*)).mp. | 5832 |
| 112 | exp Obesity/ or obes*.mp. | 45008 |
| 113 | exp Osteoporosis/ or osteoporo*.mp. or bone loss.mp. or osteolysis.mp. or bone resorption.mp. | 2582 |
| 114 | exp Parkinson's Disease/ or parkinson*.mp. or paralysis agitans.mp. | 36156 |
| 115 | exp Arthritis/ or arthriti*.mp. or polyarthriti*.mp. or rheumarthriti*.mp. | 7166 |
| 116 | exp Kidney Diseases/ or (kidney adj (disease* or disorder*)).mp. | 2902 |
| 117 | exp Liver Disorders/ or (liver adj (disease* or disorder* or dysfunction*)).mp. | 5355 |
| 118 | exp Hypertension/ or high blood pressure*.mp. or hypertens*.mp. | 20303 |
| 119 | (hyperlipem* or hyperlipidem* or lipem* or lipidem*).mp. | 1309 |
| 120 | (((high* or elevat*) adj cholesterol*) or hypercholesterem* or hypercholesterolem*).mp. | 1634 |

| | | |
|---|---|---|
| 121 | hypertriglyceridem*.mp. | 293 |
| 122 | exp Thyroid Disorders/ or (thyroid adj (disease* or disorder*)).mp. or exp Hyperthyroidism/ or hyperthyroid*.mp. or exp Hypothyroidism/ or hypothyroid*.mp. or ((thyroid-stimulating hormone* or tsh) adj deficien*).mp. | 2918 |
| 123 | exp Nervous System Disorders/ or motor neuron* disease*.mp. or lateral scleros*.mp. or motor system disease*.mp. | 318241 |
| 124 | exp Multiple Sclerosis/ or multiple sclerosis.mp. or disseminated sclerosis.mp. | 16467 |
| 125 | exp Pulmonary Emphysema/ or emphysema*.mp. | 284 |
| 126 | exp Bronchial Disorders/ or bronchit*.mp. | 475 |
| 127 | 96 or 97 or 98 or 99 or 100 or 101 or 102 or 103 or 104 or 105 or 106 or 107 or 108 or 109 or 110 or 111 or 112 or 113 or 114 or 115 or 116 or 117 or 118 or 119 or 120 or 121 or 122 or 123 or 124 or 125 or 126 | 565246 |
| 128 | exp Mental Disorders/ or exp Psychosis/ or ((mental* or psychiatr* or psycho*) adj (disorder* or disease* or illness*)).mp. | 918609 |
| 129 | exp Major Depression/ or exp "Depression (Emotion)"/ or Depress*.mp. or MDD.mp. | 372735 |
| 130 | exp Anxiety Disorders/ or exp Anxiety/ or anxi*.mp. | 272212 |
| 131 | exp Phobias/ or phobi*.mp. | 24164 |
| 132 | exp Schizophrenia/ or schizophreni*.mp. or hebephreni*.mp. | 140086 |
| 133 | exp Somatoform Disorders/ or ((somatoform* or somati* or medically unexplained or briquet or pain) adj (disorder* or syndrome* or symptom*)).mp. | 27266 |
| 134 | exp Dissociative Disorders/ or (dissociative adj (disorder* or hysteri* or reaction*)).mp. or dissociation*.mp. | 24719 |
| 135 | exp Hysteria/ or hysteri*.mp. | 8248 |
| 136 | exp Affective Disorders/ or ((affective* or mood*) adj (disorder* or disease* or illness* or symptom*)).mp. | 170186 |
| 137 | exp Posttraumatic Stress Disorder/ or PTSD.mp. or ((post trauma* or posttrauma*) adj (stress* or neurose*)).mp. or combat disorder*.mp. or war disorder*.mp. | 51301 |
| 138 | exp Cognitive Impairment/ or ((cognitive or cognition or mental or neurocognitive) adj (dysfunction* or decline* or impairment* or deterioration* or disorder* or illness* or disease*)).mp. | 220755 |

| | | |
|---|---|---|
| 139 | exp Personality Disorders/ or personality disorder*.mp. | 49496 |
| 140 | exp Impulse Control Disorders/ or impulse control disorder*.mp. or intermittent explosive disorder*.mp. | 2226 |
| 141 | exp Eating Disorders/ or ((eating or appetite or feeding) adj disorder*).mp. | 38899 |
| 142 | exp Bipolar Disorder/ or ((bipolar or mani*) adj (disorder* or illness* or disease*)).mp. | 38226 |
| 143 | exp Obsessive Compulsive Disorder/ or OCD*.mp. or ((obsess*-compulsi* or obsess* or compulsi*) adj (disorder* or illness* or disease* or neuros*)).mp. | 21395 |
| 144 | exp Panic Disorder/ or (panic adj (attack* or disorder*)).mp. | 13622 |
| 145 | exp Agoraphobia/ or agoraphobi*.mp. | 6088 |
| 146 | exp Neurosis/ or neuros*.mp. or neurotic disorder*.mp. or psychoneuros*.mp. | 80933 |
| 147 | 128 or 129 or 130 or 131 or 132 or 133 or 134 or 135 or 136 or 137 or 138 or 139 or 140 or 141 or 142 or 143 or 144 or 145 or 146 | 1281162 |
| 148 | exp Infectious Disorders/ or ((communic* or contag* or transmi* or infect*) adj (disease* or infection* or illness*)).mp. | 69749 |
| 149 | exp Bacterial Disorders/ or bacteri* infection*.mp. | 2726 |
| 150 | exp Eye Disorders/ or conjunctivitis.mp. | 4941 |
| 151 | exp HIV/ or hiv.mp. or Human immuno deficiency virus.mp. | 57258 |
| 152 | exp AIDS/ or AIDS.mp. or immunodeficiency associated virus.mp. or immun* deficiency associated virus.mp. or acquired immunodeficiency syndrome*.mp. or acquired immun* deficiency syndrome*.mp. | 48618 |
| 153 | (Bairnsdale or Buruli).mp. | 7 |
| 154 | onchocer*.mp. | 47 |
| 155 | hepatitis.mp. or exp Hepatitis/ | 4725 |
| 156 | leishmania*.mp. | 61 |
| 157 | (lepros* or hansen*).mp. | 1152 |
| 158 | (elephantias* or filaria*).mp. | 75 |
| 159 | (egyptian ophthalmia* or trachoma*).mp. | 309 |

| | | |
|---|---|---|
| 160 | (chickungunya or chikungunya).mp. | 39 |
| 161 | taenia*.mp. | 160 |
| 162 | cysticercos*.mp. | 102 |
| 163 | (hydatid* or echinococc*).mp. | 39 |
| 164 | (trypanosom* or chagas).mp. | 217 |
| 165 | sleeping sickness.mp. | 50 |
| 166 | (japanese adj3 encephalitis).mp. | 72 |
| 167 | exp Syphilis/ or syphilis.mp. | 1866 |
| 168 | 148 or 149 or 150 or 151 or 152 or 153 or 154 or 155 or 156 or 157 or 158 or 159 or 160 or 161 or 162 or 163 or 164 or 165 or 166 or 167 | 107431 |
| 169 | exp Tuberculosis/ | 1220 |
| 170 | Tuberculos*.mp. | 2791 |
| 171 | TB.mp. | 1351 |
| 172 | koch*.mp. | 1045 |
| 173 | exp Tuberculosis/ or Tuberculos*.mp. or TB.mp. or koch*.mp. | 4445 |
| 174 | (multiple adj (ill* or disease* or condition* or syndrom* or disorder*)).mp. | 909 |
| 175 | ((Cooccur* or co-occur* or coexist* or co-exist* or multipl* or concord* or discord* or long-term or physical*) adj3 (disease* or ill* or care or condition* or disorder* or health* or medication* or symptom* or syndrom*)).mp. | 115733 |
| 176 | (comorbid* or multimorbid* or co-occurren* or co-morbid* or multi-morbid* or Multidisease* or multi-disease*).mp. | 87752 |
| 177 | exp Comorbidity/ | 32938 |
| 178 | 174 or 175 or 176 or 177 | 191815 |
| 179 | exp "Systematic Review"/ | 442 |
| 180 | "systematic review*".m_titl. | 21369 |
| 181 | exp Meta Analysis/ | 4803 |
| 182 | "meta-analys*".m_titl. | 18471 |
| 183 | exp "Systematic Review"/ or "systematic review*".m_titl. or exp Meta Analysis/ or "meta-analys*".m_titl. | 36905 |

| | | |
|---|---|---|
| 184 | 127 or 147 or 168 | 1660437 |
| 185 | (127 or 147 or 168) and 173 | 3041 |
| **186** | **(127 or 147 or 168) and 173 and 178** | **361** |

Web of Science (October 1st 2020)

| Set | Results | Save History / Create AlertOpen Saved History |
|---|---|---|
| # 88 | 3,306 | (#74 OR #52 OR #32 ) AND #82 AND #78<br>*Indexes=SCI-EXPANDED, SSCI, A&HCI, CPCI-S, CPCI-SSH, ESCI Timespan=All years* |
| # 87 | 87,389 | (#74 OR #52 OR #32 ) AND #82<br>*Indexes=SCI-EXPANDED, SSCI, A&HCI, CPCI-S, CPCI-SSH, ESCI Timespan=All years* |
| # 86 | 11,036,526 | #74 OR #52 OR #32<br>*Indexes=SCI-EXPANDED, SSCI, A&HCI, CPCI-S, CPCI-SSH, ESCI Timespan=All years* |
| # 85 | 224,241 | #84 OR #83<br>*Indexes=SCI-EXPANDED, SSCI, A&HCI, CPCI-S, CPCI-SSH, ESCI Timespan=All years* |
| # 84 | 133,549 | TI="Meta-analysis"<br>*Indexes=SCI-EXPANDED, SSCI, A&HCI, CPCI-S, CPCI-SSH, ESCI Timespan=All years* |
| # 83 | 145,562 | TI = "Systematic Review"<br>*Indexes=SCI-EXPANDED, SSCI, A&HCI, CPCI-S, CPCI-SSH, ESCI Timespan=All years* |
| # 82 | 233,516 | #81 OR #80 OR #79<br>*Indexes=SCI-EXPANDED, SSCI, A&HCI, CPCI-S, CPCI-SSH, ESCI Timespan=All years* |
| # 81 | 19,258 | TS=koch*<br>*Indexes=SCI-EXPANDED, SSCI, A&HCI, CPCI-S, CPCI-SSH, ESCI Timespan=All years* |
| # 80 | 80,335 | TS=TB<br>*Indexes=SCI-EXPANDED, SSCI, A&HCI, CPCI-S, CPCI-SSH, ESCI Timespan=All years* |
| # 79 | 169,896 | TS=Tuberculos*<br>*Indexes=SCI-EXPANDED, SSCI, A&HCI, CPCI-S, CPCI-SSH, ESCI Timespan=All years* |

| | | |
|---|---|---|
| # 78 | 654,553 | #77 OR #76 OR #75<br><br>*Indexes=SCI-EXPANDED, SSCI, A&HCI, CPCI-S, CPCI-SSH, ESCI Timespan=All years* |
| # 77 | 236,371 | TS=(comorbid* or multimorbid* or co-occurren* or co-morbid* or multi-morbid* or Multidisease* or multi-disease*)<br><br>*Indexes=SCI-EXPANDED, SSCI, A&HCI, CPCI-S, CPCI-SSH, ESCI Timespan=All years* |
| # 76 | 323,507 | TS=((Cooccur* or co-occur* or coexist* or co-exist* or multipl* or concord* or discord* or long-term or physical*) NEAR/3 (disease* or ill* or care or condition* or disorder* or health* or medication* or symptom* or syndrom*) )<br><br>*Indexes=SCI-EXPANDED, SSCI, A&HCI, CPCI-S, CPCI-SSH, ESCI Timespan=All years* |
| # 75 | 190,356 | TS=(multiple NEAR (ill* or disease* or condition* or syndrom* or disorder*) )<br><br>*Indexes=SCI-EXPANDED, SSCI, A&HCI, CPCI-S, CPCI-SSH, ESCI Timespan=All years* |
| # 74 | 2,665,421 | #73 OR #72 OR #71 OR #70 OR #69 OR #68 OR #67 OR #66 OR #65 OR #64 OR #63 OR #62 OR #61 OR #60 OR #59 OR #58 OR #57 OR #56 OR #55 OR #54 OR #53<br><br>*Indexes=SCI-EXPANDED, SSCI, A&HCI, CPCI-S, CPCI-SSH, ESCI Timespan=All years* |
| # 73 | 23,326 | TS=syphilis<br><br>*Indexes=SCI-EXPANDED, SSCI, A&HCI, CPCI-S, CPCI-SSH, ESCI Timespan=All years* |
| # 72 | 5,967 | TS=(japanese NEAR/3 encephalitis)<br><br>*Indexes=SCI-EXPANDED, SSCI, A&HCI, CPCI-S, CPCI-SSH, ESCI Timespan=All years* |
| # 71 | 4,247 | TS=sleeping sickness<br><br>*Indexes=SCI-EXPANDED, SSCI, A&HCI, CPCI-S, CPCI-SSH, ESCI Timespan=All years* |
| # 70 | 49,886 | TS=trypanosom* or TS=chagas<br><br>*Indexes=SCI-EXPANDED, SSCI, A&HCI, CPCI-S, CPCI-SSH, ESCI Timespan=All years* |
| # 69 | 19,739 | TS=hydatid* OR TS=echinococc* |

| | | |
|---|---|---|
| | | *Indexes=SCI-EXPANDED, SSCI, A&HCI, CPCI-S, CPCI-SSH, ESCI Timespan=All years* |
| # 68 | 4,647 | TS=cysticercos* |
| | | *Indexes=SCI-EXPANDED, SSCI, A&HCI, CPCI-S, CPCI-SSH, ESCI Timespan=All years* |
| # 67 | 8,237 | TS=taenia* |
| | | *Indexes=SCI-EXPANDED, SSCI, A&HCI, CPCI-S, CPCI-SSH, ESCI Timespan=All years* |
| # 66 | 6,578 | TS=chickungunya Or TS=chikungunya |
| | | *Indexes=SCI-EXPANDED, SSCI, A&HCI, CPCI-S, CPCI-SSH, ESCI Timespan=All years* |
| # 65 | 19,944 | TS=trachoma* |
| | | *Indexes=SCI-EXPANDED, SSCI, A&HCI, CPCI-S, CPCI-SSH, ESCI Timespan=All years* |
| # 64 | 5 | TS=egyptian ophthalmia* |
| | | *Indexes=SCI-EXPANDED, SSCI, A&HCI, CPCI-S, CPCI-SSH, ESCI Timespan=All years* |
| # 63 | 11,234 | TS=elephantias* OR TS=filaria* |
| | | *Indexes=SCI-EXPANDED, SSCI, A&HCI, CPCI-S, CPCI-SSH, ESCI Timespan=All years* |
| # 62 | 30,013 | TS=lepros* OR TS=hansen* |
| | | *Indexes=SCI-EXPANDED, SSCI, A&HCI, CPCI-S, CPCI-SSH, ESCI Timespan=All years* |
| # 61 | 41,697 | TS=leishmania* |
| | | *Indexes=SCI-EXPANDED, SSCI, A&HCI, CPCI-S, CPCI-SSH, ESCI Timespan=All years* |
| # 60 | 267,219 | TS=hepatitis |
| | | *Indexes=SCI-EXPANDED, SSCI, A&HCI, CPCI-S, CPCI-SSH, ESCI Timespan=All years* |
| # 59 | 6,450 | TS=onchocer* |
| | | *Indexes=SCI-EXPANDED, SSCI, A&HCI, CPCI-S, CPCI-SSH, ESCI Timespan=All years* |
| # 58 | 1,117 | TS=Bairnsdale OR TS=Buruli |

| | | |
|---|---|---|
| | | *Indexes=SCI-EXPANDED, SSCI, A&HCI, CPCI-S, CPCI-SSH, ESCI Timespan=All years* |
| # 57 | 637,613 | TS=AIDS OR TS=immunodeficiency associated virus OR TS=immun* deficiency associated virus OR TS=acquired immunodeficiency syndrome* OR TS=acquired immun* deficiency syndrome*<br>*Indexes=SCI-EXPANDED, SSCI, A&HCI, CPCI-S, CPCI-SSH, ESCI Timespan=All years* |
| # 56 | 373,281 | TS=hiv OR TS=Human immuno deficiency virus<br>*Indexes=SCI-EXPANDED, SSCI, A&HCI, CPCI-S, CPCI-SSH, ESCI Timespan=All years* |
| # 55 | 10,487 | TS=conjunctivitis<br>*Indexes=SCI-EXPANDED, SSCI, A&HCI, CPCI-S, CPCI-SSH, ESCI Timespan=All years* |
| # 54 | 239,298 | TS=bacteri* infection*<br>*Indexes=SCI-EXPANDED, SSCI, A&HCI, CPCI-S, CPCI-SSH, ESCI Timespan=All years* |
| # 53 | 1,701,693 | TS=((communic* or contag* or transmi* or infect*) NEAR (disease* or infection* or illness*) )<br>*Indexes=SCI-EXPANDED, SSCI, A&HCI, CPCI-S, CPCI-SSH, ESCI Timespan=All years* |
| # 52 | 1,882,635 | #51 OR #50 OR #49 OR #48 OR #47 OR #46 OR #45 OR #44 OR #43 OR #42 OR #41 OR #40 OR #39 OR #38 OR #37 OR #36 OR #35 OR #34 OR #33<br>*Indexes=SCI-EXPANDED, SSCI, A&HCI, CPCI-S, CPCI-SSH, ESCI Timespan=All years* |
| # 51 | 155,150 | TS=neuros* OR TS=neurotic disorder* OR TS=psychoneuros*<br>*Indexes=SCI-EXPANDED, SSCI, A&HCI, CPCI-S, CPCI-SSH, ESCI Timespan=All years* |
| # 50 | 5,159 | TS=agoraphobi*<br>*Indexes=SCI-EXPANDED, SSCI, A&HCI, CPCI-S, CPCI-SSH, ESCI Timespan=All years* |
| # 49 | 19,457 | TS=(panic NEAR (attack* or disorder*) )<br>*Indexes=SCI-EXPANDED, SSCI, A&HCI, CPCI-S, CPCI-SSH, ESCI Timespan=All years* |

### 48 32,462 TS=OCD* OR TS=((obsess*-compulsi* or obsess* or compulsi*) NEAR (disorder* or illness* or disease* or neuros*) )

*Indexes=SCI-EXPANDED, SSCI, A&HCI, CPCI-S, CPCI-SSH, ESCI Timespan=All years*

### 47 136,340 TS=((bipolar or mani*) NEAR (disorder* or illness* or disease*) )

*Indexes=SCI-EXPANDED, SSCI, A&HCI, CPCI-S, CPCI-SSH, ESCI Timespan=All years*

### 46 35,398 TS=((eating or appetite or feeding) NEAR disorder*)

*Indexes=SCI-EXPANDED, SSCI, A&HCI, CPCI-S, CPCI-SSH, ESCI Timespan=All years*

### 45 4,494 TS=impulse control disorder* OR TS=intermittent explosive disorder*

*Indexes=SCI-EXPANDED, SSCI, A&HCI, CPCI-S, CPCI-SSH, ESCI Timespan=All years*

### 44 52,209 TS=personality disorder*

*Indexes=SCI-EXPANDED, SSCI, A&HCI, CPCI-S, CPCI-SSH, ESCI Timespan=All years*

### 43 322,947 TS=((cognitive or cognition or mental or neurocognitive) NEAR (dysfunction* or decline* or impairment* or deterioration* or disorder* or illness* or disease* ) )

*Indexes=SCI-EXPANDED, SSCI, A&HCI, CPCI-S, CPCI-SSH, ESCI Timespan=All years*

### 42 68,594 TS=PTSD OR TS=(((post NEAR trauma*) or posttrauma*) NEAR (stress* or neurose*) ) OR TS=combat disorder* OR TS=war disorder*

*Indexes=SCI-EXPANDED, SSCI, A&HCI, CPCI-S, CPCI-SSH, ESCI Timespan=All years*

### 41 73,023 TS=((affective* or mood*) NEAR (disorder* or disease* or illness* or symptom*) )

*Indexes=SCI-EXPANDED, SSCI, A&HCI, CPCI-S, CPCI-SSH, ESCI Timespan=All years*

### 40 6,459 TS=hysteri*

*Indexes=SCI-EXPANDED, SSCI, A&HCI, CPCI-S, CPCI-SSH, ESCI Timespan=All years*

### 39 215,343 TS=(dissociative NEAR (disorder* or hysteri* or reaction*) ) OR TS=dissociation*

| | | |
|---|---|---|
| | | *Indexes=SCI-EXPANDED, SSCI, A&HCI, CPCI-S, CPCI-SSH, ESCI Timespan=All years* |
| # 38 | 121,351 | TS=((somatoform* or somati* or (medically NEAR unexplained) or briquet or pain) NEAR (disorder* or syndrome* or symptom*) ) <br> *Indexes=SCI-EXPANDED, SSCI, A&HCI, CPCI-S, CPCI-SSH, ESCI Timespan=All years* |
| # 37 | 199,661 | TS=schizophreni* OR TS=hebephreni* <br> *Indexes=SCI-EXPANDED, SSCI, A&HCI, CPCI-S, CPCI-SSH, ESCI Timespan=All years* |
| # 36 | 19,308 | TS=phobi* <br> *Indexes=SCI-EXPANDED, SSCI, A&HCI, CPCI-S, CPCI-SSH, ESCI Timespan=All years* |
| # 35 | 298,695 | TS=anxi* <br> *Indexes=SCI-EXPANDED, SSCI, A&HCI, CPCI-S, CPCI-SSH, ESCI Timespan=All years* |
| # 34 | 642,370 | TS=Depress* OR TS=MDD <br> *Indexes=SCI-EXPANDED, SSCI, A&HCI, CPCI-S, CPCI-SSH, ESCI Timespan=All years* |
| # 33 | 281,778 | TS=((mental* or psychiatr* or psycho*) NEAR (disorder* or disease* or illness *) ) <br> *Indexes=SCI-EXPANDED, SSCI, A&HCI, CPCI-S, CPCI-SSH, ESCI Timespan=All years* |
| # 32 | 7,445,095 | #31 OR #30 OR #29 OR #28 OR #27 OR #26 OR #25 OR #24 OR #23 OR #22 OR #21 OR #20 OR #19 OR #18 OR #17 OR #16 OR #15 OR #14 OR #13 OR #12 OR #11 OR #10 OR #9 OR #8 OR #7 OR #6 OR #5 OR #4 OR #3 OR #2 OR #1 <br> *Indexes=SCI-EXPANDED, SSCI, A&HCI, CPCI-S, CPCI-SSH, ESCI Timespan=All years* |
| # 31 | 20,508 | TS=bronchit* <br> *Indexes=SCI-EXPANDED, SSCI, A&HCI, CPCI-S, CPCI-SSH, ESCI Timespan=All years* |
| # 30 | 26,576 | TS=emphysema* <br> *Indexes=SCI-EXPANDED, SSCI, A&HCI, CPCI-S, CPCI-SSH, ESCI Timespan=All years* |
| # 29 | 128,730 | TS=multiple sclerosis OR TS=disseminated sclerosis |

*Indexes=SCI-EXPANDED, SSCI, A&HCI, CPCI-S, CPCI-SSH, ESCI Timespan=All years*

| | | |
|---|---|---|
| # 28 | 67,597 | TS=motor neuron* disease* OR TS=lateral scleros* OR TS=motor system disease*<br>*Indexes=SCI-EXPANDED, SSCI, A&HCI, CPCI-S, CPCI-SSH, ESCI Timespan=All years* |
| # 27 | 70,867 | TS=(thyroid NEAR (disease* or disorder*) ) OR TS=hyperthyroid* OR TS=hypothyroid* OR TS=(((thyroid-stimulating NEAR hormone*) or tsh) NEAR deficien*)<br>*Indexes=SCI-EXPANDED, SSCI, A&HCI, CPCI-S, CPCI-SSH, ESCI Timespan=All years* |
| # 26 | 14,058 | TS=hypertriglyceridem*<br>*Indexes=SCI-EXPANDED, SSCI, A&HCI, CPCI-S, CPCI-SSH, ESCI Timespan=All years* |
| # 25 | 113,344 | TS=((high* or elevat*) NEAR cholesterol*) Or TS=hypercholesterem* OR TS=hypercholesterolem*<br>*Indexes=SCI-EXPANDED, SSCI, A&HCI, CPCI-S, CPCI-SSH, ESCI Timespan=All years* |
| # 24 | 36,865 | TS=hyperlipem* OR TS=hyperlipidem* or TS=lipem* OR TS=lipidem*<br>*Indexes=SCI-EXPANDED, SSCI, A&HCI, CPCI-S, CPCI-SSH, ESCI Timespan=All years* |
| # 23 | 611,082 | TS=high blood pressure* OR TS=hypertens*<br>*Indexes=SCI-EXPANDED, SSCI, A&HCI, CPCI-S, CPCI-SSH, ESCI Timespan=All years* |
| # 22 | 160,331 | TS=(liver NEAR (disease* or disorder* or dysfunction*) )<br>*Indexes=SCI-EXPANDED, SSCI, A&HCI, CPCI-S, CPCI-SSH, ESCI Timespan=All years* |
| # 21 | 124,636 | TS=(kidney NEAR (disease* or disorder*) )<br>*Indexes=SCI-EXPANDED, SSCI, A&HCI, CPCI-S, CPCI-SSH, ESCI Timespan=All years* |
| # 20 | 278,113 | TS=arthriti* OR TS=polyarthriti* OR TS=rheumarthriti*<br>*Indexes=SCI-EXPANDED, SSCI, A&HCI, CPCI-S, CPCI-SSH, ESCI Timespan=All years* |
| # 19 | 179,614 | TS=parkinson* OR TS=paralysis agitans |

| | | |
|---|---|---|
| | | *Indexes=SCI-EXPANDED, SSCI, A&HCI, CPCI-S, CPCI-SSH, ESCI Timespan=All years* |
| # 18 | 194,341 | TS=osteoporo* OR TS=bone loss OR TS=osteolysis or TS=bone resorption<br>*Indexes=SCI-EXPANDED, SSCI, A&HCI, CPCI-S, CPCI-SSH, ESCI Timespan=All years* |
| # 17 | 424,614 | TS=obes*<br>*Indexes=SCI-EXPANDED, SSCI, A&HCI, CPCI-S, CPCI-SSH, ESCI Timespan=All years* |
| # 16 | 173,206 | TS=((metabolic or (insulin near resistance) ) NEAR (disorder* or disease* or syndrome*) )<br>*Indexes=SCI-EXPANDED, SSCI, A&HCI, CPCI-S, CPCI-SSH, ESCI Timespan=All years* |
| # 15 | 108,636 | TS=((autoimmun* or (auto NEAR immun*) or autoaggress* or (auto NEAR aggress*) ) NEAR (disorder* or disease*) )<br>*Indexes=SCI-EXPANDED, SSCI, A&HCI, CPCI-S, CPCI-SSH, ESCI Timespan=All years* |
| # 14 | 768,073 | TS=diabet*<br>*Indexes=SCI-EXPANDED, SSCI, A&HCI, CPCI-S, CPCI-SSH, ESCI Timespan=All years* |
| # 13 | 383,033 | TS=((lung* or respiratory or pulmonar* or airflow or airway) NEAR/2 (disease* or obstruct* or hypersensitiv*) ) OR TS=asthma*<br>*Indexes=SCI-EXPANDED, SSCI, A&HCI, CPCI-S, CPCI-SSH, ESCI Timespan=All years* |
| # 12 | 3,504,460 | TS=Cancer* or TS=neoplas* OR TS=tumor*<br>*Indexes=SCI-EXPANDED, SSCI, A&HCI, CPCI-S, CPCI-SSH, ESCI Timespan=All years* |
| # 11 | 354,992 | TS=stroke<br>*Indexes=SCI-EXPANDED, SSCI, A&HCI, CPCI-S, CPCI-SSH, ESCI Timespan=All years* |
| # 10 | 163,389 | TS=(pulmonar* NEAR (thromboembolism* or embolism* or disease* or disorder*) )<br>*Indexes=SCI-EXPANDED, SSCI, A&HCI, CPCI-S, CPCI-SSH, ESCI Timespan=All years* |
| # 9 | 34,608 | TS=(("deep vein" or "deep venous") NEAR thrombos*) OR TS=phlebothrombos* |

| | | |
|---|---|---|
| | | *Indexes=SCI-EXPANDED, SSCI, A&HCI, CPCI-S, CPCI-SSH, ESCI Timespan=All years* |
| # 8 | 52,703 | TS=(heart NEAR/3 (malform* or defect* or congeni*) )<br>*Indexes=SCI-EXPANDED, SSCI, A&HCI, CPCI-S, CPCI-SSH, ESCI Timespan=All years* |
| # 7 | 223,227 | TS=(arter* NEAR (disease* or disorder*) )<br>*Indexes=SCI-EXPANDED, SSCI, A&HCI, CPCI-S, CPCI-SSH, ESCI Timespan=All years* |
| # 6 | 131,426 | TS=(cerebrovascular NEAR (disease* or disorder* or insufficienc* or occlusion*) ) OR TS=(vascular NEAR (disease* or disorder*) ) OR TS=(carotid* NEAR (disease* or disorder*) )<br>*Indexes=SCI-EXPANDED, SSCI, A&HCI, CPCI-S, CPCI-SSH, ESCI Timespan=All years* |
| # 5 | 259,881 | TS=(coronary NEAR (disease* or disorder* or failure) )<br>*Indexes=SCI-EXPANDED, SSCI, A&HCI, CPCI-S, CPCI-SSH, ESCI Timespan=All years* |
| # 4 | 284,840 | TS=(cardiovascular NEAR (disease* or disorder* or failure) )<br>*Indexes=SCI-EXPANDED, SSCI, A&HCI, CPCI-S, CPCI-SSH, ESCI Timespan=All years* |
| # 3 | 559,562 | TS=(heart NEAR (disease* or disorder* or failure) ) OR TS=(cardiac NEAR (disease* or disorder* or failure) )<br>*Indexes=SCI-EXPANDED, SSCI, A&HCI, CPCI-S, CPCI-SSH, ESCI Timespan=All years* |
| # 2 | 497,564 | TS=((chronic or long-term) NEAR (disease* or condition* or illness*) )<br>*Indexes=SCI-EXPANDED, SSCI, A&HCI, CPCI-S, CPCI-SSH, ESCI Timespan=All years* |
| # 1 | 12,281 | TS=((Non-communicable or Noncommunicable or Non-infectious) NEAR/1 (disease* or condition* or illness*) )<br>*Indexes=SCI-EXPANDED, SSCI, A&HCI, CPCI-S, CPCI-SSH, ESCI Timespan=All years* |

#### Global Index Medicus (October 1st 2020)

tw:(((("Noncommunicable Diseases" OR ((("Non-communicable" OR noncommunicable OR "Non-infectious") AND (disease* OR condition* OR illness*)))) OR ("Chronic Disease" OR (((chronic OR "long-term") AND (disease* OR condition* OR illness*)))) OR ("Heart Diseases" OR ((heart AND (disease* OR disorder* OR failure)) OR (cardiac AND (disease* OR disorder* OR failure)))) OR ("Cardiovascular Diseases" OR ((cardiovascular AND (disease* OR disorder* OR failure)))) OR ("Coronary Disease" OR (coronary AND (disease* OR disorder* OR failure))) OR ("Cerebrovascular Disorders" OR ((cerebrovascular AND (disease* OR disorder* OR insufficienc* OR occlusion*)) OR (vascular AND (disease* OR disorder*)) OR (carotid* AND (disease* OR disorder*)))) OR ("Peripheral Arterial Disease" OR ( (arter* AND (disease* OR disorder*)))) OR ("Rheumatic Heart Disease" OR "Heart Defects, Congenital" OR ((heart AND (malform* OR defect* OR congeni*)))) OR ("Venous Thrombosis" OR ( (("deep vein" OR "deep venous") AND thrombos*) OR phlebothrombos*)) OR ("Pulmonary Embolism" OR ((pulmonar* AND (thromboembolism* OR embolism* OR disease* OR disorder*)))) OR ("Stroke" OR (stroke)) OR ("Neoplasms" OR (cancer* OR neoplas* OR tumor*)) OR ("Lung Diseases" OR "Respiratory Tract Diseases" OR "Lung Diseases, Obstructive" OR (((lung* OR respiratory OR pulmonar* OR airflow OR airway) AND (disease* OR obstruct* OR hypersensitiv*))) OR "Asthma" OR asthma* OR "Pulmonary Disease, Chronic Obstructive" OR "Respiratory Hypersensitivity") OR ("Diabetes Mellitus" OR diabet*) OR ("Autoimmune Diseases" OR ((autoimmun* OR (auto AND immun*) OR autoaggress* OR (auto AND aggress*)) AND (disorder* OR disease*))) OR ("Metabolic Syndrome" OR "Metabolic Diseases" OR (((metabolic OR "insulin resistance") AND (disorder* OR disease* OR syndrome*)))) OR ("Obesity" OR obes*) OR ("Osteoporosis" OR (osteoporo* OR "bone loss" OR "Osteolysis" OR osteolysis OR "bone resorption")) OR ("Parkinson Disease" OR (parkinson* OR "paralysis agitans")) OR ("Arthritis" OR (arthriti* OR polyarthriti* OR rheumarthriti*)) OR ("Kidney Diseases" OR ((kidney AND (disease* OR disorder*)))) OR ("Liver Diseases" OR (liver AND (disease* OR disorder* OR dysfunction*))) OR ("Hypertension" OR ("high blood pressure" OR hypertens*)) OR ("Hyperlipidemias" OR (hyperlipem* OR hyperlipidem* OR lipem* OR lipidem*)) OR ("Hypercholesterolemia" OR (((high* OR elevat*) AND cholesterol*) OR hypercholesterem* OR hypercholesterolem*)) OR ("Hypertriglyceridemia " OR (hypertriglyceridem*)) OR ("Thyroid Diseases" OR (thyroid AND (disease* OR disorder*)) OR "Hyperthyroidism " OR (hyperthyroid*) OR "Hypothyroidism" OR (hypothyroid*) OR ( (("thyroid-stimulating hormone" OR tsh) AND deficien*))) OR ("Motor Neuron Disease" OR ("motor neuron disease") OR (lateral AND scleros*) OR ("motor system disease")) OR ("Multiple Sclerosis" OR ("multiple sclerosis" OR "disseminated sclerosis")) OR ("Emphysema" OR emphysema*) OR ("Bronchitis" OR  bronchit*)) OR (("Mental Disorders" OR "Psychotic Disorders" OR ( ((mental* OR psychiatr* OR psycho*) AND (disorder* OR disease* OR illness*)))) OR ("Depressive Disorder, Major" OR "Depression" OR (depress* OR mdd)) OR ("Anxiety Disorders" OR "Anxiety" OR anxi*) OR ("Phobic Disorders" OR phobi*) OR ("Schizophrenia" OR (schizophreni* OR hebephreni*)) OR ("Somatoform Disorders" OR "Medically Unexplained Symptoms" OR ( ((somatoform* OR somati* OR (medically AND unexplained) OR briquet OR pain) AND (disorder* OR syndrome* OR symptom*)))) OR ("Dissociative Disorders" OR  (dissociative AND (disorder* OR hysteri* OR reaction*)) OR dissociation*) OR ("Hysteria" OR hysteri*) OR ("Mood Disorders" OR ((affective* OR mood*) AND (disorder* OR disease* OR illness* OR symptom*))) OR ("Stress Disorders, Post-Traumatic" OR (ptsd OR ((posttrauma* OR trauma*) AND (stress* OR neurose*)) OR "combat disorder" OR "war disorder")) OR ("Cognition Disorders" OR ( ((cognitive OR cognition OR mental OR neurocognitive) AND (dysfunction* OR decline* OR impairment* OR deterioration* OR disorder* OR illness* OR disease*)))) OR ("Personality Disorders" OR "personality disorder") OR ("Disruptive, Impulse Control, and Conduct Disorders" OR ("impulse control disorder" OR "intermittent

| |
|---|
| explosive disorder")) OR ("Feeding and Eating Disorders" OR ((eating OR appetite OR feeding) AND disorder*)) OR ("Bipolar Disorder" OR ((bipolar OR mani*) AND (disorder* OR illness* OR disease*))) OR ("Obsessive-Compulsive Disorder" OR ocd* OR ((obsess* OR compulsi*) AND (disorder* OR illness* OR disease* OR neuros*))) OR ("Panic Disorder" OR ((panic AND (attack* OR disorder*)))) OR ("Agoraphobia" OR agoraphobi*) OR ("Neurotic Disorders" OR (neuros* OR "neurotic disorder" OR psychoneuros*))) OR (("Communicable Diseases" OR (((communic* OR contag* OR transmi* OR infect*) AND (disease* OR infection* OR illness*)))) OR ("Bacterial Infections" OR ( bacteri* infection*)) OR ("Conjunctivitis") OR ("HIV" OR "Human immuno deficiency virus") OR ("Acquired Immunodeficiency Syndrome" OR (aids OR "immunodeficiency associated virus" OR (immun* deficiency associated virus) OR (acquired immunodeficiency syndrome*) OR (acquired immun* deficiency syndrome*))) OR ("Buruli Ulcer" OR ( bairnsdale OR buruli)) OR ("Onchocerciasis" OR (onchocer*)) OR ("Hepatitis B" OR "Hepatitis C" OR (hepatitis)) OR ("Leishmaniasis" OR (leishmania*)) OR ("Leprosy" OR (lepros* OR hansen*)) OR ("Elephantiasis, Filarial" OR (elephantias* OR filaria*)) OR ("Trachoma" OR ((egyptian ophthalmia*) OR trachoma*)) OR ("Chikungunya Fever" OR (chickungunya OR chikungunya)) OR ("Taeniasis" OR taenia*) OR ("Cysticercosis" OR cysticercos*) OR ("Echinococcosis" OR (hydatid OR echinococc*)) OR ("Chagas Disease" OR (trypanosom* OR chagas)) OR ("Trypanosomiasis" OR ("sleeping sickness")) OR ("Encephalitis, Japanese" OR (japanese encephalitis)) OR ("Syphilis"))) AND ("Tuberculosis" OR tuberculos* OR tb OR koch*) AND ((multiple AND (ill* OR disease* OR condition* OR syndrom* OR disorder*)) OR (((cooccur* OR co-occur* OR coexist* OR co-exist* OR multipl* OR concord* OR discord* OR long-term OR physical*) AND (disease* OR ill* OR care OR condition* OR disorder* OR health* OR medication* OR symptom* OR syndrom*))) OR (comorbid* OR multimorbid* OR co-occurren* OR co-morbid* OR multi-morbid* OR multidisease* OR multi-disease*) OR ("Comorbidity" OR "Multimorbidity" OR "Multiple Chronic Conditions"))) |
| 5131 |

OpenGrey

((("Noncommunicable Diseases" OR ((("Non-communicable" or Noncommunicable or "Non-infectious") AND (disease* or condition* or illness*)))) OR ("Chronic Disease" OR (((chronic or "long-term") AND (disease* or condition* or illness*)))) OR ("Heart Diseases" OR ((heart AND (disease* or disorder* or failure)) or (cardiac AND (disease* or disorder* or failure)))) OR ("Cardiovascular Diseases" OR ((cardiovascular AND (disease* or disorder* or failure)))) OR ("Coronary Disease" OR (coronary AND (disease* or disorder* or failure))) OR ("Cerebrovascular Disorders" OR ((cerebrovascular AND (disease* or disorder* or insufficienc* or occlusion*)) or (vascular AND (disease* or disorder*)) or (carotid* AND (disease* or disorder*)))) OR ("Peripheral Arterial Disease" OR ( (arter* AND (disease* or disorder*)))) OR ("Rheumatic Heart Disease" OR "Heart Defects, Congenital" OR ((heart AND (malform* or defect* or congeni*)))) OR ("Venous Thrombosis" OR ( (("deep vein" or "deep venous") AND thrombos*) or phlebothrombos*)) OR ("Pulmonary Embolism" OR ((pulmonar* AND (thromboembolism* or embolism* or disease* or disorder*)))) OR ("Stroke" OR (stroke)) OR ("Neoplasms" OR (Cancer* or neoplas* or tumor*)) OR ("Lung Diseases" OR "Respiratory Tract Diseases" OR "Lung Diseases, Obstructive" OR (((lung* or respiratory or pulmonar* or airflow or airway) AND (disease* or obstruct* or hypersensitiv*))) OR "Asthma" OR asthma* OR "Pulmonary Disease, Chronic Obstructive" OR "Respiratory Hypersensitivity") OR ("Diabetes Mellitus" or diabet*) OR ("Autoimmune Diseases" OR ((autoimmun* or (auto AND immun*) or autoaggress* or (auto AND aggress*)) AND (disorder* or disease*))) OR ("Metabolic Syndrome" OR "Metabolic Diseases" OR (((metabolic or "insulin resistance") AND (disorder* or disease* or syndrome*)))) OR ("Obesity" OR obes*) OR ("Osteoporosis" OR (osteoporo* or "bone loss" or "Osteolysis" or osteolysis or "bone resorption")) OR ("Parkinson Disease" OR (parkinson* or "paralysis agitans")) OR ("Arthritis" OR (arthriti* or polyarthriti* or rheumarthriti*)) OR ("Kidney Diseases" OR ((kidney AND (disease* or disorder*)))) OR ("Liver Diseases" OR (liver AND (disease* or disorder* or dysfunction*))) OR ("Hypertension" OR ("high blood pressure" or hypertens*)) OR ("Hyperlipidemias" OR (hyperlipem* or hyperlipidem* or lipem* or lipidem*)) OR ("Hypercholesterolemia" OR (((high* or elevat*) AND cholesterol*) or hypercholesterem* or hypercholesterolem*)) OR ("Hypertriglyceridemia " OR (hypertriglyceridem*)) OR ("Thyroid Diseases" OR (thyroid AND (disease* or disorder*)) OR "Hyperthyroidism " OR (hyperthyroid*) OR "Hypothyroidism" OR (hypothyroid*) OR ( (("thyroid-stimulating hormone" or tsh) AND deficien*))) OR ("Motor Neuron Disease" OR ("motor neuron disease") OR (lateral AND scleros*) OR ("motor system disease")) OR ("Multiple Sclerosis" OR ("multiple sclerosis" or "disseminated sclerosis")) OR ("Emphysema" OR emphysema*) OR ("Bronchitis" OR  bronchit*)) OR (("Mental Disorders" OR "Psychotic Disorders" OR ( ((mental* or psychiatr* or psycho*) AND (disorder* or disease* or illness*)))) OR ("Depressive Disorder, Major" OR "Depression" OR (Depress* or MDD)) OR ("Anxiety Disorders" OR "Anxiety" OR anxi*) OR ("Phobic Disorders" OR phobi*) OR ("Schizophrenia" Or (schizophreni* or hebephreni*)) OR ("Somatoform Disorders" OR "Medically Unexplained Symptoms" OR ( ((somatoform* or somati* or (medically AND unexplained) or briquet or pain) AND (disorder* or syndrome* or symptom*)))) OR ("Dissociative Disorders" OR  (dissociative AND (disorder* or hysteri* or reaction*)) or dissociation*) OR ("Hysteria" or hysteri*) OR ("Mood Disorders" OR ((affective* or mood*) AND (disorder* or disease* or illness* or symptom*))) OR ("Stress Disorders, Post-Traumatic" OR (PTSD or ((posttrauma* or trauma*) AND (stress* or neurose*)) or "combat disorder" or "war disorder")) OR ("Cognition Disorders" OR ( ((cognitive or cognition or mental or neurocognitive) AND (dysfunction* or decline* or impairment* or deterioration* or disorder* or illness* or disease*)))) OR ("Personality Disorders" OR "personality disorder") OR ("Disruptive, Impulse Control, and Conduct Disorders" OR ("impulse control disorder" or "intermittent explosive disorder")) OR ("Feeding and Eating Disorders" OR ((eating or appetite or feeding) AND disorder*)) OR ("Bipolar Disorder" OR ((bipolar or mani*) AND (disorder* or illness* or disease*))) OR ("Obsessive-Compulsive Disorder" OR OCD* or ((obsess* or compulsi*) AND (disorder* or illness* or disease* or neuros*))) OR ("Panic Disorder" OR ((panic AND (attack* or disorder*)))) OR ("Agoraphobia" OR

| |
|---|
| agoraphobi*) OR ("Neurotic Disorders" OR (neuros* or "neurotic disorder" or psychoneuros*))) OR (("Communicable Diseases" OR (((communic* or contag* or transmi* or infect*) AND (disease* or infection* or illness*)))) OR ("Bacterial Infections" OR ( bacteri* infection*)) OR ("Conjunctivitis") OR ("HIV" OR "Human immuno deficiency virus") OR ("Acquired Immunodeficiency Syndrome" OR (AIDS or "immunodeficiency associated virus" or (immun* deficiency associated virus) or (acquired immunodeficiency syndrome*) or (acquired immun* deficiency syndrome*))) OR ("Buruli Ulcer" OR ( Bairnsdale or Buruli)) OR ("Onchocerciasis" OR (onchocer*)) OR ("Hepatitis B" OR "Hepatitis C" OR (hepatitis)) OR ("Leishmaniasis" OR (leishmania*)) OR ("Leprosy" OR (lepros* or hansen*)) OR ("Elephantiasis, Filarial" OR (elephantias* or filaria*)) OR ("Trachoma" OR ((egyptian ophthalmia*) or trachoma*)) OR ("Chikungunya Fever" OR (chickungunya or chikungunya)) OR ("Taeniasis" OR taenia*) OR ("Cysticercosis" OR cysticercos*) OR ("Echinococcosis" OR (hydatid or echinococc*)) OR ("Chagas Disease" OR (trypanosom* or chagas)) OR ("Trypanosomiasis" OR ("sleeping sickness")) OR ("Encephalitis, Japanese" OR (japanese encephalitis)) OR ("Syphilis"))) AND ("Tuberculosis" OR Tuberculos* OR TB OR koch*) AND ((multiple AND (ill* or disease* or condition* or syndrom* or disorder*)) OR (((Cooccur* or co-occur* or coexist* or co-exist* or multipl* or concord* or discord* or long-term or physical*) AND (disease* or ill* or care or condition* or disorder* or health* or medication* or symptom* or syndrom*))) OR (comorbid* or multimorbid* or co-occurren* or co-morbid* or multi-morbid* or Multidisease* or multi-disease*) OR ("Comorbidity" OR "Multimorbidity" OR "Multiple Chronic Conditions")) |
| 0 |
